# Patient2Sentence: Large Language Model-based Semantic Compression for Oncology Trial Eligibility Screening

**DOI:** 10.1101/2025.11.14.25340276

**Authors:** Gerson Hiroshi Yoshinari Júnior, William Caetano Silva Goulart, Ana Beatriz Oliveira Urbano, Maressa Mouty Rabello, Marina Moreira Zorzetto, Sanderson Oliveira de Macedo, Luciano Magalhães Vitorino

**Author notes:** Corresponding author: **Gerson Hiroshi Yoshinari Júnior, MD, PhD**, Professor, Faculty of Medicine of Itajubá-FMIT, Itajubá, Minas Gerais, Brazil.

## Abstract

Efficient clinical trial recruitment in oncology is constrained by the need to interpret long, heterogeneous electronic health records (EHRs) that remain largely unstructured and difficult to automate. We present Patient2Sentence (P2S), a large language model–based framework that performs semantic compression of full oncology EHRs into concise, standardized natural-language “patient sentences” while preserving eligibility-defining clinical logic. Using 75 fully synthetic EHRs modeled after the KATHERINE, MONARCH-E, and OLYMPIA breast cancer trials, we compared trial eligibility classifications derived from full clinical narratives with those obtained exclusively from compressed patient sentences. Eligibility decisions were evaluated against expert adjudication using agreement metrics and paired statistical testing. Sentence-based classifications achieved 94.7% concordance with expert judgments (Cohen’s κ = 0.83), with no statistically significant difference in diagnostic accuracy compared to full-record assessments (McNemar’s p = 1.00), demonstrating non-inferiority of semantic compression for eligibility screening. Trial-specific agreement reached 100% for MONARCH-E, 96% for OLYMPIA, and 88% for KATHERINE, indicating robust performance across heterogeneous eligibility criteria. Semantic compression reduced token consumption by an average of 67.1%, corresponding to a threefold gain in computational efficiency without loss of reasoning fidelity. By reformulating complex oncologic records into human-interpretable, semantically dense sentences, P2S provides an explainable, scalable, and privacy-preserving approach to AI-assisted trial screening. This framework advances biomedical computing by enabling interoperable processing of unstructured clinical narratives and supports the integration of large language models into translational research workflows aimed at accelerating clinical trial recruitment.

## Introduction

Clinical decision-making generates a vast and complex body of unstructured data, ranging from free-text medical notes and pathology reports to semi-structured electronic health records (EHRs) ^1^. Despite this abundance, the heterogeneity and fragmentation of clinical information often prevent artificial intelligence (AI) systems from fully leveraging it ^1, 2^. In recent years, large language models (LLMs) have emerged as a transformative paradigm in medicine, capable of understanding clinical narratives, synthesizing evidence, and assisting in diagnostic reasoning ^2, 3^. These models have redefined how clinical information can be represented and processed, suggesting that language-based representations may bridge the gap between human reasoning and computational inference^3^.

However, existing clinical AI frameworks, such as those based on EHR embeddings, ontology-driven systems, or task-specific architectures like ClinicalBERT and GatorTron, still depend on rigid data structures and narrow vocabularies, which hinder generalization and interoperability across specialties and institutions ^4^. While models such as BioGPT and MedAlpaca have expanded the applicability of LLMs to biomedical and clinical text through domain-specific pretraining and instruction tuning, they continue to face challenges in integrating structured numerical data with the narrative and temporal complexity of clinical reasoning^5^. Moreover, most existing models neglect the causal and contextual dimensions that physicians naturally employ when describing a patient’s evolution over time^6^. As a result, the bridge between human clinical language and machine-interpretable data remains incomplete.

Integrating heterogeneous clinical information streams therefore requires frameworks capable of representing both factual and contextual elements of patient care in a unified way^7^. Recent review have demonstrated that AI can effectively support clinical decision-making when multimodal data sources are harmonized, reinforcing the need for robust data-integration frameworks^8^. Inspired by this premise and by the Cell2Sentence (C2S)^9^ paradigm, which encodes transcriptomic profiles as natural-language “cell sentences” for LLM training, we hypothesized that each patient could be represented as a single, semantically dense sentence that preserves eligibility-defining information for oncology trials.

Based on this hypothesis, we propose Patient2Sentence (P2S): a conceptual and computational framework that represents individual patients as structured textual sentences capturing clinical, temporal, and contextual information in a standardized format. In P2S, each “patient sentence” condenses key features from structured data (such as biomarkers, comorbidities, and treatments) and unstructured text (such as progress notes and clinical impressions) into a compact natural-language representation interpretable by foundation models. This transformation enables LLMs originally trained for general language understanding to process, reason, and generalize across diverse clinical contexts, effectively linking free-text narratives with structured health data. Emerging research on the generation of synthetic patient records using LLMs supports this direction, demonstrating that textual encoding can preserve essential clinical logic while protecting patient privacy^10^.

As a proof of concept, we applied the P2S framework to three major breast cancer trials—KATHERINE^11^, MONARCH-E^12^, and OLYMPIA^13^—simulating synthetic patient records that either met or failed inclusion and exclusion criteria. For each synthetic record, a corresponding patient sentence was generated, and the eligibility classification obtained from the sentence alone was compared with that derived from the full clinical record. This initial experiment tests whether critical eligibility logic can be retained through textual compression, supporting the feasibility of P2S as a faithful and explainable representation of patient-level data.

By translating complex clinical information into standardized natural language, Patient2Sentence lays the groundwork for interoperable, interpretable, and privacy-preserving clinical AI. This perspective aligns with recent discussions that LLMs can serve as medical knowledge bases, provided they maintain transparency, reliability, and ethical safeguards^3, 14^. Beyond research applications, such as eligibility screening, audit automation, and virtual clinical trials—P2S contributes to a broader conceptual shift in medical AI: one where LLMs not only process clinical data but also “speak the language of patients” with fidelity, accountability, and reproducibility.

## Methods

### Study design and reporting guideline

This work adopts a simulation-based diagnostic accuracy design to evaluate whether Patient2Sentence (P2S) preserves trial-eligibility logic after semantic compression of oncology electronic health records (EHRs). Only fully synthetic data were used, with no access to real patient records or identifiable information at any stage. We designed and reported the study in accordance with the STARD-AI guideline for AI-centered diagnostic test accuracy studies, with explicit description of dataset provenance, the AI index test, the reference standard, and the statistical analysis^15^.

### Data sources and trial eligibility logic

We derived all synthetic cases from the published inclusion and exclusion criteria of three pivotal adjuvant breast cancer trials: KATHERINE (HER2-positive disease)^11^, MONARCH-E (high-risk HR+/HER2− disease)^12^, and OLYMPIA (germline BRCA1/2-mutated disease)^13^. The eligibility rules from these clinical trial protocols served as the medical source for determining which cases should be classified as eligible or ineligible. No hospital EHR systems, cancer registries, or real-world datasets were consulted or linked at any stage.

#### Synthetic EHR Generation and Model Configuration

Long-form synthetic EHRs were generated using GPT-5 (OpenAI, San Francisco, CA, USA) through the ChatGPT web interface between October 20 and November 5, 2025, using the October 2025 model snapshot with default parameters, so that temperature, top-p, and other stochastic controls remained unmodified to mirror standard user interaction. The CO-STAR prompting strategy requested detailed oncology records including demographics, breast cancer subtype, stage, risk features, comorbidities, prior treatments, and time-resolved clinical information, with explicit variation in ethnicity (White, Black, Asian, Mixed-race, Indigenous) and age ranges typical of adult breast cancer; because the selected trials focus on breast cancer, all records described female patients. All runs used the same GPT-5 model snapshot and the same web-based inference environment, and the sampling process was not seeded and not averaged across multiple generations: each synthetic record was generated and then classified for trial eligibility in a single pass. The synthetic EHRs were intentionally designed without missing fields relevant to eligibility, each record underwent manual inspection to confirm basic clinical coherence before inclusion, and no indeterminate outputs were observed—GPT-5 consistently returned a binary eligibility label in the requested format.

### Sample size and case spectrum

A total of 75 synthetic EHRs were generated, with 25 cases per trial. Each trial included 5 cases that met all inclusion criteria and 20 cases that violated at least one key inclusion or exclusion rule, intentionally creating a 5:20 ratio to stress-test exclusion logic rather than approximate real-world prevalence. The sample size was defined pragmatically as a proof-of-concept dataset without a formal power calculation. All 75 records demonstrated clinical coherence and proceeded to the final analysis set.

### Reference standard (human expert)

A board-certified radiation oncologist with more than 10 years of clinical experience in breast cancer served as the reference standard reader. The oncologist reviewed each full synthetic EHR and issued a binary eligibility judgment (“Included” or “Excluded”) based on the corresponding trial protocol. The oncologist knew that each subset contained both eligible and ineligible cases and was not formally blinded to the study design, which may introduce verification bias that we address in the Discussion. We treated these human labels as the ground truth for all accuracy analyses.

#### AI index test: Patient2Sentence (P2S)

The index test consisted of the P2S semantic compression followed by eligibility assessment using only the compressed representation. In the first step, GPT-5 converted each long-form synthetic EHR into a single standardized “patient sentence” summarizing tumor subtype, stage, biomarker status, comorbidities, treatments, temporal relationships, and major risk modifiers. In the second step, the same GPT-5 instance classified trial eligibility based solely on the P2S sentence. A fixed, pre-specified eligibility prompt instructed the model to return “Included” or “Excluded” and a brief justification. The identical eligibility prompt was applied to the full EHRs and to the P2S sentences to isolate the effect of semantic compression. No training, fine-tuning, or model adaptation was performed; evaluation occurred in a zero-shot setting using predefined prompts.

#### Outcomes

The primary outcome was the concordance between the P2S-based eligibility classification (index test) and the human expert classification (reference standard) at the individual-case level, including agreement stratified by trial (KATHERINE^13^, MONARCH-E^12^, OLYMPIA^13^). Secondary outcomes included overall and trial-specific percent agreement between AI-based classification on full EHRs and the reference standard, and the reduction in token count between full EHRs and P2S sentences expressed as percentage reduction.

Discordant patterns (false positives and false negatives) were described by trial in an exploratory manner. No formal subgroup or fairness analyses across demographic strata were performed because the dataset was small and entirely synthetic, and this limitation is explicitly reported.

### Statistical analysis

Agreement between the index test and the reference standard was summarized using percent agreement and Cohen’s kappa (κ). Confidence intervals (95% CIs) were calculated using the Wilson score method for proportions and standard large-sample approximations for κ. McNemar’s test for paired nominal data compared eligibility decisions derived from the full EHR and from the P2S representation, adopting a two-sided significance threshold of α = 0.05. All analyses were conducted at the case level, with results reported for each trial and for the pooled dataset. No adjustment for multiple comparisons was applied because this proof-of-concept study has an exploratory aim and a small sample size.

## Results

**Table 1.**
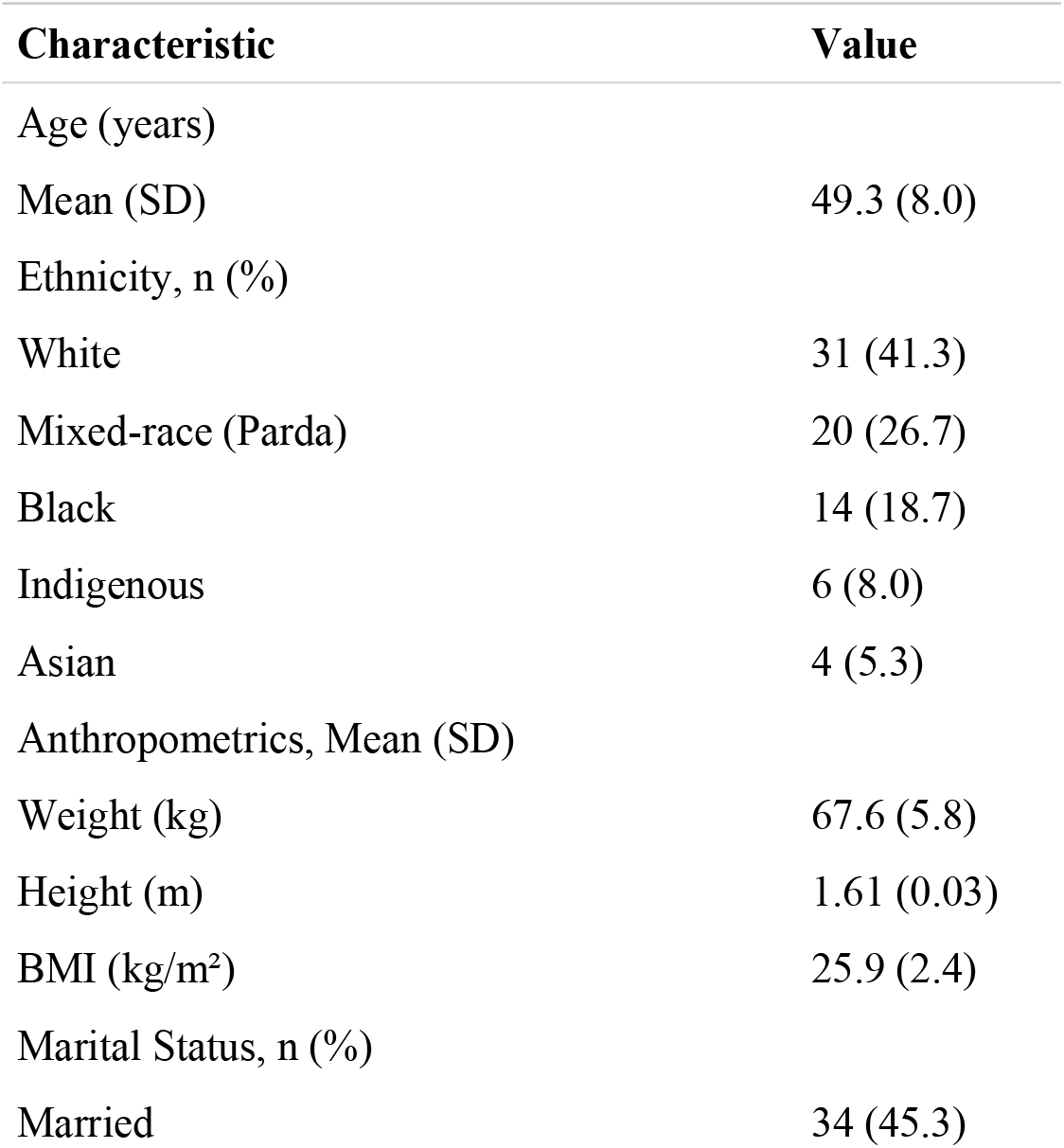
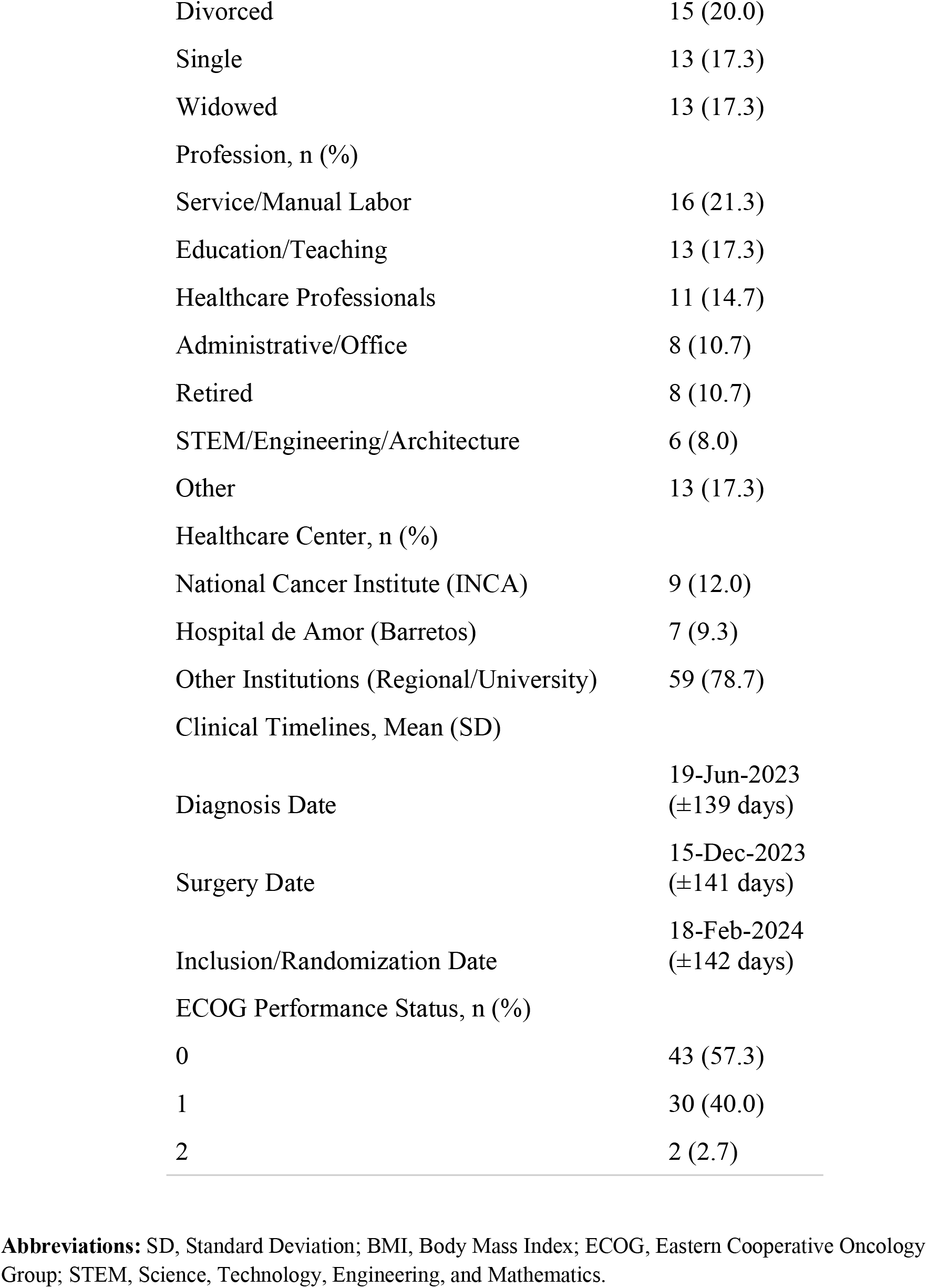
Demographic and Clinical Characteristics of the Synthetic Patient Population (N = 75).

| **Characteristic** | **Value** |
| --- | --- |
| Age (years) |  |
| Mean (SD) | 49.3 (8.0) |
| Ethnicity, n (%) |  |
| White | 31 (41.3) |
| Mixed-race (Parda) | 20 (26.7) |
| Black | 14 (18.7) |
| Indigenous | 6 (8.0) |
| Asian | 4 (5.3) |
| Anthropometrics, Mean (SD) |  |
| Weight (kg) | 67.6 (5.8) |
| Height (m) | 1.61 (0.03) |
| BMI (kg/m<sup>2</sup>) | 25.9 (2.4) |
| Marital Status, n (%) |  |
| Married | 34 (45.3) |
| Divorced | 15 (20.0) |
| Single | 13 (17.3) |
| Widowed | 13 (17.3) |
| Profession, n (%) |  |
| Service/Manual Labor | 16 (21.3) |
| Education/Teaching | 13 (17.3) |
| Healthcare Professionals | 11 (14.7) |
| Administrative/Office | 8 (10.7) |
| Retired | 8 (10.7) |
| STEM/Engineering/Architecture | 6 (8.0) |
| Other | 13 (17.3) |
| Healthcare Center, n (%) |  |
| National Cancer Institute (INCA) | 9 (12.0) |
| Hospital de Amor (Barretos) | 7 (9.3) |
| Other Institutions (Regional/University) | 59 (78.7) |
| Clinical Timelines, Mean (SD) |  |
| Diagnosis Date | 19-Jun-2023<br>(±139 days) |
| Surgery Date | 15-Dec-2023<br>(±141 days) |
| Inclusion/Randomization Date | 18-Feb-2024<br>(±142 days) |
| ECOG Performance Status, n (%) |  |
| 0 | 43 (57.3) |
| 1 | 30 (40.0) |
| 2 | 2 (2.7) |

**Abbreviations:** SD, Standard Deviation; BMI, Body Mass Index; ECOG, Eastern Cooperative Oncology Group; STEM, Science, Technology, Engineering, and Mathematics.

Seventy-five synthetic clinical cases were generated across the KATHERINE, MONARCH-E, and OLYMPIA breast cancer trials, each comprising five eligible and twenty ineligible virtual patients. All records represented adult female oncology cases showing variability in age, ethnicity, comorbidities, and ECOG performance status, consistent with expected clinical diversity in adjuvant breast cancer populations. The Patient2Sentence (P2S) framework successfully converted every full synthetic electronic health record (EHR) into a single sentence encoding all eligibility-relevant clinical attributes.

Across the pooled dataset, sentence-based eligibility decisions demonstrated 94.7% agreement with the validated full-record standard (71/75; 95% CI 87.1–97.9%), corresponding to a Cohen’s κ of 0.83, indicating almost-perfect agreement. McNemar’s test showed no statistically significant difference between eligibility classifications derived from full records and P2S sentences (p = 1.00), supporting the non-inferiority of semantic compression for trial screening tasks.

Trial-stratified performance mirrored these findings: concordance reached 88.0% in KATHERINE (κ = 0.65), 100% in MONARCH-E (κ = 1.00), and 96.0% in OLYMPIA (κ = 0.86). Four discrepancies occurred in total—three in KATHERINE (two false-positives and one false-negative) and one in OLYMPIA (false-negative); no mismatches were identified in MONARCH-E. All P2S generations complied with the predefined prompting templates, and no indeterminate AI outputs were observed.

**Table 2.**
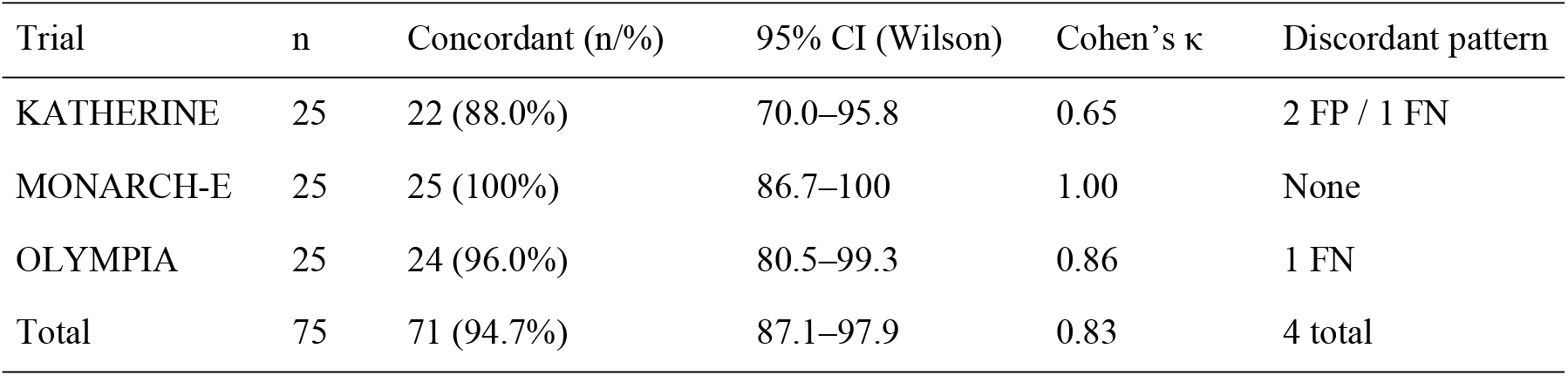
Agreement between full-record and sentence-based eligibility classifications across three breast cancer trials.

Computational efficiency improved substantially following semantic compression. The sentence representation reduced token consumption by 64.2% in KATHERINE (17,462 → 6,247 tokens), 69.0% in MONARCH-E (18,243 → 5,662 tokens), and 68.2% in OLYMPIA (18,329 → 5,826 tokens). Across all trials, P2S achieved an average 67.1% reduction in processing tokens—equivalent to a threefold improvement in computational cost while maintaining high fidelity in eligibility outcomes.

## Discussion

The Patient2Sentence (P2S) framework demonstrated that large language models can compress complex oncology records into concise textual representations without compromising clinical trial screening accuracy. High concordance with full-record eligibility assessments shows that semantic compression can retain essential reasoning pathways, supporting the use of LLMs as reliable mediators between unstructured clinical language and automated inference^16^. In oncology—where timely and accurate screening determines whether patients access potentially beneficial therapies, this ability to convert heterogeneous data into standardized reasoning units represents a practical breakthrough for trial recruitment pipelines^17^.

Operationally, P2S offers a path to near–real-time prescreening within electronic health record (EHR) environments. Continuous eligibility triage can expand the pool of candidates screened per day, reduce manual chart burden, shorten activation timelines, and help research centers meet recruitment targets that are historically difficult to achieve^18, 19^. For patients, earlier identification may translate into accelerated access to innovative therapies and inclusion in high-impact protocols, reinforcing the translational value of this approach across diverse oncology settings^18, 20^.

Conceptually, P2S advances toward interoperable and multimodal clinical reasoning. Although this proof of concept used only textual information, the same architecture can incorporate radiology, digital pathology, and genomics when paired with multimodal transformer models. Encoding visual and quantitative findings in natural-language form enables unified reasoning across modalities and supports pipelines based on clinical “digital twins,” including probabilistic simulation of disease progression and therapeutic response^21^.

Future work will extend the Patient2Sentence (P2S) framework into a dynamic Digital Twin architecture: the Narrative Inference Twin (NIT). This novel DT subclass is defined by its strict reliance on LLM-based inference, utilizing semantic compression exclusively from unstructured clinical narratives, thereby intentionally circumventing the dependence on direct structured data integration. The research focus is validating the NIT’s capacity to accurately infer quantifiable, time-resolved parameters solely from text, positioning it as a scalable solution for prognostic modeling in data-limited environments. This architectural shift necessitates rigorous development of Explainable AI (XAI) to ensure auditable traceability of all inferred parameters back to the source text, a prerequisite for clinical safety and responsible DT adoption^3^.

Explainability and ethical transparency also emerge as strengths. Unlike latent embeddings or proprietary classification heuristics, each “patient sentence” preserves human-readable logic, increasing interpretability and auditability—two criteria frequently cited as barriers to clinical adoption of AI systems^22^. The exclusive use of synthetic textual data reduces privacy and re-identification risk and aligns with current expectations for privacy-preserving validation in multicenter studies and educational environments^23^.

The lower concordance observed in the KATHERINE dataset highlights an important methodological insight^11^. Eligibility for this trial requires contextual interpretation of neoadjuvant timing, residual disease, HER2-directed therapy sequences, and margin status. When compressed into a single sentence, omission or weakening of temporal markers may disproportionately affect classification accuracy^24^. This pattern suggests that prompts designed for complex neoadjuvant scenarios should explicitly encode timelines and sequence-dependent clinical events to preserve decision logic^3^.

This study has limitations: the dataset is small, synthetic, and focused on three oncology trials, limiting generalizability. Validation using real-world EHRs and across additional clinical domains is necessary. Future work should evaluate latency, scalability, robustness across LLM platforms, and the resilience of P2S to demographic and clinical diversity. Deployment using open-weight models trained locally is also recommended to support institutional privacy requirements and regulatory compliance^10^.

Despite these constraints, P2S emerges as a robust and scalable strategy for semantic compression, multimodal integration, and transparent AI-assisted decision support. By transforming unstructured clinical information into standardized natural-language reasoning, P2S provides a foundation for faster, explainable, and privacy-conscious automation of patient-trial matching with direct clinical consequences for oncology research and care delivery^17^.

## Conclusion

The Patient2Sentence (P2S) framework maintained high clinical decision accuracy while reducing computational demand by more than two-thirds, confirming that complex oncologic information can be compressed into concise and standardized natural-language representations without loss of eligibility logic. The approach goes beyond operational gains in trial recruitment: it establishes a reproducible and interpretable pathway for integrating unstructured clinical narratives into AI-assisted decision support. By enabling scalable, explainable, and privacy-preserving automation of patient-trial matching, P2S supports faster identification of eligible candidates, reduces manual workload, and promotes timely access to innovative therapies. These strengths position P2S as a foundational component for future multimodal medical AI—capable of linking text, pathology, imaging, and genomic reasoning within ethical and transparent data ecosystems.

## Data Availability

All data generated in this study are included in the supplementary material.

## Supplementary Material

Supplementary tables with full screening matrices for all synthetic patients and original eligibility assessments are available.

## Credit authorship contribution statement

**BLINDE:** Conceptualization, Supervision, Project administration, Methodology, Investigation, Writing – review & editing. **BLINDE:** Data curation, Investigation, Writing – original draft. **BLINDE:** Data curation, Investigation, Writing – original draft. **BLINDE:** Data curation, Investigation, Writing – original draft. **BLINDE:** Validation, Data curation, Investigation, Writing – review & editing. **BLINDE:** Writing – review & editing, Critical revision of the manuscript, Validation, Approval of the final version. **BLINDE:** Critical revision of the manuscript, Writing – review & editing, Validation, Final approval.

