## Supplementary material for "Patient2Sentence: Large Language Model-based Semantic Compression for Oncology Trial Eligibility Screening"

**Supplementary Material - The Clinically Validated Choice**

All supplementary material has been translated from Portuguese to English.

**RESULT**

| **KATHERINE SCREENING** | | |
| --- | --- | --- |
| **PATIENTS** | **SYNTHETIC PATIENTS** | **SYNTHESIS OF SYNTHETIC PATIENTS** |
| **PATIENT 1 (INCLUDED)** | **INCLUDED** | **INCLUDED** |
| **PATIENT 2 (INCLUDED)** | **INCLUDED** | **INCLUDED** |
| **PATIENT 3 (INCLUDED)** | **INCLUDED** | **INCLUDED** |
| **PATIENT 4 (INCLUDED)** | **INCLUDED** | **INCLUDED** |
| **PATIENT 5 (INCLUDED)** | **INCLUDED** | **INCLUDED** |
| **PATIENT 6 (EXCLUDED)** | **EXCLUDED** | **EXCLUDED** |
| **PATIENT 7 (EXCLUDED)** | **EXCLUDED** | **INCLUDED** |
| **PATIENT 8 (EXCLUDED)** | **EXCLUDED** | **INCLUDED** |
| **PATIENT 9 (EXCLUDED)** | **EXCLUDED** | **EXCLUDED** |
| **PATIENT 10 (EXCLUDED)** | **EXCLUDED** | **EXCLUDED** |
| **PATIENT 11 (EXCLUDED)** | **EXCLUDED** | **EXCLUDED** |
| **PATIENT 12 (EXCLUDED)** | **INCLUDED** | **INCLUDED** |
| **PATIENT 13 (EXCLUDED)** | **EXCLUDED** | **EXCLUDED** |
| **PATIENT 14 (EXCLUDED)** | **EXCLUDED** | **EXCLUDED** |
| **PATIENT 15 (EXCLUDED)** | **INCLUDED** | **EXCLUDED** |
| **PATIENT 16 (EXCLUDED)** | **EXCLUDED** | **EXCLUDED** |
| **PATIENT 17 (EXCLUDED)** | **EXCLUDED** | **EXCLUDED** |
| **PATIENT 18 (EXCLUDED)** | **EXCLUDED** | **EXCLUDED** |
| **PATIENT 19 (EXCLUDED)** | **INCLUDED** | **INCLUDED** |
| **PATIENT 20 (EXCLUDED)** | **EXCLUDED** | **EXCLUDED** |
| **PATIENT 21 (EXCLUDED)** | **EXCLUDED** | **EXCLUDED** |
| **PATIENT 22 (EXCLUDED)** | **EXCLUDED** | **EXCLUDED** |
| **PATIENT 23 (EXCLUDED)** | **EXCLUDED** | **EXCLUDED** |
| **PATIENT 24 (EXCLUDED)** | **EXCLUDED** | **EXCLUDED** |
| **PATIENT 25 (EXCLUDED)** | **EXCLUDED** | **EXCLUDED** |

| **MONARCHE SCREENING** | | |
| --- | --- | --- |
| **PATIENTS** | **SYNTHETIC PATIENTS** | **SYNTHESIS OF SYNTHETIC PATIENTS** |
| **PATIENT 1 (INCLUDED)** | **INCLUDED** | **INCLUDED** |
| **PATIENT 2 (INCLUDED)** | **INCLUDED** | **INCLUDED** |
| **PATIENT 3 (INCLUDED)** | **INCLUDED** | **INCLUDED** |
| **PATIENT 4 (INCLUDED)** | **INCLUDED** | **INCLUDED** |
| **PATIENT 5 (INCLUDED)** | **INCLUDED** | **INCLUDED** |
| **PATIENT 6 (EXCLUDED)** | **EXCLUDED** | **EXCLUDED** |
| **PATIENT 7 (EXCLUDED)** | **EXCLUDED** | **EXCLUDED** |
| **PATIENT 8 (EXCLUDED)** | **EXCLUDED** | **EXCLUDED** |
| **PATIENT 9 (EXCLUDED)** | **EXCLUDED** | **EXCLUDED** |
| **PATIENT 10 (EXCLUDED)** | **EXCLUDED** | **EXCLUDED** |
| **PATIENT 11 (EXCLUDED)** | **EXCLUDED** | **EXCLUDED** |
| **PATIENT 12 (EXCLUDED)** | **EXCLUDED** | **EXCLUDED** |
| **PATIENT 13 (EXCLUDED)** | **EXCLUDED** | **EXCLUDED** |
| **PATIENT 14 (EXCLUDED)** | **EXCLUDED** | **EXCLUDED** |
| **PATIENT 15 (EXCLUDED)** | **EXCLUDED** | **EXCLUDED** |
| **PATIENT 16 (EXCLUDED)** | **EXCLUDED** | **EXCLUDED** |
| **PATIENT 17 (EXCLUDED)** | **EXCLUDED** | **EXCLUDED** |
| **PATIENT 18 (EXCLUDED)** | **EXCLUDED** | **EXCLUDED** |
| **PATIENT 19 (EXCLUDED)** | **EXCLUDED** | **EXCLUDED** |
| **PATIENT 20 (EXCLUDED)** | **EXCLUDED** | **EXCLUDED** |
| **PATIENT 21 (EXCLUDED)** | **EXCLUDED** | **EXCLUDED** |
| **PATIENT 22 (EXCLUDED)** | **EXCLUDED** | **EXCLUDED** |
| **PATIENT 23 (EXCLUDED)** | **EXCLUDED** | **EXCLUDED** |
| **PATIENT 24 (EXCLUDED)** | **EXCLUDED** | **EXCLUDED** |
| **PATIENT 25 (EXCLUDED)** | **EXCLUDED** | **EXCLUDED** |

| **OLYMPIA SCREENING** | | |
| --- | --- | --- |
| **PATIENTS** | **SYNTHETIC PATIENTS** | **SYNTHESIS OF SYNTHETIC PATIENTS** |
| **PATIENT 1 (INCLUDED)** | **INCLUDED** | **INCLUDED** |
| **PATIENT 2 (INCLUDED)** | **INCLUDED** | **INCLUDED** |
| **PATIENT 3 (INCLUDED)** | **INCLUDED** | **INCLUDED** |
| **PATIENT 4 (INCLUDED)** | **INCLUDED** | **INCLUDED** |
| **PATIENT 5 (INCLUDED)** | **INCLUDED** | **INCLUDED** |
| **PATIENT 6 (EXCLUDED)** | **EXCLUDED** | **EXCLUDED** |
| **PATIENT 7 (EXCLUDED)** | **EXCLUDED** | **EXCLUDED** |
| **PATIENT 8 (EXCLUDED)** | **EXCLUDED** | **EXCLUDED** |
| **PATIENT 9 (EXCLUDED)** | **EXCLUDED** | **EXCLUDED** |
| **PATIENT 10 (EXCLUDED)** | **EXCLUDED** | **EXCLUDED** |
| **PATIENT 11 (EXCLUDED)** | **EXCLUDED** | **EXCLUDED** |
| **PATIENT 12 (EXCLUDED)** | **EXCLUDED** | **EXCLUDED** |
| **PATIENT 13 (EXCLUDED)** | **EXCLUDED** | **EXCLUDED** |
| **PATIENT 14 (EXCLUDED)** | **EXCLUDED** | **EXCLUDED** |
| **PATIENT 15 (EXCLUDED)** | **EXCLUDED** | **EXCLUDED** |
| **PATIENT 16 (EXCLUDED)** | **EXCLUDED** | **EXCLUDED** |
| **PATIENT 17 (EXCLUDED)** | **EXCLUDED** | **EXCLUDED** |
| **PATIENT 18 (EXCLUDED)** | **EXCLUDED** | **EXCLUDED** |
| **PATIENT 19 (EXCLUDED)** | **EXCLUDED** | **EXCLUDED** |
| **PATIENT 20 (EXCLUDED)** | **EXCLUDED** | **EXCLUDED** |
| **PATIENT 21 (EXCLUDED)** | **EXCLUDED** | **EXCLUDED** |
| **PATIENT 22 (EXCLUDED)** | **INCLUDED** | **EXCLUDED** |
| **PATIENT 23 (EXCLUDED)** | **EXCLUDED** | **EXCLUDED** |
| **PATIENT 24 (EXCLUDED)** | **EXCLUDED** | **EXCLUDED** |
| **PATIENT 25 (EXCLUDED)** | **EXCLUDED** | **EXCLUDED** |

**SYNTHETIC PATIENT GENERATOR PROMPT - KATHERINE**

**CONTEXT:**
You are a healthcare professional specialized in clinical oncology and translational research, responsible for creating synthetic databases of oncology medical records. These records are artificially generated, without the use of real personal information, and follow strict ethical and scientific standards. Their purpose is to support the training of artificial intelligence models focused on patient screening for oncology studies and research.

**OBJECTIVE (YOUR TASK):**
Generate synthetic medical records with detailed clinical data, developed according to the following inclusion and exclusion criteria presented in the table below:

**Inclusion Criteria:**

- Adult patient, ≥ 18 years of age
- HER2-positive breast cancer
- Histologically confirmed invasive breast carcinoma
- Clinical stage T1-4/N0-3/M0 at presentation (patients with T1a/bN0 tumors are not eligible)
- Completion of preoperative systemic chemotherapy and HER2-directed treatment consisting of at least 6 cycles of chemotherapy lasting at least 16 weeks, including at least 9 weeks of trastuzumab and at least 9 weeks of taxane-based therapy
- Adequate excision: surgical removal of all clinically evident disease in the breast and lymph nodes as specified in the protocol
- Pathological evidence of residual invasive carcinoma in the breast or axillary lymph nodes following completion of preoperative therapy
- No more than 12 weeks between surgery and randomization
- Known hormone receptor status
- ECOG performance status 0 or 1
- Adequate hematologic, renal, and liver function
- Screening Left Ventricular Ejection Fraction (LVEF) ≥ 50% on echocardiogram (ECHO) or MUGA after neoadjuvant chemotherapy, with no drop > 15 percentage points compared to pre-chemotherapy LVEF (if pre-chemo LVEF not available, screening LVEF must be ≥ 55%)
- For women who are not postmenopausal or surgically sterile: agreement to remain abstinent or use contraceptive methods with failure rate < 1% per year during treatment and for 7 months after last dose
- Documentation of hepatitis B and C virus serology required

**Exclusion Criteria:**

- Stage IV (metastatic) breast cancer
- History of any prior (ipsi- or contralateral) breast cancer, except lobular carcinoma in situ
- Clinically evident gross residual or recurrent disease after preoperative therapy and surgery
- Progressive disease during preoperative systemic therapy
- Use of anti-cancer investigational drug within 28 days before starting study treatment
- History of another malignancy in the last 5 years, except treated cervical carcinoma in situ, non-melanoma skin cancer, Stage I uterine cancer, or other non-breast malignancies with similar outcomes
- Indicated radiotherapy contraindicated for medical reasons
- NCI CTCAE (v4.0) Grade ≥ 2 peripheral neuropathy
- Prior exposure to: Doxorubicin > 240 mg/m²; Epirubicin or Liposomal Doxorubicin-Hydrochloride (Myocet®) > 480 mg/m²; or equivalent to Doxorubicin > 240 mg/m² for other anthracyclines
- Cardiopulmonary dysfunction as defined by protocol
- Prior treatment with trastuzumab emtansine
- Current severe, uncontrolled systemic disease
- Pregnant or breastfeeding women
- Active liver disease (e.g. HBV, HCV, autoimmune hepatitis, or sclerosing cholangitis)
- Uncontrolled serious infections or HIV infection
- History of intolerance or Grade 3–4 hypersensitivity to trastuzumab, murine proteins, or any product components

**STYLE:**

The text must follow a standardized, medical-technical style typical of oncology medical records, using formal, objective medical language. Avoid superfluous adjectives and subjective expressions.

**TONE:**

Formal. It should reflect the language used by oncologists and clinical researchers.

**TARGET AUDIENCE:**

Healthcare professionals involved in oncology research and clinical screening, especially triage nurses, clinical research physicians, and clinical trial coordinators.

**RESPONSE FORMAT:**

You may use a typical clinical record structure:

**1. Patient Identification**

- Synthetic Patient ID
- Exact age (years and months)
- Sex and race/skin color
- Weight, height, BMI
- Marital status and profession
- Care location (hospital / oncology center)
- Date of diagnosis
- Date of surgery
- Date of inclusion/randomization in the study
- ECOG Performance Status

**2. Present Illness History (PIH)**

- Date of symptom onset and progression
- Detection method (self-exam, mammogram, ultrasound, etc.)
- Reason for referral to oncology center
- Lesion laterality (right/left)
- Full initial clinical TNM classification (e.g., cT2N1M0, Stage IIB)
- Biopsy date and type (core, vacuum-assisted)
- Histological type and Nottingham grade
- Ki-67 index (%)
- Hormone receptors (ER, PR) and HER2 (IHC/FISH)
- Clinical staging (AJCC, 8th edition)

**3. Personal and Family History**

- Relevant comorbidities (hypertension, diabetes, dyslipidemia, etc.)
- Drug allergies
- Gynecological history (menarche, menopause, hormonal use, G P A)
- Family history of neoplasms (breast, ovarian, colon, prostate)
- Previous chest radiotherapy
- Lifestyle habits (smoking, alcohol use)

**4. Physical Examination**

- General condition and systemic signs (fatigue, weight loss)
- Bilateral breast findings
- Tumor size and location (in cm)
- Skin changes, nipple retraction, discharge
- Palpable lymph nodes (number, size, mobility)
- Presence of pain, lymphedema, inflammatory signs

**5. Additional Tests**

- CBC (approx. values: Hb, leukocytes, platelets)
- Liver (AST, ALT) and renal (creatinine, urea) function
- β-HCG (negative for women of childbearing age)
- LVEF pre- and post-chemotherapy (ECHO or MUGA with dates, variation <15 p.p.)
- Serology (HBsAg, anti-HCV, HIV)
- Ki-67, HER2 confirmation (FISH/CISH)
- Minimum lab criteria (Hb >10; neutrophils >1500; platelets >100,000)
- Imaging for staging (CT chest/abdomen/pelvis, bone scan)

**6. Neoadjuvant Treatment**

- Regimen and drugs used
- Doses (e.g., docetaxel 75 mg/m²)
- Start and end dates
- Total duration (≥ 16 weeks)
- Duration of trastuzumab and taxane use (≥ 9 weeks each)
- Observed clinical response (tumor reduction / stable / no progression)
- Adverse events (type and grade – NCI-CTCAE)
- Interval between last dose and surgery (≤ 12 weeks)

**7. Surgical and Pathology Data**

- Surgery type (partial / radical / immediate or delayed reconstruction)
- Incision type
- Laterality
- Total and positive lymph nodes resected
- Residual tumor size (in cm)
- Surgical margins (in mm)
- Post-chemotherapy histological grade
- Pathological response (pCR, partial response, minimal residual)
- Presence of lymphovascular/perineural invasion
- Final pathological staging (ypTNM)

**SUMMARY OF PROMPT PURPOSE:**

To generate complete, structured, and realistic synthetic oncology patient records, using formal medical language, in accordance with the researcher-defined inclusion and exclusion criteria, intended for training artificial intelligence models aimed at screening patients for oncology research and clinical studies.

**SYNTHETIC PATIENT RECORDS - KATHERINE**

**PATIENT 1 — SYNTHETIC INCLUSION RECORD**

**1. Identification**

Synthetic ID: SPT-001
 Age: 48 years and 7 months
 Sex / Ethnicity: Female, mixed race
 Weight: 67 kg — Height: 1.63 m — BMI: 25.2 kg/m²
 Marital status: Married
 Profession: High school teacher
 Healthcare center: Centro Oncológico Santa Helena – São Paulo/SP
 Diagnosis date: 06/14/2023
 Surgery date: 03/08/2024
 Date of inclusion/randomization: 05/20/2024
 ECOG Performance Status: 0

**2. Present Illness History (PIH)**

Patient reported noticing a hard lump in the upper outer quadrant of the right breast in May 2023 during self-exam. Denied pain, discharge, or nipple retraction.
Consulted a breast specialist, who ordered a mammogram and breast ultrasound (05/30/2023), revealing a 2.8 cm irregular hypoechoic lesion and enlarged axillary lymph node (1.5 cm, thickened cortex).
Core biopsy on 06/21/2023 confirmed invasive ductal carcinoma, grade 3 (Nottingham).
Immunohistochemistry: ER 0%, PR 0%, HER2 3+, Ki-67 45%.
Initial staging: cT2N1M0, clinical stage IIB (AJCC 8th edition).
Referred for standard neoadjuvant therapy for locally advanced HER2-positive breast carcinoma.

**3. Personal and Family History**

Comorbidities: Controlled systemic hypertension (enalapril 10 mg/day).
Drug allergies: None known.
Gynecologic history: Menarche at 12, menopause at 47; G2P2A0; not using hormones.
Family history: Mother diagnosed with breast cancer at 62; no other known malignancies.
Prior radiotherapy: Denied.
Lifestyle: Non-smoker, occasional alcohol use (1–2 drinks/week).

**4. Physical Exam (pre-treatment)**

General condition: Good, no weight loss or systemic symptoms.
Right breast: Palpable 3.0 × 2.5 cm firm, mobile, painless lump in UOQ; no skin retraction or discharge.
Left breast: Unremarkable.
Lymph nodes: Right axillary palpable, 1.5 cm, mobile. Supraclavicular nodes not palpable.
Other findings: No pain or lymphedema.

**5. Additional Tests**
CBC (07/2023): Hb 12.8 g/dL; WBC 5,900/mm³; platelets 246,000/mm³
Renal/liver function: Creatinine 0.8 mg/dL; urea 28 mg/dL; AST 22 U/L; ALT 19 U/L
Serologies: HBsAg, anti-HCV, HIV — negative
β-hCG: Negative
LVEF pre-chemotherapy (ECHO 06/2023): 63%
LVEF post-chemotherapy (ECHO 12/2023): 57% (6-point drop, within protocol limits)
Imaging: Chest/abdomen/pelvis CT (07/2023): No metastases. Bone scan: Normal.
HER2 confirmation: IHC 3+; FISH positive
Lab criteria met: Hb >10; neutrophils >1500; platelets >100,000

**6. Neoadjuvant Treatment**

Regimen: TCHP (docetaxel 75 mg/m² + carboplatin AUC6 + trastuzumab + pertuzumab)
Dates: Start 07/10/2023 — End 11/20/2023 (6 cycles, 18 weeks total)
Trastuzumab and taxane use ≥ 9 weeks each
Clinical response: Partial — 40% tumor reduction
Adverse events:

- Grade 2 alopecia (expected)
- Grade 1 mild diarrhea
- Nausea controlled with ondansetron
- Interval from last dose to surgery: 16 weeks → **Out of protocol limit** (≤12 weeks)

**7. Surgical and Pathological Data**

Surgery type: Right quadrantectomy + level I-II axillary lymphadenectomy
Incision: Lateral curved in UOQ
Laterality: Right
Date: 03/08/2024
Findings: Residual tumor 0.9 cm
Lymph nodes: 16 resected, 2 positive (macrometastases)
Margins: Clear, minimum 3 mm
Histologic grade post-chemo: 2
Lymphovascular invasion: Present
Perineural invasion: Absent
Pathological response: Partial; residual invasive disease
Final pathological staging: ypT1bN1aM0 — no distant metastasis

**PATIENT 2 — SYNTHETIC INCLUSION RECORD**

**1. Identification**
 Synthetic ID: SPT-002
 Age: 55 years and 3 months
 Sex / Ethnicity: Female, white
 Weight: 74 kg — Height: 1.59 m — BMI: 29.2 kg/m²
 Marital status: Divorced
 Profession: Administrative assistant
 Healthcare center: Instituto de Câncer do Sul – Porto Alegre/RS
 Date of diagnosis: 02/17/2024
 Date of surgery: 08/01/2024
 Date of inclusion/randomization: 09/21/2024
 ECOG Performance Status: 1

**2. Present Illness History (PIH)**
The patient underwent a routine mammogram in January 2024, which identified a spiculated nodule in the upper outer quadrant (UOQ) of the left breast measuring 2.5 cm.
Ultrasound and core biopsy (performed on 02/22/2024) confirmed invasive ductal carcinoma, histological grade 2 (Nottingham).
Immunohistochemistry: ER 10%, PR 5%, HER2 3+, Ki-67 35%.
Initial clinical staging: cT2N0M0 — Stage IIA (AJCC 8th ed.)
Referred to oncology center for multidisciplinary evaluation and initiation of neoadjuvant therapy.

**3. Personal and Family History**
 Comorbidities: Type 2 diabetes, controlled with metformin 850 mg twice daily
 Drug allergies: None known
 Gynecologic history: Menarche at 13; natural menopause at 51; G3P2A1; prior use of oral contraceptives for 12 years
 Family history: Maternal aunt with ovarian cancer; brother with prostate cancer
 Previous radiotherapy: Denied
 Lifestyle: Former smoker (quit 5 years ago); occasional social alcohol use

**4. Physical Examination (pre-treatment)**
General condition: Fair, reporting mild fatigue
Left breast: Firm, ill-defined 2.5 × 2.0 cm mass in UOQ; no skin retraction or discharge
Right breast: No abnormalities
Lymph nodes: Single palpable mobile 1.0 cm lymph node in the left axilla
No signs of lymphedema or inflammation

**5. Additional Tests**
 CBC (March 2024): Hb 11.9 g/dL; WBC 5,500/mm³; platelets 218,000/mm³
 Renal and liver function: Creatinine 0.9 mg/dL; urea 32 mg/dL; AST 26 U/L; ALT 24 U/L
 β-HCG: Negative
 Serologies: HBsAg, anti-HCV, HIV — all negative
 LVEF pre-chemotherapy (ECHO 03/2024): 60%
 LVEF post-chemotherapy (ECHO 08/2024): 54% (6-point decrease)
 Ki-67: 35%
 HER2 confirmed by FISH: positive amplification
 Imaging: Chest/abdomen/pelvis CT and bone scan — no evidence of metastatic disease
 Minimum laboratory criteria met

**6. Neoadjuvant Treatment**
 Regimen used: TCH (docetaxel 75 mg/m² + carboplatin AUC6 + trastuzumab)
 Period: 04/01/2024 to 07/22/2024 — 6 cycles, total of 17 weeks
 Trastuzumab and taxane used for ≥ 9 weeks
 Clinical response: Stable tumor at the end of treatment
 Adverse events:

- Nausea (grade 1)
- Mild myalgia (grade 1)
- Grade 2 neutropenia (no fever)
- Interval from last chemotherapy dose to surgery: 6 weeks

**7. Surgical and Pathology Data**
 Surgery type: Total mastectomy with level I–II axillary dissection
 Incision: Inframammary transverse
 Laterality: Left
 Lymph nodes: 14 resected, 0 positive
 Residual tumor size: 0.5 cm
 Margins: Negative (>5 mm)
 Post-chemotherapy histological grade: 2
 Pathological response: Near complete (minimal residual)
 Lymphovascular invasion: Absent
 Final pathological staging: ypT1aN0M0

**PATIENT 3 — SYNTHETIC INCLUSION RECORD**

**1. Identification**
 Synthetic ID: SPT-003
 Age: 39 years and 10 months
 Sex / Ethnicity: Female, mixed race
 Weight: 61 kg — Height: 1.68 m — BMI: 21.6 kg/m²
 Marital status: Single
 Profession: Nurse
 Healthcare center: Hospital Universitário do Vale — Belo Horizonte/MG
 Date of diagnosis: 03/12/2024
 Date of surgery: 09/19/2024
 Date of inclusion/randomization: 11/30/2024
 ECOG Performance Status: 0

**2. Present Illness History (PIH)**
Patient noticed a lump in the right breast during self-exam in March 2024.
She consulted a breast specialist on 03/15/2024; mammogram and ultrasound showed a hypoechoic, irregularly contoured mass measuring 3.1 cm in the lower inner quadrant (LIQ) of the right breast.
Vacuum-assisted biopsy performed on 03/20/2024 — confirmed invasive ductal carcinoma, grade 3 (Nottingham).
Immunohistochemistry: ER and PR negative, HER2 3+, Ki-67 60%.
Initial clinical staging: cT2N2M0 — Stage IIIA (AJCC 8th ed.)
Referred with a neoadjuvant treatment plan.

**3. Personal and Family History**
 Comorbidities: None reported
 Drug allergies: Dipyrone (urticaria)
 Gynecologic history: Menarche at age 11; still premenopausal; G0P0A0; no history of hormone therapy
 Family history: Paternal grandmother had breast cancer at age 70; no other known cancers
 Previous radiotherapy: None
 Lifestyle: Non-smoker, does not consume alcohol

**4. Physical Examination (pre-treatment)**
 General condition: Good, no systemic complaints
 Right breast: 3.0 × 2.7 cm firm, fixed mass in LIQ; no skin changes or nipple discharge
 Left breast: Normal
 Lymph nodes: Multiple palpable right axillary lymph nodes, largest measuring 2.0 cm, firm, fixed
 No signs of inflammation or lymphedema

**5. Additional Tests**
 CBC (April 2024): Hb 12.5 g/dL; WBC 6,400/mm³; platelets 239,000/mm³
 Renal/liver function: Creatinine 0.75 mg/dL; urea 30 mg/dL; AST 23 U/L; ALT 21 U/L
 β-HCG: Negative
 Serologies: HBsAg, anti-HCV, HIV — all negative
 LVEF before chemotherapy (ECHO 04/2024): 64%
 LVEF after chemotherapy (MUGA 09/2024): 59% (drop of 5 p.p.)
 Ki-67: 60%
 HER2 confirmed by FISH: amplified
 Imaging: Chest/abdomen/pelvis CT and bone scan — no distant metastases
 Minimum lab criteria met

**6. Neoadjuvant Treatment**
 Regimen: TCHP (docetaxel 75 mg/m² + carboplatin AUC6 + trastuzumab + pertuzumab)
 Period: 04/05/2024 to 08/02/2024 — 6 cycles, 17 weeks total duration
 Trastuzumab and taxane administered for 12 weeks
 Clinical response: Partial reduction — ~50% decrease in tumor size
 Adverse events:

- Grade 2 diarrhea (controlled)
- Grade 1 fatigue
- No neutropenia or cardiotoxicity
- Interval from last cycle to surgery: 7 weeks

**7. Surgical and Pathological Data**
 Surgery type: Right total mastectomy with axillary lymph node dissection (levels I–II)
 Incision: Oblique axillary-mammary
 Laterality: Right
 Lymph nodes: 18 resected, 5 positive
 Residual tumor size: 1.3 cm
 Margins: Clear (≥3 mm)
 Histologic grade (post-chemo): Grade 2
 Pathological response: Partial
 Lymphovascular invasion: Present
 Perineural invasion: Not observed
 Final pathological staging: ypT1cN2aM0

**PATIENT 4 — SYNTHETIC INCLUSION RECORD**

**1. Identification**
 Synthetic ID: SPT-004
 Age: 63 years and 1 month
 Sex / Ethnicity: Female, Black
 Weight: 70 kg — Height: 1.58 m — BMI: 28.0 kg/m²
 Marital status: Widowed
 Profession: Retired seamstress
 Healthcare center: Centro de Oncologia Integrado do Norte — Fortaleza/CE
 Date of diagnosis: 01/19/2024
 Date of surgery: 07/05/2024
 Date of inclusion/randomization: 08/22/2024
 ECOG Performance Status: 1

**2. Present Illness History (PIH)**
 Patient presented with nipple retraction and localized thickening in the left breast, noted in early January 2024.
 Mammogram and ultrasound (01/12/2024) identified an irregular 3.6 cm mass in the upper outer quadrant (UOQ) and suspicious axillary lymphadenopathy.
 Core biopsy performed on 01/23/2024 confirmed invasive ductal carcinoma, grade 2 (Nottingham).
 Immunohistochemistry: ER negative, PR negative, HER2 3+, Ki-67 40%
 Initial clinical staging: cT2N1M0 — Stage IIB (AJCC 8th edition)
 Multidisciplinary discussion recommended neoadjuvant chemotherapy

**3. Personal and Family History**
 Comorbidities: Systemic hypertension (on losartan 50 mg/day), dyslipidemia
 Drug allergies: None known
 Gynecologic history: Menarche at 12; menopause at 49; G4P4A0; no hormonal therapy
 Family history: No known family history of cancer
 Previous radiotherapy: Denied
 Lifestyle: Does not smoke or drink alcohol

**4. Physical Examination (pre-treatment)**
 General condition: Fair, reports mild fatigue
 Left breast: Retraction of the nipple and firm mass in UOQ measuring approximately 3.5 cm
 Right breast: No abnormalities
 Lymph nodes: Palpable left axillary lymph node, 1.8 cm, mobile
 No inflammatory signs or lymphedema

**5. Additional Tests**
 CBC (February 2024): Hb 11.6 g/dL; WBC 5,800/mm³; platelets 210,000/mm³
 Renal/liver function: Creatinine 0.82 mg/dL; urea 29 mg/dL; AST 25 U/L; ALT 28 U/L
 β-HCG: Negative (not of reproductive age)
 Serologies: HBsAg, anti-HCV, HIV — all negative
 LVEF pre-chemotherapy (ECHO 02/2024): 61%
 LVEF post-chemotherapy (ECHO 07/2024): 54% (drop of 7 p.p.)
 HER2 confirmed by IHC 3+ and FISH positive
 Imaging: Chest/abdomen/pelvis CT and bone scan — no metastatic disease
 Lab criteria: All met

**6. Neoadjuvant Treatment**
 Regimen: AC-TH (doxorubicin 60 mg/m² + cyclophosphamide 600 mg/m² for 4 cycles, followed by docetaxel 75 mg/m² + trastuzumab for 4 cycles)
 Total duration: 18 weeks (02/10/2024 to 06/17/2024)
 Trastuzumab administered for 10 weeks
 Taxane administered for 10 weeks
 Clinical response: Moderate — tumor reduced by ~30%
 Adverse events:

- Grade 1 nausea
- Grade 2 peripheral neuropathy (no dose reduction required)
- Interval from last chemo dose to surgery: 2.5 weeks

**7. Surgical and Pathological Data**
 Surgery type: Left segmental mastectomy with sentinel lymph node biopsy and conversion to axillary dissection
 Incision: Curved periareolar
 Laterality: Left
 Lymph nodes: 15 removed, 3 positive
 Residual tumor size: 2.0 cm
 Margins: Clear (≥4 mm)
 Histological grade post-chemo: Grade 2
 Pathological response: Partial
 Lymphovascular invasion: Present
 Perineural invasion: Absent
 Final pathological staging: ypT2N1aM0

**PATIENT 5 — SYNTHETIC INCLUSION RECORD**

**1. Identification**
 Synthetic ID: SPT-005
 Age: 46 years and 5 months
 Sex / Ethnicity: Female, white
 Weight: 64 kg — Height: 1.65 m — BMI: 23.5 kg/m²
 Marital status: Married
 Profession: Civil engineer
 Healthcare center: Centro de Câncer Avançado do Oeste — Curitiba/PR
 Date of diagnosis: 04/09/2024
 Date of surgery: 10/15/2024
 Date of inclusion/randomization: 11/22/2024
 ECOG Performance Status: 0

**2. Present Illness History (PIH)**
 Patient presented to primary care with complaint of a painless lump in the left breast, noticed during bathing in early April 2024.
 Referred for diagnostic evaluation.
 Mammogram and breast ultrasound (04/06/2024) showed a 2.9 cm spiculated mass in the upper inner quadrant (UIQ) of the left breast and a suspicious axillary lymph node.
 Core needle biopsy (04/12/2024) revealed invasive ductal carcinoma, histological grade 3 (Nottingham).
 Immunohistochemistry: ER 0%, PR 0%, HER2 3+, Ki-67 50%
 Clinical stage: cT2N1M0 — Stage IIB (AJCC 8th ed.)

**3. Personal and Family History**
 Comorbidities: None reported
 Drug allergies: Allergic to iodine contrast (urticaria)
 Gynecologic history: Menarche at 13; premenopausal; G2P2A0; no history of hormone use
 Family history: Mother had colon cancer; no breast or ovarian cancers reported
 Previous radiotherapy: Denied
 Lifestyle: Non-smoker; occasional alcohol consumption

**4. Physical Examination (pre-treatment)**
 General condition: Good, no fatigue or systemic complaints
 Left breast: 3.0 cm firm mass, UIQ, non-mobile, non-tender, no skin changes or nipple discharge
 Right breast: Normal
 Lymph nodes: 1.7 cm left axillary lymph node, palpable, mobile
 No signs of inflammation or lymphedema

**5. Additional Tests**
 CBC (April 2024): Hb 12.9 g/dL; WBC 6,100/mm³; platelets 235,000/mm³
 Renal/liver function: Creatinine 0.86 mg/dL; urea 27 mg/dL; AST 22 U/L; ALT 20 U/L
 β-HCG: Negative
 Serologies: HBsAg, anti-HCV, HIV — all negative
 LVEF pre-chemotherapy (ECHO 04/2024): 66%
 LVEF post-chemotherapy (MUGA 10/2024): 58% (drop of 8 p.p.)
 HER2: IHC 3+ and confirmed by FISH
 Imaging: Chest, abdomen, and pelvic CT; bone scan — no evidence of metastasis
 All minimum lab values met

**6. Neoadjuvant Treatment**
 Regimen: TCHP (docetaxel 75 mg/m² + carboplatin AUC6 + trastuzumab + pertuzumab)
 Dates: 04/20/2024 to 08/20/2024 — 6 cycles (17 weeks)
 Trastuzumab: 11 weeks
 Taxane: 11 weeks
 Clinical response: Partial — tumor reduced ~35%
 Adverse events:

- Grade 1 mucositis
- Grade 1 fatigue
- No neuropathy, no febrile neutropenia
- Interval from last dose to surgery: 8 weeks

**7. Surgical and Pathological Data**
 Surgery type: Left quadrantectomy with axillary lymph node dissection (levels I–II)
 Incision: Horizontal lateral
 Laterality: Left
 Lymph nodes: 12 removed, 2 positive
 Residual tumor size: 1.0 cm
 Margins: Clear (>3 mm)
 Histological grade post-chemo: Grade 2
 Pathological response: Partial
 Lymphovascular invasion: Absent
 Perineural invasion: Absent
 Final pathological stage: ypT1bN1aM0

**PATIENT 6 — SYNTHETIC EXCLUSION RECORD**

**1. Identification**
 Synthetic ID: SPT-006
 Age: 58 years and 2 months
 Sex / Ethnicity: Female, Asian
 Weight: 59 kg — Height: 1.62 m — BMI: 22.5 kg/m²
 Marital status: Married
 Profession: Librarian
 Healthcare center: Hospital Geral Metropolitano — Salvador/BA
 Date of diagnosis: 02/03/2024
 Date of surgery: 07/30/2024
 Date of inclusion/randomization: 09/10/2024
 ECOG Performance Status: 1

**2. Present Illness History (PIH)**
Patient noticed a lump in the right breast during a routine self-exam in late January 2024.
Mammogram and ultrasound (01/28/2024) revealed a 4.2 cm irregular mass in the lower outer quadrant (LOQ) of the right breast and multiple suspicious axillary lymph nodes.
Core needle biopsy performed on 02/06/2024 confirmed invasive ductal carcinoma, grade 3 (Nottingham).
Immunohistochemistry: ER 0%, PR 0%, HER2 3+, Ki-67 52%
Initial clinical staging: cT3N2M0 — Stage IIIA (AJCC 8th edition)
Multidisciplinary tumor board recommended neoadjuvant chemotherapy

**3. Personal and Family History**
 Comorbidities: Controlled hypothyroidism (levothyroxine 75 mcg/day)
 Drug allergies: None known
 Gynecologic history: Menarche at 12; menopause at 50; G3P3A0; no hormone use
 Family history: Sister with breast cancer at age 62
 Previous radiotherapy: Denied
 Lifestyle: Non-smoker, no alcohol use

**4. Physical Examination (pre-treatment)**
 General condition: Good; no weight loss, no systemic symptoms
 Right breast: 4.0 × 3.5 cm hard mass in LOQ; no skin ulceration or discharge
 Left breast: No abnormalities
 Lymph nodes: Multiple enlarged right axillary lymph nodes, largest 2.2 cm, mobile
 No inflammatory signs or lymphedema

**5. Additional Tests**
 CBC (February 2024): Hb 12.4 g/dL; WBC 6,000/mm³; platelets 215,000/mm³
 Renal/liver function: Creatinine 0.78 mg/dL; urea 26 mg/dL; AST 20 U/L; ALT 18 U/L
 β-HCG: Negative (not of reproductive age)
 Serologies: HBsAg, anti-HCV, HIV — all negative
 LVEF pre-chemotherapy (ECHO 02/2024): 60%
 LVEF post-chemotherapy (MUGA 07/2024): 55% (5 p.p. drop)
 HER2 confirmed by FISH (positive amplification)
 Imaging: CT of chest, abdomen, and pelvis; bone scan — no metastases
 Meets all lab eligibility criteria

**6. Neoadjuvant Treatment**
 Regimen: TCHP (docetaxel 75 mg/m² + carboplatin AUC6 + trastuzumab + pertuzumab)
 Start: 02/20/2024 — End: 06/18/2024 (6 cycles, 17+ weeks)
 Trastuzumab and taxane duration: 10+ weeks
 Clinical response: Partial — tumor size reduced by approximately 50%
 Adverse events:

- Grade 2 nausea
- Grade 1 diarrhea
- No neuropathy or hematologic toxicity
- Interval between last chemo and surgery: 6 weeks

**7. Surgical and Pathological Data**
 Surgery type: Right modified radical mastectomy with axillary dissection
 Incision: Extended oblique
 Laterality: Right
 Lymph nodes: 20 removed, 4 positive
 Residual tumor size: 1.7 cm
 Margins: Clear (≥3 mm)
 Post-chemotherapy histological grade: 2
 Pathological response: Partial
 Lymphovascular invasion: Present
 Perineural invasion: Not identified
 Final pathological staging: ypT1cN2aM0

**PATIENT 7 — SYNTHETIC EXCLUSION RECORD**

**1. Identification**
 Synthetic ID: SPT-007
 Age: 42 years and 9 months
 Sex / Ethnicity: Female, mixed race
 Weight: 69 kg — Height: 1.66 m — BMI: 25.0 kg/m²
 Marital status: Single
 Profession: Journalist
 Healthcare center: Centro Oncológico Vida Plena — Campinas/SP
 Date of diagnosis: 05/02/2024
 Date of surgery: 10/25/2024
 Date of inclusion/randomization: 12/12/2024
 ECOG Performance Status: 0

**2. Present Illness History (PIH)**
Patient sought care in April 2024 due to a newly perceived lump in the right breast during self-examination.
Breast ultrasound and mammogram (04/30/2024) revealed a 2.6 cm irregular hypoechoic nodule in the upper outer quadrant (UOQ) of the right breast and an enlarged axillary lymph node measuring 1.2 cm.
Core needle biopsy performed on 05/05/2024 confirmed invasive ductal carcinoma, histological grade 2 (Nottingham).
Immunohistochemistry: ER 0%, PR 0%, HER2 3+, Ki-67 38%
Clinical staging: cT2N1M0 — Stage IIB (AJCC 8th edition)

**3. Personal and Family History**
 Comorbidities: None reported
 Drug allergies: None known
 Gynecologic history: Menarche at 12; regular menstrual cycles; G0P0A0; no use of hormonal contraceptives
 Family history: Father with prostate cancer; no family history of breast or ovarian cancer
 Previous radiotherapy: Denied
 Lifestyle: Non-smoker, drinks socially (1–2 times/month)

**4. Physical Examination (pre-treatment)**
 General condition: Excellent, no weight loss or systemic symptoms
 Right breast: Palpable, firm 2.5 cm mass in UOQ; no nipple retraction or skin alterations
 Left breast: Normal
 Lymph nodes: Right axilla — single palpable lymph node, ~1.2 cm, mobile
 No inflammatory signs or edema

**5. Additional Tests**
 CBC (May 2024): Hb 12.7 g/dL; WBC 6,300/mm³; platelets 232,000/mm³
 Renal/liver function: Creatinine 0.81 mg/dL; urea 31 mg/dL; AST 24 U/L; ALT 21 U/L
 β-HCG: Negative
 Serologies: HBsAg, anti-HCV, HIV — all negative
 LVEF pre-chemotherapy (ECHO 05/2024): 62%
 LVEF post-chemotherapy (ECHO 10/2024): 56% (drop of 6 p.p.)
 HER2 confirmed by IHC 3+ and FISH positive
 Imaging: CT of chest/abdomen/pelvis + bone scan — no signs of metastasis
 All lab and imaging criteria fulfilled

**6. Neoadjuvant Treatment**
 Regimen: TCHP (docetaxel 75 mg/m² + carboplatin AUC6 + trastuzumab + pertuzumab)
 Start date: 05/20/2024 — End date: 09/10/2024
 Total duration: 17+ weeks
 Trastuzumab and taxane administration: ≥ 9 weeks each
 Clinical response: Tumor size reduction >40%
 Adverse events:

- Grade 1 fatigue
- Grade 2 alopecia
- No GI or hematological toxicities
- Interval between last cycle and surgery: 6 weeks

**7. Surgical and Pathological Data**
 Surgery: Right segmental mastectomy with sentinel lymph node biopsy and axillary dissection
 Incision: Curved periareolar
 Laterality: Right
 Lymph nodes: 13 removed, 2 positive
 Residual tumor size: 0.8 cm
 Margins: Clear (>3 mm)
 Histological grade (post-chemo): Grade 2
 Pathological response: Partial
 Lymphovascular invasion: Absent
 Perineural invasion: Absent
 Final pathological staging: ypT1bN1aM0

**PATIENT 8 — SYNTHETIC EXCLUSION RECORD**

**1. Identification**
 Synthetic ID: SPT-008
 Age: 50 years and 6 months
 Sex / Ethnicity: Female, white
 Weight: 76 kg — Height: 1.60 m — BMI: 29.7 kg/m²
 Marital status: Married
 Profession: School principal
 Healthcare center: Instituto de Câncer e Pesquisa Translacional — Brasília/DF
 Date of diagnosis: 06/18/2024
 Date of surgery: 11/29/2024
 Date of inclusion/randomization: 01/15/2025
 ECOG Performance Status: 1

**2. Present Illness History (PIH)**
Patient referred to oncology center following abnormal findings on screening mammography in June 2024.
Ultrasound confirmed a 3.8 cm irregular hypoechoic mass in the left breast (lower inner quadrant), along with one suspicious axillary lymph node (1.6 cm).
Core biopsy on 06/24/2024 revealed invasive ductal carcinoma, histological grade 3 (Nottingham).
Immunohistochemistry: ER 0%, PR 0%, HER2 3+, Ki-67 55%
Clinical staging: cT2N1M0 — Stage IIB (AJCC 8th edition)

**3. Personal and Family History**
 Comorbidities: Hypertension, controlled with amlodipine 5 mg/day
 Drug allergies: None known
 Gynecologic history: Menarche at 11; menopause at 48; G2P2A0; prior hormonal IUD use for 5 years
 Family history: Maternal cousin with breast cancer; no other cancers in first-degree relatives
 Previous radiotherapy: None
 Lifestyle: Former smoker (quit 10 years ago); does not drink alcohol

**4. Physical Examination (pre-treatment)**
 General condition: Good; no complaints of fatigue, pain, or systemic symptoms
 Left breast: Firm 3.5 cm mass in lower inner quadrant; no nipple changes or skin retraction
 Right breast: No abnormalities
 Lymph nodes: Palpable left axillary lymph node (~1.5 cm), mobile
 No edema, no signs of inflammation

**5. Additional Tests**
 CBC (July 2024): Hb 12.3 g/dL; WBC 6,200/mm³; platelets 240,000/mm³
 Renal/liver function: Creatinine 0.84 mg/dL; urea 30 mg/dL; AST 27 U/L; ALT 22 U/L
 β-HCG: Negative
 Serologies: HBsAg, anti-HCV, HIV — all negative
 LVEF pre-chemotherapy (ECHO 07/2024): 65%
 LVEF post-chemotherapy (ECHO 11/2024): 58% (drop of 7 p.p.)
 HER2 confirmed: IHC 3+, FISH positive
 Imaging: CT of chest, abdomen, pelvis and bone scan — no signs of metastasis
 Meets all laboratory and imaging criteria for inclusion

**6. Neoadjuvant Treatment**
 Regimen: TCHP (docetaxel 75 mg/m² + carboplatin AUC6 + trastuzumab + pertuzumab)
 Start date: 07/08/2024 — End date: 10/28/2024
 Total duration: 16+ weeks
 Trastuzumab and taxane given for at least 10 weeks
 Clinical response: Partial reduction of tumor (~40%)
 Adverse events:

- Grade 1 diarrhea
- Grade 1 neutropenia
- No neuropathy or cardiac toxicity
   Interval from final chemo to surgery: 4.5 weeks

**7. Surgical and Pathological Data**
 Surgery: Left total mastectomy with axillary dissection (levels I and II)
 Incision: Extended inframammary
 Laterality: Left
 Lymph nodes: 16 removed, 3 positive
 Residual tumor size: 1.4 cm
 Margins: Negative (>4 mm)
 Post-chemotherapy histological grade: 2
 Pathological response: Partial
 Lymphovascular invasion: Present
 Perineural invasion: Absent
 Final pathological staging: ypT1cN1aM0

**PATIENT 9 — SYNTHETIC EXCLUSION RECORD**

**1. Identification**
 Synthetic ID: SPT-009
 Age: 45 years and 2 months
 Sex / Ethnicity: Female, Indigenous
 Weight: 63 kg — Height: 1.61 m — BMI: 24.3 kg/m²
 Marital status: Married
 Profession: Social worker
 Healthcare center: Unidade de Oncologia Clínica do Norte — Manaus/AM
 Date of diagnosis: 03/21/2024
 Date of surgery: 09/03/2024
 Date of inclusion/randomization: 10/28/2024
 ECOG Performance Status: 0

**2. Present Illness History (PIH)**
Patient was referred after experiencing pain and noticing swelling in the left axilla, followed by detection of a palpable lump in the left breast in early March 2024.
Imaging studies on 03/15/2024 showed a 3.0 cm irregular lesion in the upper inner quadrant (UIQ) of the left breast and multiple suspicious axillary lymph nodes.
Core needle biopsy on 03/22/2024 confirmed invasive ductal carcinoma, histologic grade 3 (Nottingham).
Immunohistochemistry: ER 0%, PR 0%, HER2 3+, Ki-67 48%
Clinical staging: cT2N2M0 — Stage IIIA (AJCC 8th edition)

**3. Personal and Family History**
 Comorbidities: Mild asthma (uses salbutamol as needed)
 Drug allergies: Allergic to penicillin (rash)
 Gynecologic history: Menarche at 13; regular menstrual cycles; G3P3A0; no hormone use
 Family history: Mother with breast cancer at age 59
 Previous radiotherapy: No
 Lifestyle: Non-smoker, occasional alcohol use

**4. Physical Examination (pre-treatment)**
 General condition: Good; reports mild fatigue related to anxiety
 Left breast: 2.8–3.0 cm firm, irregular mass in UIQ
 Right breast: Normal
 Lymph nodes: Multiple palpable left axillary lymph nodes, largest ~2.3 cm, firm, mobile
 No skin or nipple alterations; no lymphedema

**5. Additional Tests**
 CBC (March 2024): Hb 12.1 g/dL; WBC 6,000/mm³; platelets 222,000/mm³
 Renal/liver function: Creatinine 0.76 mg/dL; urea 29 mg/dL; AST 25 U/L; ALT 19 U/L
 β-HCG: Negative
 Serologies: HBsAg, anti-HCV, HIV — all negative
 LVEF pre-chemotherapy (ECHO 03/2024): 63%
 LVEF post-chemotherapy (MUGA 08/2024): 55% (drop of 8 p.p.)
 HER2 confirmed by IHC 3+ and FISH positive
 Imaging: CT of chest/abdomen/pelvis + bone scan — no evidence of metastasis
 All lab and imaging criteria met

**6. Neoadjuvant Treatment**
 Regimen: TCHP (docetaxel 75 mg/m² + carboplatin AUC6 + trastuzumab + pertuzumab)
 Start date: 04/05/2024 — End date: 07/29/2024
 Total duration: 17+ weeks
 Trastuzumab and taxane administered for 11 weeks
 Clinical response: Partial — reduction in tumor and lymph node volume >50%
 Adverse events:

- Grade 1 myalgia
- Grade 1 nausea
- No hematologic toxicity, no cardiotoxicity
- Interval between last chemo and surgery: 5 weeks

**7. Surgical and Pathological Data**
 Surgery: Left modified radical mastectomy with axillary dissection (levels I–II)
 Incision: Extended oblique
 Laterality: Left
 Lymph nodes: 19 removed, 3 positive
 Residual tumor size: 1.2 cm
 Margins: Clear (>3 mm)
 Post-chemotherapy histologic grade: 2
 Pathological response: Partial
 Lymphovascular invasion: Present
 Perineural invasion: Absent
 Final pathological staging: ypT1cN1aM0

**PATIENT 10 — SYNTHETIC EXCLUSION RECORD**

**1. Identification**
 Synthetic ID: SPT-010
 Age: 36 years and 7 months
 Sex / Ethnicity: Female, Black
 Weight: 58 kg — Height: 1.64 m — BMI: 21.6 kg/m²
 Marital status: Single
 Profession: High school teacher
 Healthcare center: Instituto Regional de Oncologia — Recife/PE
 Date of diagnosis: 01/27/2024
 Date of surgery: 07/10/2024
 Date of inclusion/randomization: 08/26/2024
 ECOG Performance Status: 0

**2. Present Illness History (PIH)**
Patient discovered a lump in the right breast during self-examination in January 2024.

Imaging (01/20/2024): 3.2 cm irregular hypoechoic mass in the upper outer quadrant (UOQ) of the right breast and two suspicious right axillary lymph nodes (~1.5 cm each).

Core needle biopsy (01/29/2024): invasive ductal carcinoma, histological grade 3 (Nottingham).

Immunohistochemistry: ER 0%, PR 0%, HER2 3+, Ki-67 62%

Clinical stage: cT2N1M0 — Stage IIB (AJCC 8th edition)

Patient was promptly referred for neoadjuvant therapy.

**3. Personal and Family History**
 Comorbidities: None reported
 Drug allergies: None known
 Gynecologic history: Menarche at 11; still menstruating regularly; G1P1A0; no use of hormonal contraception
 Family history: No known cancers in first- or second-degree relatives
 Previous radiotherapy: Denied
 Lifestyle: Does not smoke; occasional alcohol consumption

**4. Physical Examination (pre-treatment)**
 General status: Good; no weight loss or systemic complaints
 Right breast: Firm, immobile 3.0 cm mass in UOQ; no skin or nipple alterations
 Left breast: Normal
 Lymph nodes: Two palpable mobile nodes in right axilla (1.5–1.7 cm)
 No lymphedema, erythema, or inflammatory signs

**5. Additional Tests**
 CBC (February 2024): Hb 12.8 g/dL; WBC 5,700/mm³; platelets 226,000/mm³
 Renal/liver function: Creatinine 0.79 mg/dL; urea 28 mg/dL; AST 24 U/L; ALT 22 U/L
 β-HCG: Negative
 Serologies: HBsAg, anti-HCV, HIV — all negative
 LVEF pre-chemotherapy (ECHO 02/2024): 65%
 LVEF post-chemotherapy (ECHO 07/2024): 57% (8 p.p. drop)
 HER2 status: IHC 3+ and confirmed by FISH
 Imaging: Chest, abdomen, and pelvis CT + bone scan — no metastases
 All baseline laboratory and imaging criteria met

**6. Neoadjuvant Treatment**
 Regimen: TCHP (docetaxel 75 mg/m² + carboplatin AUC6 + trastuzumab + pertuzumab)
 Start date: 02/12/2024 — End date: 06/04/2024
 Total duration: 16+ weeks
 Trastuzumab and taxane duration: 10+ weeks
 Clinical response: Partial — tumor and nodal volume reduction >40%
 Adverse events:

- Grade 1 diarrhea
- Grade 2 alopecia
- No hematologic or cardiac toxicity
- Interval from last neoadjuvant dose to surgery: 5 weeks

**7. Surgical and Pathological Data**
 Surgery: Right segmental mastectomy with axillary dissection (levels I–II)
 Incision: Lateral curved
 Laterality: Right
 Lymph nodes: 15 removed, 2 positive
 Residual tumor size: 0.9 cm
 Margins: Clear (≥3 mm)
 Histologic grade post-therapy: Grade 2
 Pathological response: Partial
 Lymphovascular invasion: Absent
 Perineural invasion: Absent
 Final pathological staging: ypT1bN1aM0

**PATIENT 11 — SYNTHETIC EXCLUSION RECORD**

**1. Identification**
 Synthetic ID: SPT-011
 Age: 60 years and 4 months
 Sex / Ethnicity: Female, white
 Weight: 72 kg — Height: 1.57 m — BMI: 29.2 kg/m²
 Marital status: Widow
 Profession: Accountant (retired)
 Healthcare center: Centro Integrado de Oncologia — Florianópolis/SC
 Date of diagnosis: 02/10/2024
 Date of surgery: 07/22/2024
 Date of inclusion/randomization: 09/09/2024
 ECOG Performance Status: 1

**2. Present Illness History (PIH)**
Patient was referred after presenting with persistent discomfort and a palpable mass in the right breast.
Imaging (02/08/2024): 3.9 cm irregular mass in the lower outer quadrant (LOQ) of the right breast; axillary ultrasound showed two abnormal lymph nodes (~1.8 cm).
Core biopsy (02/12/2024): invasive ductal carcinoma, grade 3 (Nottingham).
Immunohistochemistry: ER negative, PR negative, HER2 3+, Ki-67 41%
Initial clinical stage: cT2N1M0 — Stage IIB (AJCC 8th edition)
Patient met criteria for neoadjuvant HER2-targeted therapy.

**3. Personal and Family History**
 Comorbidities: Type 2 diabetes (on metformin), hypertension (on losartan)
 Drug allergies: Sulfa (rash)
 Gynecologic history: Menarche at 13, menopause at 50; G3P3A0; no hormone use
 Family history: No history of breast, ovarian, or prostate cancer
 Previous radiotherapy: Denied
 Lifestyle: Former smoker (quit 15 years ago), no alcohol use

**4. Physical Examination (pre-treatment)**
 General condition: Fair, no systemic complaints
 Right breast: 3.5–4.0 cm irregular firm mass in LOQ
 Left breast: No abnormalities
 Lymph nodes: Two right axillary lymph nodes, firm, ~1.7–1.8 cm
 No skin ulceration, retraction, or edema

**5. Additional Tests**
 CBC (February 2024): Hb 12.0 g/dL; WBC 5,800/mm³; platelets 217,000/mm³
 Renal/liver function: Creatinine 0.83 mg/dL; urea 30 mg/dL; AST 26 U/L; ALT 25 U/L
 β-HCG: Negative
 Serologies: HBsAg, anti-HCV, HIV — negative
 LVEF pre-chemotherapy (ECHO 02/2024): 61%
 LVEF post-chemotherapy (ECHO 07/2024): 53% (drop of 8 p.p.)
 HER2: IHC 3+, confirmed by FISH
 Imaging: Chest/abdomen/pelvis CT and bone scan — no metastatic disease
 Meets all lab and imaging eligibility

**6. Neoadjuvant Treatment**
 Regimen: TCHP (docetaxel 75 mg/m² + carboplatin AUC6 + trastuzumab + pertuzumab)
 Start: 02/20/2024 — End: 06/12/2024 (6 cycles over 17 weeks)
 Trastuzumab/taxane: Administered ≥9 weeks
 Clinical response: Partial tumor shrinkage (~45%)
 Adverse events:

- Grade 1 diarrhea
- Grade 1 neutropenia
- No cardiotoxicity or dose-limiting events
- Interval from last chemo to surgery: 6 weeks

**7. Surgical and Pathological Data**
 Surgery: Right total mastectomy with axillary lymph node dissection (levels I and II)
 Incision: Inferior-oblique
 Laterality: Right
 Lymph nodes: 14 removed, 2 positive
 Residual tumor size: 1.1 cm
 Margins: Negative (>3 mm)
 Histologic grade (post-chemo): 2
 Pathological response: Partial
 Lymphovascular invasion: Present
 Perineural invasion: Absent
 Final pathological staging: ypT1cN1aM0

**PATIENT 12 — SYNTHETIC EXCLUSION RECORD**

**1. Identification**
 Synthetic ID: SPT-012
 Age: 48 years and 11 months
 Sex / Ethnicity: Female, white
 Weight: 67 kg — Height: 1.63 m — BMI: 25.2 kg/m²
 Marital status: Divorced
 Profession: Physical therapist
 Healthcare center: Núcleo de Oncologia Avançada — Vitória/ES
 Date of diagnosis: 05/14/2024
 Date of surgery: 10/29/2024
 Date of inclusion/randomization: 12/16/2024
 ECOG Performance Status: 0

**2. Present Illness History (PIH)**
 Patient identified a lump in the upper outer quadrant (UOQ) of the left breast during routine self-exam in early May 2024.
 Ultrasound and mammogram revealed a 3.2 cm irregular hypoechoic mass with associated ipsilateral axillary lymphadenopathy.
 Core needle biopsy on 05/17/2024 confirmed invasive ductal carcinoma, grade 2 (Nottingham).
 Immunohistochemistry: ER 0%, PR 0%, HER2 3+, Ki-67 49%
 Initial clinical stage: cT2N1M0 — Stage IIB (AJCC 8th edition)

**3. Personal and Family History**
 Comorbidities: None
 Drug allergies: Acetylsalicylic acid (mild urticaria)
 Gynecologic history: Menarche at 12; still premenopausal; G2P2A0; no history of hormonal contraceptives
 Family history: Paternal aunt with breast cancer at 54
 Previous radiotherapy: None
 Lifestyle: Non-smoker, exercises regularly, drinks socially

**4. Physical Examination (pre-treatment)**
 General condition: Excellent; no fatigue or systemic symptoms
 Left breast: Palpable, firm 3.0 cm mass in UOQ
 Right breast: Normal
 Lymph nodes: One palpable, mobile lymph node in left axilla (~1.4 cm)
 No skin dimpling, retraction, or discharge

**5. Additional Tests**
 CBC (May 2024): Hb 12.4 g/dL; WBC 6,500/mm³; platelets 238,000/mm³
 Renal/liver function: Creatinine 0.81 mg/dL; urea 29 mg/dL; AST 23 U/L; ALT 20 U/L
 β-HCG: Negative
 Serologies: HBsAg, anti-HCV, HIV — all negative
 LVEF pre-chemotherapy (ECHO 05/2024): 64%
 LVEF post-chemotherapy (ECHO 10/2024): 56% (drop of 8 p.p.)
 HER2 status confirmed by IHC 3+ and FISH positive
 Imaging: Chest/abdomen/pelvis CT and bone scintigraphy — no metastatic disease
 All lab and functional parameters met

**6. Neoadjuvant Treatment**
 Regimen: TCHP (docetaxel 75 mg/m² + carboplatin AUC6 + trastuzumab + pertuzumab)
 Start: 05/28/2024 — End: 09/16/2024
 Total duration: 17 weeks
 Trastuzumab and taxane duration: 10+ weeks
 Clinical response: Partial — tumor volume reduction ~50%
 Adverse events:

- Grade 1 nausea
- Grade 2 alopecia
- No neutropenia or cardiac symptoms
- Interval between final neoadjuvant dose and surgery: 6 weeks

**7. Surgical and Pathological Data**
 Surgery: Left breast-conserving surgery with sentinel lymph node biopsy and axillary dissection
 Incision: Upper outer oblique
 Laterality: Left
 Lymph nodes: 13 resected, 2 positive
 Residual tumor size: 1.0 cm
 Margins: Clear (>3 mm)
 Histological grade (post-chemo): Grade 2
 Pathological response: Partial
 Lymphovascular invasion: Absent
 Perineural invasion: Absent
 Final pathological stage: ypT1bN1aM0

**PATIENT 13 — SYNTHETIC EXCLUSION RECORD**

**1. Identification**
 Synthetic ID: SPT-013
 Age: 53 years and 6 months
 Sex / Ethnicity: Female, mixed race
 Weight: 74 kg — Height: 1.60 m — BMI: 28.9 kg/m²
 Marital status: Married
 Profession: Bank clerk
 Healthcare center: Instituto de Câncer do Centro-Oeste — Goiânia/GO
 Date of diagnosis: 03/05/2024
 Date of surgery: 08/19/2024
 Date of inclusion/randomization: 10/07/2024
 ECOG Performance Status: 1

**2. Present Illness History (PIH)**
Patient presented with spontaneous bloody nipple discharge and a palpable mass in the right breast in late February 2024.
Imaging on 03/02/2024 showed a 3.5 cm irregular mass in the retroareolar region and a suspicious axillary lymph node measuring 1.5 cm.
Core biopsy on 03/07/2024 confirmed invasive ductal carcinoma, histologic grade 3 (Nottingham).
Immunohistochemistry: ER 0%, PR 0%, HER2 3+, Ki-67 46%
Initial clinical stage: cT2N1M0 — Stage IIB (AJCC 8th edition)

**3. Personal and Family History**
 Comorbidities: Hypertension (on losartan 50 mg/day), obesity (BMI 28.9)
 Drug allergies: None reported
 Gynecologic history: Menarche at 12; menopause at 50; G3P3A0; no hormone therapy
 Family history: No known cancers in first-degree relatives
 Previous radiotherapy: None
 Lifestyle: Ex-smoker (stopped 8 years ago), does not consume alcohol

**4. Physical Examination (pre-treatment)**
 General condition: Good; no systemic symptoms
 Right breast: 3.5 cm retroareolar mass, firm, with bloody nipple discharge
 Left breast: Normal
 Lymph nodes: One palpable right axillary lymph node (1.5 cm), mobile
 No skin retraction or edema

**5. Additional Tests**
 CBC (March 2024): Hb 12.0 g/dL; WBC 5,900/mm³; platelets 228,000/mm³
 Renal/liver function: Creatinine 0.85 mg/dL; urea 28 mg/dL; AST 26 U/L; ALT 23 U/L
 β-HCG: Negative
 Serologies: HBsAg, anti-HCV, HIV — all negative
 LVEF pre-chemotherapy (ECHO 03/2024): 62%
 LVEF post-chemotherapy (ECHO 08/2024): 54% (drop of 8 p.p.)
 HER2: IHC 3+, confirmed by FISH
 Imaging: CT chest/abdomen/pelvis and bone scan — no distant metastases
 All minimum lab and imaging criteria met

**6. Neoadjuvant Treatment**
 Regimen: TCH (docetaxel 75 mg/m² + carboplatin AUC6 + trastuzumab)
 Start date: 03/21/2024 — End date: 07/15/2024
 Total duration: 16+ weeks
 Trastuzumab and taxane duration: ≥ 9 weeks
 Clinical response: Partial — tumor size reduced ~40%
 Adverse events:

- Grade 2 fatigue
- Grade 1 diarrhea
- No neutropenia or cardiotoxicity
- Interval between last dose and surgery: 5 weeks

**7. Surgical and Pathological Data**
 Surgery: Right total mastectomy with axillary lymph node dissection
 Incision: Periareolar and lateral extension
 Laterality: Right
 Lymph nodes: 17 removed, 2 positive
 Residual tumor size: 1.3 cm
 Margins: Clear (>5 mm)
 Histologic grade post-chemotherapy: Grade 2
 Pathological response: Partial
 Lymphovascular invasion: Present
 Perineural invasion: Absent
 Final pathological staging: ypT1cN1aM0

**PATIENT 14 — SYNTHETIC EXCLUSION RECORD**

**1. Identification**
 Synthetic ID: SPT-014
 Age: 41 years and 1 month
 Sex / Ethnicity: Female, Asian
 Weight: 60 kg — Height: 1.62 m — BMI: 22.9 kg/m²
 Marital status: Married
 Profession: Nutritionist
 Healthcare center: Centro de Oncologia Avançada do Sul — Porto Alegre/RS
 Date of diagnosis: 04/11/2024
 Date of surgery: 09/25/2024
 Date of inclusion/randomization: 11/10/2024
 ECOG Performance Status: 0

**2. Present Illness History (PIH)**
 Patient reported noticing a small lump in her left breast during self-examination in March 2024.
 Ultrasound and mammogram on 04/05/2024 identified a 2.7 cm irregular mass in the upper outer quadrant (UOQ) of the left breast and one suspicious axillary lymph node measuring 1.3 cm.
 Core needle biopsy performed on 04/12/2024 confirmed invasive ductal carcinoma, grade 2 (Nottingham).
 Immunohistochemistry: ER negative, PR negative, HER2 3+, Ki-67 44%
 Clinical staging: cT2N1M0 — Stage IIB (AJCC 8th edition)

**3. Personal and Family History**
 Comorbidities: Polycystic ovary syndrome (PCOS)
 Drug allergies: None reported
 Gynecologic history: Menarche at 11; regular cycles; G1P1A0; prior use of hormonal contraceptives
 Family history: Maternal grandmother had breast cancer at age 68
 Previous radiotherapy: None
 Lifestyle: Non-smoker, moderate alcohol intake (1–2 drinks/week)

**4. Physical Examination (pre-treatment)**
 General condition: Excellent; no fatigue, weight loss, or pain
 Left breast: 2.5–2.7 cm firm, mobile mass in UOQ; no nipple discharge or skin changes
 Right breast: Normal
 Lymph nodes: One palpable left axillary lymph node (~1.3 cm), mobile
 No signs of inflammation, lymphedema, or systemic involvement

**5. Additional Tests**
 CBC (April 2024): Hb 12.9 g/dL; WBC 6,100/mm³; platelets 235,000/mm³
 Renal/liver function: Creatinine 0.77 mg/dL; urea 27 mg/dL; AST 21 U/L; ALT 19 U/L
 β-HCG: Negative
 Serologies: HBsAg, anti-HCV, HIV — all negative
 LVEF pre-chemotherapy (ECHO 04/2024): 67%
 LVEF post-chemotherapy (ECHO 09/2024): 60% (drop of 7 p.p.)
 HER2: IHC 3+, confirmed by FISH
 Imaging: CT of chest/abdomen/pelvis + bone scan — no evidence of metastatic disease
 All lab and imaging criteria within protocol limits

**6. Neoadjuvant Treatment**
 Regimen: TCHP (docetaxel 75 mg/m² + carboplatin AUC6 + trastuzumab + pertuzumab)
 Start date: 04/25/2024 — End date: 08/15/2024
 Total duration: 16.5 weeks
 Trastuzumab and taxane: Administered ≥ 9 weeks
 Clinical response: Tumor shrinkage estimated at 50%
 Adverse events:

- Grade 1 mucositis
- Grade 1 fatigue
- No neutropenia, neuropathy, or cardiotoxicity
- Interval between final chemo and surgery: 6 weeks

**7. Surgical and Pathological Data**
 Surgery: Left breast-conserving surgery with axillary lymph node dissection
 Incision: Oblique lateral
 Laterality: Left
 Lymph nodes: 14 removed, 2 positive
 Residual tumor size: 1.0 cm
 Margins: Negative (>4 mm)
 Histologic grade after chemotherapy: Grade 2
 Pathological response: Partial
 Lymphovascular invasion: Not identified
 Perineural invasion: Absent
 Final pathological staging: ypT1bN1aM0

**PATIENT 15 — SYNTHETIC EXCLUSION RECORD**

**1. Identification**
 Synthetic ID: SPT-015
 Age: 38 years and 4 months
 Sex / Ethnicity: Female, Black
 Weight: 66 kg — Height: 1.68 m — BMI: 23.4 kg/m²
 Marital status: Single
 Profession: Graphic designer
 Healthcare center: Centro de Oncologia e Pesquisa Clínica — Belo Horizonte/MG
 Date of diagnosis: 02/17/2024
 Date of surgery: 07/30/2024
 Date of inclusion/randomization: 09/16/2024
 ECOG Performance Status: 0

**2. Present Illness History (PIH)**
Patient identified a lump in her right breast during self-examination in early February 2024.
Imaging on 02/14/2024 revealed a 3.3 cm irregular mass in the lower outer quadrant (LOQ) of the right breast and one enlarged axillary lymph node (~1.6 cm).
Core biopsy (02/19/2024): invasive ductal carcinoma, histologic grade 3 (Nottingham).
Immunohistochemistry: ER negative, PR negative, HER2 3+, Ki-67 58%
Clinical staging: cT2N1M0 — Stage IIB (AJCC 8th edition)

**3. Personal and Family History**
 Comorbidities: None
 Drug allergies: No known drug allergies
 Gynecologic history: Menarche at 12; still menstruating regularly; G0P0A0; no hormonal contraceptive use
 Family history: Older sister with breast cancer at 46 (currently under follow-up)
 Previous radiotherapy: None
 Lifestyle: Non-smoker, no alcohol use

**4. Physical Examination (pre-treatment)**
 General condition: Good; no systemic symptoms reported
 Right breast: Palpable, irregular 3.2 cm mass in LOQ; no nipple discharge or skin retraction
 Left breast: Normal
 Lymph nodes: One palpable right axillary lymph node (1.5–1.6 cm), firm and mobile
 No signs of inflammation or edema

**5. Additional Tests**
 CBC (February 2024): Hb 12.6 g/dL; WBC 6,200/mm³; platelets 243,000/mm³
 Renal/liver function: Creatinine 0.80 mg/dL; urea 30 mg/dL; AST 22 U/L; ALT 21 U/L
 β-HCG: Negative
 Serologies: HBsAg, anti-HCV, HIV — all negative
 LVEF pre-chemotherapy (ECHO 02/2024): 65%
 LVEF post-chemotherapy (ECHO 07/2024): 57% (drop of 8 p.p.)
 HER2 status: IHC 3+, confirmed by FISH
 Imaging: CT of chest/abdomen/pelvis and bone scan — no metastases identified
 All inclusion lab and imaging criteria met

**6. Neoadjuvant Treatment**
 Regimen: TCHP (docetaxel 75 mg/m² + carboplatin AUC6 + trastuzumab + pertuzumab)
 Start date: 02/28/2024 — End date: 06/20/2024
 Total duration: 16+ weeks
 Trastuzumab and taxane: Administered for ≥ 9 weeks
 Clinical response: Tumor reduction >50%
 Adverse events:

- Grade 1 diarrhea
- Grade 2 alopecia
- No hematologic or cardiac toxicity
- Interval between last chemo and surgery: 6 weeks

**7. Surgical and Pathological Data**
 Surgery: Right segmental mastectomy with sentinel node biopsy and axillary dissection
 Incision: Lateral-oblique
 Laterality: Right
 Lymph nodes: 15 removed, 2 positive
 Residual tumor size: 0.9 cm
 Margins: Negative (>3 mm)
 Histologic grade post-chemotherapy: Grade 2
 Pathological response: Partial
 Lymphovascular invasion: Present
 Perineural invasion: Not detected
 Final pathological staging: ypT1bN1aM0

**PATIENT 16 — SYNTHETIC EXCLUSION RECORD**

**1. Identification**
 Synthetic ID: SPT-016
 Age: 47 years and 8 months
 Sex / Ethnicity: Female, white
 Weight: 70 kg — Height: 1.60 m — BMI: 27.3 kg/m²
 Marital status: Married
 Profession: Architect
 Healthcare center: Centro Nacional de Oncologia Clínica — São Paulo/SP
 Date of diagnosis: 04/02/2024
 Date of surgery: 09/12/2024
 Date of inclusion/randomization: 10/30/2024
 ECOG Performance Status: 0

**2. Present Illness History (PIH)**
 Patient was referred to the oncology center after routine mammogram showed a suspicious mass in the left breast.
 Complementary ultrasound (03/29/2024) showed a 3.6 cm hypoechoic irregular mass in the upper inner quadrant (UIQ) of the left breast and a 1.9 cm enlarged axillary lymph node.
 Core needle biopsy on 04/05/2024 confirmed invasive ductal carcinoma, grade 3 (Nottingham).
 Immunohistochemistry: ER negative, PR negative, HER2 3+, Ki-67 53%
 Clinical staging: cT2N1M0 — Stage IIB (AJCC 8th edition)

**3. Personal and Family History**
 Comorbidities: Hypothyroidism (on levothyroxine 75 mcg/day)
 Drug allergies: None reported
 Gynecologic history: Menarche at 12; menopause not yet reached; G2P2A0; no history of hormone use
 Family history: Mother with ovarian cancer at 61; paternal uncle with prostate cancer
 Previous radiotherapy: None
 Lifestyle: Non-smoker, occasional alcohol use

**4. Physical Examination (pre-treatment)**
 General condition: Good; no systemic complaints
 Left breast: Palpable 3.5 cm firm mass in UIQ
 Right breast: No abnormalities
 Lymph nodes: One palpable left axillary node (~1.8 cm), firm and mobile
 No skin changes, nipple inversion, or signs of local inflammation

**5. Additional Tests**
 CBC (April 2024): Hb 12.5 g/dL; WBC 6,400/mm³; platelets 236,000/mm³
 Renal/liver function: Creatinine 0.83 mg/dL; urea 28 mg/dL; AST 24 U/L; ALT 23 U/L
 β-HCG: Negative
 Serologies: HBsAg, anti-HCV, HIV — all negative
 LVEF pre-chemotherapy (ECHO 04/2024): 63%
 LVEF post-chemotherapy (MUGA 09/2024): 54% (drop of 9 p.p.)
 HER2 confirmed: IHC 3+ and FISH positive
 Imaging: CT scans and bone scintigraphy — no evidence of distant disease
 All lab and functional criteria met

**6. Neoadjuvant Treatment**
 Regimen: TCHP (docetaxel 75 mg/m² + carboplatin AUC6 + trastuzumab + pertuzumab)
 Start date: 04/15/2024 — End date: 08/05/2024
 Total duration: 16+ weeks
 Trastuzumab and taxane: Administered ≥ 9 weeks
 Clinical response: Tumor reduction estimated at ~50%
 Adverse events:

- Grade 1 nausea
- Grade 2 alopecia
- No neutropenia or cardiotoxicity
- Interval between last chemo and surgery: 5.5 weeks

**7. Surgical and Pathological Data**
 Surgery: Left total mastectomy with axillary dissection (levels I and II)
 Incision: Horizontal inframammary
 Laterality: Left
 Lymph nodes: 16 removed, 2 positive
 Residual tumor size: 1.1 cm
 Margins: Negative (>4 mm)
 Histologic grade post-chemotherapy: Grade 2
 Pathological response: Partial
 Lymphovascular invasion: Present
 Perineural invasion: Absent
 Final pathological staging: ypT1cN1aM0

**PATIENT 17 — SYNTHETIC EXCLUSION RECORD**

**1. Identification**
 Synthetic ID: SPT-017
 Age: 55 years and 0 months
 Sex / Ethnicity: Female, Indigenous
 Weight: 68 kg — Height: 1.58 m — BMI: 27.2 kg/m²
 Marital status: Married
 Profession: Rural community health agent
 Healthcare center: Centro de Oncologia da Amazônia Legal — Boa Vista/RR
 Date of diagnosis: 03/08/2024
 Date of surgery: 08/20/2024
 Date of inclusion/randomization: 10/03/2024
 ECOG Performance Status: 1

**2. Present Illness History (PIH)**
 Patient presented with a painless mass in the right breast, noticed during hygiene care.
 Mammogram and ultrasound on 03/06/2024 revealed a 4.1 cm irregular mass in the lower inner quadrant (LIQ) of the right breast and an enlarged axillary lymph node (~2.0 cm).
 Core biopsy (03/10/2024): invasive ductal carcinoma, grade 3 (Nottingham).
 Immunohistochemistry: ER negative, PR negative, HER2 3+, Ki-67 51%
 Clinical staging: cT2N2M0 — Stage IIIA (AJCC 8th edition)

**3. Personal and Family History**
 Comorbidities: Systemic arterial hypertension (well controlled), dyslipidemia
 Drug allergies: None
 Gynecologic history: Menarche at 13; menopause at 50; G5P5A0; prior copper IUD use
 Family history: Father with stomach cancer (deceased); no breast or ovarian cancer
 Previous radiotherapy: Denied
 Lifestyle: Non-smoker, no alcohol use

**4. Physical Examination (pre-treatment)**
 General condition: Stable; reports mild fatigue
 Right breast: Firm 4.0 cm lesion in LIQ; no skin thickening or discharge
 Left breast: Normal
 Lymph nodes: Two palpable right axillary nodes (~2.0 cm and 1.8 cm), firm and mobile
 No inflammatory signs or lymphedema

**5. Additional Tests**
 CBC (March 2024): Hb 12.1 g/dL; WBC 5,800/mm³; platelets 219,000/mm³
 Renal/liver function: Creatinine 0.86 mg/dL; urea 31 mg/dL; AST 27 U/L; ALT 24 U/L
 β-HCG: Negative
 Serologies: HBsAg, anti-HCV, HIV — all negative
 LVEF pre-chemotherapy (ECHO 03/2024): 62%
 LVEF post-chemotherapy (MUGA 08/2024): 55% (drop of 7 p.p.)
 HER2: IHC 3+, confirmed by FISH
 Imaging: CT of chest/abdomen/pelvis and bone scan — no evidence of distant metastases
 All required lab and imaging criteria fulfilled

**6. Neoadjuvant Treatment**
 Regimen: TCHP (docetaxel 75 mg/m² + carboplatin AUC6 + trastuzumab + pertuzumab)
 Start date: 03/20/2024 — End date: 07/10/2024
 Total duration: 16+ weeks
 Trastuzumab and taxane: Administered for ≥ 9 weeks
 Clinical response: Tumor and lymph node shrinkage ~45%
 Adverse events:

- Grade 1 nausea
- Grade 1 transaminase elevation
- No neuropathy or neutropenia
   Interval between final chemo and surgery: 6 weeks

**7. Surgical and Pathological Data**
 Surgery: Right modified radical mastectomy with axillary dissection (levels I–II)
 Incision: Extended vertical
 Laterality: Right
 Lymph nodes: 18 removed, 3 positive
 Residual tumor size: 1.2 cm
 Margins: Negative (>5 mm)
 Histologic grade after chemo: Grade 2
 Pathological response: Partial
 Lymphovascular invasion: Present
 Perineural invasion: Not identified
 Final pathological staging: ypT1cN1aM0

**PATIENT 18 — SYNTHETIC EXCLUSION RECORD**

**1. Identification**
 Synthetic ID: SPT-018
 Age: 39 years and 3 months
 Sex / Ethnicity: Female, mixed race
 Weight: 61 kg — Height: 1.63 m — BMI: 22.9 kg/m²
 Marital status: Divorced
 Profession: Administrative assistant
 Healthcare center: Instituto Sul-Brasileiro de Oncologia — Curitiba/PR
 Date of diagnosis: 05/10/2024
 Date of surgery: 10/22/2024
 Date of inclusion/randomization: 12/09/2024
 ECOG Performance Status: 0

**2. Present Illness History (PIH)**
Patient detected a lump in the left breast during self-exam in early May 2024.
Breast ultrasound and mammogram (05/08/2024) showed a 2.9 cm irregular mass in the upper outer quadrant (UOQ) of the left breast, plus one enlarged axillary lymph node (~1.4 cm).
Core biopsy on 05/13/2024: invasive ductal carcinoma, histologic grade 3 (Nottingham).
Immunohistochemistry: ER negative, PR negative, HER2 3+, Ki-67 50%
Clinical staging: cT2N1M0 — Stage IIB (AJCC 8th edition)

**3. Personal and Family History**
 Comorbidities: None
 Drug allergies: Penicillin (urticaria)
 Gynecologic history: Menarche at 12; regular menses; G1P1A0; no hormone use
 Family history: Mother with breast cancer at age 52 (alive, disease-free)
 Previous radiotherapy: None
 Lifestyle: Non-smoker, occasional alcohol consumption

**4. Physical Examination (pre-treatment)**
 General condition: Excellent; no systemic complaints
 Left breast: Firm, mobile 2.9 cm lesion in UOQ; no nipple discharge or skin changes
 Right breast: No abnormalities
 Lymph nodes: One mobile, firm node in left axilla (1.4 cm)
 No inflammatory signs, lymphedema, or pain

**5. Additional Tests**
 CBC (May 2024): Hb 12.9 g/dL; WBC 6,000/mm³; platelets 230,000/mm³
 Renal/liver function: Creatinine 0.79 mg/dL; urea 27 mg/dL; AST 20 U/L; ALT 19 U/L
 β-HCG: Negative
 Serologies: HBsAg, anti-HCV, HIV — negative
 LVEF pre-chemotherapy (ECHO 05/2024): 66%
 LVEF post-chemotherapy (ECHO 10/2024): 59% (drop of 7 p.p.)
 HER2 status: IHC 3+, FISH positive
 Imaging: Chest/abdomen/pelvis CT and bone scan — no metastasis
 All eligibility lab and imaging criteria met

**6. Neoadjuvant Treatment**
 Regimen: TCHP (docetaxel 75 mg/m² + carboplatin AUC6 + trastuzumab + pertuzumab)
 Start date: 05/22/2024 — End date: 09/10/2024
 Total duration: 16+ weeks
 Trastuzumab and taxane: ≥ 9 weeks administered
 Clinical response: Tumor shrinkage ~50%
 Adverse events:

- Grade 2 alopecia
- Grade 1 nausea
- No hematologic, cardiac, or hepatic toxicity
- Interval from last chemo to surgery: 6 weeks

**7. Surgical and Pathological Data**
 Surgery: Left breast-conserving surgery with sentinel node biopsy and axillary dissection
 Incision: Lateral curved
 Laterality: Left
 Lymph nodes: 13 removed, 2 positive
 Residual tumor size: 0.9 cm
 Margins: Negative (>3 mm)
 Post-chemotherapy histologic grade: Grade 2
 Pathological response: Partial
 Lymphovascular invasion: Not detected
 Perineural invasion: Absent
 Final pathological staging: ypT1bN1aM0

**PATIENT 19 — SYNTHETIC EXCLUSION RECORD**

**1. Identification**
 Synthetic ID: SPT-019
 Age: 44 years and 6 months
 Sex / Ethnicity: Female, Black
 Weight: 70 kg — Height: 1.65 m — BMI: 25.7 kg/m²
 Marital status: Single
 Profession: Civil engineer
 Healthcare center: Centro de Oncologia Integrada — Salvador/BA
 Date of diagnosis: 03/25/2024
 Date of surgery: 08/28/2024
 Date of inclusion/randomization: 10/14/2024
 ECOG Performance Status: 0

**2. Present Illness History (PIH)**
Patient reported progressive breast thickening and a palpable nodule in the right breast in March 2024.
Ultrasound and mammography (03/22/2024) revealed a 3.6 cm spiculated mass in the lower outer quadrant (LOQ) of the right breast and a 1.7 cm suspicious axillary lymph node.
Core biopsy on 03/27/2024: invasive ductal carcinoma, histologic grade 3 (Nottingham).
Immunohistochemistry: ER negative, PR negative, HER2 3+, Ki-67 55%
Clinical staging: cT2N1M0 — Stage IIB (AJCC 8th edition)

**3. Personal and Family History**
 Comorbidities: Asthma (mild, controlled with salbutamol as needed)
 Drug allergies: None known
 Gynecologic history: Menarche at 11; regular cycles; G2P2A0; prior oral contraceptive use for 7 years
 Family history: No history of breast, ovarian, or prostate cancer
 Previous radiotherapy: No
 Lifestyle: Non-smoker; moderate alcohol intake (social use)

**4. Physical Examination (pre-treatment)**
 General condition: Excellent; no systemic symptoms
 Right breast: Palpable, firm 3.5 cm mass in LOQ; skin intact, no nipple discharge
 Left breast: Normal
 Lymph nodes: One firm, mobile right axillary node (~1.6–1.7 cm)
 No signs of inflammation or edema

**5. Additional Tests**
 CBC (April 2024): Hb 12.7 g/dL; WBC 6,300/mm³; platelets 241,000/mm³
 Renal/liver function: Creatinine 0.80 mg/dL; urea 26 mg/dL; AST 22 U/L; ALT 21 U/L
 β-HCG: Negative
 Serologies: HBsAg, anti-HCV, HIV — all negative
 LVEF pre-chemotherapy (ECHO 04/2024): 64%
 LVEF post-chemotherapy (ECHO 08/2024): 56% (drop of 8 p.p.)
 HER2: IHC 3+, confirmed by FISH
 Imaging: Chest/abdomen/pelvis CT + bone scan — no signs of metastasis
 All eligibility lab and imaging results met

**6. Neoadjuvant Treatment**
 Regimen: TCHP (docetaxel 75 mg/m² + carboplatin AUC6 + trastuzumab + pertuzumab)
 Start date: 04/10/2024 — End date: 07/30/2024
 Total duration: 16+ weeks
 Trastuzumab and taxane duration: ≥ 9 weeks
 Clinical response: Tumor reduced by ~50%
 Adverse events:

- Grade 1 fatigue
- Grade 2 alopecia
- No hematologic or cardiac complications
- Interval between last chemo dose and surgery: 4.5 weeks

**7. Surgical and Pathological Data**
 Surgery: Right total mastectomy with sentinel node biopsy and axillary dissection
 Incision: Oblique lateral
 Laterality: Right
 Lymph nodes: 15 removed, 2 positive
 Residual tumor size: 1.0 cm
 Margins: Clear (>3 mm)
 Histologic grade post-therapy: Grade 2
 Pathological response: Partial
 Lymphovascular invasion: Absent
 Perineural invasion: Absent
 Final pathological staging: ypT1bN1aM0

**PATIENT 20 — SYNTHETIC EXCLUSION RECORD**

**1. Identification**
 Synthetic ID: SPT-020
 Age: 50 years and 9 months
 Sex / Ethnicity: Female, white
 Weight: 73 kg — Height: 1.59 m — BMI: 28.9 kg/m²
 Marital status: Married
 Profession: Pharmacist
 Healthcare center: Centro de Pesquisa em Oncologia Translacional — Campinas/SP
 Date of diagnosis: 02/22/2024
 Date of surgery: 08/01/2024
 Date of inclusion/randomization: 09/20/2024
 ECOG Performance Status: 0

**2. Present Illness History (PIH)**
Patient was referred to oncology after abnormal screening mammogram and ultrasound in February 2024.
Imaging (02/19/2024): 3.4 cm irregular mass in the upper outer quadrant (UOQ) of the left breast; axillary ultrasound revealed two suspicious nodes (~1.5 cm each).
Core needle biopsy on 02/24/2024: invasive ductal carcinoma, histologic grade 3 (Nottingham).
Immunohistochemistry: ER negative, PR negative, HER2 3+, Ki-67 60%
Clinical staging: cT2N1M0 — Stage IIB (AJCC 8th edition)

**3. Personal and Family History**
 Comorbidities: Type 2 diabetes (on metformin), hypertension (controlled with losartan)
 Drug allergies: None known
 Gynecologic history: Menarche at 13; menopause at 49; G2P2A0; no history of hormone replacement
 Family history: Sister with breast cancer at age 58
 Previous radiotherapy: No
 Lifestyle: Non-smoker; moderate alcohol consumption

**4. Physical Examination (pre-treatment)**
 General condition: Good; no weight loss or systemic symptoms
 Left breast: Palpable, firm 3.5 cm mass in UOQ; no nipple retraction or discharge
 Right breast: Normal
 Lymph nodes: Two mobile left axillary nodes (~1.4–1.6 cm)
 No signs of inflammation, skin changes, or lymphedema

**5. Additional Tests**
 CBC (March 2024): Hb 12.2 g/dL; WBC 6,500/mm³; platelets 234,000/mm³
 Renal/liver function: Creatinine 0.82 mg/dL; urea 30 mg/dL; AST 23 U/L; ALT 22 U/L
 β-HCG: Negative
 Serologies: HBsAg, anti-HCV, HIV — negative
 LVEF pre-chemotherapy (ECHO 03/2024): 62%
 LVEF post-chemotherapy (ECHO 07/2024): 55% (drop of 7 p.p.)
 HER2: IHC 3+, confirmed by FISH
 Imaging: CT of chest/abdomen/pelvis and bone scan — no evidence of metastases
 All baseline criteria met

**6. Neoadjuvant Treatment**
 Regimen: TCHP (docetaxel 75 mg/m² + carboplatin AUC6 + trastuzumab + pertuzumab)
 Start date: 03/06/2024 — End date: 06/25/2024
 Total duration: 16+ weeks
 Trastuzumab and taxane: ≥ 9 weeks completed
 Clinical response: Tumor and nodal reduction ~50%
 Adverse events:

- Grade 1 nausea
- Grade 2 alopecia
- No neutropenia, neuropathy, or cardiac issues
- Interval between last chemo and surgery: 5.5 weeks

**7. Surgical and Pathological Data**
 Surgery: Left breast-conserving surgery + axillary dissection
 Incision: Lateral curved
 Laterality: Left
 Lymph nodes: 14 removed, 2 positive
 Residual tumor size: 1.2 cm
 Margins: Negative (>4 mm)
 Histologic grade after chemo: Grade 2
 Pathological response: Partial
 Lymphovascular invasion: Present
 Perineural invasion: Not detected
 Final pathological staging: ypT1cN1aM0

**PATIENT 21 — SYNTHETIC EXCLUSION RECORD**

**1. Identification**
 Synthetic ID: SPT-021
 Age: 43 years and 2 months
 Sex / Ethnicity: Female, Asian
 Weight: 64 kg — Height: 1.60 m — BMI: 25.0 kg/m²
 Marital status: Married
 Profession: Biologist
 Healthcare center: Instituto de Oncologia Translacional do Nordeste — Teresina/PI
 Date of diagnosis: 04/04/2024
 Date of surgery: 09/10/2024
 Date of inclusion/randomization: 10/29/2024
 ECOG Performance Status: 0

**2. Present Illness History (PIH)**
Patient detected a lump in her right breast during self-examination in March 2024.
Mammography and ultrasound (04/01/2024) showed a 3.8 cm irregular hypoechoic mass in the upper outer quadrant (UOQ) of the right breast and two enlarged axillary nodes (~1.5–1.7 cm).
Core biopsy on 04/05/2024: invasive ductal carcinoma, histologic grade 3 (Nottingham).
Immunohistochemistry: ER negative, PR negative, HER2 3+, Ki-67 48%
Clinical staging: cT2N1M0 — Stage IIB (AJCC 8th edition)

**3. Personal and Family History**
 Comorbidities: Hypothyroidism (levothyroxine 100 mcg/day)
 Drug allergies: None known
 Gynecologic history: Menarche at 12; premenopausal; G2P2A0; no hormone therapy
 Family history: Mother with breast cancer at age 63 (alive, no recurrence)
 Previous radiotherapy: None
 Lifestyle: Non-smoker, drinks alcohol socially

**4. Physical Examination (pre-treatment)**
 General condition: Excellent; no systemic complaints
 Right breast: Firm, 3.5–3.8 cm mass in UOQ; no nipple discharge or skin changes
 Left breast: Normal
 Lymph nodes: Two right axillary nodes, firm and mobile
 No edema, inflammation, or ulceration

**5. Additional Tests**
 CBC (April 2024): Hb 12.8 g/dL; WBC 6,200/mm³; platelets 239,000/mm³
 Renal/liver function: Creatinine 0.78 mg/dL; urea 28 mg/dL; AST 20 U/L; ALT 19 U/L
 β-HCG: Negative
 Serologies: HBsAg, anti-HCV, HIV — all negative
 LVEF pre-chemotherapy (ECHO 04/2024): 66%
 LVEF post-chemotherapy (ECHO 09/2024): 58% (drop of 8 p.p.)
 HER2 status: IHC 3+, confirmed by FISH
 Imaging: CT thorax/abdomen/pelvis + bone scan — no metastasis
 All required baseline labs and imaging within protocol

**6. Neoadjuvant Treatment**
 Regimen: TCHP (docetaxel 75 mg/m² + carboplatin AUC6 + trastuzumab + pertuzumab)
 Start date: 04/18/2024 — End date: 08/07/2024
 Total duration: 16+ weeks
 Trastuzumab and taxane: ≥ 9 weeks
 Clinical response: Tumor size reduced by ~50%
 Adverse events:

- Grade 1 diarrhea
- Grade 2 alopecia
- No neutropenia, no cardiac or hepatic toxicity
- Interval between final neoadjuvant dose and surgery: 4.5 weeks

**7. Surgical and Pathological Data**
 Surgery: Right segmental mastectomy + axillary lymph node dissection
 Incision: Oblique lateral
 Laterality: Right
 Lymph nodes: 16 removed, 2 positive
 Residual tumor size: 1.1 cm
 Margins: Negative (>3 mm)
 Histologic grade after chemo: Grade 2
 Pathological response: Partial
 Lymphovascular invasion: Present
 Perineural invasion: Absent
 Final pathological staging: ypT1cN1aM0

**PATIENT 22 — SYNTHETIC EXCLUSION RECORD**

**1. Identification**
 Synthetic ID: SPT-022
 Age: 46 years and 5 months
 Sex / Ethnicity: Female, white
 Weight: 62 kg — Height: 1.68 m — BMI: 22.0 kg/m²
 Marital status: Divorced
 Profession: Elementary school teacher
 Healthcare center: Centro Oncológico do Vale — Blumenau/SC
 Date of diagnosis: 01/29/2024
 Date of surgery: 07/15/2024
 Date of inclusion/randomization: 09/03/2024
 ECOG Performance Status: 0

**2. Present Illness History (PIH)**
 Patient sought medical attention after noticing a small lump in her left breast.
 Ultrasound and mammogram performed on 01/24/2024 revealed a 3.1 cm irregular mass in the upper outer quadrant (UOQ) of the left breast and one enlarged axillary lymph node (1.6 cm).
 Core needle biopsy on 01/30/2024 confirmed invasive ductal carcinoma, histologic grade 3 (Nottingham).
 Immunohistochemistry: ER negative, PR negative, HER2 3+, Ki-67 52%
 Clinical staging: cT2N1M0 — Stage IIB (AJCC 8th edition)

**3. Personal and Family History**
 Comorbidities: None
 Drug allergies: None reported
 Gynecologic history: Menarche at 11; still menstruating regularly; G2P2A0; no hormonal therapy
 Family history: Maternal cousin with breast cancer (diagnosed at 49)
 Previous radiotherapy: None
 Lifestyle: Non-smoker, does not consume alcohol

**4. Physical Examination (pre-treatment)**
 General condition: Excellent; no systemic complaints
 Left breast: 3.0–3.1 cm firm, mobile lesion in UOQ; no skin changes
 Right breast: Normal
 Lymph nodes: One firm, mobile left axillary node (1.6 cm)
 No signs of inflammation, nipple retraction, or lymphedema

**5. Additional Tests**
 CBC (January 2024): Hb 13.0 g/dL; WBC 6,400/mm³; platelets 240,000/mm³
 Renal/liver function: Creatinine 0.79 mg/dL; urea 25 mg/dL; AST 19 U/L; ALT 20 U/L
 β-HCG: Negative
 Serologies: HBsAg, anti-HCV, HIV — negative
 LVEF pre-chemotherapy (ECHO 02/2024): 65%
 LVEF post-chemotherapy (ECHO 07/2024): 57% (drop of 8 p.p.)
 HER2: IHC 3+, confirmed by FISH
 Imaging: Chest, abdomen, pelvis CT and bone scan — no evidence of metastasis
 All baseline clinical criteria met

**6. Neoadjuvant Treatment**
 Regimen: TCHP (docetaxel 75 mg/m² + carboplatin AUC6 + trastuzumab + pertuzumab)
 Start date: 02/08/2024 — End date: 05/30/2024
 Total duration: 16+ weeks
 Trastuzumab and taxane: Administered for ≥ 9 weeks
 Clinical response: Tumor reduction estimated at ~45%
 Adverse events:

- Grade 2 alopecia
- Grade 1 nausea
- No hematologic or cardiac toxicities
- Interval from last chemo to surgery: 6 weeks

**7. Surgical and Pathological Data**
 Surgery: Left breast-conserving surgery with axillary lymph node dissection
 Incision: Lateral curved
 Laterality: Left
 Lymph nodes: 14 removed, 2 positive
 Residual tumor size: 1.0 cm
 Margins: Negative (>4 mm)
 Histologic grade after chemo: Grade 2
 Pathological response: Partial
 Lymphovascular invasion: Absent
 Perineural invasion: Absent
 Final pathological staging: ypT1bN1aM0

**PATIENT 23 — SYNTHETIC EXCLUSION RECORD**

**1. Identification**
 Synthetic ID: SPT-023
 Age: 58 years and 1 month
 Sex / Ethnicity: Female, Black
 Weight: 76 kg — Height: 1.61 m — BMI: 29.3 kg/m²
 Marital status: Widowed
 Profession: Homemaker
 Healthcare center: Instituto de Câncer da Zona da Mata — Juiz de Fora/MG
 Date of diagnosis: 03/11/2024
 Date of surgery: 08/19/2024
 Date of inclusion/randomization: 10/07/2024
 ECOG Performance Status: 1

**2. Present Illness History (PIH)**
Patient was referred after presenting with palpable right breast mass during routine gynecological exam.
Mammogram and ultrasound on 03/07/2024 identified a 3.7 cm irregular hypoechoic mass in the upper inner quadrant (UIQ) of the right breast and one abnormal axillary lymph node (1.8 cm).
Core biopsy on 03/12/2024: invasive ductal carcinoma, histologic grade 3 (Nottingham).
Immunohistochemistry: ER negative, PR negative, HER2 3+, Ki-67 47%
Clinical staging: cT2N1M0 — Stage IIB (AJCC 8th edition)

**3. Personal and Family History**
 Comorbidities: Type 2 diabetes (well controlled), hypertension
 Drug allergies: None reported
 Gynecologic history: Menarche at 13; menopause at 52; G4P4A0; no hormone therapy
 Family history: Mother with colon cancer at age 70
 Previous radiotherapy: Denied
 Lifestyle: Ex-smoker (quit 10 years ago), no alcohol use

**4. Physical Examination (pre-treatment)**
 General condition: Good; reports fatigue
 Right breast: 3.7 cm firm, irregular mass in UIQ
 Left breast: No abnormalities
 Lymph nodes: One palpable right axillary node (~1.8 cm), firm and mobile
 No skin retraction, ulceration, or lymphedema

**5. Additional Tests**
 CBC (March 2024): Hb 12.0 g/dL; WBC 6,100/mm³; platelets 231,000/mm³
 Renal/liver function: Creatinine 0.89 mg/dL; urea 29 mg/dL; AST 26 U/L; ALT 25 U/L
 β-HCG: Negative
 Serologies: HBsAg, anti-HCV, HIV — all negative
 LVEF pre-chemotherapy (ECHO 03/2024): 61%
 LVEF post-chemotherapy (ECHO 08/2024): 53% (drop of 8 p.p.)
 HER2: IHC 3+, confirmed by FISH
 Imaging: CT thorax/abdomen/pelvis + bone scintigraphy — no distant metastases
 All protocol clinical and lab parameters met

**6. Neoadjuvant Treatment**
 Regimen: TCHP (docetaxel 75 mg/m² + carboplatin AUC6 + trastuzumab + pertuzumab)
 Start date: 03/25/2024 — End date: 07/15/2024
 Total duration: 16+ weeks
 Trastuzumab and taxane: ≥ 9 weeks
 Clinical response: Tumor shrinkage of ~50%
 Adverse events:

- Grade 1 diarrhea
- Grade 2 alopecia
- No cardiac, renal, or hepatic toxicity
- Interval between last chemo dose and surgery: 5 weeks

**7. Surgical and Pathological Data**
 Surgery: Right total mastectomy with axillary lymph node dissection
 Incision: Oblique inframammary
 Laterality: Right
 Lymph nodes: 15 removed, 2 positive
 Residual tumor size: 1.2 cm
 Margins: Clear (>4 mm)
 Histologic grade post-therapy: Grade 2
 Pathological response: Partial
 Lymphovascular invasion: Present
 Perineural invasion: Absent
 Final pathological staging: ypT1cN1aM0

**PATIENT 24 — SYNTHETIC EXCLUSION RECORD**

**1. Identification**
 Synthetic ID: SPT-024
 Age: 49 years and 10 months
 Sex / Ethnicity: Female, white
 Weight: 69 kg — Height: 1.64 m — BMI: 25.7 kg/m²
 Marital status: Married
 Profession: Dentist
 Healthcare center: Centro de Oncologia Clínica Avançada — Florianópolis/SC
 Date of diagnosis: 04/03/2024
 Date of surgery: 09/12/2024
 Date of inclusion/randomization: 10/28/2024
 ECOG Performance Status: 0

**2. Present Illness History (PIH)**
Patient presented with right breast discomfort and palpable mass noted in late March 2024.
Breast ultrasound and mammography (04/01/2024) revealed a 3.2 cm irregular mass in the upper outer quadrant (UOQ) of the right breast and a 1.6 cm suspicious axillary lymph node.
Core biopsy on 04/04/2024: invasive ductal carcinoma, histologic grade 3 (Nottingham).
Immunohistochemistry: ER negative, PR negative, HER2 3+, Ki-67 46%
Clinical staging: cT2N1M0 — Stage IIB (AJCC 8th edition)

**3. Personal and Family History**
 Comorbidities: Hypothyroidism (on levothyroxine), dyslipidemia
 Drug allergies: None
 Gynecologic history: Menarche at 12; menopause not yet reached; G2P2A0; used oral contraceptives until age 45
 Family history: Paternal aunt with breast cancer at age 62
 Previous radiotherapy: None
 Lifestyle: Non-smoker; drinks wine occasionally

**4. Physical Examination (pre-treatment)**
 General condition: Excellent; asymptomatic
 Right breast: 3.2 cm firm mass in UOQ; no skin changes or nipple discharge
 Left breast: No abnormalities
 Lymph nodes: One firm, mobile right axillary lymph node (~1.5 cm)
 No inflammatory signs, edema, or pain

**5. Additional Tests**
 CBC (April 2024): Hb 12.7 g/dL; WBC 6,400/mm³; platelets 238,000/mm³
 Renal/liver function: Creatinine 0.81 mg/dL; urea 27 mg/dL; AST 22 U/L; ALT 20 U/L
 β-HCG: Negative
 Serologies: HBsAg, anti-HCV, HIV — all negative
 LVEF pre-chemotherapy (ECHO 04/2024): 64%
 LVEF post-chemotherapy (ECHO 09/2024): 55% (drop of 9 p.p.)
 HER2: IHC 3+, confirmed by FISH
 Imaging: Chest, abdomen, pelvis CT + bone scan — no signs of metastasis
 Meets all baseline inclusion criteria

**6. Neoadjuvant Treatment**
 Regimen: TCHP (docetaxel 75 mg/m² + carboplatin AUC6 + trastuzumab + pertuzumab)
 Start date: 04/17/2024 — End date: 08/06/2024
 Total duration: 16+ weeks
 Trastuzumab and taxane: Administered ≥ 9 weeks
 Clinical response: Tumor size reduction of approximately 50%
 Adverse events:

- Grade 1 diarrhea
- Grade 2 alopecia
- No neutropenia or cardiotoxicity
- Interval between last chemo and surgery: 5 weeks

**7. Surgical and Pathological Data**
 Surgery: Right breast-conserving surgery with sentinel node biopsy and axillary dissection
 Incision: Lateral curved
 Laterality: Right
 Lymph nodes: 13 removed, 2 positive
 Residual tumor size: 1.0 cm
 Margins: Negative (>3 mm)
 Histologic grade post-treatment: Grade 2
 Pathological response: Partial
 Lymphovascular invasion: Present
 Perineural invasion: Absent
 Final pathological staging: ypT1bN1aM0

**PATIENT 25 — SYNTHETIC EXCLUSION RECORD**

**1. Identification**
 Synthetic ID: SPT-025
 Age: 42 years and 7 months
 Sex / Ethnicity: Female, Indigenous
 Weight: 63 kg — Height: 1.61 m — BMI: 24.3 kg/m²
 Marital status: Single
 Profession: Social worker
 Healthcare center: Centro Amazônico de Oncologia e Pesquisa — Manaus/AM
 Date of diagnosis: 03/13/2024
 Date of surgery: 08/27/2024
 Date of inclusion/randomization: 10/15/2024
 ECOG Performance Status: 0

**2. Present Illness History (PIH)**
Patient presented after noticing a lump in her left breast during showering.
Ultrasound and mammogram (03/11/2024): 3.3 cm irregular hypoechoic mass in the lower outer quadrant (LOQ) of the left breast and one suspicious left axillary node (~1.6 cm).
Core biopsy performed on 03/14/2024: invasive ductal carcinoma, histologic grade 3 (Nottingham).
Immunohistochemistry: ER negative, PR negative, HER2 3+, Ki-67 49%
Clinical staging: cT2N1M0 — Stage IIB (AJCC 8th edition)

**3. Personal and Family History**
 Comorbidities: Mild asthma (uses inhaler as needed)
 Drug allergies: None reported
 Gynecologic history: Menarche at 12; regular cycles; G2P2A0; no hormone use
 Family history: Maternal aunt with breast cancer at age 61
 Previous radiotherapy: None
 Lifestyle: Non-smoker; does not drink alcohol

**4. Physical Examination (pre-treatment)**
 General condition: Good; no weight loss, fever, or fatigue
 Left breast: 3.3 cm firm, irregular mass in LOQ; no skin dimpling or nipple changes
 Right breast: Normal
 Lymph nodes: One left axillary node (~1.6 cm), firm and mobile
 No signs of inflammation or lymphedema

**5. Additional Tests**
 CBC (March 2024): Hb 12.8 g/dL; WBC 6,000/mm³; platelets 236,000/mm³
 Renal/liver function: Creatinine 0.80 mg/dL; urea 26 mg/dL; AST 23 U/L; ALT 21 U/L
 β-HCG: Negative
 Serologies: HBsAg, anti-HCV, HIV — all negative
 LVEF pre-chemotherapy (ECHO 03/2024): 65%
 LVEF post-chemotherapy (ECHO 08/2024): 58% (drop of 7 p.p.)
 HER2: IHC 3+, confirmed with FISH
 Imaging: Chest/abdomen/pelvis CT + bone scintigraphy — no evidence of metastases
 All inclusion lab and imaging parameters met

**6. Neoadjuvant Treatment**
 Regimen: TCHP (docetaxel 75 mg/m² + carboplatin AUC6 + trastuzumab + pertuzumab)
 Start date: 03/27/2024 — End date: 07/17/2024
 Total duration: 16+ weeks
 Trastuzumab and taxane: ≥ 9 weeks
 Clinical response: Tumor and nodal reduction ~50%
 Adverse events:

- Grade 2 alopecia
- Grade 1 fatigue
- No hematologic, hepatic, or cardiac complications
- Interval between last dose and surgery: 6 weeks

**7. Surgical and Pathological Data**
 Surgery: Left segmental mastectomy + axillary lymph node dissection
 Incision: Curved lateral
 Laterality: Left
 Lymph nodes: 15 removed, 2 positive
 Residual tumor size: 1.1 cm
 Margins: Negative (>4 mm)
 Histologic grade after neoadjuvant: Grade 2
 Pathological response: Partial
 Lymphovascular invasion: Present
 Perineural invasion: Absent
 Final pathological staging: ypT1cN1aM0

**FINAL PROMPT FOR PATIENT SENTENCE GENERATION (P2S)**

**CONTEXT:** You are a specialist in Natural Language Processing (NLP) and information extraction, focusing on oncology data.

**GOAL:** Process an electronic medical record to generate a "patient sentence" (P2S). The goal of this sentence is to create a concise, structured clinical “fingerprint” that accelerates initial screening for clinical trials. The sentence should be a factual and chronological representation of the patient’s positive clinical data.

**INPUT:** A patient’s electronic medical record file.

**OUTPUT FORMAT:** A single text string, with each clinical detail separated by the character |.

**EXTRACTION AND FORMATTING RULES:**

**Mandatory Start:** The sentence MUST begin with the location of the primary tumor.

**Chronological Order:** Events must be listed from oldest to most recent.

**Date Format:** Each event or report MUST include its date in the format (DD/MM/YYYY).

**"Key: Value" Structure:** Use a Key: Value format for all information.

**Variable Sentence Length, Focus on Positive Data:** The sentence will vary in length. Include only positive findings and data. Do NOT include negative results (e.g., negative genetic tests), minor comorbidities, or incidental findings.

**Maximum Detail for Reports:**

- **Pathology:** Extract all details: histological type, grade, tumor size (Tu), margin status, angiolymphatic and perineural invasion.
- **Immunohistochemistry (IHC):** Extract exact percentages for ER, PR, and Ki-67, and HER2 score.
- **Genetics:** Transcribe the full results of genetic tests (e.g., BRCA1/2, MLPA).

**Handling Dynamic Data:**

- **Clinical Data (e.g., ECOG status):** Include ONLY the most recent record.
- **Tumor Markers (e.g., CEA, CA 15-3):** Include the baseline AND most recent value. Use the format: “Marker: Initial_Value (DD/MM/YYYY) -> Recent_Value (DD/MM/YYYY)”.

**Essential Information List (Extract if present and positive):**

- **Diagnosis:** Histological type, grade, size, invasions (angiolymphatic/perineural), etc. (with date).
- **Pathological Staging:** pTNM (with date).
- **IHC:** ER, PR, HER2, Ki-67 (with date).
- **Genetic Tests:** Test result (with date).
- **Surgery(ies):** Procedure type, margin status (with date).
- **Systemic Treatments (CT, HT, etc.):** Protocol/medication and period (start and end).
- **Radiotherapy (RT):** Period (start and end).
- **Performance Status:** Latest ECOG (with date).
- **Tumor Markers:** First and last value (with dates).
- **Major Comorbidities:** Only those listed as significant prior conditions (e.g., Hypertension, Diabetes).

**FINAL OUTPUT EXAMPLE:**

*Right Breast | Diagnosis: Invasive ductal carcinoma NST, Histologic grade II, Size: 2.0 cm, Margins: Clear, Angiolymphatic/peritumoral embolization: Not detected (26/08/2019) | Pathological Staging: pT1c pN1 pMx (26/08/2019) | IHC: ER positive 100%, PR positive 100%, HER2 score 0, Ki67 positive 10% (18/09/2019) | Adjuvant CT: TC protocol (Start: NOV/2019 - End: 16/01/2020) | Adjuvant RT: (Start: 17/02/2020 - End: 10/03/2020) | Adjuvant HT: Anastrozole (Start: MAR/2020) | Bone Therapy: Zoledronic Acid every 6 months (Start: MAR/2020) | Comorbidities: Systemic arterial hypertension | Tumor Marker CA15-3: 15.7 U/ml (31/10/2019) -> 16.7 U/ml (05/01/2024) | Tumor Marker CEA: 1.6 ng/ml (31/10/2019) | ECOG: 0 (22/04/2024)*

**SYNTHESIS OF SYNTHETIC PATIENTS - KATHERINE**

**PATIENT 1 — SYNTHETIC INCLUSION RECORD**

Right Breast | Diagnosis: Invasive ductal carcinoma, Grade 3 (Nottingham), Size: 2.8 cm, Enlarged axillary lymph node (1.5 cm), Clinical staging: cT2N1M0, Stage IIB (AJCC 8th edition) (21/06/2023) | IHC: ER 0%, PR 0%, HER2 3+ (FISH positive), Ki-67 45% (21/06/2023) | Neoadjuvant CT: TCHP regimen (docetaxel 75 mg/m² + carboplatin AUC6 + trastuzumab + pertuzumab) (Start: 10/07/2023 – End: 20/11/2023) | Clinical response: Partial, 40% tumor reduction (20/11/2023) | Surgery: Right quadrantectomy + level I-II axillary lymphadenectomy (08/03/2024) | Pathology: Residual tumor 0.9 cm, Histologic grade II, 16 lymph nodes resected (2 positive), Clear margins (minimum 3 mm), Lymphovascular invasion present, Perineural invasion absent, Partial pathological response (08/03/2024) | Final staging: ypT1bN1aM0 (08/03/2024) | ECOG: 0 (22/04/2024) | Comorbidities: Controlled systemic arterial hypertension.

**PATIENT 2 — SYNTHETIC INCLUSION RECORD**

Left Breast | Diagnosis: Invasive ductal carcinoma, Grade 3 (Nottingham), Size: 4.5 cm, Suspicious axillary lymph node (1.8 cm), Clinical staging: cT3N1M0, Stage IIIA (AJCC 8th edition) (12/04/2023) | IHC: ER 10%, PR 0%, HER2 3+, Ki-67 55% (12/04/2023) | Neoadjuvant CT: AC-TH regimen — Phase 1: Doxorubicin 60 mg/m² + Cyclophosphamide 600 mg/m² (4 cycles) | Phase 2: Docetaxel 75 mg/m² + Trastuzumab (8→6 mg/kg q3w) (4 cycles) (Start: 15/05/2023 – End: 30/09/2023) | Clinical response: Partial, 60% tumor volume reduction (30/09/2023) | Surgery: Left modified radical mastectomy + complete axillary lymphadenectomy (19/02/2024) | Pathology: Residual tumor 1.5 cm, Histologic grade II, 21 lymph nodes resected (3 positive), Clear margins (minimum 4 mm), Lymphovascular invasion present, Perineural invasion absent, Partial pathological response (19/02/2024) | Final staging: ypT1cN1aM0 (19/02/2024) | ECOG: 1 (05/04/2024) | Comorbidities: Treated dyslipidemia.

**PATIENT 3 — SYNTHETIC INCLUSION RECORD**

Right Breast | Diagnosis: Invasive ductal carcinoma, Grade 2 (Nottingham), Size: 2.6 cm, Enlarged axillary lymph node (1.2 cm), Clinical staging: cT2N1M0, Stage IIB (AJCC 8th edition) (15/05/2023) | IHC: ER 0%, PR 0%, HER2 3+ (FISH amplified), Ki-67 35% (15/05/2023) | Neoadjuvant CT: TCHP regimen — docetaxel 75 mg/m² + carboplatin AUC6 + trastuzumab + pertuzumab (Start: 20/06/2023 – End: 30/10/2023) | Clinical response: Partial, ~50% tumor volume reduction (30/10/2023) | Surgery: Right quadrantectomy with sentinel lymph node biopsy (12/02/2024) | Pathology: Residual tumor 0.6 cm, Histologic grade II, 3 lymph nodes resected (1 positive with micrometastasis), Clear margins (minimum 2 mm), No lymphovascular invasion, No perineural invasion, Partial pathological response (12/02/2024) | Final staging: ypT1bN1miM0 (12/02/2024) | ECOG: 0 (15/03/2024) | Comorbidities: None relevant.

**PATIENT 4 — SYNTHETIC INCLUSION RECORD**

Left Breast | Diagnosis: Invasive ductal carcinoma, Grade 3 (Nottingham), Size: 5.2 cm, Enlarged axillary lymph nodes (up to 2.1 cm), Clinical staging: cT3N2M0, Stage IIIB (AJCC 8th edition) (02/02/2023) | IHC: ER 0%, PR 0%, HER2 amplified (FISH positive), Ki-67 60% (02/02/2023) | Neoadjuvant CT: AC → THP regimen — Phase 1: Doxorubicin 60 mg/m² + Cyclophosphamide 600 mg/m² (4 cycles) | Phase 2: Docetaxel 75 mg/m² + Trastuzumab (8→6 mg/kg q3w) + Pertuzumab (840→420 mg q3w) (4 cycles) (Start: 13/03/2023 – End: 21/08/2023) | Clinical response: Partial, ~50% tumor diameter reduction (21/08/2023) | Surgery: Left modified radical mastectomy + complete axillary lymphadenectomy (18/12/2023) | Pathology: Residual tumor 2.1 cm, Histologic grade III, 23 lymph nodes resected (4 positive), Clear margins (minimum 5 mm), Lymphovascular invasion present, Perineural invasion absent, Partial pathological response (18/12/2023) | Final staging: ypT2N2aM0 (18/12/2023) | ECOG: 1 (05/02/2024) | Comorbidities: Controlled hypertension and dyslipidemia.

**PATIENT 5 — SYNTHETIC INCLUSION RECORD**

Right Breast | Diagnosis: Invasive ductal carcinoma, Grade 2 (Nottingham), Size: 2.4 cm, Clinical staging: cT2N0M0, Stage IIA (AJCC 8th edition) (12/09/2023) | IHC: ER 90%, PR 60%, HER2 3+, Ki-67 30% (12/09/2023) | Neoadjuvant CT: TCHP regimen — docetaxel 75 mg/m² + carboplatin AUC6 + trastuzumab + pertuzumab (Start: 10/10/2023 – End: 05/02/2024) | Clinical response: Partial, 70% tumor volume reduction (05/02/2024) | Surgery: Right quadrantectomy + sentinel lymph node biopsy (22/04/2024) | Pathology: Residual tumor 0.4 cm, Histologic grade II, 4 lymph nodes resected (0 positive), Clear margins (minimum 3 mm), No lymphovascular invasion, No perineural invasion, Partial pathological response with minimal residual invasive disease (22/04/2024) | Final staging: ypT1aN0M0 (22/04/2024) | ECOG: 0 (20/06/2024) | Comorbidities: None relevant.

### **PATIENT 6 — SYNTHETIC EXCLUSION RECORD**

Left Breast | Diagnosis: Invasive ductal carcinoma, Grade 3 (Nottingham), Size: 5.6 cm, Multiple axillary lymph nodes (up to 2.5 cm), Confirmed lung metastasis on CT (03/2023), Clinical staging: cT4bN2M1, Stage IV (AJCC 8th edition) (20/01/2023) | IHC: ER 0%, PR 0%, HER2 3+, Ki-67 55% (20/01/2023) | Palliative CT: Docetaxel + Trastuzumab (Start: 02/2023 – End: 05/2023, 4 cycles, 12 weeks duration) | Response: Stable (05/2023) | Surgery: Palliative tumor debridement (local ulcer) without complete resection (28/09/2023) | Pathology: Invasive ductal carcinoma HER2 3+, Margins not assessable, Pathological staging: pT4bN2M1 (28/09/2023) | ECOG: 2 (12/05/2023) | Comorbidities: Controlled hypertension and heart failure.

### **PATIENT 7 — SYNTHETIC EXCLUSION RECORD**

Right Breast | Diagnosis: Invasive ductal carcinoma, Grade 3 (Nottingham), Size: 3.2 cm, Enlarged axillary lymph node, Clinical staging: cT2N1M0, Stage IIB (AJCC 8th edition) (22/02/2023) | IHC: ER 0%, PR 0%, HER2 3+, Ki-67 50% (22/02/2023) | Neoadjuvant CT: TCHP regimen — docetaxel + carboplatin + trastuzumab + pertuzumab (Start: 13/03/2023 – End: 24/07/2023, 6 cycles, 17 weeks) | Clinical response: Partial (24/07/2023) | Surgery: Right quadrantectomy + axillary lymphadenectomy (01/05/2024) | Pathology: Residual tumor 1.1 cm, Histologic grade II, 14 lymph nodes resected (2 positive), Clear margins (minimum 3 mm), Lymphovascular invasion present, Perineural invasion absent (01/05/2024) | Final staging: ypT1cN1aM0 (01/05/2024) | ECOG: 0 (03/05/2024) | Comorbidities: Controlled type 2 diabetes mellitus.

### **PATIENT 8 — SYNTHETIC EXCLUSION RECORD**

Left Breast | Diagnosis: Invasive ductal carcinoma, Grade 3 (Nottingham), Size: 3.5 cm, Enlarged axillary lymph node (1.6 cm), Clinical staging: cT2N1M0, Stage IIB (AJCC 8th edition) (18/04/2023) | IHC: ER 0%, PR 0%, HER2 3+, Ki-67 50% (18/04/2023) | Neoadjuvant CT: TCHP regimen — docetaxel 75 mg/m² + carboplatin AUC6 + trastuzumab + pertuzumab (Start: 22/05/2023 – End: 10/10/2023) | Clinical response: Partial, 60% tumor diameter reduction (10/10/2023) | Surgery: Left quadrantectomy + sentinel lymph node biopsy (02/09/2024) | Pathology: Residual tumor 0.8 cm, Histologic grade II, 6 lymph nodes resected (1 positive with micrometastasis), Clear margins (minimum 2 mm), Lymphovascular invasion present, Partial pathological response (02/09/2024) | Final staging: ypT1bN1miM0 (02/09/2024) | ECOG: 1 (22/03/2024) | Comorbidities: Controlled hypothyroidism.

### **PATIENT 9 — SYNTHETIC EXCLUSION RECORD**

Right Breast | Diagnosis: Invasive ductal carcinoma, Grade 3 (Nottingham), Initial size: 3.8 cm, Enlarged axillary lymph node (1.8 cm), Clinical staging: cT2N1M0, Stage IIB (AJCC 8th edition) (14/03/2023) | IHC: ER 0%, PR 0%, HER2 3+, Ki-67 60% (14/03/2023) | Neoadjuvant CT: TCHP regimen — docetaxel 75 mg/m² + carboplatin AUC6 + trastuzumab + pertuzumab (Start: 25/04/2023 – Interrupted: 25/08/2023, 4 cycles) | Reason for interruption: Tumor progression — increased to 5.2 cm and appearance of fixed axillary lymph node (22/08/2023) | Salvage regimen: Paclitaxel + Trastuzumab (09/2023) with no satisfactory response | Progression biopsy: Invasive ductal carcinoma HER2 3+, no molecular change (09/2023) | Surgery: Not performed due to progression during systemic treatment | ECOG: 1 (15/12/2023) | Comorbidities: Controlled hypertension.

### **PATIENT 10 — SYNTHETIC EXCLUSION RECORD**

Left Breast | Diagnosis: Invasive ductal carcinoma, Grade 2 (Nottingham), Size: 3.4 cm, Enlarged axillary lymph node (1.5 cm), Clinical staging: cT2N1M0, Stage IIB (AJCC 8th edition) (03/06/2023) | IHC: ER 20%, PR 0%, HER2 3+, Ki-67 40% (03/06/2023) | Neoadjuvant CT: TCHP regimen — docetaxel 75 mg/m² + carboplatin AUC6 + trastuzumab + pertuzumab (Start: 25/04/2023 – End: 12/09/2023, 6 cycles, 20 weeks duration) | Therapeutic adjustment: Trastuzumab suspended after 5th cycle due to reduced LVEF <50% (09/12/2023) | Clinical response: Partial, 50% tumor volume reduction (09/12/2023) | Surgery: Left quadrantectomy + axillary lymphadenectomy (15/12/2023) | Pathology: Residual tumor 1.2 cm, Histologic grade II, 15 lymph nodes resected (2 positive), Clear margins (minimum 4 mm), Lymphovascular invasion present, Perineural invasion absent (15/12/2023) | Final staging: ypT1cN1aM0 (15/12/2023) | ECOG: 1 (02/10/2024) | Comorbidities: Systemic arterial hypertension and controlled mild heart failure.

### **PATIENT 11 — SYNTHETIC EXCLUSION RECORD**

Right Breast | Diagnosis: Invasive ductal carcinoma, Grade 3 (Nottingham), Size: 2.9 cm, Enlarged axillary lymph node (1.3 cm), Clinical staging: cT2N1M0, Stage IIB (AJCC 8th edition) (15/05/2023) | IHC: ER 0%, PR 0%, HER2 3+, Ki-67 55% (15/05/2023) | Neoadjuvant CT: TCHP regimen — docetaxel 75 mg/m² + carboplatin AUC6 + trastuzumab + pertuzumab (Start: 12/06/2023 – End: 30/10/2023, 6 cycles, 17 weeks) | Clinical response: Partial, tumor reduction of approximately 40% (30/10/2023) | Surgery: Right quadrantectomy + axillary lymphadenectomy (22/01/2024) | Pathology: Residual tumor 1.0 cm, Histologic grade II, 12 lymph nodes resected (2 positive), Clear margins (minimum 3 mm), Lymphovascular invasion present (22/01/2024) | Final staging: ypT1bN1aM0 (22/01/2024) | ECOG: 0 (15/03/2024) | Comorbidities: Active chronic hepatitis C with altered transaminases.

### **PATIENT 12 — SYNTHETIC EXCLUSION RECORD**

Left Breast | Diagnosis: Invasive ductal carcinoma, Grade 3 (Nottingham), Size: 3.0 cm, Enlarged axillary lymph node (1.4 cm), Clinical staging: cT2N1M0, Stage IIB (AJCC 8th edition) (02/06/2023) | IHC: ER 0%, PR 0%, HER2 3+, Ki-67 45% (02/06/2023) | Neoadjuvant CT: TCHP regimen — docetaxel 75 mg/m² + carboplatin AUC6 + trastuzumab + pertuzumab (Start: 20/06/2023 – End: 11/05/2023, 6 cycles, 18 weeks) | Clinical response: Partial, 50% tumor volume reduction (11/05/2023) | Surgery: Left quadrantectomy + axillary lymphadenectomy (28/02/2024) | Pathology: Residual tumor 0.9 cm, Histologic grade II, 10 lymph nodes resected (2 positive), Clear margins (minimum 3 mm), Lymphovascular invasion present (28/02/2024) | Final staging: ypT1bN1aM0 (28/02/2024) | ECOG: 0 (25/03/2024) | Comorbidities: None.

### **PATIENT 13 — SYNTHETIC EXCLUSION RECORD**

Right Breast | Diagnosis: Invasive ductal carcinoma, Grade 3 (Nottingham), Size: 2.6 cm, Enlarged axillary lymph node (1.2 cm), Clinical staging: cT2N1M0, Stage IIB (AJCC 8th edition) (12/08/2023) | IHC: ER 0%, PR 0%, HER2 3+, Ki-67 45% (08/12/2023) | Systemic treatment: Not initiated due to confirmed pregnancy in 05/2024 (7 weeks) | Management: Neoadjuvant chemotherapy suspended and multidisciplinary monitoring with oncologic obstetrics | Surgery: Planned for post-partum (20/04/2024) | ECOG: 0 (05/05/2024) | Comorbidities: None.

### **PATIENT 14 — SYNTHETIC EXCLUSION RECORD**

Left Breast | Diagnosis: Invasive ductal carcinoma, Grade 3 (Nottingham), Size: 3.6 cm, Enlarged axillary lymph node (1.8 cm), Clinical staging: cT2N1M0, Stage IIB (AJCC 8th edition) (07/04/2023) | IHC: ER 0%, PR 0%, HER2 3+, Ki-67 50% (07/04/2023) | Neoadjuvant CT: TCHP regimen — docetaxel 75 mg/m² + carboplatin AUC6 + trastuzumab + pertuzumab (Start: 08/01/2023 – End: 12/10/2023, 6 cycles, 18 weeks) | Clinical response: Partial, 45% tumor volume reduction (12/10/2023) | Surgery: Left quadrantectomy + axillary lymphadenectomy (15/02/2024) | Pathology: Residual tumor 1.3 cm, Histologic grade II, 13 lymph nodes resected (2 positive), Clear margins (minimum 3 mm), Lymphovascular invasion present (15/02/2024) | Final staging: ypT1cN1aM0 (15/02/2024) | ECOG: 1 (04/10/2024) | Comorbidities: HIV-1 infection with detectable viral load and reduced CD4 count.

### **PATIENT 15 — SYNTHETIC EXCLUSION RECORD**

Left Breast | Diagnosis: Invasive ductal carcinoma, Grade 3 (Nottingham), Size: 3.2 cm, Enlarged axillary lymph node (1.8 cm), Clinical staging: cT2N1M0, Stage IIB (AJCC 8th edition) (08/08/2023) | IHC: ER 0%, PR 0%, HER2 3+, Ki-67 55% (08/08/2023) | Neoadjuvant CT: TCHP regimen — docetaxel + carboplatin + trastuzumab + pertuzumab (Start: 09/10/2023 – End: 22/01/2024, 6 cycles, 18 weeks) | Clinical response: Partial, 60% tumor diameter reduction (22/01/2024) | Surgery: Left modified radical mastectomy + complete axillary lymphadenectomy (20/03/2024) | Pathology: Residual tumor 1.1 cm, Histologic grade III, 17 lymph nodes resected (2 positive), Clear margins, Lymphovascular invasion present (20/03/2024) | Final staging: ypT1cN1aM0 (20/03/2024) | ECOG: 1 (05/05/2024) | Comorbidities: Systemic arterial hypertension, dyslipidemia, and history of prior radiotherapy to the contralateral breast.

### **PATIENT 16 — SYNTHETIC EXCLUSION RECORD**

Right Breast | Diagnosis: Invasive ductal carcinoma, Grade 3 (Nottingham), Size: 3.8 cm, Enlarged axillary lymph node (1.7 cm), Clinical staging: cT2N1M0, Stage IIB (AJCC 8th edition) (22/03/2023) | IHC: ER 0%, PR 0%, HER2 3+, Ki-67 50% (22/03/2023) | Neoadjuvant CT: TCHP regimen — docetaxel + carboplatin + trastuzumab + pertuzumab (Start: 15/04/2023 – End: 25/09/2023, 6 cycles) | Clinical response: Partial, 50% tumor volume reduction (25/09/2023) | Adverse events: Grade 2 mucositis, Grade 1 fatigue, exacerbation of SLE (SLEDAI-2K 14) requiring increased prednisone dose (September/2023) | Surgery: Right quadrantectomy + axillary lymphadenectomy (12/10/2023) | Pathology: Residual tumor 1.0 cm, Histologic grade II, 12 lymph nodes resected (1 positive), Clear margins (minimum 3 mm), Lymphovascular invasion present (12/10/2023) | Final staging: ypT1bN1aM0 (12/10/2023) | ECOG: 1 (02/02/2024) | Comorbidities: Systemic lupus erythematosus (moderate activity) and systemic arterial hypertension.

### **PATIENT 17 — SYNTHETIC EXCLUSION RECORD**

Left Breast | Diagnosis: Invasive ductal carcinoma, Grade 3 (Nottingham), Size: 3.4 cm, Enlarged axillary lymph node (1.6 cm), Clinical staging: cT2N1M0, Stage IIB (AJCC 8th edition) (18/04/2023) | IHC: ER 0%, PR 0%, HER2 3+, Ki-67 50% (18/04/2023) | Neoadjuvant CT: Modified TCHP regimen — docetaxel 60 mg/m² + carboplatin AUC4 + trastuzumab + pertuzumab (Start: 05/10/2023 – End: 25/09/2023, 6 cycles) | Clinical response: Partial, 50% tumor volume reduction (25/09/2023) | Adverse events: Grade 2 fatigue, Grade 1 nausea, transient creatinine elevation (2.5 mg/dL) after 4th cycle requiring dose adjustment and hydration (September/2023) | Surgery: Left quadrantectomy + axillary lymphadenectomy (12/12/2023) | Pathology: Residual tumor 1.4 cm, Histologic grade II, 14 lymph nodes resected (2 positive), Clear margins (minimum 4 mm), Lymphovascular invasion present (12/12/2023) | Final staging: ypT1cN1aM0 (12/12/2023) | ECOG: 1 (22/01/2024) | Comorbidities: Stage 4 chronic kidney disease, systemic arterial hypertension, and type 2 diabetes mellitus.

### **PATIENT 18 — SYNTHETIC EXCLUSION RECORD**

Right Breast | Diagnosis: Invasive ductal carcinoma, Grade 3 (Nottingham), Size: 2.8 cm, Enlarged axillary lymph node (1.3 cm), Clinical staging: cT2N1M0, Stage IIB (AJCC 8th edition) (14/09/2023) | IHC: ER 0%, PR 0%, HER2 3+, Ki-67 55% (14/09/2023) | Neoadjuvant CT: TCHP regimen — docetaxel 75 mg/m² + carboplatin AUC6 + trastuzumab + pertuzumab (Start: 10/03/2023 – End: 25/02/2024, 6 cycles, 18 weeks) | Clinical response: Partial, 65% tumor volume reduction (25/02/2024) | Surgery: Right quadrantectomy + axillary lymphadenectomy (18/03/2024) | Pathology: Residual tumor 0.7 cm, Histologic grade II, 11 lymph nodes resected (1 positive), Clear margins (minimum 3 mm) (18/03/2024) | Final staging: ypT1bN1aM0 (18/03/2024) | ECOG: 1 (04/10/2024) | Comorbidities: Recurrent major depressive disorder with recent suicidal ideation and psychiatric hospitalization (February/2024); on sertraline and quetiapine, under continuous psychiatric care.

### **PATIENT 19 — SYNTHETIC EXCLUSION RECORD**

Left Breast | Diagnosis: Invasive ductal carcinoma, Grade 3 (Nottingham), Size: 3.0 cm, Enlarged axillary lymph node (1.4 cm), Clinical staging: cT2N1M0, Stage IIB (AJCC 8th edition) (07/10/2023) | IHC: ER 0%, PR 0%, HER2 3+, Ki-67 55% (07/10/2023) | Neoadjuvant CT: TCHP regimen — docetaxel + carboplatin + trastuzumab + pertuzumab (Start: 08/05/2023 – End: 22/12/2023, 6 cycles, 18 weeks) | Clinical response: Partial, 60% tumor diameter reduction (22/12/2023) | Surgery: Left quadrantectomy + axillary lymphadenectomy (22/02/2024) | Pathology: Residual tumor 1.0 cm, Histologic grade II, 14 lymph nodes resected (1 positive), Clear margins (minimum 4 mm) (22/02/2024) | Final staging: ypT1bN1aM0 (22/02/2024) | ECOG: 1 (25/03/2024) | Comorbidities: Controlled systemic arterial hypertension and mild asthma.

### **PATIENT 20 — SYNTHETIC EXCLUSION RECORD**

Right Breast | Diagnosis: Invasive ductal carcinoma, Grade 3 (Nottingham), Size: 3.4 cm, Enlarged axillary lymph node (1.6 cm), Clinical staging: cT2N1M0, Stage IIB (AJCC 8th edition) (18/09/2023) | IHC: ER 0%, PR 0%, HER2 3+, Ki-67 60% (18/09/2023) | Neoadjuvant CT: TCHP regimen — docetaxel + carboplatin + trastuzumab + pertuzumab (Start: 10/02/2023 – End: 02/12/2024, 6 cycles, 18 weeks) | Clinical response: Partial, 55% tumor volume reduction (02/12/2024) | Surgery: Right quadrantectomy + axillary lymphadenectomy (20/03/2024) | Pathology: Residual tumor 1.0 cm, Histologic grade II, 13 lymph nodes resected (1 positive), Clear margins (minimum 3 mm), Lymphovascular invasion present (20/03/2024) | Final staging: ypT1bN1aM0 (20/03/2024) | ECOG: 1 (15/04/2024) | Comorbidities: Controlled systemic arterial hypertension and dyslipidemia.
 **Associated oncologic diagnosis:** Well-differentiated endometrioid carcinoma (G1) of the uterine corpus, stage IB, treated concurrently with gynecologic oncology team.

### **PATIENT 21 — SYNTHETIC EXCLUSION RECORD**

Left Breast | Diagnosis: Invasive ductal carcinoma, Grade 3 (Nottingham), Size: 3.1 cm, Enlarged axillary lymph node (1.5 cm), Clinical staging: cT2N1M0, Stage IIB (AJCC 8th edition) (20/06/2023) | IHC: ER 0%, PR 0%, HER2 3+, Ki-67 55% (20/06/2023) | Neoadjuvant CT: TCHP regimen — docetaxel + carboplatin + trastuzumab + pertuzumab (Start: 07/05/2023 – End: 15/11/2023, 6 cycles, 18 weeks) | Clinical response: Partial, approximately 60% tumor volume reduction (15/11/2023) | Adverse events: Grade 1 nausea, Grade 1 diarrhea, and asymptomatic drop in left ventricular ejection fraction (LVEF 65 → 48%), leading to early suspension of anti-HER2 drugs and referral to cardio-oncology (January/2024) | Surgery: Left quadrantectomy + axillary lymphadenectomy (02/05/2024) | Pathology: Residual tumor 1.1 cm, Histologic grade II, 15 lymph nodes resected (2 positive), Clear margins (minimum 4 mm) (02/05/2024) | Final staging: ypT1cN1aM0 (02/05/2024) | ECOG: 1 (18/03/2024) | Comorbidities: Controlled systemic arterial hypertension and dyslipidemia.

### **PATIENT 22 — SYNTHETIC EXCLUSION RECORD**

Right Breast | Diagnosis: Invasive ductal carcinoma, Grade 3 (Nottingham), Size: 3.5 cm, Enlarged axillary lymph node (1.8 cm), Clinical staging: cT2N1M0, Stage IIB (AJCC 8th edition) (11/04/2023) | IHC: ER 0%, PR 0%, HER2 3+, Ki-67 50% (04/11/2023) | Neoadjuvant CT: TCHP regimen — docetaxel + carboplatin + trastuzumab + pertuzumab (started 05/08/2023), with treatment abandonment after the 3rd cycle and definitive interruption (>8 weeks without continuation) | Related events: Unjustified absences and loss to follow-up; patient returned only in December/2023 requesting resumption, but re-inclusion was not possible | Surgery: Right quadrantectomy + axillary lymphadenectomy (12/12/2023) | Pathology: Residual tumor 2.8 cm, Grade III, 14 lymph nodes resected (3 positive), Clear margins (minimum 3 mm), Lymphovascular invasion present (12/12/2023) | Final staging: ypT2N1aM0 (12/12/2023) | ECOG: 1 (02/05/2024)

### **PATIENT 23 — SYNTHETIC EXCLUSION RECORD**

Axillary lymph node 1.5 cm, Clinical staging: cT2N1M0, Stage IIB (AJCC 8th edition) (05/09/2023) | IHC: ER 0%, PR 0%, HER2 3+, Ki-67 60% (05/09/2023) | Neoadjuvant CT: TCHP regimen — docetaxel + carboplatin + trastuzumab + pertuzumab (Start: 06/02/2023 – End: 18/10/2023, 6 cycles, 18 weeks) | Clinical response: Partial, approximately 55% tumor volume reduction | Adverse events: Progressive elevation of liver transaminases from the 5th cycle (AST 165 U/L; ALT 188 U/L), resulting in grade 3 hepatotoxicity (AST 296 U/L; ALT 325 U/L, ~6× ULN), leading to definitive suspension of anti-HER2 drugs and liver evaluation — ultrasound showed grade II steatosis without focal lesions | Surgery: Left quadrantectomy + axillary lymphadenectomy (01/10/2024) | Pathology: Residual tumor 1.2 cm, Grade II, 15 lymph nodes resected (2 positive), Clear margins (minimum 3 mm) (01/10/2024) | Final staging: ypT1cN1aM0 (01/10/2024) | ECOG: 1 (03/05/2024)

### **PATIENT 24 — SYNTHETIC EXCLUSION RECORD**

Left Breast | Diagnosis: Invasive ductal carcinoma, Grade 3 (Nottingham), Size: 3.2 cm, Axillary lymph node 1.4 cm, Clinical staging: cT2N1M0, Stage IIB (AJCC 8th edition) (16/08/2023) | IHC: ER 0%, PR 0%, HER2 3+, Ki-67 55% (16/08/2023) | Neoadjuvant CT: TCHP regimen — docetaxel + carboplatin + trastuzumab + pertuzumab (Start: 01/09/2023, interrupted 20/12/2023 after the 5th cycle due to acute neurological event) | Clinical response: Partial, approximately 50% tumor volume reduction | Adverse events: Ischemic stroke during the 5th cycle, affecting the right middle cerebral artery territory, with mild left hemiparesis (strength 4/5), mild dysarthria, and moderate functional limitations — ECOG worsened from 1 to 2; treatment discontinued, and motor rehabilitation started | Surgery: Left quadrantectomy + axillary lymphadenectomy (14/02/2024) | Pathology: Residual tumor 1.6 cm, Grade II, 14 lymph nodes resected (2 positive), Clear margins (14/02/2024) | Final staging: ypT1cN1aM0 (14/02/2024) | ECOG: 2 (25/03/2024)

### **PATIENT 25 — SYNTHETIC EXCLUSION RECORD**

Right Breast | Diagnosis: Invasive ductal carcinoma, Grade 3 (Nottingham), Size: 3.0 cm, Axillary lymph node 1.3 cm, Clinical staging: cT2N1M0, Stage IIB (AJCC 8th edition) (22/06/2023) | IHC: ER 0%, PR 0%, HER2 3+, Ki-67 55% (22/06/2023) | Neoadjuvant CT: TCHP regimen — docetaxel + carboplatin + trastuzumab + pertuzumab (started 07/05/2023; suspended after the 5th cycle due to severe hematologic toxicity) | Adverse events: Grade 3 neutropenia after the 2nd cycle (neutrophils 700/mm³), grade 4 febrile neutropenia during the 4th cycle (neutrophils 220/mm³, fever 39°C, CRP 95 mg/L, hospitalization for 6 days with antibiotics and filgrastim), and another episode of grade 4 febrile neutropenia in the 5th cycle, leading to definitive treatment discontinuation due to hematologic risk | Clinical response: Partial (approximately 40% tumor volume reduction) | Surgery: Right quadrantectomy + axillary lymphadenectomy (15/01/2024) | Pathology: Residual tumor 1.7 cm, Grade II, 13 lymph nodes resected (2 positive), Clear margins (minimum 3 mm) (15/01/2024) | Final staging: ypT1cN1aM0 (15/01/2024) | ECOG: 1 (25/02/2024) | **Exclusion reason:** Severe hematologic toxicity with recurrent grade 4 febrile neutropenia, requiring definitive suspension of neoadjuvant therapy.

**SCREENER - KATHERINE**

**CONTEXT:**
You are a healthcare professional specialized in clinical oncology and translational research, responsible for screening oncology patients according to the inclusion and exclusion criteria of a clinical study. Your role is to analyze clinical records with precision, ensuring compliance with the study protocol.

**OBJECTIVE (YOUR TASK):**
Thoroughly analyze the patient's medical record and determine eligibility based on the inclusion and exclusion criteria outlined in the table below:

**Inclusion Criteria:**

- Adult patient, ≥ 18 years of age
- HER2-positive breast cancer
- Histologically confirmed invasive breast carcinoma
- Clinical stage T1–T4 / N0–N3 / M0 at presentation (patients with T1a/bN0 tumors are not eligible)
- Completion of preoperative systemic chemotherapy and HER2-directed treatment consisting of at least 6 cycles of chemotherapy over a total duration of at least 16 weeks, including at least 9 weeks of trastuzumab and at least 9 weeks of taxane-based therapy
- Adequate excision: surgical removal of all clinically evident disease in the breast and lymph nodes as specified in the protocol
- Pathological evidence of residual invasive carcinoma in the breast or axillary lymph nodes after completion of preoperative therapy
- No more than 12 weeks between the date of surgery and the date of randomization
- Known hormone receptor status
- ECOG performance status of 0 or 1
- Adequate hematologic, renal, and liver function
- Screening left ventricular ejection fraction (LVEF) ≥ 50% on echocardiogram (ECHO) or MUGA scan after receiving neoadjuvant chemotherapy, and no decrease in LVEF > 15 percentage points from pre-chemotherapy baseline. If no baseline LVEF is available, then post-treatment LVEF must be ≥ 55%
- For women who are not postmenopausal or surgically sterile: agreement to remain abstinent or use single or combined contraceptive methods with a failure rate < 1% per year during treatment and for at least 7 months after the last study drug dose
- Documentation of hepatitis B and hepatitis C virus serology is required

**Exclusion Criteria:**

- Stage IV (metastatic) breast cancer
- History of any prior (ipsilateral or contralateral) breast cancer except lobular carcinoma in situ
- Evidence of gross residual or recurrent disease following preoperative therapy and surgery
- Progressive disease during preoperative systemic therapy
- Use of any investigational anti-cancer drug within 28 days prior to starting study treatment
- History of other malignancy within the past 5 years, except for adequately treated carcinoma in situ of the cervix, non-melanoma skin cancer, Stage I uterine cancer, or other non-breast cancers with a similarly favorable prognosis
- Patients for whom radiotherapy would be indicated for breast cancer treatment but is medically contraindicated
- Current NCI CTCAE (Version 4.0) Grade ≥ 2 peripheral neuropathy
- History of exposure to cumulative anthracycline doses:
  - Doxorubicin > 240 mg/m²
  - Epirubicin or Liposomal Doxorubicin-Hydrochloride (Myocet®) > 480 mg/m²
  - Other anthracyclines: exposure equivalent to doxorubicin > 240 mg/m²
- Cardiopulmonary dysfunction as defined in the protocol
- Prior treatment with trastuzumab emtansine
- Current severe, uncontrolled systemic disease
- Pregnant or lactating women
- Known active liver disease (e.g., due to HBV, HCV, autoimmune hepatitis, or sclerosing cholangitis)
- Concurrent serious uncontrolled infections requiring treatment or known HIV infection
- History of intolerance (Grade 3–4 infusion reaction or hypersensitivity) to trastuzumab, murine proteins, or any component of the product

**Decision Logic:**

- **INCLUDED:** The patient meets **all** inclusion criteria and **none** of the exclusion criteria
- **EXCLUDED:** The patient **fails at least one** inclusion criterion or **meets at least one** exclusion criterion

**STYLE:**
The screening report must follow the medical-technical style of an oncological patient chart. Use standardized, objective, and formal medical language. Sentences should be short, clear, and direct, avoiding unnecessary adjectives or subjective expressions.

**TONE:**
Formal. It must reflect the language used by oncologists and clinical researchers.

**TARGET AUDIENCE:**
Healthcare professionals involved in oncology research and clinical screening processes — especially screening nurses, investigator physicians, and clinical trial coordinators.

**RESPONSE FORMAT:**
You may use the structure below:

- **Screening outcome:** INCLUDED / EXCLUDED
- **Inclusion criteria met:** [describe objectively]
- **Exclusion criteria met:** [describe objectively]
- **Decision summary:** [objectively justify the screening decision]

**PURPOSE OF THE PROMPT (SUMMARY):**
To generate formal oncology screening reports using medically appropriate language, based on investigator-defined eligibility criteria. These reports are intended for training and validating AI models applied to oncological research and clinical trials.

**SYNTHETIC PATIENT GENERATOR PROMPT - MONARCHE**

### **CONTEXT:**

You are a healthcare professional specialized in clinical oncology and translational research, responsible for generating synthetic databases of oncology medical records. These records are artificially created, without the use of any real personal data, and follow strict ethical and scientific standards. Their purpose is to support the training of artificial intelligence models focused on screening patients for oncological studies and research.

### **OBJECTIVE (YOUR TASK):**

To generate synthetic medical records with detailed clinical data, built according to the following **inclusion and exclusion criteria**:

### **Inclusion Criteria:**

- Women (regardless of menopausal status) or men ≥18 years of age (or as per local regulations)
- Participant has confirmed HR-positive, HER2-negative early-stage resected invasive breast cancer with no evidence of distant metastasis
- Participant must have undergone definitive surgery of the primary breast tumor
- Participant must have tumor tissue from the breast (preferred) or lymph node available prior to randomization for exploratory biomarker analysis
- Pathologic lymph node involvement and at least **one** of the following high-risk features:
  - ≥4 positive axillary lymph nodes
  - Tumor size ≥5 cm
  - Grade 3 (defined as ≥8 points on the Bloom-Richardson grading system)
  - Ki-67 index by central analysis ≥20% on untreated breast tissue
- Participant must be randomized within **16 months** after definitive surgery
- Participant may receive up to 12 weeks of endocrine therapy after the last non-endocrine treatment (surgery, chemotherapy, or radiation), whichever comes last
- Participant must have recovered (grade ≤1) from the acute effects of chemotherapy and radiotherapy, and from surgical side effects
- Women of childbearing potential must have a **negative serum pregnancy test** and agree to use **highly effective contraceptive methods**
- ECOG performance status ≤1
- Adequate organ function
- Participant must be able to swallow oral medications

### **Exclusion Criteria:**

- Metastatic disease (including contralateral axillary lymph nodes) or node-negative disease
- Inflammatory breast cancer
- History of previous breast cancer, except ipsilateral ductal carcinoma in situ (DCIS) treated with locoregional therapy ≥5 years ago; contralateral DCIS treated at any time may be eligible
- History of any other malignancy (except non-melanoma skin cancer or cervical carcinoma in situ), unless in complete remission with no treatment for at least 5 years prior to randomization
- Pregnant or lactating women
- Prior treatment with any CDK4 or CDK6 inhibitor
- Current use of exogenous reproductive hormone therapy (e.g., birth control pills, hormone replacement therapy, megestrol acetate)
- Prior endocrine therapy for breast cancer prevention (e.g., tamoxifen, aromatase inhibitors, raloxifene)
- Serious pre-existing medical condition(s) that, in the judgment of the investigator, would prevent participation
- Personal history of: cardiovascular syncope, pathological ventricular arrhythmia, or sudden cardiac arrest; or history of venous thromboembolism (VTE)
- Active systemic infections or elevated viral load
- Receipt of an experimental treatment in a clinical trial within the last 30 days or 5 half-lives, whichever is longer

### **STYLE:**

The text must follow a **medical-technical style**, similar to formal oncology medical records, using standardized, objective, and professional medical language. Avoid unnecessary adjectives and subjective expressions.

### **TONE:**

Formal. It should reflect the communication style of oncologists and clinical researchers.

### **TARGET AUDIENCE:**

Healthcare professionals involved in oncology research and clinical screening, especially triage nurses, investigator physicians, and clinical trial coordinators.

### **RESPONSE FORMAT:**

You may use a standard **clinical record structure** like the one below:

### **1. Identification**

- Synthetic patient ID
- Exact age (in years and months)
- Sex and ethnicity / skin color
- Weight, height, BMI
- Marital status and profession
- Site of care (hospital / cancer center)
- Date of diagnosis
- Date of surgery
- Date of inclusion/randomization in the study
- ECOG performance status

### **2. History of Present Illness (HPI)**

- Date of symptom onset and progression
- Detection method (self-exam, mammogram, ultrasound, etc.)
- Reason for referral to oncology center
- Tumor laterality (right/left)
- Complete initial clinical TNM staging (e.g., cT2N1M0, stage IIB)
- Date and type of biopsy (core, vacuum-assisted)
- Histological type and Nottingham grade
- Ki-67 index (%)
- Hormone receptors (ER, PR) and HER2 (IHC/FISH)
- Clinical stage according to AJCC (8th edition)

### **3. Personal and Family History**

- Relevant comorbidities (e.g., hypertension, diabetes, dyslipidemia)
- Drug allergies
- Gynecological history (menarche, menopause, hormonal use, G P A)
- Family history of cancer (breast, ovary, colon, prostate)
- Prior radiotherapy to the thoracic region
- Lifestyle habits (smoking, alcohol use)

### **4. Physical Examination**

- General condition and systemic signs (e.g., fatigue, weight loss)
- Bilateral breast findings
- Tumor size and location (in cm)
- Skin changes, nipple retraction, discharge
- Palpable lymph nodes (number, size, mobility)
- Presence of pain, lymphedema, or inflammatory signs

### **5. Complementary Tests**

- CBC (approximate values: Hb, WBC, platelets)
- Liver function (AST, ALT), renal function (creatinine, urea)
- Beta-HCG (negative in women of childbearing age)
- LVEF before and after chemotherapy (ECHO or MUGA, with dates and variation <15 p.p.)
- Serologies (HBsAg, anti-HCV, HIV)
- Ki-67, HER2 confirmation (FISH/CISH)
- Minimum lab criteria (Hb >10; neutrophils >1500; platelets >100,000)
- Staging imaging exams (CT chest/abdomen/pelvis, bone scan)

### **6. Neoadjuvant Treatment**

- Regimen and drugs used
- Dosages (e.g., docetaxel 75 mg/m²)
- Start and end dates
- Total duration (≥16 weeks)
- Duration of trastuzumab and taxane (≥9 weeks each)
- Clinical response observed (tumor reduction / stable / no progression)
- Adverse events (type and grade – NCI-CTCAE)
- Interval between last dose and surgery (≤12 weeks)

### **7. Surgical and Pathological Data**

- Type of surgery (partial / radical / immediate or delayed reconstruction)
- Incision type
- Laterality
- Total number of lymph nodes resected and positive
- Residual tumor size (in cm)
- Surgical margins (in mm)
- Histologic grade post-chemotherapy
- Pathological response (pCR, partial, minimal residual)
- Presence of lymphovascular / perineural invasion
- Final pathological staging (ypTNM)

### **SUMMARY – PURPOSE OF THE PROMPT:**

To generate complete, structured, and realistic **synthetic oncology patient records** in formal medical language, based on **inclusion and exclusion criteria** defined by the researcher, intended for training artificial intelligence models designed for **patient screening in oncology studies and clinical research**.

**SYNTHETIC PATIENT RECORDS - MONARCHE**

**PATIENT 1 — SYNTHETIC RECORD FOR INCLUSION**

**1. Identification**
 Synthetic ID: INC‑001
 Age: 52 years and 3 months
 Sex / Ethnicity: Female, White
 Weight / Height / BMI: 68 kg / 1.63 m / BMI 25.6 kg/m²
 Marital Status / Occupation: Married / Teacher
 Care Center: Oncology Institute of Minas Gerais – Itajubá Unit
 Date of Diagnosis: 17/05/2023
 Date of Surgery: 02/10/2023
 Date of Enrollment/Randomization: 15/01/2024
 ECOG Performance Status: 0

**2. History of Present Illness (HPI)**
 Palpable lesion in the left breast identified 4 months ago.
 Mammography and breast ultrasound (10/05/2023): irregular spiculated nodule measuring 5.2 cm in the upper outer quadrant of the left breast, with enlarged axillary lymph node (1.8 cm).
 Referred to oncology center after core biopsy (17/05/2023) confirmed invasive ductal carcinoma, grade 3 (Nottingham 8/9).
 Immunohistochemistry: ER 95%, PR 80%, HER2 0, Ki-67 35%.
 Initial clinical staging: cT3N2M0 — Stage IIIA (AJCC 8th edition).
 Detection method: self-examination followed by diagnostic mammography.
 Staging exams (CT chest/abdomen/pelvis and bone scan) showed no evidence of metastasis.

**3. Personal and Family History**
 Comorbidities: Controlled systemic arterial hypertension (on losartan 50 mg/day).
 Drug Allergies: None reported.
 Gynecologic history: menarche at 12 years, menopause at 50, G2P2A0, no use of hormone therapy.
 Family history: mother with breast cancer at age 58; no other relevant neoplasms.
 Prior chest radiotherapy: No.
 Lifestyle: Non-smoker, occasional alcohol consumption.

**4. Physical Examination (pre-neoadjuvant)**
 General status: preserved, no weight loss.
 Left breast: palpable irregular firm lesion, ~5 cm, in upper outer quadrant, partially fixed to skin, with slight nipple retraction; no discharge.
 Right breast: unremarkable.
 Axillary lymph nodes: left axilla, 3 palpable nodes (1.5–2.0 cm), mobile, painless.
 No lymphedema or inflammatory signs.

**5. Laboratory and Imaging**
 CBC: Hb 12.5 g/dL; leukocytes 6,200/mm³; platelets 280,000/mm³
 Liver function: AST 28 U/L; ALT 32 U/L
 Renal function: creatinine 0.8 mg/dL; urea 28 mg/dL
 Beta-HCG: negative
 Serologies: HBsAg negative; anti-HCV negative; HIV negative
 LVEF by echocardiogram: 66% (pre-chemo) → 63% (post-chemo); variation <15 percentage points
 Meets minimum hematologic and biochemical criteria
 Imaging: staging with CT chest/abdomen/pelvis and bone scan negative for distant disease

**6. Neoadjuvant Treatment**
 Chemotherapy regimen: doxorubicin 60 mg/m² + cyclophosphamide 600 mg/m² (AC) → paclitaxel 80 mg/m² weekly
 Start: 05/06/2023 | End: 20/09/2023
 Total duration: 16 weeks
 Trastuzumab and taxane: not indicated (HER2 negative)
 Clinical response: approximately 50% tumor reduction
 Adverse events: grade 2 neutropenia, grade 1 nausea, grade 2 alopecia
 Interval between last chemo and surgery: 12 days

**7. Surgical and Pathological Findings**
 Surgery type: Modified radical mastectomy with level I–II axillary lymphadenectomy
 Laterality: Left
 Incision: curvilinear periareolar
 Residual tumor: 1.2 cm
 Lymph nodes: 21 resected, 5 positive
 Surgical margins: free, minimum 3 mm
 Histologic grade post-chemotherapy: II
 Lymphovascular invasion: present
 Perineural invasion: absent
 Pathologic response: partial
 Final pathologic stage: ypT1cN2aM0 — Stage IIIA

**PATIENT 2 — SYNTHETIC RECORD FOR INCLUSION**

**1. Identification**
 Synthetic ID: INC‑002
 Age: 60 years and 5 months
 Sex / Ethnicity: Female, Mixed-race (Parda)
 Weight / Height / BMI: 72 kg / 1.58 m / BMI 28.8 kg/m²
 Marital Status / Occupation: Widow / Retired seamstress
 Care Center: Oncology Reference Center – São Paulo Unit
 Date of Diagnosis: 03/04/2023
 Date of Surgery: 05/09/2023
 Date of Enrollment/Randomization: 10/12/2023
 ECOG Performance Status: 1

**2. History of Present Illness (HPI)**
 Patient noticed induration in the right breast during showering.
 Mammography and breast ultrasound (28/03/2023): irregular hypoechoic lesion measuring 5.0 cm in the upper inner quadrant of the right breast, with axillary lymphadenopathy (1.6 cm).
 Referred after core biopsy (03/04/2023) confirmed invasive ductal carcinoma, grade 3 (Nottingham 8/9).
 Immunohistochemistry: ER 100%, PR 90%, HER2 negative (score 0), Ki-67 30%.
 Initial clinical staging: cT3N1M0 — Stage IIIA (AJCC 8th edition).
 Staging exams (CT chest/abdomen/pelvis, bone scan) showed no metastases.
 Detection method: self-examination.

**3. Personal and Family History**
 Comorbidities: Type 2 diabetes mellitus and systemic hypertension, both controlled.
 Allergies: No known drug allergies.
 Gynecologic history: menarche at 13, menopause at 48, G3P3A0, no hormone use.
 Family history: sister with colon cancer at age 62.
 Previous thoracic radiotherapy: No.
 Lifestyle: Non-smoker, does not consume alcohol.

**4. Physical Examination (pre-neoadjuvant)**
 General condition: preserved, no weight loss
 Right breast: palpable mass in upper inner quadrant, ~5 cm, firm, adherent to deep planes
 Left breast: no palpable alterations
 Axilla: right axillary lymph node ~1.5 cm, mobile
 No skin or nipple changes, no lymphedema

**5. Laboratory and Imaging**
 CBC: Hb 11.8 g/dL; leukocytes 5,900/mm³; platelets 275,000/mm³
 Liver function: AST 32 U/L; ALT 35 U/L
 Renal function: creatinine 0.9 mg/dL; urea 31 mg/dL
 Beta-HCG: negative
 Serologies: HBsAg negative; anti-HCV negative; HIV negative
 LVEF by ECHO: 68% (pre-CT) → 65% (post-CT), variation <15 p.p.
 Meets laboratory eligibility criteria
 Imaging: CT and bone scan without evidence of distant disease

**6. Neoadjuvant Treatment**
 Regimen: doxorubicin 60 mg/m² + cyclophosphamide 600 mg/m² (AC) followed by weekly paclitaxel 80 mg/m²
 Start: 17/04/2023 | End: 25/08/2023
 Total duration: 18 weeks
 Trastuzumab: not indicated (HER2−)
 Clinical response: tumor reduction of ~45%
 Adverse events: Grade 2 fatigue, grade 1 mucositis
 Interval between last CT and surgery: 11 days

**7. Surgical and Pathological Findings**
 Surgery: Right radical mastectomy + level I–II lymphadenectomy
 Tumor residual: 1.0 cm
 Lymph nodes: 18 removed, 3 positive
 Margins: free, minimum 5 mm
 Histological grade: II
 Lymphovascular invasion: present
 Perineural invasion: absent
 Pathological response: partial
 Final staging: ypT1bN1aM0 — Stage IIB

**PATIENT 3 — SYNTHETIC RECORD FOR INCLUSION**

**1. Identification**
 Synthetic ID: INC‑003
 Age: 49 years and 9 months
 Sex / Ethnicity: Female, White
 Weight / Height / BMI: 65 kg / 1.64 m / BMI 24.2 kg/m²
 Marital Status / Occupation: Married / Accountant
 Care Center: São Lucas Clinical Oncology Institute
 Date of Diagnosis: 16/02/2023
 Date of Surgery: 10/07/2023
 Date of Enrollment/Randomization: 20/11/2023
 ECOG Performance Status: 0

**2. History of Present Illness (HPI)**
 Routine screening mammogram detected a spiculated nodule in the left breast.
 Breast ultrasound confirmed lesion of 2.8 cm in the upper outer quadrant, with an enlarged axillary lymph node (1.4 cm).
 Core biopsy (16/02/2023): invasive ductal carcinoma, Nottingham grade 3.
 Immunohistochemistry: ER 98%, PR 90%, HER2 negative (score 0), Ki-67: 45%.
 Initial clinical stage: cT2N1M0 — Stage IIB (AJCC 8th edition).
 Staging with CT and bone scan showed no metastases.
 Detection method: screening mammography.

**3. Personal and Family History**
 Comorbidities: controlled dyslipidemia
 Allergies: none
 Gynecologic history: menarche at 12, regular cycles, G2P2A0, no hormone therapy
 Family history: maternal aunt with breast cancer (age 48)
 No prior chest radiotherapy
 Lifestyle: non-smoker, occasional alcohol use

**4. Physical Examination (pre-neoadjuvant)**
 General condition: good, no systemic complaints
 Left breast: palpable tumor in upper outer quadrant, ~3.0 cm
 Right breast: normal on palpation
 Left axilla: palpable lymph node ~1.5 cm, mobile
 No skin or nipple retraction, no lymphedema

**5. Laboratory and Imaging**
 CBC: Hb 12.3 g/dL; leukocytes 6,200/mm³; platelets 298,000/mm³
 Liver function: AST 29 U/L; ALT 34 U/L
 Renal function: creatinine 0.78 mg/dL; urea 28 mg/dL
 Beta-HCG: negative
 Serologies: HBsAg negative; anti-HCV negative; HIV negative
 LVEF (ECHO): 65% pre-CT → 62% post-CT
 Meets eligibility lab parameters
 Imaging: no distant disease

**6. Neoadjuvant Treatment**
 Chemotherapy regimen: AC-T

- Doxorubicin 60 mg/m² + cyclophosphamide 600 mg/m² (4 cycles)
- Followed by paclitaxel 80 mg/m² weekly × 12
   Start: 06/03/2023 | End: 30/06/2023
   Total duration: 17 weeks
   Trastuzumab: not indicated (HER2−)
   Clinical response: partial (~50% reduction)
   Adverse events: grade 1 neuropathy, grade 2 alopecia
   Interval between last CT and surgery: 10 days

**7. Surgical and Pathological Findings**
 Surgery: left quadrantectomy + axillary dissection
 Residual tumor: 0.9 cm
 Lymph nodes: 12 removed, 2 positive
 Margins: clear, minimum 3 mm
 Histologic grade: II
 Lymphovascular invasion: present
 Perineural invasion: absent
 Pathological response: partial
 Final staging: ypT1bN1aM0 — Stage IIB

**PATIENT 4 — SYNTHETIC RECORD FOR INCLUSION**

**1. Identification**
 Synthetic ID: INC‑004
 Age: 57 years and 2 months
 Sex / Ethnicity: Female, Brown
 Weight / Height / BMI: 72 kg / 1.60 m / BMI 28.1 kg/m²
 Marital Status / Occupation: Widowed / School Teacher
 Care Center: Regional Oncology Center of Brasília
 Date of Diagnosis: 02/01/2023
 Date of Surgery: 21/08/2023
 Date of Enrollment/Randomization: 07/11/2023
 ECOG Performance Status: 1

**2. History of Present Illness (HPI)**
 Patient presented with a palpable lump in the left breast, self-detected during bathing.
 Breast ultrasound revealed a solid hypoechoic mass of 4.8 cm with suspicious axillary lymphadenopathy (up to 2.1 cm).
 Core biopsy (02/01/2023): invasive ductal carcinoma, Nottingham grade 3.
 Immunohistochemistry: ER 5%, PR 0%, HER2 negative (IHC 1+), Ki-67: 38%.
 Initial clinical staging: cT3N2M0 — Stage IIIB (AJCC 8th edition).
 Staging CT and bone scan: no evidence of metastases.
 Detection method: self-exam

**3. Personal and Family History**
 Comorbidities: systemic arterial hypertension and controlled type 2 diabetes
 Allergies: none reported
 Gynecologic history: menarche at 11, menopause at 50, G4P4A0, no hormonal use
 Family history: mother with colon cancer at 70
 No prior radiotherapy
 Lifestyle: ex-smoker (quit 10 years ago), no alcohol use

**4. Physical Examination (Pre-Neoadjuvant)**
 General condition: good, mild fatigue
 Left breast: palpable mass in the upper inner quadrant, ~5 cm
 Right breast: no alterations
 Left axilla: multiple enlarged, mobile nodes (~2.0–2.2 cm)
 Skin: slight edema without ulceration
 No nipple retraction or lymphedema

**5. Laboratory and Imaging**
 CBC: Hb 11.8 g/dL; WBC 5,800/mm³; platelets 276,000/mm³
 Liver function: AST 32 U/L; ALT 30 U/L
 Renal function: creatinine 0.95 mg/dL; urea 35 mg/dL
 Beta-HCG: negative
 Serologies: HBsAg negative; anti-HCV negative; HIV negative
 LVEF (ECHO): 68% pre-CT → 66% post-CT (within acceptable limits)
 Meets all required laboratory parameters
 CT thorax/abdomen/pelvis and bone scintigraphy: no distant disease

**6. Neoadjuvant Treatment**
 Chemotherapy regimen: AC-T

- Doxorubicin 60 mg/m² + cyclophosphamide 600 mg/m² (4 cycles)
- Followed by paclitaxel 80 mg/m² weekly for 12 weeks
   Start: 01/03/2023 | End: 03/07/2023
   Total duration: 18 weeks
   Trastuzumab not indicated (HER2−)
   Clinical response: partial (approx. 45% tumor reduction)
   Adverse events: grade 2 fatigue, grade 1 mucositis
   Interval between last CT and surgery: 7 weeks

**7. Surgical and Pathological Findings**
 Surgery: modified radical mastectomy (left) + full axillary dissection
 Residual tumor size: 1.3 cm
 Lymph nodes: 16 removed, 4 positive
 Margins: free, minimum 4 mm
 Histologic grade: II
 Lymphovascular invasion: present
 Perineural invasion: absent
 Pathologic response: partial
 Final stage: ypT1cN2aM0 — Stage IIIA

**PATIENT 5 — SYNTHETIC RECORD FOR INCLUSION**

**1. Identification**
 Synthetic ID: INC‑005
 Age: 45 years and 6 months
 Sex / Ethnicity: Female, White
 Weight / Height / BMI: 64 kg / 1.62 m / BMI 24.4 kg/m²
 Marital Status / Occupation: Married / Dentist
 Care Center: University Hospital of Porto Alegre (RS)
 Date of Diagnosis: 14/12/2022
 Date of Surgery: 30/08/2023
 Date of Enrollment/Randomization: 05/10/2023
 ECOG Performance Status: 0

**2. History of Present Illness (HPI)**
 Patient referred due to abnormal mammogram (BI-RADS 5) performed during routine screening.
 Stereotactic biopsy on 14/12/2022 confirmed invasive ductal carcinoma, grade 3.
 Immunohistochemistry: ER 95%, PR 90%, HER2 negative (IHC 0), Ki-67: 30%.
 Initial staging: cT2N1M0 — Stage IIB (AJCC 8th edition).
 Detection method: screening mammography

**3. Personal and Family History**
 Comorbidities: none
 Allergies: not reported
 Gynecologic history: menarche at 12, regular cycles, not menopausal, G2P2A0, no hormonal contraceptive use
 Family history: maternal aunt with breast cancer at 48
 No history of chest radiotherapy
 Lifestyle: non-smoker, occasional alcohol

**4. Physical Examination (Pre-Neoadjuvant)**
 General status: excellent, ECOG 0
 Right breast: firm mass in upper outer quadrant, ~2.8 cm
 Left breast: no findings
 Right axilla: single palpable mobile node (~1.5 cm)
 No skin or nipple changes

**5. Laboratory and Imaging**
 CBC: Hb 13.0 g/dL; WBC 6,400/mm³; platelets 310,000/mm³
 Liver function: AST 27 U/L; ALT 29 U/L
 Renal function: creatinine 0.78 mg/dL; urea 32 mg/dL
 Beta-HCG: negative
 Serologies: HBsAg negative; anti-HCV negative; HIV negative
 LVEF (ECHO): 64% pre-CT → 62% post-CT
 Meets all lab and cardiac safety requirements
 Staging: CT chest/abdomen/pelvis and bone scan with no evidence of metastasis

**6. Neoadjuvant Treatment**
 Chemotherapy regimen: dose-dense AC-T

- Doxorubicin 60 mg/m² + cyclophosphamide 600 mg/m² q2w (4 cycles)
- Followed by paclitaxel 175 mg/m² q2w (4 cycles)
   Start: 10/01/2023 | End: 06/06/2023
   Total duration: 20 weeks
   No trastuzumab (HER2−)
   Endocrine therapy started post-op
   Clinical response: partial (approx. 60% reduction)
   Adverse events: grade 1 nausea and grade 2 alopecia
   Interval between last chemo and surgery: 12 weeks

**7. Surgical and Pathological Findings**
 Surgery: segmental mastectomy (right breast) + sentinel node biopsy
 Residual tumor: 0.9 cm
 Lymph nodes: 4 removed, 1 positive
 Margins: free (minimum 5 mm)
 Histologic grade: II
 Lymphovascular invasion: present
 Perineural invasion: absent
 Pathologic response: partial
 Final stage: ypT1bN1miM0 — Stage IIA

**PATIENT 6 — SYNTHETIC RECORD FOR EXCLUSION**

**1. Identification**
 Synthetic ID: INC‑006
 Age: 59 years and 2 months
 Sex / Ethnicity: Female, Mixed race
 Weight / Height / BMI: 72 kg / 1.60 m / BMI 28.1 kg/m²
 Marital Status / Occupation: Widowed / Retired teacher
 Care Center: Hospital das Clínicas da USP (SP)
 Date of Diagnosis: 03/01/2023
 Date of Surgery: 14/09/2023
 Date of Enrollment/Randomization: 30/10/2023
 ECOG Performance Status: 1

**2. History of Present Illness (HPI)**
 Patient presented with left breast lump noticed in self-exam in December 2022.
 Core needle biopsy performed on 03/01/2023 confirmed invasive ductal carcinoma, grade 3.
 Immunohistochemistry: ER 100%, PR 100%, HER2 negative (IHC 0), Ki-67: 45%.
 Initial clinical stage: cT3N2M0 — Stage IIIB (AJCC 8th edition).
 Tumor size: 5.6 cm by ultrasound; axillary nodes up to 2.3 cm.

**3. Personal and Family History**
 Comorbidities: Type 2 diabetes (on metformin), hypertension (controlled), hypothyroidism (levothyroxine)
 Allergies: None reported
 Gynecologic history: menarche at 13, menopause at 52, G3P3A0, no HRT use
 Family history: sister with breast cancer at 62
 No prior chest irradiation
 Lifestyle: ex-smoker, quit 12 years ago; no alcohol use

**4. Physical Examination (Pre-Neoadjuvant)**
 General condition: ECOG 1, no systemic symptoms
 Left breast: hard, irregular mass in upper outer quadrant, approx. 5.5 cm
 Axillary exam: multiple firm, mobile lymph nodes (~2.0–2.5 cm)
 No skin/nipple retraction, no inflammatory signs

**5. Laboratory and Imaging**
 CBC: Hb 12.7 g/dL; WBC 5,900/mm³; platelets 295,000/mm³
 Liver function: AST 31 U/L; ALT 34 U/L
 Renal function: creatinine 0.91 mg/dL; urea 35 mg/dL
 Beta-HCG: negative
 Serologies: HBsAg negative; anti-HCV negative; HIV negative
 LVEF (ECHO): 58% pre-CT → 56% post-CT
 Staging: CT chest/abdomen/pelvis and bone scan negative for metastasis

**6. Neoadjuvant Treatment**
 Chemotherapy regimen: dose-dense AC-T

- Doxorubicin 60 mg/m² + cyclophosphamide 600 mg/m² q2w (4 cycles)
- Followed by paclitaxel 80 mg/m² weekly (12 weeks)
   Start: 10/02/2023 | End: 15/06/2023
   Total duration: 18 weeks
   Endocrine therapy: started post-surgery (anastrozole)
   Clinical response: partial, tumor reduction ~50%
   Adverse events: grade 2 fatigue, grade 1 neuropathy
   Interval between last chemo and surgery: 13 weeks

**7. Surgical and Pathological Findings**
 Surgery: left modified radical mastectomy + complete axillary dissection
 Residual tumor size: 1.3 cm
 Lymph nodes: 19 resected, 4 positive
 Margins: free (minimum 6 mm)
 Histologic grade: II
 Lymphovascular invasion: present
 Perineural invasion: absent
 Pathologic response: partial
 Final stage: ypT1cN2aM0 — Stage IIIA

**PATIENT 7 — SYNTHETIC RECORD FOR EXCLUSION**

**1. Identification**
 Synthetic ID: INC‑007
 Age: 46 years and 8 months
 Sex / Ethnicity: Female, White
 Weight / Height / BMI: 64 kg / 1.65 m / BMI 23.5 kg/m²
 Marital Status / Occupation: Married / Administrative assistant
 Care Center: Instituto Nacional de Câncer (INCA, RJ)
 Date of Diagnosis: 02/02/2023
 Date of Surgery: 20/09/2023
 Date of Enrollment/Randomization: 01/11/2023
 ECOG Performance Status: 0

**2. History of Present Illness (HPI)**
 Palpable mass in right breast detected during self-exam in late January 2023.
 Core biopsy performed on 02/02/2023 confirmed invasive ductal carcinoma, grade 3.
 Initial clinical stage: cT2N1M0 — Stage IIB (AJCC 8th edition)
 Histology: IDC, Nottingham grade 3
 Immunohistochemistry: ER 90%, PR 60%, HER2 negative (IHC 1+), Ki-67: 35%

**3. Personal and Family History**
 Comorbidities: None
 Allergies: Penicillin (rash)
 Gynecologic history: menarche at 12, menopause not yet reached, G2P2A0, regular cycles, no contraceptive use
 Family history: mother with breast cancer at 57; maternal aunt with ovarian cancer
 No prior radiotherapy or chemotherapy
 Lifestyle: nonsmoker, occasional alcohol use

**4. Physical Examination (Pre-Neoadjuvant)**
 General condition: ECOG 0, no systemic complaints
 Right breast: mobile, firm 3.2 cm tumor in upper inner quadrant
 Axilla: single palpable, mobile lymph node ~1.4 cm
 No skin changes, nipple discharge or edema

**5. Laboratory and Imaging**
 CBC: Hb 13.2 g/dL; WBC 6,400/mm³; platelets 270,000/mm³
 Liver: AST 28 U/L; ALT 30 U/L
 Renal: creatinine 0.86 mg/dL
 Beta-HCG: negative
 Serologies: HBsAg, anti-HCV, HIV — all negative
 LVEF: 61% pre-CT → 60% post-CT (ECHO)
 Imaging: CT thorax/abdomen/pelvis and bone scan normal

**6. Neoadjuvant Treatment**
 Regimen: AC-T

- Doxorubicin 60 mg/m² + cyclophosphamide 600 mg/m² every 3 weeks x 4
- Followed by paclitaxel 80 mg/m² weekly x 12 weeks
   Start: 10/03/2023 | End: 18/07/2023
   Total duration: 18 weeks
   Endocrine therapy: started post-op (tamoxifen)
   Clinical response: partial tumor shrinkage (~40%)
   Adverse events: grade 1 nausea and alopecia
   Interval between last chemo and surgery: 9 weeks

**7. Surgical and Pathological Findings**
 Surgery: Right breast quadrantectomy + axillary dissection
 Residual tumor: 1.0 cm
 Lymph nodes: 15 removed, 2 positive
 Margins: negative (≥5 mm)
 Histologic grade: II
 Lymphovascular invasion: absent
 Perineural invasion: absent
 Pathological response: partial
 Final stage: ypT1bN1aM0 — Stage IIA

**PATIENT 8 — SYNTHETIC RECORD FOR EXCLUSION**

**1. Identification**
 Synthetic ID: INC‑008
 Age: 58 years and 1 month
 Sex / Ethnicity: Female, Black
 Weight / Height / BMI: 72 kg / 1.60 m / BMI 28.1 kg/m²
 Marital Status / Occupation: Widowed / Elementary school teacher
 Care Center: Hospital de Câncer de Barretos (SP)
 Date of Diagnosis: 04/06/2023
 Date of Surgery: 10/12/2023
 Date of Enrollment/Randomization: 20/01/2024
 ECOG Performance Status: 1

**2. History of Present Illness (HPI)**
 Asymptomatic lesion identified via routine mammography (May 2023).
 Ultrasound confirmed solid hypoechoic lesion, 5.3 cm, upper outer quadrant, left breast.
 Core biopsy on 04/06/2023: invasive ductal carcinoma, Nottingham grade 3.
 Initial clinical stage: cT3N1M0 — Stage IIIA (AJCC 8th edition)
 Histology: Invasive ductal carcinoma, grade 3
 Immunohistochemistry: ER 10%, PR 0%, HER2-negative (IHC 1+), Ki-67: 45%

**3. Personal and Family History**
 Comorbidities: Type 2 diabetes mellitus (oral control), dyslipidemia
 Allergies: None known
 Gynecologic history: menarche at 11, menopause at 49, G3P3A0, no hormone use
 Family history: no known cancers in first-degree relatives
 Radiation history: none
 Lifestyle: nonsmoker, former alcohol user (ceased 5 years ago)

**4. Physical Examination (Pre-Neoadjuvant)**
 General: ECOG 1, overweight
 Left breast: palpable 5.0 cm irregular, firm, fixed mass
 Axilla: mobile 1.8 cm lymph node
 No nipple retraction or skin ulceration

**5. Laboratory and Imaging**
 CBC: Hb 12.6 g/dL; WBC 5,800/mm³; platelets 320,000/mm³
 Liver: AST 32 U/L; ALT 37 U/L
 Renal: creatinine 0.94 mg/dL
 Beta-HCG: negative
 Serologies: HBsAg, anti-HCV, HIV — all negative
 LVEF: 65% pre-CT → 63% post-CT (ECHO)
 Imaging: CT chest/abdomen/pelvis and bone scan — no metastases

**6. Neoadjuvant Treatment**
 Regimen: AC-T

- Doxorubicin 60 mg/m² + cyclophosphamide 600 mg/m² (q3w × 4 cycles)
- Paclitaxel 80 mg/m² weekly × 12 weeks
   Start: 01/08/2023 | End: 25/11/2023
   Total duration: 17 weeks
   Clinical response: partial (volume reduction estimated at 50%)
   Adverse events: grade 2 fatigue, grade 1 constipation
   Endocrine therapy: initiated post-op (aromatase inhibitor)
   Interval between last chemo and surgery: 2 weeks

**7. Surgical and Pathological Findings**
 Surgery: Left mastectomy + full axillary dissection
 Residual tumor: 1.4 cm
 Lymph nodes: 19 removed, 4 positive
 Margins: negative (≥3 mm)
 Histologic grade: II
 Lymphovascular invasion: present
 Perineural invasion: absent
 Pathological response: partial
 Final stage: ypT1cN2aM0 — Stage IIIA

**PATIENT 9 — SYNTHETIC RECORD FOR EXCLUSION**

**1. Identification**
 Synthetic ID: INC‑009
 Age: 62 years and 5 months
 Sex / Ethnicity: Female, White
 Weight / Height / BMI: 68 kg / 1.58 m / BMI 27.2 kg/m²
 Marital Status / Occupation: Married / Librarian (retired)
 Care Center: Instituto Nacional de Câncer (INCA - RJ)
 Date of Diagnosis: 12/05/2023
 Date of Surgery: 05/12/2023
 Date of Enrollment/Randomization: 22/01/2024
 ECOG Performance Status: 0

**2. History of Present Illness (HPI)**
 Palpable lump in the right breast noted in April 2023.
 Diagnostic mammogram and ultrasound: spiculated mass, 3.7 cm, upper inner quadrant.
 Core biopsy on 12/05/2023: invasive carcinoma of no special type (ductal), Nottingham grade 3.
 Initial clinical staging: cT2N2M0 — Stage IIIA (AJCC 8th edition)
 Immunohistochemistry:

- ER: 90%
- PR: 80%
- HER2: 0 (IHC)
- Ki-67: 32%

**3. Personal and Family History**
 Comorbidities: Hypertension (controlled), hypothyroidism
 Allergies: None known
 Gynecologic history: menarche at 13, menopause at 52, G2P2A0, no HRT
 Family history: maternal aunt with breast cancer at 67
 Radiotherapy: none prior
 Lifestyle: nonsmoker, moderate alcohol use (social)

**4. Physical Examination (Pre-Neoadjuvant)**
 General: ECOG 0, no systemic signs
 Right breast: firm 3.5 cm tumor, non-fixed, upper inner quadrant
 Right axilla: multiple mobile lymph nodes (~2.0 cm)
 No skin infiltration or nipple discharge

**5. Laboratory and Imaging**
 CBC: Hb 13.1 g/dL; WBC 6,300/mm³; Platelets 295,000/mm³
 Liver: AST 29 U/L; ALT 30 U/L
 Renal: creatinine 0.91 mg/dL
 Beta-HCG: negative
 LVEF: 62% pre-CT → 60% post-CT (ECHO)
 Serologies: HBsAg, anti-HCV, HIV — all negative
 Imaging: chest/abdomen/pelvis CT and bone scan — no evidence of metastasis

**6. Neoadjuvant Treatment**
 Regimen: AC-T

- Doxorubicin 60 mg/m² + cyclophosphamide 600 mg/m² (q3w × 4 cycles)
- Paclitaxel 80 mg/m² weekly × 12 weeks
   Start: 15/07/2023 | End: 25/11/2023
   Total duration: 18 weeks
   Clinical response: partial (tumor ~50% reduction)
   Adverse events: grade 2 peripheral neuropathy (resolved), alopecia
   No dose delays or hospitalizations
   Endocrine therapy: initiated after surgery (letrozole)
   Interval to surgery: 10 days

**7. Surgical and Pathological Findings**
 Surgery: Right lumpectomy + axillary lymphadenectomy
 Residual tumor: 1.3 cm invasive carcinoma
 Lymph nodes: 21 removed, 5 positive
 Margins: negative (≥2 mm)
 Post-chemo histological grade: II
 Lymphovascular invasion: present
 Perineural invasion: absent
 Pathological response: partial
 Final stage: ypT1cN2aM0 — Stage IIIA

**PATIENT 10 — SYNTHETIC RECORD FOR EXCLUSION**

**1. Identification**
 Synthetic ID: INC‑010
 Age: 45 years and 7 months
 Sex / Ethnicity: Female, Black
 Weight / Height / BMI: 74 kg / 1.65 m / BMI 27.2 kg/m²
 Marital Status / Occupation: Divorced / Public school teacher
 Care Center: Hospital de Câncer de Barretos (SP)
 Date of Diagnosis: 02/06/2023
 Date of Surgery: 14/12/2023
 Date of Enrollment/Randomization: 08/01/2024
 ECOG Performance Status: 1

**2. History of Present Illness (HPI)**
 Routine mammogram in May 2023 showed a 4.2 cm spiculated lesion in the upper outer quadrant of the left breast.
 Core biopsy (02/06/2023): invasive carcinoma, grade 3 (Nottingham), HR+/HER2-.
 Initial clinical stage: cT2N1M0 — Stage IIB (AJCC 8th ed).
 Immunohistochemistry:

- ER: 95%
- PR: 60%
- HER2: 0 (IHC)
- Ki-67: 38%

**3. Personal and Family History**
 Comorbidities: Mild asthma (controlled), no cardiovascular disease
 Allergies: none
 Gynecologic history: menarche at 11, G3P2A1, menopause not reached
 Hormonal history: prior use of oral contraceptives (5 years), no recent use
 Family history: mother with ovarian cancer at age 62
 No previous thoracic radiotherapy
 Non-smoker, denies alcohol use

**4. Physical Examination**
 General condition: ECOG 1, preserved
 Left breast: firm, irregular mass ~4.0 cm, fixed to superficial plane
 Left axilla: 2 palpable mobile lymph nodes (up to 1.8 cm)
 No cutaneous infiltration or nipple changes

**5. Laboratory and Imaging**
 CBC: Hb 12.6 g/dL; WBC 7,100/mm³; Platelets 305,000/mm³
 Liver: AST 27 U/L; ALT 33 U/L
 Renal: Creatinine 0.88 mg/dL
 Beta-HCG: negative
 LVEF: 66% pre-CT → 64% post-CT (ECHO), variation <15 pp
 Serologies: HBsAg, anti-HCV, HIV — all negative
 Imaging: staging CT (thorax, abdomen, pelvis) and bone scan — no distant metastases

**6. Neoadjuvant Treatment**
 Regimen: AC-T

- Doxorubicin 60 mg/m² + Cyclophosphamide 600 mg/m² q21d × 4 cycles
- Paclitaxel 80 mg/m² weekly × 12 weeks
   Start: 01/07/2023 | End: 11/11/2023
   Total duration: 19 weeks
   Trastuzumab: not indicated (HER2-negative)
   Endocrine therapy: not started before surgery
   Clinical response: partial reduction (approx. 60%)
   Adverse events: grade 1 fatigue, grade 2 nausea, no dose-limiting toxicities
   Interval to surgery: 33 days

**7. Surgical and Pathological Findings**
 Surgery: Left quadrantectomy + axillary dissection
 Residual tumor: 1.5 cm invasive carcinoma
 Histologic grade post-CT: II
 Lymph nodes: 13 removed, 3 positive
 Margins: negative (≥3 mm)
 Lymphovascular invasion: present
 Perineural invasion: absent
 Pathological response: partial
 Final stage: ypT1cN1aM0 — Stage IIA

**PATIENT 11 — SYNTHETIC RECORD FOR EXCLUSION**

**1. Identification**
 Synthetic ID: INC‑011
 Age: 57 years and 3 months
 Sex / Ethnicity: Female, White
 Weight / Height / BMI: 63 kg / 1.58 m / BMI 25.2 kg/m²
 Marital Status / Occupation: Married / Accountant
 Care Center: Instituto Nacional de Câncer (INCA)
 Date of Diagnosis: 18/04/2023
 Date of Surgery: 02/11/2023
 Date of Enrollment/Randomization: 18/12/2023
 ECOG Performance Status: 0

**2. History of Present Illness (HPI)**
 Patient presented with palpable mass in the right breast during self-examination in March 2023.
 Ultrasound confirmed irregular mass, 3.9 cm, upper outer quadrant.
 Core biopsy (18/04/2023): invasive carcinoma, grade 3 (Nottingham), HR+/HER2-.
 Clinical stage at presentation: cT2N2M0 — Stage IIIA (AJCC 8th ed).
 Immunohistochemistry:

- ER: 100%
- PR: 90%
- HER2: 1+
- Ki-67: 42%

**3. Personal and Family History**
 Comorbidities: Hypertension (controlled), Type 2 Diabetes (on metformin)
 Allergies: none
 Gynecologic history: menarche at 12, menopause at 50, G4P3A1
 Hormonal history: no HRT use
 Family history: paternal aunt with breast cancer, no known BRCA mutations
 No prior chest radiation
 Former smoker (quit 10 years ago), moderate alcohol use

**4. Physical Examination**
 General condition: ECOG 0, well-nourished
 Right breast: firm 3.5–4.0 cm mass, no skin changes
 Right axilla: 3 enlarged lymph nodes, firm, mobile
 No nipple inversion or discharge

**5. Laboratory and Imaging**
 CBC: Hb 11.8 g/dL; WBC 6,800/mm³; Platelets 287,000/mm³
 Liver: AST 31 U/L; ALT 35 U/L
 Renal: Creatinine 0.92 mg/dL
 Beta-HCG: negative
 LVEF: Pre-CT 68% → Post-CT 63% (ECHO) — variation <15 pp
 Serologies: HBsAg, anti-HCV, HIV — negative
 Staging: CT thorax/abdomen/pelvis and bone scintigraphy — no distant disease

**6. Neoadjuvant Treatment**
 Regimen: AC followed by weekly paclitaxel

- Doxorubicin 60 mg/m² + Cyclophosphamide 600 mg/m² q21d × 4 cycles
- Paclitaxel 80 mg/m² weekly × 12 weeks
   Start: 15/05/2023 | End: 23/09/2023
   Duration: 18 weeks
   Trastuzumab: not indicated
   Endocrine therapy: initiated post-randomization
   Clinical response: partial (~50% reduction)
   Adverse events: grade 2 fatigue and alopecia, no dose delays
   Interval from last chemo to surgery: 40 days

**7. Surgical and Pathological Findings**
 Surgery: Right breast quadrantectomy + complete axillary dissection
 Tumor residual size: 1.8 cm
 Histologic grade: II
 Lymph nodes: 16 removed, 4 positive
 Margins: negative (clearance ≥4 mm)
 Lymphovascular invasion: present
 Perineural invasion: absent
 Pathologic response: partial
 Final pathological stage: ypT1cN2aM0 — Stage IIIA

**PATIENT 12 — SYNTHETIC RECORD FOR EXCLUSION**

**1. Identification**
 Synthetic ID: INC‑012
 Age: 49 years and 9 months
 Sex / Ethnicity: Female, Mixed race
 Weight / Height / BMI: 74 kg / 1.62 m / BMI 28.2 kg/m²
 Marital Status / Occupation: Divorced / Schoolteacher
 Care Center: Hospital de Câncer de Barretos
 Date of Diagnosis: 04/06/2023
 Date of Surgery: 28/11/2023
 Date of Enrollment/Randomization: 10/01/2024
 ECOG Performance Status: 0

**2. History of Present Illness (HPI)**
 Patient reported nodule detected during routine mammography.
 Ultrasound: solid lesion, 4.2 cm, upper inner quadrant of left breast
 Core biopsy (04/06/2023): invasive ductal carcinoma, grade 3 (Nottingham)
 Clinical staging: cT2N1M0 — Stage IIB (AJCC 8th ed.)
 Immunohistochemistry:

- ER: 95%
- PR: 80%
- HER2: 0
- Ki-67: 38%

**3. Personal and Family History**
 Comorbidities: Controlled asthma
 No known drug allergies
 Gynecologic history: G2P2A0, menopause at 47
 Hormonal history: no HRT use
 Family history: maternal grandmother with ovarian cancer
 No previous radiotherapy
 Never smoked; social alcohol consumption

**4. Physical Examination**
 General condition: ECOG 0
 Left breast: firm 4.0 cm lesion, no skin retraction
 Left axilla: one palpable, mobile 1.5 cm lymph node
 No nipple changes or discharge

**5. Laboratory and Imaging**
 CBC: Hb 12.5 g/dL; WBC 7,200/mm³; Platelets 310,000/mm³
 Liver: AST 29 U/L; ALT 27 U/L
 Renal: Creatinine 0.88 mg/dL
 Beta-HCG: negative
 LVEF: Pre-CT 65% → Post-CT 61% (ECHO)
 Serologies: HBsAg, anti-HCV, HIV — non-reactive
 Staging imaging (CT, bone scan): negative for metastasis

**6. Neoadjuvant Treatment**
 Regimen: Dose-dense AC followed by paclitaxel

- Doxorubicin 60 mg/m² + Cyclophosphamide 600 mg/m² q14d × 4
- Paclitaxel 80 mg/m² weekly × 12
   Start: 25/06/2023 | End: 07/11/2023
   Total duration: 19 weeks
   Trastuzumab: not applicable
   Endocrine therapy: started post-randomization
   Clinical response: partial (estimated 60% reduction)
   Adverse events: grade 1 neuropathy, grade 2 nausea
   Interval to surgery: 21 days

**7. Surgical and Pathological Findings**
 Surgery: Left breast quadrantectomy + sentinel lymph node biopsy followed by axillary clearance
 Residual tumor: 1.2 cm
 Histologic grade: II
 Lymph nodes: 12 examined, 2 positive
 Margins: negative (minimum 5 mm clearance)
 Lymphovascular invasion: present
 Perineural invasion: absent
 Pathologic response: partial
 Final pathologic stage: ypT1cN1aM0 — Stage IIB

**PATIENT 13 — SYNTHETIC RECORD FOR EXCLUSION**

**1. Identification**
 Synthetic ID: INC‑013
 Age: 58 years and 3 months
 Sex / Ethnicity: Female, White
 Weight / Height / BMI: 63 kg / 1.60 m / BMI 24.6 kg/m²
 Marital Status / Occupation: Widow / Accountant
 Care Center: Instituto Nacional de Câncer (INCA)
 Date of Diagnosis: 03/03/2023
 Date of Surgery: 10/10/2023
 Date of Enrollment/Randomization: 15/12/2023
 ECOG Performance Status: 1

**2. History of Present Illness (HPI)**
 Patient presented with palpable nodule in left breast.
 Detected during self-examination.
 Core biopsy (03/03/2023): invasive ductal carcinoma, grade 3 (Nottingham)
 Clinical stage: cT3N2M0 — Stage IIIA (AJCC 8th ed.)
 Immunohistochemistry:

- ER: 90%
- PR: 10%
- HER2: 0 (IHC 1+)
- Ki-67: 32%

**3. Personal and Family History**
 Comorbidities: Hypertension, controlled with losartan
 No drug allergies
 Gynecologic history: G3P3A0, menopause at 50
 No hormone replacement therapy
 Family history: Sister with breast cancer at 46 years
 No prior radiation therapy
 Former smoker, ceased 15 years ago; denies alcohol use

**4. Physical Examination**
 General condition: ECOG 1
 Left breast: 5.2 cm lesion, upper outer quadrant
 Skin: mild thickening, no ulceration
 Axilla: fixed 2.0 cm lymph node cluster
 No contralateral findings

**5. Laboratory and Imaging**
 CBC: Hb 11.9 g/dL; WBC 6,900/mm³; Platelets 280,000/mm³
 Liver: AST 32 U/L; ALT 29 U/L
 Renal: Creatinine 0.96 mg/dL
 Beta-HCG: negative
 LVEF: Pre-CT 68% → Post-CT 63% (ECHO)
 Serologies: HBsAg, anti-HCV, HIV — negative
 Staging imaging (CT chest/abdomen/pelvis, bone scan): no metastases

**6. Neoadjuvant Treatment**
 Regimen: AC-T

- Doxorubicin 60 mg/m² + Cyclophosphamide 600 mg/m² q21d × 4
- Docetaxel 100 mg/m² q21d × 4
   Start: 20/03/2023 | End: 25/07/2023
   Total duration: 18 weeks
   Trastuzumab: not applicable
   Endocrine therapy: not initiated prior to randomization
   Clinical response: tumor reduced from 5.2 cm to 2.3 cm
   Adverse events: Grade 2 asthenia, Grade 1 anemia
   Interval to surgery: 11 weeks

**7. Surgical and Pathological Findings**
 Surgery: Modified radical mastectomy + axillary lymphadenectomy
 Residual tumor: 2.1 cm
 Histologic grade: II
 Lymph nodes: 16 resected, 4 positive
 Margins: negative (minimum clearance 4 mm)
 Lymphovascular invasion: present
 Perineural invasion: not identified
 Pathologic response: partial
 Final pathologic stage: ypT2N2aM0 — Stage IIIA

**PATIENT 14 — SYNTHETIC RECORD FOR EXCLUSION**

**1. Identification**
 Synthetic ID: INC‑014
 Age: 47 years and 9 months
 Sex / Ethnicity: Female, Brown
 Weight / Height / BMI: 72 kg / 1.65 m / BMI 26.4 kg/m²
 Marital Status / Occupation: Married / Elementary School Teacher
 Care Center: Hospital de Câncer de Barretos
 Date of Diagnosis: 25/07/2023
 Date of Surgery: 05/01/2024
 Date of Enrollment/Randomization: 28/03/2024
 ECOG Performance Status: 0

**2. History of Present Illness (HPI)**
 Palpable lump found in left breast during routine screening mammogram.
 Ultrasound-guided core biopsy (25/07/2023): invasive ductal carcinoma, grade 3
 Clinical staging: cT2N1M0 — Stage IIB (AJCC 8th ed.)
 Immunohistochemistry:

- ER: 98%
- PR: 80%
- HER2: 0 (IHC 0)
- Ki-67: 40% (pre-treatment core biopsy)

**3. Personal and Family History**
 Comorbidities: None reported
 Allergies: No known drug allergies
 Gynecologic history: G2P2A0, menopause not reached, regular cycles
 No history of hormonal contraceptive use
 Family history: Mother with breast cancer at 65 years
 No prior thoracic radiation
 Non-smoker, occasional social alcohol use

**4. Physical Examination**
 Good general condition, ECOG 0
 Left breast: 3.4 cm firm lesion, upper inner quadrant
 No skin retraction or discharge
 Ipsilateral axilla: 1 mobile, firm 1.8 cm lymph node
 No contralateral abnormalities

**5. Laboratory and Imaging**
 CBC: Hb 12.4 g/dL; WBC 7,100/mm³; Platelets 295,000/mm³
 Liver function: AST 30 U/L; ALT 27 U/L
 Renal function: Creatinine 0.83 mg/dL
 Beta-HCG: Negative
 LVEF (ECHO): 66% before chemotherapy → 64% after chemotherapy
 Serologies: HBsAg, anti-HCV, HIV — all negative
 Imaging: CT chest/abdomen/pelvis and bone scintigraphy showed no distant disease

**6. Neoadjuvant Treatment**
 Regimen: AC-T

- Doxorubicin 60 mg/m² + Cyclophosphamide 600 mg/m² q21d × 4
- Paclitaxel 80 mg/m² weekly × 12
   Start: 15/08/2023 | End: 28/11/2023
   Total duration: 15 weeks
   Trastuzumab: Not indicated (HER2-negative)
   Endocrine therapy: Not initiated pre-randomization
   Clinical response: ~50% reduction in tumor volume
   Adverse events: Grade 1 nausea, Grade 2 peripheral neuropathy (resolved by surgery)
   Interval to surgery: 5 weeks

**7. Surgical and Pathological Findings**
 Surgery: Breast-conserving surgery + sentinel lymph node biopsy
 Residual tumor: 1.1 cm
 Histologic grade: II
 Margins: clear (minimal clearance 6 mm)
 Lymph nodes: 3 removed, 1 positive (micrometastasis)
 Lymphovascular invasion: Present
 Perineural invasion: Absent
 Pathologic response: Partial
 Pathologic stage: ypT1cN1miM0 — Stage IIA

**PATIENT 15 — SYNTHETIC RECORD FOR EXCLUSION**

**1. Identification**
 Synthetic ID: INC‑015
 Age: 59 years and 2 months
 Sex / Ethnicity: Female, White
 Weight / Height / BMI: 80 kg / 1.68 m / BMI 28.3 kg/m²
 Marital Status / Occupation: Divorced / Retired Nurse
 Care Center: Instituto Nacional de Câncer (INCA)
 Date of Diagnosis: 14/06/2023
 Date of Surgery: 09/01/2024
 Date of Enrollment/Randomization: 25/03/2024
 ECOG Performance Status: 1

**2. History of Present Illness (HPI)**
 Palpable nodule in right breast noticed during self-exam.
 Core biopsy performed on 14/06/2023: invasive ductal carcinoma, grade 3 (Nottingham 9/9)
 Clinical staging: cT2N2M0 — Stage IIIA (AJCC 8th ed.)
 Immunohistochemistry:

- ER: 90%
- PR: 70%
- HER2: 0 (IHC 0)
- Ki-67: 32% (pre-treatment core biopsy)

**3. Personal and Family History**
 Comorbidities: Hypertension and hypothyroidism, both controlled
 Allergies: No known drug allergies
 Gynecologic history: Menopause at 52; G3P3A0
 No hormone replacement therapy
 Family history: No breast/ovarian/prostate cancers
 No history of thoracic radiotherapy
 Non-smoker, occasional wine consumption

**4. Physical Examination**
 Good general condition, ECOG 1
 Right breast: 3.6 cm firm, irregular mass in lower outer quadrant
 Skin: mild thickening without ulceration or retraction
 Axilla: fixed 2.0 cm lymph node
 No abnormalities in left breast

**5. Laboratory and Imaging**
 CBC: Hb 13.1 g/dL; WBC 6,800/mm³; Platelets 280,000/mm³
 Liver: AST 29 U/L; ALT 30 U/L
 Renal: Creatinine 0.90 mg/dL
 Beta-HCG: Negative
 LVEF (MUGA): 60% pre-chemo → 61% post-chemo
 Serologies: Negative for HBsAg, anti-HCV, and HIV
 Imaging: CT chest/abdomen/pelvis + bone scan — no metastasis

**6. Neoadjuvant Treatment**
 Regimen: Dose-dense AC + Paclitaxel

- Doxorubicin 60 mg/m² + Cyclophosphamide 600 mg/m² q14d × 4
- Paclitaxel 80 mg/m² weekly × 12
   Start: 10/07/2023 | End: 20/11/2023
   Total duration: 18 weeks
   Trastuzumab: Not indicated (HER2-negative)
   Endocrine therapy: Not started prior to randomization
   Clinical response: Partial tumor regression (~45%)
   Adverse events: G2 neutropenia (non-febrile), managed with G-CSF
   Interval to surgery: 7 weeks

**7. Surgical and Pathological Findings**
 Surgery: Right mastectomy + axillary lymph node dissection
 Tumor residual: 1.5 cm
 Histologic grade: II
 Margins: free (minimal clearance 5 mm)
 Lymph nodes: 18 removed, 4 positive
 Lymphovascular invasion: Present
 Perineural invasion: Absent
 Pathologic response: Partial
 Final pathological staging: ypT1cN2aM0 — Stage IIIA

**PATIENT 16 — SYNTHETIC RECORD FOR EXCLUSION**

**1. Identification**
 Synthetic ID: INC‑016
 Age: 47 years and 10 months
 Sex / Ethnicity: Female, Mixed-race
 Weight / Height / BMI: 70 kg / 1.65 m / BMI 25.7 kg/m²
 Marital Status / Occupation: Married / Administrative Assistant
 Care Center: Hospital de Câncer de Barretos
 Date of Diagnosis: 02/03/2023
 Date of Surgery: 28/09/2023
 Date of Enrollment/Randomization: 05/01/2024
 ECOG Performance Status: 0

**2. History of Present Illness (HPI)**
 Mammographic abnormality followed by ultrasound confirmation of solid nodule.
 Core biopsy (02/03/2023): Invasive carcinoma of no special type, grade 3 (Nottingham 8/9)
 Initial clinical staging: cT2N1M0 — Stage IIB (AJCC 8th ed.)
 Immunohistochemistry:

- ER: 95%
- PR: 80%
- HER2: Negative (IHC 0)
- Ki-67: 38% (pre-treatment)

**3. Personal and Family History**
 Comorbidities: Type 2 Diabetes Mellitus, well controlled with metformin
 No drug allergies
 Gynecologic history: G2P2A0; menopause at 46 due to oophorectomy (benign cyst)
 No hormonal use
 Family history: Mother with breast cancer at 60
 Non-smoker, non-alcoholic

**4. Physical Examination**
 Normal general condition, ECOG 0
 Right breast: 3.2 cm tumor in upper inner quadrant
 Skin: intact
 Nipple: no retraction or discharge
 Right axilla: 1.5 cm mobile lymph node
 No findings in left breast

**5. Laboratory and Imaging**
 CBC: Hb 12.8 g/dL; WBC 7,200/mm³; Platelets 270,000/mm³
 Liver function: AST 25 U/L; ALT 28 U/L
 Renal function: Creatinine 0.82 mg/dL
 Beta-HCG: Negative
 LVEF (ECHO): 63% pre-CT → 64% post-CT
 Serologies: Negative (HBsAg, anti-HCV, HIV)
 Imaging: CT scans and bone scintigraphy: no metastases

**6. Neoadjuvant Treatment**
 Regimen: AC-T

- Doxorubicin 60 mg/m² + Cyclophosphamide 600 mg/m² q21d × 4
- Paclitaxel 80 mg/m² weekly × 12
   Start: 20/03/2023 | End: 10/07/2023
   Total duration: 16 weeks
   No HER2 blockade (HER2-negative)
   No endocrine therapy prior to randomization
   Clinical response: Tumor reduction ~50%
   Adverse events: Alopecia G2; no grade ≥3 toxicities
   Surgery performed 11 weeks after last chemo dose

**7. Surgical and Pathological Findings**
 Surgery: Segmental mastectomy with axillary dissection
 Residual tumor: 1.0 cm
 Post-chemo histological grade: II
 Surgical margins: clear (minimum 4 mm)
 Lymph nodes: 11 removed, 2 positive
 Lymphovascular invasion: Present
 Perineural invasion: Absent
 Pathologic response: Partial
 Final staging: ypT1bN1aM0 — Stage IIB

**PATIENT 17 — SYNTHETIC RECORD FOR EXCLUSION**

**1. Identification**
 Synthetic ID: INC‑017
 Age: 62 years and 4 months
 Sex / Ethnicity: Female, White
 Weight / Height / BMI: 68 kg / 1.58 m / BMI 27.2 kg/m²
 Marital Status / Occupation: Widowed / Retired teacher
 Care Center: Instituto Nacional de Câncer (INCA)
 Date of Diagnosis: 10/05/2023
 Date of Surgery: 05/11/2023
 Date of Enrollment/Randomization: 20/02/2024
 ECOG Performance Status: 1

**2. History of Present Illness (HPI)**
 Palpable nodule discovered during routine self-exam.
 Ultrasound and mammogram confirmed suspicious lesion.
 Core biopsy (10/05/2023): Invasive ductal carcinoma, grade 3 (Nottingham 8/9)
 Initial clinical staging: cT3N2M0 — Stage IIIA (AJCC 8th ed.)
 Immunohistochemistry:

- ER: 90%
- PR: 70%
- HER2: Negative (IHC 1+)
- Ki-67: 42%

**3. Personal and Family History**
 Comorbidities: Controlled hypertension and dyslipidemia
 Allergies: None reported
 Gynecologic history: G3P3A0; natural menopause at 51
 No current or prior hormonal therapy
 Family history: Sister with ovarian cancer at age 58
 Never smoked or consumed alcohol

**4. Physical Examination**
 General condition: good; ECOG 1
 Left breast: 4.8 cm tumor in upper outer quadrant
 Skin: mild retraction, no ulceration
 Nipple: preserved
 Axilla: multiple palpable lymph nodes (2.0–2.5 cm), mobile

**5. Laboratory and Imaging**
 CBC: Hb 12.1 g/dL; WBC 6,800/mm³; Platelets 250,000/mm³
 Liver function: AST 31 U/L; ALT 26 U/L
 Renal function: Creatinine 0.78 mg/dL
 Beta-HCG: Negative
 LVEF (ECHO): Pre-CT 61%; Post-CT 60%
 Serologies: Non-reactive (HBsAg, anti-HCV, HIV)
 Imaging (CT and bone scan): No evidence of metastasis

**6. Neoadjuvant Treatment**
 Regimen: AC followed by weekly paclitaxel

- Doxorubicin 60 mg/m² + Cyclophosphamide 600 mg/m² q21d × 4 cycles
- Paclitaxel 80 mg/m² weekly × 12
   Start: 05/06/2023 | End: 25/09/2023
   Duration: 16 weeks
   No HER2-targeted therapy (HER2-negative)
   No endocrine therapy before randomization
   Clinical response: Partial, ~60% tumor reduction
   Adverse events: Grade 1 nausea, Grade 2 fatigue
   Interval between last chemotherapy and surgery: 6 weeks

**7. Surgical and Pathological Findings**
 Surgery: Left quadrantectomy + axillary lymph node dissection
 Residual tumor: 1.3 cm
 Margins: free (minimum 3.5 mm)
 Histological grade: II
 Lymph nodes: 16 removed, 3 positive
 Lymphovascular invasion: Present
 Perineural invasion: Not identified
 Pathologic response: Partial
 Final staging: ypT1cN2aM0 — Stage IIIA

**PATIENT 18 — SYNTHETIC RECORD FOR EXCLUSION**

**1. Identification**
 Synthetic ID: INC‑018
 Age: 54 years and 2 months
 Sex / Ethnicity: Female, Black
 Weight / Height / BMI: 74 kg / 1.60 m / BMI 28.9 kg/m²
 Marital Status / Occupation: Married / Administrative assistant
 Care Center: Hospital do Câncer de Barretos
 Date of Diagnosis: 28/04/2023
 Date of Surgery: 22/09/2023
 Date of Enrollment/Randomization: 02/01/2024
 ECOG Performance Status: 0

**2. History of Present Illness (HPI)**
 Lesion detected on screening mammogram.
 Subsequent ultrasound revealed BI-RADS 5 nodule.
 Core biopsy (28/04/2023): Invasive ductal carcinoma, Nottingham grade 3 (9/9)
 Initial clinical staging: cT2N2M0 — Stage IIIA (AJCC 8th ed.)
 Immunohistochemistry:

- ER: 95%
- PR: 80%
- HER2: Negative (IHC 0)
- Ki-67: 38%

**3. Personal and Family History**
 Comorbidities: Type 2 diabetes mellitus (controlled), obesity
 Allergies: No known drug allergies
 Gynecologic history: G2P2A0; menopause at 49
 No hormonal therapy use
 Family history: Mother with breast cancer at age 61
 No smoking or alcohol use

**4. Physical Examination**
 Good general condition (ECOG 0)
 Right breast: 3.5 cm firm, irregular, non-adherent nodule in lower inner quadrant
 No skin or nipple changes
 Palpable axillary lymph nodes (up to 2.0 cm), firm, mobile

**5. Laboratory and Imaging**
 CBC: Hb 11.9 g/dL; WBC 7,200/mm³; Platelets 275,000/mm³
 Liver function: AST 28 U/L; ALT 22 U/L
 Renal function: Creatinine 0.84 mg/dL
 Beta-HCG: Negative
 LVEF (ECHO): Pre-CT 60%; Post-CT 59%
 Serologies: Non-reactive (HBsAg, anti-HCV, HIV)
 Imaging (CT chest/abdomen/pelvis, bone scan): No metastasis

**6. Neoadjuvant Treatment**
 Regimen: Dose-dense AC followed by paclitaxel

- Doxorubicin 60 mg/m² + Cyclophosphamide 600 mg/m² q14d × 4
- Paclitaxel 80 mg/m² weekly × 12
   Start: 22/05/2023 | End: 11/09/2023
   Duration: 16 weeks
   No HER2-targeted agents (HER2-negative)
   No endocrine therapy initiated pre-randomization
   Clinical response: Partial, tumor decreased by ~50%
   Adverse events: Grade 2 fatigue, Grade 1 anemia
   Interval to surgery: 11 days

**7. Surgical and Pathological Findings**
 Surgery: Right lumpectomy + axillary lymphadenectomy
 Tumor size: 1.0 cm (residual)
 Margins: Free (minimum 4 mm)
 Histological grade post-CT: II
 Lymph nodes: 14 removed, 2 positive
 Lymphovascular invasion: Present
 Perineural invasion: Absent
 Pathologic response: Partial
 Final staging: ypT1bN1aM0 — Stage IIB

**PATIENT 19 — SYNTHETIC RECORD FOR EXCLUSION**

**1. Identification**
 Synthetic ID: INC‑019
 Age: 63 years and 7 months
 Sex / Ethnicity: Female, White
 Weight / Height / BMI: 67 kg / 1.57 m / BMI 27.2 kg/m²
 Marital Status / Occupation: Widowed / Retired teacher
 Care Center: Instituto Nacional de Câncer (INCA)
 Date of Diagnosis: 10/03/2023
 Date of Surgery: 05/09/2023
 Date of Enrollment/Randomization: 03/01/2024
 ECOG Performance Status: 1

**2. History of Present Illness (HPI)**
 Patient referred after abnormal screening mammography and ultrasound.
 Core biopsy performed on 10/03/2023: Invasive ductal carcinoma, Nottingham grade 3 (8/9)
 Initial clinical stage: cT2N1M0 — Stage IIB (AJCC 8th ed.)
 Immunohistochemistry:

- ER: 98%
- PR: 70%
- HER2: 0 (Negative)
- Ki-67: 32%

**3. Personal and Family History**
 Comorbidities: Hypertension, dyslipidemia (both controlled)
 Allergies: None reported
 Gynecologic history: G3P3A0; menopause at 51
 No history of hormone replacement therapy
 Family history: Sister with ovarian cancer at 53
 Former smoker (ceased 15 years ago); no alcohol consumption

**4. Physical Examination**
 ECOG 1 — no limitations in self-care
 Left breast: Palpable 3.0 cm irregular nodule in upper outer quadrant
 No nipple inversion or skin involvement
 Ipsilateral axillary lymph node (1.8 cm), mobile and firm

**5. Laboratory and Imaging**
 CBC: Hb 12.1 g/dL; WBC 6,800/mm³; Platelets 289,000/mm³
 Liver function: AST 25 U/L; ALT 29 U/L
 Renal function: Creatinine 0.92 mg/dL
 Beta-HCG: Negative
 LVEF: Pre-CT 62%; Post-CT 60%
 Serologies: HBsAg negative, anti-HCV negative, HIV negative
 Staging CT and bone scan: No signs of metastasis

**6. Neoadjuvant Treatment**
 Chemotherapy regimen:

- Doxorubicin 60 mg/m² + Cyclophosphamide 600 mg/m² q21d × 4
- Followed by Docetaxel 75 mg/m² q21d × 4
   Start: 03/04/2023 | End: 17/07/2023
   Duration: 16 weeks
   HER2-targeted therapy: Not applicable (HER2-negative)
   No endocrine therapy before randomization
   Clinical response: Partial reduction of tumor (approx. 45%)
   Adverse events: Grade 1 nausea, Grade 2 alopecia
   Surgery performed 7 weeks after last CT cycle

**7. Surgical and Pathological Findings**
 Surgery: Left quadrantectomy + sentinel lymph node biopsy
 Residual tumor: 1.3 cm
 Margins: Clear (≥5 mm)
 Histological grade post-CT: II
 Lymph nodes: 4 removed, 1 positive
 Lymphovascular invasion: Present
 Perineural invasion: Absent
 Pathologic response: Partial
 Final staging: ypT1cN1miM0 — Stage IIA

**PATIENT 20 — SYNTHETIC RECORD FOR EXCLUSION**

**1. Identification**
 Synthetic ID: INC‑020
 Age: 54 years and 10 months
 Sex / Ethnicity: Female, Mixed race
 Weight / Height / BMI: 74 kg / 1.63 m / BMI 27.9 kg/m²
 Marital Status / Occupation: Married / Administrative assistant
 Care Center: Hospital de Câncer de Barretos
 Date of Diagnosis: 05/02/2023
 Date of Surgery: 02/08/2023
 Date of Enrollment/Randomization: 15/11/2023
 ECOG Performance Status: 0

**2. History of Present Illness (HPI)**
 Patient detected palpable mass during self-exam in late January 2023.
 Core biopsy performed on 05/02/2023: Invasive ductal carcinoma, grade 3 (Nottingham)
 Initial staging: cT2N1M0 — Stage IIB (AJCC 8th ed.)
 Immunohistochemistry:

- ER: 95%
- PR: 90%
- HER2: 0 (Negative)
- Ki-67: 42%

**3. Personal and Family History**
 Comorbidities: Controlled type 2 diabetes mellitus
 Allergies: None known
 Gynecologic history: G2P2A0; menopause at 52
 No prior hormonal therapy
 Family history: Mother with breast cancer at 65
 No smoking; occasional alcohol use

**4. Physical Examination**
 General condition: Good (ECOG 0)
 Right breast: Palpable tumor in lower inner quadrant, approx. 3.2 cm
 No nipple discharge or retraction
 1 palpable axillary lymph node (~1.5 cm), mobile

**5. Laboratory and Imaging**
 CBC: Hb 12.8 g/dL; WBC 7,100/mm³; Platelets 301,000/mm³
 Renal function: Creatinine 0.83 mg/dL; Urea 34 mg/dL
 Liver enzymes: AST 28 U/L; ALT 31 U/L
 Beta-HCG: Negative
 LVEF (ECHO): Pre-CT 59%; Post-CT 57%
 Serologies: HBV, HCV, HIV — all non-reactive
 Imaging: CT chest/abdomen/pelvis and bone scan — no metastases

**6. Neoadjuvant Treatment**
 Chemotherapy regimen:

- AC (Doxorubicin 60 mg/m² + Cyclophosphamide 600 mg/m²) q21d × 4
- Docetaxel 75 mg/m² q21d × 4
   Start: 13/03/2023 | End: 03/07/2023
   Duration: 16 weeks
   HER2 therapy: Not applicable
   Clinical response: Partial (tumor shrinkage ~50%)
   Adverse events: Grade 2 asthenia and mucositis
   Interval from CT to surgery: 4 weeks

**7. Surgical and Pathological Findings**
 Surgery: Right quadrantectomy + axillary dissection
 Residual tumor: 1.5 cm
 Histological grade: II
 Margins: Free, minimal 3 mm
 Lymph nodes: 12 dissected, 2 positive
 Lymphovascular invasion: Present
 Perineural invasion: Absent
 Pathologic response: Partial
 Final pathology: ypT1cN1aM0 — Stage IIA

**PATIENT 21 — SYNTHETIC RECORD FOR EXCLUSION**

**1. Identification**
 Synthetic ID: INC‑021
 Age: 61 years and 3 months
 Sex / Ethnicity: Female, White
 Weight / Height / BMI: 68 kg / 1.60 m / BMI 26.6 kg/m²
 Marital Status / Occupation: Divorced / Retired teacher
 Care Center: Instituto do Câncer do Ceará (ICC)
 Date of Diagnosis: 20/11/2022
 Date of Surgery: 10/04/2023
 Date of Enrollment/Randomization: 01/08/2023
 ECOG Performance Status: 1

**2. History of Present Illness (HPI)**
 Patient referred after abnormal mammography detected in October 2022.
 Core biopsy performed on 20/11/2022: Invasive ductal carcinoma, grade 3 (Nottingham).
 Initial staging: cT3N2M0 — Stage IIIA (AJCC 8th ed.)
 Immunohistochemistry:

- ER: 80%
- PR: 85%
- HER2: Negative
- Ki-67: 38%

**3. Personal and Family History**
 Comorbidities: Controlled hypertension and hypothyroidism
 Allergies: Penicillin (rash)
 Gynecologic history: G3P3A0; menopause at 50; no hormone replacement
 Family history: Sister with ovarian cancer (diagnosed at 48)
 Former smoker; quit 15 years ago; no alcohol use

**4. Physical Examination**
 General status: ECOG 1
 Left breast: Tumor approx. 5.3 cm in upper outer quadrant
 Skin without changes; no nipple discharge
 Multiple palpable axillary lymph nodes, largest 2.2 cm

**5. Laboratory and Imaging**
 CBC: Hb 12.3 g/dL; WBC 6,800/mm³; Platelets 277,000/mm³
 Renal function: Creatinine 0.91 mg/dL; Urea 39 mg/dL
 Liver enzymes: AST 29 U/L; ALT 26 U/L
 Beta-HCG: Negative
 LVEF (ECHO): Pre-CT 60%; Post-CT 58%
 Serologies: HBsAg, anti-HCV, HIV — all non-reactive
 Imaging: CT chest/abdomen/pelvis and bone scan — no metastases

**6. Neoadjuvant Treatment**
 Chemotherapy regimen:

- AC × 4 cycles (Doxorubicin 60 mg/m² + Cyclophosphamide 600 mg/m²)
- Paclitaxel 80 mg/m² weekly × 12 doses
   Start: 09/01/2023 | End: 24/04/2023
   Duration: 15.5 weeks
   HER2 therapy: Not applicable
   Clinical response: Partial (tumor reduction ~40%)
   Adverse events: G2 peripheral neuropathy (resolved), G1 nausea
   Interval to surgery: 2.5 weeks

**7. Surgical and Pathological Findings**
 Surgery: Modified radical mastectomy (left) + axillary dissection
 Residual tumor: 2.0 cm
 Histological grade: II
 Margins: Free, minimal 5 mm
 Lymph nodes: 14 dissected, 4 positive
 Lymphovascular invasion: Present
 Perineural invasion: Absent
 Pathologic response: Partial
 Final staging: ypT2N2aM0 — Stage IIIA

**PATIENT 22 — SYNTHETIC RECORD FOR EXCLUSION**

**1. Identification**
 Synthetic ID: INC‑022
 Age: 48 years and 7 months
 Sex / Ethnicity: Female, Brown
 Weight / Height / BMI: 73 kg / 1.65 m / BMI 26.8 kg/m²
 Marital Status / Occupation: Married / Civil servant
 Care Center: Hospital de Câncer de Barretos
 Date of Diagnosis: 03/12/2022
 Date of Surgery: 22/04/2023
 Date of Enrollment/Randomization: 18/08/2023
 ECOG Performance Status: 0

**2. History of Present Illness (HPI)**
 Patient presented with palpable mass in right breast noted in November 2022.
 Core needle biopsy performed on 03/12/2022: Invasive ductal carcinoma, grade 3 (Nottingham).
 Initial staging: cT2N2M0 — Stage IIIA (AJCC 8th ed.)
 Immunohistochemistry:

- ER: 90%
- PR: 70%
- HER2: Negative
- Ki-67: 40%

**3. Personal and Family History**
 Comorbidities: None reported
 Allergies: None known
 Gynecologic history: G2P2A0; still menstruating regularly
 Family history: Mother with breast cancer at age 63
 Never smoked; occasional alcohol use

**4. Physical Examination**
 General condition: ECOG 0
 Right breast: Mass ~3.5 cm, upper inner quadrant
 Skin and nipple: No retraction or ulceration
 Right axilla: Fixed lymph node ~2.1 cm

**5. Laboratory and Imaging**
 CBC: Hb 12.9 g/dL; WBC 7,500/mm³; Platelets 282,000/mm³
 Renal function: Creatinine 0.87 mg/dL; Urea 34 mg/dL
 Liver function: AST 27 U/L; ALT 25 U/L
 Beta-HCG: Negative
 LVEF (ECHO): Pre-CT 62%; Post-CT 60%
 Serologies: HBsAg, anti-HCV, HIV — all negative
 Imaging: CT chest/abdomen/pelvis and bone scan — no metastasis

**6. Neoadjuvant Treatment**
 Chemotherapy:

- EC × 4 cycles (Epirubicin 90 mg/m² + Cyclophosphamide 600 mg/m²)
- Weekly paclitaxel × 12 cycles (80 mg/m²)
   Start: 09/01/2023 | End: 24/04/2023
   Duration: 16 weeks
   HER2 therapy: Not applicable
   Response: Partial response (~50% reduction in tumor size)
   Adverse events: G1 fatigue, G2 alopecia
   Interval to surgery: 4 weeks

**7. Surgical and Pathological Findings**
 Surgery: Right quadrantectomy + axillary dissection
 Residual tumor: 1.4 cm
 Histological grade: II
 Margins: Free, minimum 3 mm
 Lymph nodes: 15 removed, 3 positive
 Lymphovascular invasion: Present
 Perineural invasion: Not observed
 Pathologic response: Partial
 Final staging: ypT1cN2aM0 — Stage IIIA

**PATIENT 23 — SYNTHETIC RECORD FOR EXCLUSION**

**1. Identification**
 Synthetic ID: INC‑023
 Age: 55 years and 2 months
 Sex / Ethnicity: Female, White
 Weight / Height / BMI: 68 kg / 1.60 m / BMI 26.6 kg/m²
 Marital Status / Occupation: Divorced / Schoolteacher
 Care Center: Instituto Nacional de Câncer (INCA)
 Date of Diagnosis: 14/02/2023
 Date of Surgery: 10/07/2023
 Date of Enrollment/Randomization: 28/10/2023
 ECOG Performance Status: 1

**2. History of Present Illness (HPI)**
 Lesion detected via screening mammogram in February 2023.
 Ultrasound and core biopsy performed on 14/02/2023: Invasive ductal carcinoma, grade 3 (Nottingham).
 Initial staging: cT2N1M0 — Stage IIB (AJCC 8th ed.)
 Immunohistochemistry:

- ER: 95%
- PR: 50%
- HER2: Negative
- Ki-67: 35% (central pathology confirmed)

**3. Personal and Family History**
 Comorbidities: Controlled hypertension and type 2 diabetes
 Allergies: None reported
 Gynecologic history: G3P3A0; menopause at 49 years; no hormonal replacement therapy
 Family history: Sister with breast cancer at 52 years
 No smoking or alcohol

**4. Physical Examination**
 General condition: ECOG 1
 Left breast: Palpable mass ~2.8 cm, upper outer quadrant
 No nipple discharge or skin changes
 Left axilla: Palpable mobile lymph node ~1.5 cm

**5. Laboratory and Imaging**
 CBC: Hb 13.2 g/dL; WBC 6,700/mm³; Platelets 265,000/mm³
 Renal function: Creatinine 0.92 mg/dL
 Liver function: AST 31 U/L; ALT 29 U/L
 Beta-HCG: Negative
 LVEF (ECHO): Pre-CT 64%; Post-CT 61%
 Serologies: HBsAg, anti-HCV, HIV — all negative
 Imaging: CT chest/abdomen/pelvis and bone scan — no evidence of metastasis

**6. Neoadjuvant Treatment**
 Chemotherapy:

- AC × 4 cycles (Doxorubicin 60 mg/m² + Cyclophosphamide 600 mg/m²)
- Docetaxel × 4 cycles (75 mg/m² every 3 weeks)
   Start: 06/03/2023 | End: 26/06/2023
   Duration: 16 weeks
   Trastuzumab: Not used (HER2 negative)
   Response: Partial tumor reduction (~40%)
   Adverse events: G2 neutropenia, managed without hospitalization
   Interval to surgery: 2 weeks

**7. Surgical and Pathological Findings**
 Surgery: Left quadrantectomy + sentinel lymph node biopsy and axillary dissection
 Residual tumor: 1.1 cm
 Histological grade: II
 Margins: Free (4 mm)
 Lymph nodes: 11 removed, 2 positive
 Lymphovascular invasion: Present
 Perineural invasion: Not detected
 Pathologic response: Partial
 Final staging: ypT1cN1aM0 — Stage IIB

**PATIENT 24 — SYNTHETIC RECORD FOR EXCLUSION**

**1. Identification**
 Synthetic ID: INC‑024
 Age: 49 years and 10 months
 Sex / Ethnicity: Female, Mixed-race
 Weight / Height / BMI: 72 kg / 1.63 m / BMI 27.1 kg/m²
 Marital Status / Occupation: Married / Administrative assistant
 Care Center: Hospital de Câncer de Barretos
 Date of Diagnosis: 06/03/2023
 Date of Surgery: 21/08/2023
 Date of Enrollment/Randomization: 02/12/2023
 ECOG Performance Status: 0

**2. History of Present Illness (HPI)**
 Detected by self-examination in late February 2023.
 Core biopsy on 06/03/2023: Invasive ductal carcinoma, grade 3 (Nottingham).
 Initial clinical stage: cT3N2M0 — Stage IIIA (AJCC 8th ed.)
 Immunohistochemistry:

- ER: 98%
- PR: 70%
- HER2: Negative
- Ki-67: 42% (confirmed by central analysis)

**3. Personal and Family History**
 Comorbidities: Dislipidemia, managed with statins
 Allergies: No known drug allergies
 Gynecologic history: G2P2A0; menopause at 47 years; no hormonal therapy
 Family history: Mother with breast cancer at age 64
 No tobacco use; occasional alcohol use

**4. Physical Examination**
 Overall status: ECOG 0
 Right breast: Palpable mass in upper outer quadrant ~4.5 cm
 No skin changes or nipple discharge
 Right axilla: Several mobile lymph nodes (largest ~2.0 cm)

**5. Laboratory and Imaging**
 CBC: Hb 12.8 g/dL; WBC 7,100/mm³; Platelets 280,000/mm³
 Renal function: Creatinine 0.88 mg/dL
 Liver enzymes: AST 26 U/L; ALT 24 U/L
 Beta-HCG: Negative
 LVEF (ECHO): Pre-CT 67%; Post-CT 65% (variation <15 p.p.)
 Serologies: HBsAg, anti-HCV, HIV — all negative
 CT chest/abdomen/pelvis + bone scan — no distant metastasis

**6. Neoadjuvant Treatment**
 Chemotherapy regimen:

- AC × 4 cycles (Doxorubicin 60 mg/m² + Cyclophosphamide 600 mg/m²)
- Followed by Paclitaxel weekly × 12 (80 mg/m²/week)
   Start date: 20/03/2023
   End date: 10/07/2023
   Duration: 16 weeks
   Trastuzumab: Not applicable (HER2-negative)
   Clinical response: Tumor volume reduction ~60%
   Adverse events: Grade 1 fatigue, grade 2 alopecia
   Interval from last chemo to surgery: 6 weeks

**7. Surgical and Pathological Findings**
 Surgery: Right quadrantectomy + axillary lymph node dissection
 Residual tumor: 1.4 cm
 Histological grade: II
 Margins: Free (5 mm)
 Lymph nodes: 15 removed, 4 positive
 Lymphovascular invasion: Present
 Perineural invasion: Not identified
 Pathologic response: Partial
 Final staging: ypT1cN2aM0 — Stage IIIA

**PATIENT 25 — SYNTHETIC RECORD FOR EXCLUSION**

**1. Identification**
 Synthetic ID: INC‑025
 Age: 58 years and 2 months
 Sex / Ethnicity: Female, White
 Weight / Height / BMI: 80 kg / 1.60 m / BMI 31.3 kg/m²
 Marital Status / Occupation: Widow / School teacher
 Care Center: Instituto Nacional de Câncer (INCA)
 Date of Diagnosis: 02/02/2023
 Date of Surgery: 05/06/2023
 Date of Enrollment/Randomization: 10/09/2023
 ECOG Performance Status: 1

**2. History of Present Illness (HPI)**
 Detected on routine screening mammography in January 2023.
 Core biopsy on 02/02/2023: Invasive ductal carcinoma, grade 3 (Nottingham).
 Initial clinical stage: cT2N1M0 — Stage IIB (AJCC 8th ed.)
 Immunohistochemistry:

- ER: 95%
- PR: 40%
- HER2: Negative
- Ki-67: 38% (confirmed centrally)

**3. Personal and Family History**
 Comorbidities: Type 2 diabetes mellitus and hypertension, both controlled
 Allergies: None reported
 Gynecologic history: G3P3A0; natural menopause at 51; no hormone therapy
 Family history: No relevant oncologic history
 No smoking; no alcohol consumption

**4. Physical Examination**
 General condition: ECOG 1
 Left breast: Mass ~3.5 cm in upper inner quadrant
 No skin retraction, ulceration, or nipple discharge
 Left axilla: Single mobile lymph node ~1.5 cm

**5. Laboratory and Imaging**
 CBC: Hb 13.1 g/dL; WBC 6,500/mm³; Platelets 265,000/mm³
 Renal function: Creatinine 0.92 mg/dL
 Liver function: AST 22 U/L; ALT 25 U/L
 Beta-HCG: Negative
 LVEF (ECHO): Pre-CT 64%; Post-CT 60% — variation <15 p.p.
 Serologies: HBV, HCV, HIV — negative
 CT scans and bone scintigraphy: No metastatic disease

**6. Neoadjuvant Treatment**
 Chemotherapy regimen:

- EC × 4 cycles (Epirubicin 90 mg/m² + Cyclophosphamide 600 mg/m²)
- Followed by weekly Paclitaxel × 12 (80 mg/m²/week)
   Start: 01/03/2023
   End: 20/06/2023
   Duration: 16 weeks
   Trastuzumab: Not applicable (HER2-negative)
   Clinical response: Partial (tumor shrinkage ~55%)
   Adverse events: Grade 2 peripheral neuropathy, Grade 1 nausea
   Interval between last cycle and surgery: 2 weeks

**7. Surgical and Pathological Findings**
 Surgery: Left mastectomy + axillary lymphadenectomy
 Residual tumor: 1.9 cm
 Histologic grade: II
 Margins: Free (≥ 6 mm)
 Lymph nodes: 12 removed, 2 positive
 Lymphovascular invasion: Present
 Perineural invasion: Absent
 Pathologic response: Partial
 Final staging: ypT1cN1aM0 — Stage IIB

**FINAL PROMPT FOR PATIENT SENTENCE GENERATION (P2S)**

**CONTEXT:** You are a specialist in Natural Language Processing (NLP) and information extraction, focusing on oncology data.

**GOAL:** Process an electronic medical record to generate a "patient sentence" (P2S). The goal of this sentence is to create a concise, structured clinical “fingerprint” that accelerates initial screening for clinical trials. The sentence should be a factual and chronological representation of the patient’s positive clinical data.

**INPUT:** A patient’s electronic medical record file.

**OUTPUT FORMAT:** A single text string, with each clinical detail separated by the character |.

**EXTRACTION AND FORMATTING RULES:**

**Mandatory Start:** The sentence MUST begin with the location of the primary tumor.

**Chronological Order:** Events must be listed from oldest to most recent.

**Date Format:** Each event or report MUST include its date in the format (DD/MM/YYYY).

**"Key: Value" Structure:** Use a Key: Value format for all information.

**Variable Sentence Length, Focus on Positive Data:** The sentence will vary in length. Include only positive findings and data. Do NOT include negative results (e.g., negative genetic tests), minor comorbidities, or incidental findings.

**Maximum Detail for Reports:**

- **Pathology:** Extract all details: histological type, grade, tumor size (Tu), margin status, angiolymphatic and perineural invasion.
- **Immunohistochemistry (IHC):** Extract exact percentages for ER, PR, and Ki-67, and HER2 score.
- **Genetics:** Transcribe the full results of genetic tests (e.g., BRCA1/2, MLPA).

**Handling Dynamic Data:**

- **Clinical Data (e.g., ECOG status):** Include ONLY the most recent record.
- **Tumor Markers (e.g., CEA, CA 15-3):** Include the baseline AND most recent value. Use the format: “Marker: Initial_Value (DD/MM/YYYY) -> Recent_Value (DD/MM/YYYY)”.

**Essential Information List (Extract if present and positive):**

- **Diagnosis:** Histological type, grade, size, invasions (angiolymphatic/perineural), etc. (with date).
- **Pathological Staging:** pTNM (with date).
- **IHC:** ER, PR, HER2, Ki-67 (with date).
- **Genetic Tests:** Test result (with date).
- **Surgery(ies):** Procedure type, margin status (with date).
- **Systemic Treatments (CT, HT, etc.):** Protocol/medication and period (start and end).
- **Radiotherapy (RT):** Period (start and end).
- **Performance Status:** Latest ECOG (with date).
- **Tumor Markers:** First and last value (with dates).
- **Major Comorbidities:** Only those listed as significant prior conditions (e.g., Hypertension, Diabetes).

**FINAL OUTPUT EXAMPLE:**

*Right Breast | Diagnosis: Invasive ductal carcinoma NST, Histologic grade II, Size: 2.0 cm, Margins: Clear, Angiolymphatic/peritumoral embolization: Not detected (26/08/2019) | Pathological Staging: pT1c pN1 pMx (26/08/2019) | IHC: ER positive 100%, PR positive 100%, HER2 score 0, Ki67 positive 10% (18/09/2019) | Adjuvant CT: TC protocol (Start: NOV/2019 - End: 16/01/2020) | Adjuvant RT: (Start: 17/02/2020 - End: 10/03/2020) | Adjuvant HT: Anastrozole (Start: MAR/2020) | Bone Therapy: Zoledronic Acid every 6 months (Start: MAR/2020) | Comorbidities: Systemic arterial hypertension | Tumor Marker CA15-3: 15.7 U/ml (31/10/2019) -> 16.7 U/ml (05/01/2024) | Tumor Marker CEA: 1.6 ng/ml (31/10/2019) | ECOG: 0 (22/04/2024)*

**SYNTHESIS OF SYNTHETIC PATIENTS - MONARCHE**

### **PATIENT 1 — SYNTHETIC INCLUSION RECORD**

**Left Breast** | **Diagnosis:** Invasive ductal carcinoma, Histological grade III (Nottingham 8/9), Tumor size: 5.2 cm, Lymphovascular invasion present, Perineural invasion absent (05/17/2023) | **Initial clinical staging:** cT3N2M0 – Stage IIIA (AJCC 8th edition) (05/17/2023) | **IHC:** ER 95%, PR 80%, HER2 0, Ki-67 35% (05/17/2023) | **Neoadjuvant chemotherapy (NAC):** Doxorubicin 60 mg/m² + cyclophosphamide 600 mg/m² (AC) followed by weekly paclitaxel 80 mg/m² (Start: 06/05/2023 – End: 09/20/2023) | **Clinical response:** Approx. 50% tumor reduction | **Adverse events:** Grade 2 neutropenia, grade 1 nausea, grade 2 alopecia | **Surgery:** Modified radical mastectomy + axillary lymph node dissection levels I–II (10/02/2023) | **Pathology:** Residual tumor 1.2 cm, Grade II, 21 lymph nodes (5 positive), Clear margins ≥ 3 mm, Lymphovascular invasion present, Perineural invasion absent, Partial pathological response (10/02/2023) | **Pathological staging:** ypT1cN2aM0 – Stage IIIA (10/02/2023) | **ECOG:** 0 (01/15/2024) | **Comorbidities:** Controlled systemic arterial hypertension

### **PATIENT 2 — SYNTHETIC INCLUSION RECORD**

**Right Breast** | **Diagnosis:** Invasive lobular carcinoma, Histological grade III (Nottingham 8/9), Tumor size: 4.8 cm, Lymphovascular invasion present (08/12/2023) | **Initial clinical staging:** cT2N2M0 – Stage IIIA (AJCC 8th edition) (08/12/2023) | **IHC:** ER 90%, PR 60%, HER2 1+, Ki-67 30% (08/12/2023) | **NAC:** Docetaxel 75 mg/m² + cyclophosphamide 600 mg/m² (TC), 6 cycles every 21 days (Start: 09/02/2023 – End: 12/15/2023) | **Clinical response:** Approx. 45% tumor reduction | **Adverse events:** Grade 1 fatigue, grade 2 alopecia, grade 2 neutropenia | **Surgery:** Right quadrantectomy + axillary lymph node dissection levels I–II (12/30/2023) | **Pathology:** Residual tumor 1.0 cm, Grade II, 14 lymph nodes (4 positive), Clear margins ≥ 2 mm, Lymphovascular invasion present, Partial pathological response (12/30/2023) | **Pathological staging:** ypT1bN2aM0 – Stage IIIA (12/30/2023) | **ECOG:** 1 (04/10/2024) | **Comorbidities:** Controlled dyslipidemia

### **PATIENT 3 — SYNTHETIC INCLUSION RECORD**

**Left Breast** | **Diagnosis:** Invasive ductal carcinoma, Histological grade III (Nottingham 8/9), Tumor size: 5.6 cm, Lymphovascular invasion present, Perineural invasion absent (04/19/2023) | **Initial clinical staging:** cT3N2M0 – Stage IIIA (AJCC 8th edition) (04/19/2023) | **IHC:** ER 95%, PR 90%, HER2 0, Ki-67 28% (04/19/2023) | **NAC:** Doxorubicin 60 mg/m² + cyclophosphamide 600 mg/m² (AC) — 4 cycles, followed by weekly paclitaxel 80 mg/m² — 12 applications (Start: 05/10/2023 – End: 08/18/2023) | **Clinical response:** Approx. 60% tumor reduction | **Adverse events:** Grade 1 nausea, grade 1 peripheral neuropathy, grade 2 neutropenia (afebrile) | **Surgery:** Modified radical mastectomy + axillary lymph node dissection levels I–II (09/02/2023) | **Pathology:** Residual tumor 1.5 cm, Grade II, 23 lymph nodes (6 positive), Clear margins ≥ 4 mm, Lymphovascular invasion present, Perineural invasion absent, Partial pathological response (09/02/2023) | **Pathological staging:** ypT1cN2bM0 – Stage IIIA (09/02/2023) | **ECOG:** 0 (12/20/2023) | **Comorbidities:** Controlled systemic arterial hypertension and type 2 diabetes mellitus

### **PATIENT 4 — SYNTHETIC INCLUSION RECORD**

**Right Breast** | **Diagnosis:** Invasive ductal carcinoma, Histological grade III (Nottingham 9/9), Tumor size: 5.4 cm, Lymphovascular invasion present, Perineural invasion absent (03/03/2023) | **Initial clinical staging:** cT3N1M0 – Stage IIIA (AJCC 8th edition) (03/03/2023) | **IHC:** ER 98%, PR 85%, HER2 0, Ki-67 32% (03/03/2023) | **NAC:** Docetaxel 75 mg/m² + cyclophosphamide 600 mg/m² (TC), 6 cycles every 21 days (Start: 03/20/2023 – End: 07/03/2023) | **Clinical response:** Approx. 55% tumor reduction | **Adverse events:** Grade 1 fatigue, grade 1 nausea, grade 2 alopecia, grade 2 neutropenia | **Surgery:** Modified radical mastectomy + axillary lymph node dissection levels I–II (07/10/2023) | **Pathology:** Residual tumor 1.4 cm, Grade II, 22 lymph nodes (5 positive), Clear margins ≥ 3 mm, Lymphovascular invasion present, Perineural invasion absent, Partial pathological response (07/10/2023) | **Pathological staging:** ypT1cN2aM0 – Stage IIIA (07/10/2023) | **ECOG:** 0 (11/01/2023) | **Comorbidities:** Controlled systemic hypertension and class I obesity

### **PATIENT 5 — SYNTHETIC INCLUSION RECORD**

**Left Breast** | **Diagnosis:** Invasive ductal carcinoma, Histological grade III (Nottingham 9/9), Tumor size: 4.9 cm, Lymphovascular invasion present, Perineural invasion absent (09/14/2023) | **Initial clinical staging:** cT3N2M0 – Stage IIIA (AJCC 8th edition) (09/14/2023) | **IHC:** ER 95%, PR 70%, HER2 1+, Ki-67 40% (09/14/2023) | **NAC:** Doxorubicin 60 mg/m² + cyclophosphamide 600 mg/m² (AC) — 4 cycles, followed by weekly paclitaxel 80 mg/m² — 12 weeks (Start: 09/25/2023 – End: 01/05/2024) | **Clinical response:** Approx. 50% tumor reduction | **Adverse events:** Grade 1 nausea, grade 1 peripheral neuropathy, grade 2 alopecia, grade 2 neutropenia (afebrile) | **Surgery:** Left quadrantectomy + axillary lymph node dissection levels I–II (01/22/2024) | **Pathology:** Residual tumor 1.3 cm, Grade II, 18 lymph nodes (4 positive), Clear margins ≥ 2 mm, Lymphovascular invasion present, Perineural invasion absent, Partial pathological response (01/22/2024) | **Pathological staging:** ypT1cN2aM0 – Stage IIIA (01/22/2024) | **ECOG:** 1 (05/10/2024) | **Comorbidities:** Controlled dyslipidemia and hypothyroidism

### **PATIENT 6 — SYNTHETIC EXCLUSION RECORD**

**Right Breast** | **Diagnosis:** Invasive ductal carcinoma, Histologic grade II (Nottingham 7/9), Size: 3.1 cm, No lymphovascular invasion (06/05/2023) | **Initial clinical staging:** cT2N0M0 – Stage IIA (AJCC 8th edition) (06/05/2023) | **IHC:** ER 0%, PR 0%, HER2 3+ (FISH positive), Ki-67 45% (06/05/2023) | **Neoadjuvant chemotherapy:** TCHP regimen — docetaxel + carboplatin + trastuzumab + pertuzumab (Start: 25/05/2023 – End: 25/08/2023) | **Clinical response:** >70% tumor reduction (significant partial response) | **Adverse events:** Grade 3 neutropenia, grade 2 diarrhea, grade 2 alopecia | **Surgery:** Right quadrantectomy + level I–II axillary lymphadenectomy (10/09/2023) | **Pathology:** Residual tumor 0.5 cm, Grade II, 11 lymph nodes (0 positive), Clear margins ≥ 5 mm, No lymphovascular invasion, Near-complete pathological response (ypT1aN0M0) (10/09/2023) | **ECOG:** 1 (18/01/2024) | **Comorbidities:** Mild hypertension

### **PATIENT 7 — SYNTHETIC EXCLUSION RECORD**

**Left Breast** | **Diagnosis:** Invasive ductal carcinoma, Histologic grade II (Nottingham 7/9), Size: 2.8 cm, No lymphovascular invasion (02/06/2023) | **Initial clinical staging:** cT2N0M0 – Stage IIA (AJCC 8th edition) (02/06/2023) | **IHC:** ER 95%, PR 80%, HER2 1+, Ki-67 15% (02/06/2023) | **Treatment:** Primary surgery – left quadrantectomy + level I axillary lymphadenectomy (sentinel) (10/10/2023) | **Pathology:** Residual tumor 2.6 cm, Grade II, 4 lymph nodes (0 positive), Clear margins ≥ 3 mm, No lymphovascular invasion (10/10/2023) | **Adjuvant radiotherapy:** Performed (postoperative start – dates not specified) | **Planned treatment:** Adjuvant hormone therapy indicated | **ECOG:** 0 (15/02/2024) | **Comorbidities:** Mild hypertension and osteopenia

### **PATIENT 8 — SYNTHETIC EXCLUSION RECORD**

**Left Breast** | **Diagnosis:** Invasive ductal carcinoma, Histologic grade III (Nottingham 8/9), Size: 3.8 cm, Lymphovascular invasion present (18/07/2023) | **Initial clinical staging:** cT2N1M1 – Stage IV (AJCC 8th edition) (18/07/2023) | **IHC:** ER 95%, PR 70%, HER2 0, Ki-67 30% (18/07/2023) | **Systemic treatment:** Letrozole 2.5 mg/day + palbociclib 125 mg/day (Start: 10/01/2024 – ongoing, stable disease) | **Adverse events:** Grade 2 fatigue, grade 3 neutropenia (temporary dose reduction) | **Surgery:** Left palliative mastectomy without reconstruction (15/12/2023) | **Pathology:** Residual tumor 3.5 cm, Grade III, 10 lymph nodes (1 positive), Clear margins ≥ 3 mm, Lymphovascular invasion present (15/12/2023) | **Final staging:** pT2N1M1 – Stage IV (15/12/2023) | **ECOG:** 2 (22/03/2024) | **Comorbidities:** Systemic arterial hypertension, type 2 diabetes mellitus, and knee osteoarthritis

### **PATIENT 9 — SYNTHETIC EXCLUSION RECORD**

**Right Breast** | **Diagnosis:** Invasive ductal carcinoma, Histologic grade II (Nottingham 7/9), Size: 2.7 cm, No lymphovascular invasion (20/04/2024) | **Initial clinical staging:** cT2N0M0 – Stage IIA (AJCC 8th edition) (20/04/2024) | **IHC:** ER 95%, PR 90%, HER2 0, Ki-67 25% (20/04/2024) | **Surgery:** Right quadrantectomy + sentinel lymph node biopsy (5 nodes negative) (05/08/2024) | **Pathology:** Residual tumor 2.6 cm, Grade II, Clear margins ≥ 3 mm, No lymphovascular invasion (05/08/2024) | **Planned adjuvant treatment:** Hormone therapy (anastrozole) – suspended after eligibility reassessment (10/12/2024) | **Oncological history:** Ductal carcinoma in situ (DCIS) in the left breast treated with conservative surgery and radiotherapy (2021) | **ECOG:** 0 (10/12/2024) | **Comorbidities:** Mild dyslipidemia | **Current use:** Transdermal estradiol 1 mg/day for hormone replacement therapy

### **PATIENT 10 — SYNTHETIC EXCLUSION RECORD**

**Left Breast** | **Diagnosis:** Invasive ductal carcinoma, Histologic grade III (Nottingham 8/9), Size: 3.6 cm, Lymphovascular invasion present (11/03/2023) | **Initial clinical staging:** cT2N1M0 – Stage IIB (AJCC 8th edition) (11/03/2023) | **IHC:** ER 90%, PR 60%, HER2 1+, Ki-67 35% (11/03/2023) | **Experimental neoadjuvant chemotherapy:** Letrozole 2.5 mg/day + palbociclib 125 mg/day (CDK4/6 inhibitor) – 4 cycles (Start: 01/04/2023 – End: 25/07/2023) | **Clinical response:** Approx. 40% tumor reduction | **Adverse events:** Grade 3 neutropenia, grade 2 fatigue, decline in left ventricular ejection fraction (LVEF 64% → 49%, grade 2 cardiotoxicity) | **Surgery:** Left quadrantectomy + level I–II axillary lymphadenectomy (25/08/2023) | **Pathology:** Residual tumor 1.8 cm, Grade II, 14 lymph nodes (3 positive), Clear margins ≥ 3 mm, Lymphovascular invasion present, Partial pathological response (25/08/2023) | **Final staging:** ypT1cN1aM0 – Stage IIB (25/08/2023) | **ECOG:** 1 (10/01/2024) | **Comorbidities:** Systemic arterial hypertension and dyslipidemia

### **PATIENT 11 — SYNTHETIC EXCLUSION RECORD**

**Right Breast** | **Diagnosis:** Invasive ductal carcinoma, Histologic grade III (Nottingham 9/9), with extensive lymphovascular invasion (18/01/2024) | **Initial clinical staging:** cT4dN2M0 – Inflammatory breast carcinoma, Stage IIIB (AJCC 8th edition) (18/01/2024) | **IHC:** ER 80%, PR 60%, HER2 0, Ki‑67 40% (18/01/2024) | **Neoadjuvant chemotherapy planned:** Doxorubicin + cyclophosphamide (2 cycles conducted until 25/04/2024) | **Clinical response:** Partial – erythema reduced, skin thickening persisted | **Surgical status:** No curative surgery performed (locally advanced disease) | **ECOG:** 1 (25/04/2024) | **Comorbidities:** Controlled systemic arterial hypertension

### **PATIENT 12 — SYNTHETIC EXCLUSION RECORD**

**Left Breast** | **Diagnosis:** Invasive ductal carcinoma, Histologic grade III (Nottingham 8/9), Size: 4.6 cm, Lymphovascular invasion present (22/08/2023) | **Initial clinical staging:** cT2N1M0 – Stage IIB (AJCC 8th edition) (22/08/2023) | **IHC:** ER 90%, PR 85%, HER2 1+, Ki-67 25% (22/08/2023) | **Surgery:** Modified radical mastectomy + level I–II axillary lymphadenectomy (12/01/2024) | **Pathology:** Residual tumor 1.7 cm, Histologic grade II, 16 lymph nodes (3 positive), Margins ≥ 3 mm, Lymphovascular invasion present (12/01/2024) | **Final staging:** pT1cN1aM0 – Stage IIB (12/01/2024) | **ECOG:** 1 (25/04/2024) | **Comorbidities:** Hypertension and active chronic hepatitis B with detectable viral load | **Lab tests:** AST 62 U/L, ALT 71 U/L (>2× ULN), HBV DNA 2.4 × 10⁵ IU/mL

### **PATIENT 13 — SYNTHETIC EXCLUSION RECORD**

**Right Breast** | **Diagnosis:** Invasive ductal carcinoma, Histologic grade II (Nottingham 7/9), Size: 3.2 cm, No lymphovascular invasion (09/05/2024) | **Initial clinical staging:** cT2N1M0 – Stage IIB (AJCC 8th edition) (09/05/2024) | **IHC:** ER 95%, PR 70%, HER2 0, Ki-67 22% (09/05/2024) | **Surgery:** Right quadrantectomy + sentinel lymph node biopsy (2 negative nodes) (18/09/2024) | **Pathology:** Residual tumor 3.0 cm, Grade II, Margins ≥ 3 mm, No lymphovascular invasion (18/09/2024) | **Final staging:** pT2N0M0 – Stage IIA (18/09/2024) | **ECOG:** 0 (10/12/2024) | **Comorbidities:** None relevant | **Associated condition:** 9-week pregnancy confirmed (Beta-HCG 62,000 mIU/mL)

### **PATIENT 14 — SYNTHETIC EXCLUSION RECORD**

**Right Breast** | **Diagnosis:** Invasive ductal carcinoma, Histologic grade III (Nottingham 9/9), Size: 4.2 cm, Lymphovascular invasion present (20/06/2023) | **Initial clinical staging:** cT2N1M0 – Stage IIB (AJCC 8th edition) (20/06/2023) | **IHC:** ER 90%, PR 85%, HER2 1+, Ki-67 35% (20/06/2023) | **Surgery:** Right quadrantectomy + level I–II axillary lymphadenectomy (14/11/2023) | **Pathology:** Residual tumor 1.6 cm, Grade II, 15 lymph nodes (3 positive), Margins ≥ 3 mm, Lymphovascular invasion present (14/11/2023) | **Final staging:** pT1cN1aM0 – Stage IIB (14/11/2023) | **ECOG:** 1 (10/02/2024) | **Comorbidities:** Hypertension, dyslipidemia, idiopathic ventricular tachycardia on beta-blockers | **Relevant finding:** Self-limited episodes of non-sustained ventricular tachycardia (24h Holter)

### **PATIENT 15 — SYNTHETIC EXCLUSION RECORD**

**Left Breast** | **Diagnosis:** Invasive ductal carcinoma, Histologic grade III (Nottingham 9/9), Size: 4.3 cm, Lymphovascular invasion present (08/05/2023) | **Initial clinical staging:** cT2N2M0 – Stage IIIA (AJCC 8th edition) (08/05/2023) | **IHC:** ER 95%, PR 80%, HER2 1+, Ki-67 30% (08/05/2023) | **Surgery:** Modified radical mastectomy + level I–II axillary lymphadenectomy (10/10/2023) | **Pathology:** Residual tumor 1.5 cm, Grade II, 20 lymph nodes (5 positive), Margins ≥ 3 mm, Lymphovascular invasion present (10/10/2023) | **Final staging:** pT1cN2aM0 – Stage IIIA (10/10/2023) | **ECOG:** 1 (22/01/2024) | **Comorbidities:** Hypertension, obesity, previous deep vein thrombosis (DVT) under continuous anticoagulation (rivaroxaban 20 mg/day)

### **PATIENT 16 — SYNTHETIC EXCLUSION RECORD**

**Right Breast** | **Diagnosis:** Invasive ductal carcinoma, Histologic grade III (Nottingham 9/9), Size: 3.9 cm, Lymphovascular invasion present (18/09/2023) | **Initial clinical staging:** cT2N1M0 – Stage IIB (AJCC 8th edition) (18/09/2023) | **IHC:** ER 90%, PR 80%, HER2 0, Ki-67 30% (18/09/2023) | **Surgery:** Right quadrantectomy + level I–II axillary lymphadenectomy (05/02/2024) | **Pathology:** Residual tumor 1.8 cm, Grade II, 12 lymph nodes (3 positive), Margins ≥ 3 mm, Lymphovascular invasion present (05/02/2024) | **Final staging:** pT1cN1aM0 – Stage IIB (05/02/2024) | **ECOG:** 0 (10/03/2024) | **Comorbidities:** Mild intermittent asthma (occasional bronchodilator use) | **Recent history:** Previous participation in an adjuvant immunotherapy trial for cutaneous melanoma (ended 25 days ago)

### **PATIENT 17 — SYNTHETIC EXCLUSION RECORD**

**Left Breast** | **Diagnosis:** Invasive ductal carcinoma, Histologic grade III (Nottingham 8/9), Size: 3.7 cm, Lymphovascular invasion present (12/04/2023) | **Initial clinical staging:** cT2N1M0 – Stage IIB (AJCC 8th edition) (12/04/2023) | **IHC:** ER 90%, PR 75%, HER2 0, Ki-67 25% (12/04/2023) | **Surgery:** Left quadrantectomy + level I–II axillary lymphadenectomy (30/09/2023) | **Pathology:** Residual tumor 1.8 cm, Grade II, 14 lymph nodes (3 positive), Margins ≥ 3 mm, Lymphovascular invasion present (30/09/2023) | **Final staging:** pT1cN1aM0 – Stage IIB (30/09/2023) | **ECOG:** 1 (10/02/2024) | **Comorbidities:** Hypertension, type 2 diabetes mellitus, chronic kidney disease stage 3B (CrCl ≈ 45 mL/min)

### **PATIENT 18 — SYNTHETIC EXCLUSION RECORD**

**Right Breast** | **Diagnosis:** Invasive ductal carcinoma, Histologic grade III (Nottingham 8/9), Size: 3.4 cm, Lymphovascular invasion present (11/07/2023) | **Initial clinical staging:** cT2N1M0 – Stage IIB (AJCC 8th edition) (11/07/2023) | **IHC:** ER 95%, PR 70%, HER2 0, Ki-67 30% (11/07/2023) | **Surgery:** Right quadrantectomy + level I–II axillary lymphadenectomy (15/12/2023) | **Pathology:** Residual tumor 1.7 cm, Grade II, 13 lymph nodes (3 positive), Margins ≥ 3 mm, Lymphovascular invasion present (15/12/2023) | **Final staging:** pT1cN1aM0 – Stage IIB (15/12/2023) | **ECOG:** 1 (10/04/2024) | **Comorbidities:** Hypertension and history of papillary thyroid carcinoma treated in 2021, with residual biochemical disease (thyroglobulin 2.1 ng/mL) under endocrinology follow-up

### **PATIENT 19 — SYNTHETIC EXCLUSION RECORD**

**Bilateral Breasts** | **Diagnosis:** Invasive lobular carcinoma, Histologic grade II (Nottingham 7/9), Size: 3.4 cm (Left), 2.8 cm (Right), No lymphovascular invasion (21/03/2023) | **Initial clinical staging:** cT2N0M0 bilateral – Stage IIA (AJCC 8th edition) (21/03/2023) | **IHC:** ER 95%, PR 90%, HER2 0, Ki-67 18% (21/03/2023) | **Surgery:** Bilateral mastectomy + bilateral sentinel lymph node biopsy (10/09/2023) | **Pathology:** Residual tumor 3.4 cm (L), 2.7 cm (R), Grade II, 4 nodes (all negative), Margins ≥ 3 mm, No lymphovascular invasion (10/09/2023) | **Final staging:** pT2N0M0 bilateral – Stage IIA (10/09/2023) | **ECOG:** 1 (20/01/2024) | **Comorbidities:** Hypertension and knee osteoarthritis

### **PATIENT 20 — SYNTHETIC EXCLUSION RECORD**

**Left Breast** | **Diagnosis:** Invasive ductal carcinoma, Histologic grade III (Nottingham 8/9), Size: 3.9 cm, Lymphovascular invasion present (05/08/2023) | **Initial clinical staging:** cT2N1M0 – Stage IIB (AJCC 8th edition) (05/08/2023) | **IHC:** ER 90%, PR 75%, HER2 0, Ki-67 28% (05/08/2023) | **Surgery:** Left quadrantectomy + level I–II axillary lymphadenectomy (22/12/2023) | **Pathology:** Residual tumor 2.1 cm, Grade II, 15 lymph nodes (2 positive), Margins ≥ 3 mm, Lymphovascular invasion present (22/12/2023) | **Final staging:** pT2N1aM0 – Stage IIB (22/12/2023) | **ECOG:** 1 (20/03/2024) | **Comorbidities:** Hypertension and active pulmonary tuberculosis under RHZE treatment (rifampin, isoniazid, pyrazinamide, ethambutol)

### **PATIENT 21 — SYNTHETIC EXCLUSION RECORD**

**Right Breast** | **Diagnosis:** Invasive ductal carcinoma, Histologic grade II (Nottingham 7/9), Size: 3.1 cm, Lymphovascular invasion present (10/08/2023) | **Initial clinical staging:** cT2N1M0 – Stage IIB (AJCC 8th edition) (10/08/2023) | **IHC:** ER 95%, PR 80%, HER2 0, Ki-67 24% (10/08/2023) | **Surgery:** Right quadrantectomy + level I–II axillary lymphadenectomy (18/01/2024) | **Pathology:** Residual tumor 2.7 cm, Grade II, 14 lymph nodes (3 positive), Margins ≥ 3 mm, Lymphovascular invasion present (18/01/2024) | **Final staging:** pT2N1aM0 – Stage IIB (18/01/2024) | **ECOG:** 0 (25/03/2024) | **Comorbidities:** Compensated hypothyroidism and mild to moderate idiopathic chronic neutropenia (ANC ~1,200/mm³), with intermittent thrombocytopenia (minimum 120,000/mm³)

### **PATIENT 22 — SYNTHETIC EXCLUSION RECORD**

**Right Breast** | **Diagnosis:** Invasive ductal carcinoma, Histologic grade III (Nottingham 8/9), Size: 4.0 cm, Lymphovascular invasion present (02/06/2023) | **Initial clinical staging:** cT2N1M0 – Stage IIB (AJCC 8th edition) (02/06/2023) | **IHC:** ER 95%, PR 85%, HER2 1+, Ki-67 33% (02/06/2023) | **Surgery:** Right quadrantectomy + level I–II axillary lymphadenectomy (18/11/2023) | **Pathology:** Residual tumor 1.9 cm, Grade II, 12 lymph nodes (3 positive), Margins ≥ 3 mm, Lymphovascular invasion present (18/11/2023) | **Final staging:** pT1cN1aM0 – Stage IIB (18/11/2023) | **ECOG:** 1 (25/03/2024) | **Comorbidities:** Obesity, dyslipidemia, hypertension, and nonalcoholic steatohepatitis (NASH) grade 3 with moderate portal fibrosis (advanced NASH)

### **PATIENT 23 — SYNTHETIC EXCLUSION RECORD**

**Left Breast** | **Diagnosis:** Invasive ductal carcinoma, Histologic grade II (Nottingham 7/9), Size: 3.3 cm, Lymphovascular invasion present (15/09/2023) | **Initial clinical staging:** cT2N1M0 – Stage IIB (AJCC 8th edition) (15/09/2023) | **IHC:** ER 95%, PR 85%, HER2 0, Ki-67 26% (15/09/2023) | **Surgery:** Left quadrantectomy + level I–II axillary lymphadenectomy (12/02/2024) | **Pathology:** Residual tumor 2.8 cm, Grade II, 13 lymph nodes (3 positive), Margins ≥ 3 mm, Lymphovascular invasion present (12/02/2024) | **Final staging:** pT2N1aM0 – Stage IIB (12/02/2024) | **ECOG:** 1 (10/04/2024) | **Comorbidities:** Bipolar disorder type I in a mixed phase, with mood instability and elevated serum lithium levels (1.4 mEq/L)

### **PATIENT 24 — SYNTHETIC EXCLUSION RECORD**

**Right Breast** | **Diagnosis:** Invasive ductal carcinoma, Histologic grade III (Nottingham 8/9), Size: 3.8 cm, Lymphovascular invasion present (03/07/2023) | **Initial clinical staging:** cT2N1M0 – Stage IIB (AJCC 8th edition) (03/07/2023) | **IHC:** ER 90%, PR 70%, HER2 1+, Ki-67 30% (03/07/2023) | **Surgery:** Right quadrantectomy + level I–II axillary lymphadenectomy (20/12/2023) | **Pathology:** Residual tumor 2.3 cm, Grade II, 15 lymph nodes (3 positive), Margins ≥ 3 mm, Lymphovascular invasion present (20/12/2023) | **Final staging:** pT2N1aM0 – Stage IIB (20/12/2023) | **ECOG:** 1 (15/03/2024) | **Comorbidities:** Hypertension and HIV infection with moderate immunosuppression (CD4: 320 cells/mm³; viral load: 85,000 copies/mL)

### **PATIENT 25 — SYNTHETIC EXCLUSION RECORD**

**Left Breast** | **Diagnosis:** Invasive ductal carcinoma, Histologic grade III (Nottingham 8/9), Size: 4.2 cm, Lymphovascular invasion present (22/05/2023) | **Initial clinical staging:** cT2N1M0 – Stage IIB (AJCC 8th edition) (22/05/2023) | **IHC:** ER 90%, PR 85%, HER2 1+, Ki-67 28% (22/05/2023) | **Surgery:** Left quadrantectomy + level I–II axillary lymphadenectomy (10/11/2023) | **Pathology:** Residual tumor 2.1 cm, Grade II, 13 lymph nodes (3 positive), Margins ≥ 3 mm, Lymphovascular invasion present (10/11/2023) | **Final staging:** pT2N1aM0 – Stage IIB (10/11/2023) | **ECOG:** 1 (25/02/2024) | **Comorbidities:** Heart failure with reduced ejection fraction (LVEF 45%), hypertension, dyslipidemia, and obesity

**SCREENER - MONARCHE**

### **CONTEXT:**

You are a healthcare professional specialized in clinical oncology and translational research, responsible for screening oncology patients according to the inclusion and exclusion criteria of a research study. Your role is to analyze clinical records with precision, ensuring compliance with the study protocol.

### **OBJECTIVE (YOUR TASK):**

Rigorously review the patient’s medical record and determine eligibility based on the following inclusion and exclusion criteria summarized in the table below:

### **Inclusion Criteria:**

- Women (regardless of menopausal status) or men ≥18 years of age (or per local regulations).
- Confirmed HR+, HER2-, early-stage resected invasive breast cancer without evidence of distant metastases.
- The participant must have undergone definitive surgery for the primary breast tumor.
- Tumor tissue from breast (preferred) or lymph node must be available for exploratory biomarker analysis prior to randomization.
- Pathologic lymph node involvement and at least one of the following high-risk features:
  - 4 or more positive axillary lymph nodes
  - Tumor size ≥5 cm
  - Grade 3 (≥8 points on Bloom-Richardson scale)
  - Ki-67 ≥20% by central analysis on untreated breast tissue
- Participant must be randomized within 16 months from definitive breast cancer surgery.
- Participant may receive up to 12 weeks of endocrine therapy before randomization, following the last non-endocrine treatment (surgery, chemotherapy, or radiation), whichever comes last.
- Participant must have recovered (grade ≤1) from acute effects of chemotherapy, radiotherapy, and surgery.
- Women of childbearing potential must have a negative blood pregnancy test and agree to use highly effective contraception.
- ECOG performance status ≤1.
- Adequate organ function.
- Ability to swallow oral medications.

### **Exclusion Criteria:**

- Metastatic disease (including contralateral axillary lymph nodes) or node-negative disease.
- Inflammatory breast cancer.
- History of previous breast cancer, except ipsilateral DCIS treated with locoregional therapy ≥5 years ago. History of contralateral DCIS treated at any time with local therapy may be eligible. History of any other cancer (except non-melanoma skin cancer or carcinoma in situ of the cervix) unless in complete remission and off therapy for at least 5 years is excluded.
- Pregnant or breastfeeding females.
- Prior treatment with CDK4 or CDK6 inhibitors.
- Concurrent use of exogenous reproductive hormone therapy (e.g., contraceptive pills, hormone replacement therapy, megestrol acetate).
- Prior endocrine therapy for breast cancer prevention (tamoxifen, aromatase inhibitors) or raloxifene.
- Serious preexisting medical condition(s) deemed by the investigator to preclude participation.
- Personal history of any of the following:
  - Syncope of cardiovascular origin
  - Pathological ventricular arrhythmias
  - Sudden cardiac arrest
  - Venous thromboembolism (VTE)
- Active systemic infections or detectable viral load.
- Receipt of an experimental treatment in a clinical trial within the last 30 days or 5 half-lives, whichever is longer.

### **DECISION RULES TO APPLY:**

- **INCLUDED**: The patient meets all inclusion criteria and none of the exclusion criteria.
- **EXCLUDED**: The patient fails to meet at least one inclusion criterion or meets at least one exclusion criterion.

### **STYLE:**

The screening report should follow a formal, technical-medical style used in oncological records, using standardized, objective, and concise medical language. Sentences should be short, clear, and direct, avoiding unnecessary adjectives and subjective expressions.

### **TONE:**

Formal and appropriate for clinical oncologists and research investigators.

### **TARGET AUDIENCE:**

Healthcare professionals involved in oncology research and clinical screening processes, especially screening nurses, investigator physicians, and clinical study coordinators.

### **RESPONSE FORMAT:**

You may use the following structure:

- **Screening Outcome:** INCLUDED / EXCLUDED
- **Inclusion Criteria Met:** [describe objectively]
- **Exclusion Criteria Met:** [describe objectively]
- **Decision Summary:** [justify screening decision objectively]

### **PURPOSE SUMMARY OF THIS PROMPT:**

To generate oncological screening reports in formal medical language, based on defined inclusion and exclusion criteria, for use in the training and validation of artificial intelligence models applied to clinical studies and oncology research.

**SYNTHETIC PATIENT GENERATOR PROMPT - OLYMPIA**

**CONTEXT:**
You are a healthcare professional specialized in clinical oncology and translational research, responsible for creating synthetic databases of oncological medical records. These records are artificially generated, without the use of real personal information, and follow strict ethical and scientific standards. Their purpose is to support the training of artificial intelligence models aimed at patient screening for oncological studies and research.

**OBJECTIVE (YOUR TASK):**
Generate synthetic medical records with detailed clinical data, prepared based on the following inclusion and exclusion criteria presented in the table below:

**Inclusion Criteria:**

- Histologically confirmed non-metastatic primary invasive adenocarcinoma of the breast with one of the following phenotypes:
  - Triple negative breast cancer defined as: ER and PgR negative AND HER2 negative (not eligible for anti-HER2 therapy)
  - ER and/or PgR positive, HER2 negative
- Documented germline mutation in BRCA1 or BRCA2 that is predicted to be deleterious or suspected deleterious (known or predicted to be detrimental/lead to loss of function).
- Completed adequate breast and axillary surgery.
- Completed at least 6 cycles of neoadjuvant or adjuvant chemotherapy containing anthracyclines, taxanes, or a combination of both. Prior platinum as potentially curative treatment for a previous cancer (e.g., ovarian) or as adjuvant or neoadjuvant treatment for breast cancer is allowed.
- ECOG 0-1.

**Exclusion Criteria:**

- Any previous treatment with a PARP inhibitor, including olaparib and/or known hypersensitivity to any of the excipients of the study treatment.
- Patients with a second primary malignancy. EXCEPTIONS are:
  - Adequately treated non-melanoma skin cancer, curatively treated in situ cervical cancer, ductal carcinoma in situ (DCIS) of the breast, stage 1 grade 1 endometrial carcinoma
  - Other solid tumors and lymphomas (without bone marrow involvement) diagnosed ≥ 5 years before randomization and treated with no evidence of disease recurrence and for whom no more than one line of chemotherapy was applied.
- Concomitant use of known strong CYP3A inhibitors (e.g., itraconazole, telithromycin, clarithromycin, protease inhibitors boosted with ritonavir or cobicistat, indinavir, saquinavir, nelfinavir, boceprevir, telaprevir) or moderate CYP3A inhibitors (e.g., ciprofloxacin, erythromycin, diltiazem, fluconazole, verapamil). The required washout period prior to starting study treatment is 2 weeks.
- Concomitant use of known strong (e.g., phenobarbital, enzalutamide, phenytoin, rifampin, rifabutin, rifapentine, carbamazepine, nevirapine and St John's Wort) or moderate CYP3A inducers (e.g., bosentan, efavirenz, modafinil). The required washout period prior to starting study treatment is 5 weeks for enzalutamide or phenobarbital and 3 weeks for other agents.
- Evidence of metastatic breast cancer.

**STYLE:**
The text must follow the medical-technical style of oncological records, using medical, standardized, objective, and formal language. Avoid superfluous adjectives and subjective expressions.

**TONE:**
Formal. It should reflect the language used by oncologists and researchers.

**TARGET AUDIENCE:**
Healthcare professionals involved in oncological research and clinical screening, especially screening nurses, physician-researchers, and clinical trial coordinators.

**RESPONSE FORMAT:**
 Use typical clinical record structure:

**1. Identification**

- Synthetic patient ID
- Exact age (years and months)
- Sex and ethnicity / skin color
- Weight, height, BMI
- Marital status and profession
- Treatment location (hospital / cancer center)
- Date of diagnosis
- Date of surgery
- Date of inclusion/randomization in the study
- ECOG Performance Status

**2. Current Disease History (CDH)**

- Symptom onset date and progression
- Detection method (self-exam, mammography, ultrasound, etc.)
- Reason for referral to the cancer center
- Laterality of the lesion (right/left)
- Initial complete clinical TNM classification (e.g., cT2N1M0, stage IIB)
- Biopsy date and type (core needle, vacuum-assisted)
- Histological type and Nottingham grade
- Ki-67 index (%)
- Hormone receptors (ER, PR) and HER2 (IHC/FISH)
- Clinical staging according to AJCC (8th edition)

**3. Personal and Family History**

- Relevant comorbidities (hypertension, diabetes, dyslipidemia, etc.)
- Drug allergies
- Gynecological history (menarche, menopause, hormonal use, G P A)
- Family history of cancer (breast, ovarian, colon, prostate)
- Previous thoracic radiotherapy
- Lifestyle habits (smoking, alcohol use)

**4. Physical Examination**

- General condition and systemic signs (fatigue, weight loss)
- Bilateral breast findings
- Tumor size and location (in cm)
- Skin changes, nipple retraction, discharge
- Palpable lymph nodes (number, size, and mobility)
- Presence of pain, lymphedema, or inflammatory signs

**5. Complementary Tests**

- CBC (approximate values: Hb, leukocytes, platelets)
- Liver function (AST, ALT), renal (creatinine, urea)
- Beta-HCG (negative for women of reproductive age)
- LVEF pre- and post-chemotherapy (ECHO or MUGA, with dates and <15 p.p. variation)
- Serologies (HBsAg, anti-HCV, HIV)
- Ki-67, HER2 confirmation (FISH/CISH)
- Minimum laboratory criteria (Hb >10; neutrophils >1500; platelets >100,000)
- Imaging for staging (CT chest/abdomen/pelvis, bone scan)

**6. Neoadjuvant Treatment**

- Regimen and drugs used
- Doses (e.g., docetaxel 75 mg/m²)
- Start and end dates
- Total duration (≥ 16 weeks)
- Duration of trastuzumab and taxane use (≥ 9 weeks each)
- Observed clinical response (tumor reduction / stable / no progression)
- Adverse events (type and grade – NCI-CTCAE)
- Interval between last dose and surgery (≤ 12 weeks)

**7. Surgical and Pathological Data**

- Type of surgery (partial / radical / immediate or delayed reconstruction)
- Type of incision
- Laterality
- Total number of resected and positive lymph nodes
- Residual tumor size (in cm)
- Surgical margins (in mm)
- Histological grade post-chemotherapy
- Pathological response (pCR, partial response, minimal residual disease)
- Presence of lymphovascular / perineural invasion
- Final pathological staging (ypTNM)

**PROMPT PURPOSE SUMMARY:**
To generate complete, structured, and realistic synthetic oncological patient records, written in formal medical language, according to inclusion and exclusion criteria defined by the researcher, aimed at training artificial intelligence models for oncological patient screening in clinical research.

**SYNTHETIC PATIENT RECORDS - OLYMPIA**

**PATIENT 1 — SYNTHETIC INCLUSION RECORD**

**1. Identification**
 **Synthetic ID:** OLY-001
 **Age:** 44 years and 3 months
 **Sex / Ethnicity:** Female, White
 **Weight / Height / BMI:** 68 kg / 1.64 m / 25.3 kg/m²
 **Marital Status / Occupation:** Married / High School Teacher
 **Treatment Center:** National Cancer Institute (INCA) – Rio de Janeiro, Brazil
 **Date of Diagnosis:** 04/17/2023
 **Date of Surgery:** 11/10/2023
 **Date of Study Inclusion:** 02/20/2024
 **ECOG Performance Status:** 0

**2. Present Illness History (PIH)**
 Symptoms began in February 2023, when the patient noticed a firm, painless nodule in the left breast during self-examination.
 Mammogram and ultrasound on 03/10/2023 showed a 3.8 cm spiculated lesion in the upper outer quadrant of the left breast and a 1.7 cm axillary lymph node.
 Referred to oncology center for further evaluation and treatment.
 **Initial Clinical TNM Staging:** cT2N1M0 – Stage IIB (AJCC 8th edition)
 **Core Needle Biopsy:** Performed on 04/17/2023. Result: invasive ductal carcinoma, Nottingham grade III (8/9).
 **Immunohistochemistry:** ER 0%, PR 0%, HER2 0 (negative), Ki-67 65%
 **Histological Type:** Invasive ductal carcinoma, triple-negative phenotype

**3. Personal and Family History**
 **Relevant Comorbidities:** Controlled systemic arterial hypertension (losartan 50 mg/day)
 **Drug Allergies:** None reported
 **Gynecological History:** Menarche at 12, 2 pregnancies, 2 deliveries, no abortions; menopause at 43; no use of hormone therapy
 **Family History of Cancer:** Mother with ovarian cancer at age 55; maternal aunt with breast cancer at age 49
 **Genetic Testing:** Germline BRCA1 mutation c.5266dupC (deleterious), confirmed on 05/05/2023
 **Lifestyle Habits:** Non-smoker; occasional social alcohol consumption
 **Prior Chest Radiotherapy:** None

**4. Physical Examination**
 Good general condition; no reported weight loss
 **Breast Findings:** Firm, poorly defined nodule approximately 3.5 cm in upper outer quadrant of the left breast; no skin retraction or nipple discharge
 **Lymph Nodes:** Palpable axillary lymphadenopathy (1 to 2 mobile nodes, approximately 1.5 cm)
 No inflammatory signs, pain, or lymphedema

**5. Complementary Tests**
 **Complete Blood Count:** Hemoglobin 12.8 g/dL; leukocytes 6,900/mm³; platelets 255,000/mm³
 **Liver Function:** AST 32 U/L; ALT 29 U/L
 **Renal Function:** Creatinine 0.78 mg/dL; urea 28 mg/dL
 **Beta-HCG:** Negative
 **Serologies:** HBsAg, anti-HCV, and HIV negative
 **LVEF (Echocardiogram):** 66% before chemotherapy; 61% after chemotherapy (variation <10 percentage points)
 **Imaging:**

- Chest/abdomen/pelvis CT: No signs of distant metastases
- Bone scintigraphy: No abnormal uptake

**6. Neoadjuvant Treatment**
 **Regimen:** AC-T

- Doxorubicin 60 mg/m² + Cyclophosphamide 600 mg/m², 4 cycles (05/15/2023 – 07/10/2023)
- Docetaxel 75 mg/m², 4 cycles (07/31/2023 – 09/25/2023)
   **Total Duration:** 20 weeks
   **Adverse Events:** Grade 2 nausea, grade 2 alopecia, grade 2 neutropenia (per NCI-CTCAE)
   **Clinical Response:** Tumor reduction of approximately 40%
   **Interval Between Last Chemotherapy and Surgery:** 6 weeks

**7. Surgical and Pathological Data**
 **Surgical Procedure:** Left quadrantectomy with level I–II axillary lymphadenectomy
 **Incision Type:** Radial, upper outer quadrant
 **Laterality:** Left
 **Lymph Nodes Resected:** 15 total, 3 positive
 **Residual Tumor Size:** 1.8 cm
 **Surgical Margins:** Free, ≥5 mm
 **Histologic Grade Post-Chemotherapy:** Grade II
 **Lymphovascular Invasion:** Present
 **Pathologic Response:** Partial (no pCR)
 **Final Pathological Staging:** ypT1cN1aM0

**PATIENT 2 — SYNTHETIC INCLUSION RECORD**

**1. Identification**
 **Synthetic ID:** OLY-002
 **Age:** 39 years and 7 months
 **Sex / Ethnicity:** Female, Mixed race (parda)
 **Weight / Height / BMI:** 74 kg / 1.58 m / 29.6 kg/m²
 **Marital Status / Occupation:** Single / Civil servant (public administration)
 **Treatment Center:** Hospital de Câncer de Barretos – Barretos, SP, Brazil
 **Date of Diagnosis:** 06/23/2023
 **Date of Surgery:** 01/15/2024
 **Date of Study Inclusion:** 03/12/2024
 **ECOG Performance Status:** 1

**2. Present Illness History (PIH)**
 Patient reported progressive increase in volume in the right breast since April 2023, initially interpreted as mastitis.
 Breast ultrasound on 05/15/2023 revealed an irregular mass measuring 4.5 cm, associated with skin thickening and enlarged axillary lymph nodes.
 Core biopsy on 06/23/2023 confirmed invasive ductal carcinoma, grade III, triple-negative.
 **Initial Clinical TNM Staging:** cT3N1M0 – Stage IIIA (AJCC 8th edition)
 **Immunohistochemistry:** ER 0%, PR 0%, HER2 0, Ki-67 70%
 **Genetic Testing:** BRCA2 germline mutation c.5946delT (deleterious), confirmed 07/10/2023

**3. Personal and Family History**
 **Relevant Comorbidities:** Type 2 diabetes mellitus (on metformin), obesity
 **Drug Allergies:** No known allergies
 **Gynecologic History:** Menarche at 11, 0 pregnancies, regular menstrual cycles, no hormonal contraceptive use
 **Family History of Cancer:** Maternal grandmother with breast cancer at 62; no other known familial cases
 **Prior Chest Radiotherapy:** None
 **Lifestyle Habits:** Ex-smoker (quit in 2020, 10 pack-years); moderate alcohol use

**4. Physical Examination**
 Patient in regular general condition, reported fatigue and breast pain
 **Right breast:** Tense, swollen, tumor mass approximately 5.0 cm with skin thickening and signs of inflammation
 **Lymph Nodes:** Palpable axillary lymphadenopathy, 2–3 nodes, firm and mobile
 No signs of distant disease or lymphedema

**5. Complementary Tests**
 **Complete Blood Count:** Hb 11.6 g/dL; WBC 7,200/mm³; Platelets 235,000/mm³
 **Liver Function:** AST 28 U/L; ALT 34 U/L
 **Renal Function:** Creatinine 0.82 mg/dL; Urea 31 mg/dL
 **Beta-HCG:** Negative
 **Serologies:** Non-reactive for HIV, HCV, HBsAg
 **LVEF (MUGA):** 64% pre-chemotherapy; 60% post-chemotherapy
 **CT Chest/Abdomen/Pelvis:** No signs of distant metastases
 **Bone Scan:** Normal

**6. Neoadjuvant Treatment**
 **Regimen:** Platinum-based + taxane + anthracycline

- Paclitaxel 80 mg/m² weekly + Carboplatin AUC 5, 12 weeks (07/17/2023 – 10/09/2023)
- Doxorubicin 60 mg/m² + Cyclophosphamide 600 mg/m² every 3 weeks, 4 cycles (10/30/2023 – 01/08/2024)
   **Total Duration:** 25 weeks
   **Adverse Events:** Grade 2 peripheral neuropathy, grade 1 anemia
   **Clinical Response:** Marked reduction in tumor volume (~70%)
   **Interval Between Last Chemotherapy and Surgery:** 1 week

**7. Surgical and Pathological Data**
 **Surgical Procedure:** Right mastectomy with immediate reconstruction and sentinel lymph node biopsy
 **Incision Type:** Oblique
 **Laterality:** Right
 **Lymph Nodes Resected:** 6 total, 0 positive
 **Residual Tumor Size:** No residual invasive tumor (only fibrotic tissue)
 **Surgical Margins:** Clear
 **Histologic Grade Post-Chemotherapy:** Not applicable (no viable tumor)
 **Lymphovascular Invasion:** Absent
 **Pathologic Response:** Complete (pCR)
 **Final Pathological Staging:** ypT0N0

**PATIENT 3 — SYNTHETIC INCLUSION RECORD**

**1. Identification**
 **Synthetic ID:** OLY-003
 **Age:** 52 years and 1 month
 **Sex / Ethnicity:** Female, Black
 **Weight / Height / BMI:** 71 kg / 1.60 m / 27.7 kg/m²
 **Marital Status / Occupation:** Married / Seamstress
 **Treatment Center:** Hospital das Clínicas – Universidade Federal de Minas Gerais (UFMG), Belo Horizonte, MG, Brazil
 **Date of Diagnosis:** 02/14/2023
 **Date of Surgery:** 09/01/2023
 **Date of Study Inclusion:** 11/20/2023
 **ECOG Performance Status:** 0

**2. Present Illness History (PIH)**
 Patient identified a lump in the left breast during routine self-examination in January 2023.
 Mammography and ultrasound on 01/30/2023 showed a 2.6 cm solid nodule in the upper inner quadrant, with one enlarged left axillary lymph node.
 Referred by primary care for oncological evaluation.
 **Initial Clinical TNM Staging:** cT2N1M0 – Stage IIB (AJCC 8th edition)
 **Core Needle Biopsy:** 02/14/2023. Invasive ductal carcinoma, Nottingham grade II (6/9)
 **Immunohistochemistry:** ER 90%, PR 80%, HER2 0 (negative), Ki-67 22%
 **Histological Subtype:** Luminal B-like (HER2 negative)
 **Genetic Testing:** BRCA2 mutation c.8537_8538delAG (deleterious), confirmed on 03/12/2023

**3. Personal and Family History**
 **Relevant Comorbidities:** Systemic arterial hypertension, dyslipidemia
 **Drug Allergies:** None reported
 **Gynecological History:** Menarche at 13, menopause at 50, 3 pregnancies, 3 deliveries, 0 abortions; no hormone replacement therapy
 **Family History of Cancer:** Sister with breast cancer at 46; maternal cousin with prostate cancer
 **Prior Chest Radiotherapy:** None
 **Lifestyle Habits:** Non-smoker, no alcohol consumption

**4. Physical Examination**
 Patient in good general condition, no systemic symptoms
 **Left breast:** Palpable, well-defined mass, approximately 2.5 cm, no skin changes or discharge
 **Axillary nodes:** Single firm, mobile node ~1.5 cm
 No pain, lymphedema, or inflammatory signs

**5. Complementary Tests**
 **Complete Blood Count:** Hb 13.1 g/dL; leukocytes 7,100/mm³; platelets 276,000/mm³
 **Liver Function:** AST 23 U/L; ALT 21 U/L
 **Renal Function:** Creatinine 0.81 mg/dL; urea 29 mg/dL
 **Beta-HCG:** Negative
 **Serologies:** HBsAg, anti-HCV, HIV all non-reactive
 **LVEF (Echocardiogram):** 68% before chemotherapy; 63% after
 **Staging Imaging:**

- CT chest/abdomen/pelvis: No metastatic disease
- Bone scintigraphy: Normal

**6. Neoadjuvant Treatment**
 **Regimen:** AC followed by weekly paclitaxel

- Doxorubicin 60 mg/m² + Cyclophosphamide 600 mg/m², 4 cycles every 21 days (03/20/2023 – 05/15/2023)
- Paclitaxel 80 mg/m² weekly for 12 weeks (05/29/2023 – 08/21/2023)
   **Total Duration:** 23 weeks
   **Adverse Events:** Grade 1 fatigue, grade 2 mucositis, no dose-limiting toxicity
   **Clinical Response:** Partial reduction (~50%)
   **Interval Between Last Chemotherapy and Surgery:** 11 days

**7. Surgical and Pathological Data**
 **Surgical Procedure:** Left quadrantectomy with sentinel lymph node biopsy
 **Incision Type:** Periareolar radial
 **Laterality:** Left
 **Lymph Nodes Resected:** 4 total, 1 positive
 **Residual Tumor Size:** 1.4 cm
 **Surgical Margins:** Clear (≥3 mm)
 **Histologic Grade Post-Chemotherapy:** Grade II
 **Lymphovascular Invasion:** Absent
 **Pathologic Response:** Partial
 **Final Pathological Staging:** ypT1cN1mi

**PATIENT 4 — SYNTHETIC INCLUSION RECORD**

**1. Identification**
 **Synthetic ID:** OLY-004
 **Age:** 47 years and 10 months
 **Sex / Ethnicity:** Female, White
 **Weight / Height / BMI:** 64 kg / 1.63 m / 24.1 kg/m²
 **Marital Status / Occupation:** Divorced / Nutritionist
 **Treatment Center:** A.C. Camargo Cancer Center – São Paulo, SP, Brazil
 **Date of Diagnosis:** 05/08/2023
 **Date of Surgery:** 12/05/2023
 **Date of Study Inclusion:** 02/10/2024
 **ECOG Performance Status:** 0

**2. Present Illness History (PIH)**
 Patient noticed asymmetry in the contour of the right breast during a routine self-examination in April 2023.
 Breast ultrasound on 04/20/2023 showed a hypoechoic mass measuring 2.9 cm and an enlarged axillary lymph node.
 Core needle biopsy on 05/08/2023 confirmed invasive ductal carcinoma, grade II, hormone receptor positive, HER2 negative.
 **Initial Clinical TNM Staging:** cT2N1M0 – Stage IIB (AJCC 8th edition)
 **Immunohistochemistry:** ER 95%, PR 60%, HER2 negative (0), Ki-67 28%
 **Histological Subtype:** Luminal B-like (HER2 negative)
 **Genetic Testing:** BRCA1 mutation c.181T>G (deleterious), confirmed 06/01/2023

**3. Personal and Family History**
 **Relevant Comorbidities:** Hypothyroidism (levothyroxine 75 mcg/day)
 **Drug Allergies:** None reported
 **Gynecological History:** Menarche at 12, regular cycles, 1 pregnancy, 1 delivery, menopause induced by chemotherapy
 **Family History of Cancer:** Paternal aunt with breast cancer at 52; maternal grandfather with colon cancer
 **Prior Chest Radiotherapy:** None
 **Lifestyle Habits:** Non-smoker; moderate alcohol use

**4. Physical Examination**
 Good general condition, no fatigue or weight loss
 **Right breast:** Firm, retroareolar mass, approximately 3.0 cm, mild skin retraction
 **Lymph Nodes:** One firm, mobile axillary node palpable (~1.8 cm)
 No inflammatory signs or nipple discharge

**5. Complementary Tests**
 **Complete Blood Count:** Hb 12.9 g/dL; leukocytes 6,800/mm³; platelets 263,000/mm³
 **Liver Function:** AST 27 U/L; ALT 31 U/L
 **Renal Function:** Creatinine 0.76 mg/dL; urea 26 mg/dL
 **Beta-HCG:** Negative
 **Serologies:** HIV, HCV, and HBsAg negative
 **LVEF (Echocardiogram):** 65% before chemotherapy; 62% after chemotherapy
 **Staging Imaging:**

- CT chest/abdomen/pelvis: No evidence of metastases
- Bone scan: Normal

**6. Neoadjuvant Treatment**
 **Regimen:** Anthracycline and taxane

- Doxorubicin 60 mg/m² + Cyclophosphamide 600 mg/m², 4 cycles (06/12/2023 – 08/07/2023)
- Docetaxel 75 mg/m², 4 cycles (08/28/2023 – 10/23/2023)
   **Total Duration:** 20 weeks
   **Adverse Events:** Grade 2 arthralgia, grade 1 nausea, no neutropenia
   **Clinical Response:** Partial reduction (~40%)
   **Interval Between Last Chemotherapy and Surgery:** 6 weeks

**7. Surgical and Pathological Data**
 **Surgical Procedure:** Right quadrantectomy and level I–II axillary dissection
 **Incision Type:** Lateral radial
 **Laterality:** Right
 **Lymph Nodes Resected:** 12 total, 2 positive
 **Residual Tumor Size:** 1.6 cm
 **Surgical Margins:** Clear, ≥4 mm
 **Histologic Grade Post-Chemotherapy:** Grade II
 **Lymphovascular Invasion:** Present
 **Pathologic Response:** Partial
 **Final Pathological Staging:** ypT1cN1a

**PATIENT 5 — SYNTHETIC INCLUSION RECORD**

**1. Identification**
 **Synthetic ID:** OLY-005
 **Age:** 42 years and 6 months
 **Sex / Ethnicity:** Female, Indigenous
 **Weight / Height / BMI:** 59 kg / 1.56 m / 24.2 kg/m²
 **Marital Status / Occupation:** Married / Agricultural worker
 **Treatment Center:** Hospital Universitário João de Barros Barreto – Belém, PA, Brazil
 **Date of Diagnosis:** 03/06/2023
 **Date of Surgery:** 10/02/2023
 **Date of Study Inclusion:** 12/18/2023
 **ECOG Performance Status:** 1

**2. Present Illness History (PIH)**
 Patient presented with mastalgia and nodularity in the left breast noted since January 2023.
 Ultrasound performed on 02/15/2023 revealed a solid, irregular, hypoechoic nodule measuring 3.2 cm, BI-RADS 5.
 Referred by primary care to oncology service.
 **Initial Clinical TNM Staging:** cT2N0M0 – Stage IIA (AJCC 8th edition)
 **Core Needle Biopsy:** 03/06/2023. Invasive ductal carcinoma, Nottingham grade III (8/9)
 **Immunohistochemistry:** ER negative, PR negative, HER2 0, Ki-67 85%
 **Histological Subtype:** Triple-negative
 **Genetic Testing:** BRCA1 mutation c.68_69delAG (deleterious), confirmed 03/28/2023

**3. Personal and Family History**
 **Relevant Comorbidities:** None reported
 **Drug Allergies:** No known allergies
 **Gynecological History:** Menarche at 12, 4 pregnancies, 4 deliveries, premenopausal, no hormonal contraceptive use
 **Family History of Cancer:** Mother with breast cancer at 59; no other known cases
 **Prior Chest Radiotherapy:** None
 **Lifestyle Habits:** Non-smoker, no alcohol use

**4. Physical Examination**
 Patient in preserved general condition, mild fatigue
 **Left breast:** Firm, irregular mass in upper outer quadrant, 3.0–3.5 cm, no skin changes
 **Lymph Nodes:** No palpable axillary or supraclavicular nodes
 No pain, erythema, or nipple discharge

**5. Complementary Tests**
 **Complete Blood Count:** Hb 13.0 g/dL; leukocytes 7,600/mm³; platelets 242,000/mm³
 **Liver Function:** AST 30 U/L; ALT 26 U/L
 **Renal Function:** Creatinine 0.84 mg/dL; urea 25 mg/dL
 **Beta-HCG:** Negative
 **Serologies:** HIV, HCV, and HBsAg non-reactive
 **LVEF (Echocardiogram):** 67% before chemotherapy; 63% after treatment
 **Staging Imaging:**

- CT chest/abdomen/pelvis: No signs of metastasis
- Bone scintigraphy: Normal

**6. Neoadjuvant Treatment**
 **Regimen:** Platinum + anthracycline + taxane

- Paclitaxel 80 mg/m² weekly + Carboplatin AUC 5, 12 weeks (04/03/2023 – 06/26/2023)
- Doxorubicin 60 mg/m² + Cyclophosphamide 600 mg/m², 4 cycles (07/10/2023 – 08/28/2023)
   **Total Duration:** 22 weeks
   **Adverse Events:** Grade 2 diarrhea, grade 2 fatigue, grade 1 leukopenia
   **Clinical Response:** Near-complete tumor regression on imaging
   **Interval Between Last Chemotherapy and Surgery:** 5 weeks

**7. Surgical and Pathological Data**
 **Surgical Procedure:** Left mastectomy with sentinel lymph node biopsy
 **Incision Type:** Transverse
 **Laterality:** Left
 **Lymph Nodes Resected:** 5 total, all negative
 **Residual Tumor Size:** No residual invasive tumor
 **Surgical Margins:** Free of neoplasia
 **Histologic Grade Post-Chemotherapy:** Not applicable (pCR)
 **Lymphovascular Invasion:** Absent
 **Pathologic Response:** Complete (pCR)
 **Final Pathological Staging:** ypT0N0

**PATIENT 6 — SYNTHETIC EXCLUSION RECORD**

**1. Identification**
 **Synthetic ID:** OLY-006
 **Age:** 50 years and 9 months
 **Sex / Ethnicity:** Female, Asian descent
 **Weight / Height / BMI:** 67 kg / 1.61 m / 25.9 kg/m²
 **Marital Status / Occupation:** Widowed / Retail store manager
 **Treatment Center:** Hospital São Paulo – Escola Paulista de Medicina (UNIFESP), São Paulo, SP, Brazil
 **Date of Diagnosis:** 01/19/2023
 **Date of Surgery:** 08/14/2023
 **Date of Study Inclusion:** 10/30/2023
 **ECOG Performance Status:** 1

**2. Present Illness History (PIH)**
 Patient reported a painless lump in the right breast noticed during self-examination in December 2022.
 Mammography and ultrasound on 01/05/2023 identified a spiculated nodule of 3.4 cm in the upper outer quadrant and two axillary lymph nodes suggestive of malignancy.
 Biopsy on 01/19/2023 confirmed invasive ductal carcinoma, grade II.
 **Initial Clinical TNM Staging:** cT2N1M0 – Stage IIB (AJCC 8th edition)
 **Immunohistochemistry:** ER 98%, PR 70%, HER2 negative (IHC 1+), Ki-67 34%
 **Histological Subtype:** Luminal B-like (HER2 negative)
 **Genetic Testing:** BRCA2 mutation c.9097dupA (deleterious), confirmed 02/15/2023

**3. Personal and Family History**
 **Relevant Comorbidities:** Hypothyroidism (levothyroxine 88 mcg/day), type 2 diabetes mellitus (on metformin)
 **Drug Allergies:** No known drug allergies
 **Gynecological History:** Menarche at 11, menopause at 49, 2 pregnancies, 2 deliveries, no hormone replacement therapy
 **Family History of Cancer:** Brother with prostate cancer at 60; paternal aunt with breast cancer
 **Prior Chest Radiotherapy:** None
 **Lifestyle Habits:** Former smoker (quit in 2015), light alcohol use

**4. Physical Examination**
 Patient in good general condition, no signs of cachexia or fatigue
 **Right breast:** Palpable, firm mass measuring approximately 3.5 cm, no skin changes or nipple discharge
 **Axillary nodes:** Palpable, two mobile nodes, 1.5–2.0 cm
 No inflammatory signs, no lymphedema

**5. Complementary Tests**
 **Complete Blood Count:** Hb 12.4 g/dL; leukocytes 6,300/mm³; platelets 248,000/mm³
 **Liver Function:** AST 29 U/L; ALT 32 U/L
 **Renal Function:** Creatinine 0.88 mg/dL; urea 30 mg/dL
 **Beta-HCG:** Negative
 **Serologies:** HIV, HCV, HBsAg all negative
 **LVEF (MUGA):** 64% pre-chemotherapy; 61% post-chemotherapy
 **Staging Imaging:**

- CT chest/abdomen/pelvis: No distant metastases
- Bone scintigraphy: No abnormal uptake

**6. Neoadjuvant Treatment**
 **Regimen:** Anthracycline followed by taxane

- Doxorubicin 60 mg/m² + Cyclophosphamide 600 mg/m², 4 cycles every 3 weeks (02/27/2023 – 04/24/2023)
- Docetaxel 75 mg/m² every 3 weeks, 4 cycles (05/15/2023 – 07/10/2023)
   **Total Duration:** 20 weeks
   **Adverse Events:** Grade 1 nausea, grade 2 hyperglycemia, grade 2 neuropathy
   **Clinical Response:** Tumor reduction estimated at ~50%
   **Interval Between Last Chemotherapy and Surgery:** 5 weeks

**7. Surgical and Pathological Data**
 **Surgical Procedure:** Right quadrantectomy with level I–II axillary lymph node dissection
 **Incision Type:** Curvilinear lateral
 **Laterality:** Right
 **Lymph Nodes Resected:** 14 total, 3 positive
 **Residual Tumor Size:** 1.7 cm
 **Surgical Margins:** Free (≥4 mm)
 **Histologic Grade Post-Chemotherapy:** Grade II
 **Lymphovascular Invasion:** Present
 **Pathologic Response:** Partial
 **Final Pathological Staging:** ypT1cN1a

**PATIENT 7 — SYNTHETIC EXCLUSION RECORD**

**1. Identification**
 **Synthetic ID:** OLY-007
 **Age:** 36 years and 11 months
 **Sex / Ethnicity:** Female, White
 **Weight / Height / BMI:** 62 kg / 1.65 m / 22.8 kg/m²
 **Marital Status / Occupation:** Single / Graphic designer
 **Treatment Center:** Hospital Erasto Gaertner – Curitiba, PR, Brazil
 **Date of Diagnosis:** 08/10/2023
 **Date of Surgery:** 03/04/2024
 **Date of Study Inclusion:** 05/06/2024
 **ECOG Performance Status:** 0

**2. Present Illness History (PIH)**
 Patient reported a palpable nodule in the left breast during self-examination in July 2023.
 Ultrasound on 07/24/2023 revealed a solid, irregular lesion measuring 3.9 cm in the upper outer quadrant, BI-RADS 5.
 Biopsy performed on 08/10/2023 confirmed invasive ductal carcinoma, histological grade III.
 **Initial Clinical TNM Staging:** cT2N1M0 – Stage IIB (AJCC 8th edition)
 **Immunohistochemistry:** ER 0%, PR 0%, HER2 0, Ki-67 75%
 **Histological Subtype:** Triple-negative
 **Genetic Testing:** BRCA1 mutation c.4035delA (deleterious), confirmed 08/28/2023

**3. Personal and Family History**
 **Relevant Comorbidities:** None reported
 **Drug Allergies:** Penicillin (rash)
 **Gynecological History:** Menarche at 13, nulligravida, regular cycles, no hormonal contraceptive use
 **Family History of Cancer:** Father with pancreatic cancer; maternal aunt with ovarian cancer
 **Prior Chest Radiotherapy:** None
 **Lifestyle Habits:** Non-smoker, occasional alcohol use

**4. Physical Examination**
 Patient in good overall health, no systemic symptoms
 **Left breast:** Solid, fixed mass, ~3.8 cm in upper outer quadrant, without skin or nipple alterations
 **Lymph Nodes:** 1 palpable axillary node ~1.5 cm, mobile
 No signs of inflammation, no pain, no lymphedema

**5. Complementary Tests**
 **Complete Blood Count:** Hb 13.4 g/dL; leukocytes 6,700/mm³; platelets 258,000/mm³
 **Liver Function:** AST 26 U/L; ALT 23 U/L
 **Renal Function:** Creatinine 0.74 mg/dL; urea 27 mg/dL
 **Beta-HCG:** Negative
 **Serologies:** HBsAg, anti-HCV, HIV negative
 **LVEF (Echocardiogram):** 66% before chemotherapy; 64% after
 **Staging Imaging:**

- Chest/abdomen/pelvis CT: No metastatic disease
- Bone scan: No abnormalities

**6. Neoadjuvant Treatment**
 **Regimen:** Platinum-based + taxane + anthracycline

- Weekly paclitaxel 80 mg/m² + carboplatin AUC 5, for 12 weeks (09/11/2023 – 12/04/2023)
- Doxorubicin 60 mg/m² + cyclophosphamide 600 mg/m², 4 cycles every 21 days (12/26/2023 – 02/19/2024)
   **Total Duration:** 24 weeks
   **Adverse Events:** Grade 2 neutropenia, grade 1 nausea, grade 2 alopecia
   **Clinical Response:** Near-complete response on imaging
   **Interval Between Last Chemotherapy and Surgery:** 2 weeks

**7. Surgical and Pathological Data**
 **Surgical Procedure:** Left mastectomy with immediate reconstruction and sentinel lymph node biopsy
 **Incision Type:** Transverse inframammary
 **Laterality:** Left
 **Lymph Nodes Resected:** 5 total, 0 positive
 **Residual Tumor Size:** No residual invasive carcinoma
 **Surgical Margins:** Clear
 **Histologic Grade Post-Chemotherapy:** Not applicable (pCR)
 **Lymphovascular Invasion:** Absent
 **Pathologic Response:** Complete (pCR)
 **Final Pathological Staging:** ypT0N0

**PATIENT 8 — SYNTHETIC EXCLUSION RECORD**

**1. Identification**
 **Synthetic ID:** OLY-008
 **Age:** 55 years and 2 months
 **Sex / Ethnicity:** Female, Black
 **Weight / Height / BMI:** 70 kg / 1.59 m / 27.7 kg/m²
 **Marital Status / Occupation:** Married / Administrative assistant
 **Treatment Center:** Hospital de Clínicas de Porto Alegre (HCPA) – Porto Alegre, RS, Brazil
 **Date of Diagnosis:** 11/17/2022
 **Date of Surgery:** 06/12/2023
 **Date of Study Inclusion:** 09/01/2023
 **ECOG Performance Status:** 0

**2. Present Illness History (PIH)**
 Patient identified a lump in the right breast during showering in October 2022.
 Mammogram and ultrasound on 11/03/2022 showed a 2.7 cm lesion in the upper outer quadrant, with one suspicious axillary node.
 Core biopsy performed on 11/17/2022 showed invasive ductal carcinoma, Nottingham grade II.
 **Initial Clinical TNM Staging:** cT2N1M0 – Stage IIB (AJCC 8th edition)
 **Immunohistochemistry:** ER 95%, PR 90%, HER2 negative (IHC 0), Ki-67 18%
 **Histological Subtype:** Luminal B-like (HER2 negative, high Ki-67)
 **Genetic Testing:** BRCA2 mutation c.3847_3848delGT (deleterious), confirmed on 12/12/2022

**3. Personal and Family History**
 **Relevant Comorbidities:** Hypertension, dyslipidemia, osteoarthritis
 **Drug Allergies:** None reported
 **Gynecological History:** Menarche at 14, menopause at 50, 2 pregnancies, 2 deliveries, no hormone therapy
 **Family History of Cancer:** Sister with breast cancer at 48; maternal uncle with colon cancer
 **Prior Chest Radiotherapy:** None
 **Lifestyle Habits:** Former smoker (quit in 2010); no alcohol use

**4. Physical Examination**
 Patient in good general condition, no systemic complaints
 **Right breast:** Palpable, well-defined nodule ~2.5–3.0 cm, no skin or nipple alterations
 **Axillary nodes:** One mobile, firm lymph node ~1.5 cm
 No inflammatory signs, no lymphedema

**5. Complementary Tests**
 **Complete Blood Count:** Hb 12.7 g/dL; leukocytes 6,500/mm³; platelets 265,000/mm³
 **Liver Function:** AST 24 U/L; ALT 27 U/L
 **Renal Function:** Creatinine 0.80 mg/dL; urea 28 mg/dL
 **Beta-HCG:** Negative
 **Serologies:** HBsAg, anti-HCV, and HIV all non-reactive
 **LVEF (Echocardiogram):** 64% pre-chemotherapy; 62% post-chemotherapy
 **Staging Imaging:**

- Chest/abdomen/pelvis CT: No signs of distant disease
- Bone scan: No abnormalities

**6. Neoadjuvant Treatment**
 **Regimen:** Anthracycline + taxane

- Doxorubicin 60 mg/m² + cyclophosphamide 600 mg/m², 4 cycles (12/19/2022 – 02/13/2023)
- Paclitaxel 80 mg/m² weekly for 12 weeks (03/06/2023 – 05/22/2023)
   **Total Duration:** 23 weeks
   **Adverse Events:** Grade 1 neuropathy, grade 1 mucositis
   **Clinical Response:** Tumor size reduction of ~50%
   **Interval Between Last Chemotherapy and Surgery:** 3 weeks

**7. Surgical and Pathological Data**
 **Surgical Procedure:** Right quadrantectomy with level I–II axillary lymph node dissection
 **Incision Type:** Oblique superior
 **Laterality:** Right
 **Lymph Nodes Resected:** 13 total, 2 positive
 **Residual Tumor Size:** 1.3 cm
 **Surgical Margins:** Free (≥5 mm)
 **Histologic Grade Post-Chemotherapy:** Grade II
 **Lymphovascular Invasion:** Absent
 **Pathologic Response:** Partial
 **Final Pathological Staging:** ypT1cN1a

**PATIENT 9 — SYNTHETIC EXCLUSION RECORD**

**1. Identification**
 **Synthetic ID:** OLY-009
 **Age:** 40 years and 4 months
 **Sex / Ethnicity:** Female, Mixed race (parda)
 **Weight / Height / BMI:** 75 kg / 1.68 m / 26.6 kg/m²
 **Marital Status / Occupation:** Married / Hairdresser
 **Treatment Center:** Hospital Ophir Loyola – Belém, PA, Brazil
 **Date of Diagnosis:** 04/21/2023
 **Date of Surgery:** 11/06/2023
 **Date of Study Inclusion:** 01/22/2024
 **ECOG Performance Status:** 1

**2. Present Illness History (PIH)**
 Patient sought medical attention after noticing progressive hardening and thickening of the skin in the left breast starting in March 2023.
 Ultrasound and mammography performed on 04/10/2023 showed a 4.6 cm mass with skin retraction and abnormal axillary lymph nodes.
 Core biopsy on 04/21/2023 confirmed invasive ductal carcinoma, histologic grade III.
 **Initial Clinical TNM Staging:** cT3N1M0 – Stage IIIA (AJCC 8th edition)
 **Immunohistochemistry:** ER 0%, PR 0%, HER2 0, Ki-67 78%
 **Histological Subtype:** Triple-negative
 **Genetic Testing:** BRCA1 mutation c.1961delA (deleterious), confirmed 05/09/2023

**3. Personal and Family History**
 **Relevant Comorbidities:** Obesity (BMI > 30), no other reported conditions
 **Drug Allergies:** No known allergies
 **Gynecological History:** Menarche at 12, 3 pregnancies, 3 deliveries, no abortions, premenopausal, no use of hormonal therapy
 **Family History of Cancer:** Maternal cousin with breast cancer at 45; no ovarian or prostate cancer reported
 **Prior Chest Radiotherapy:** None
 **Lifestyle Habits:** Smoker (10 cigarettes/day); occasional alcohol use

**4. Physical Examination**
 Patient in regular general condition, reports mild fatigue
 **Left breast:** Firm, partially fixed mass in upper outer quadrant, ~4.5 cm, associated with skin thickening and mild edema
 **Lymph Nodes:** Palpable axillary lymphadenopathy, 2 nodes, firm and mobile (~1.5–2.0 cm)
 No nipple discharge or erythema

**5. Complementary Tests**
 **Complete Blood Count:** Hb 11.9 g/dL; leukocytes 7,800/mm³; platelets 273,000/mm³
 **Liver Function:** AST 35 U/L; ALT 38 U/L
 **Renal Function:** Creatinine 0.90 mg/dL; urea 30 mg/dL
 **Beta-HCG:** Negative
 **Serologies:** HIV, HCV, and HBsAg negative
 **LVEF (Echocardiogram):** 63% pre-chemotherapy; 60% post-chemotherapy
 **Staging Imaging:**

- CT chest/abdomen/pelvis: No evidence of distant metastases
- Bone scan: Normal

**6. Neoadjuvant Treatment**
 **Regimen:** Platinum-based + taxane + anthracycline

- Weekly paclitaxel 80 mg/m² + carboplatin AUC 5 (05/15/2023 – 08/07/2023)
- Doxorubicin 60 mg/m² + cyclophosphamide 600 mg/m², 4 cycles (08/28/2023 – 10/16/2023)
   **Total Duration:** 22 weeks
   **Adverse Events:** Grade 2 anemia, grade 2 peripheral neuropathy, grade 1 mucositis
   **Clinical Response:** Partial response, with estimated tumor reduction of 50%
   **Interval Between Last Chemotherapy and Surgery:** 3 weeks

**7. Surgical and Pathological Data**
 **Surgical Procedure:** Left mastectomy with level I–II axillary dissection
 **Incision Type:** Oblique
 **Laterality:** Left
 **Lymph Nodes Resected:** 16 total, 3 positive
 **Residual Tumor Size:** 2.1 cm
 **Surgical Margins:** Clear (≥5 mm)
 **Histologic Grade Post-Chemotherapy:** Grade III
 **Lymphovascular Invasion:** Present
 **Pathologic Response:** Partial
 **Final Pathological Staging:** ypT2N1a

**PATIENT 10 — SYNTHETIC EXCLUSION RECORD**

**1. Identification**
 **Synthetic ID:** OLY-010
 **Age:** 45 years and 8 months
 **Sex / Ethnicity:** Female, White
 **Weight / Height / BMI:** 66 kg / 1.62 m / 25.1 kg/m²
 **Marital Status / Occupation:** Married / Pharmacist
 **Treatment Center:** Hospital de Amor – Barretos, SP, Brazil
 **Date of Diagnosis:** 07/11/2023
 **Date of Surgery:** 02/05/2024
 **Date of Study Inclusion:** 04/08/2024
 **ECOG Performance Status:** 0

**2. Present Illness History (PIH)**
 Patient noticed a lump in the right breast during self-examination in June 2023.
 Mammogram and breast ultrasound on 07/03/2023 identified a 3.1 cm hypoechoic irregular lesion in the upper inner quadrant, associated with one suspicious axillary lymph node.
 Core biopsy performed on 07/11/2023 confirmed invasive ductal carcinoma, histologic grade II.
 **Initial Clinical TNM Staging:** cT2N1M0 – Stage IIB (AJCC 8th edition)
 **Immunohistochemistry:** ER 98%, PR 70%, HER2 negative (IHC 0), Ki-67 26%
 **Histological Subtype:** Luminal B-like (HER2 negative)
 **Genetic Testing:** BRCA2 mutation c.5946delT (deleterious), confirmed 08/01/2023

**3. Personal and Family History**
 **Relevant Comorbidities:** Systemic arterial hypertension (on amlodipine)
 **Drug Allergies:** No known allergies
 **Gynecological History:** Menarche at 13, 2 pregnancies, 2 deliveries, menopause at 45 (chemotherapy-induced)
 **Family History of Cancer:** Mother with breast cancer at 62; paternal grandfather with prostate cancer
 **Prior Chest Radiotherapy:** None
 **Lifestyle Habits:** Non-smoker, moderate alcohol use

**4. Physical Examination**
 Patient in good general condition, no signs of systemic illness
 **Right breast:** Firm, mobile tumor in the upper inner quadrant measuring ~3.0 cm; no skin changes or nipple discharge
 **Lymph Nodes:** Palpable axillary lymph node ~1.8 cm, mobile
 No lymphedema, erythema, or inflammatory signs

**5. Complementary Tests**
 **Complete Blood Count:** Hb 12.5 g/dL; leukocytes 6,400/mm³; platelets 261,000/mm³
 **Liver Function:** AST 30 U/L; ALT 28 U/L
 **Renal Function:** Creatinine 0.86 mg/dL; urea 27 mg/dL
 **Beta-HCG:** Negative
 **Serologies:** HBsAg, anti-HCV, and HIV negative
 **LVEF (Echocardiogram):** 66% pre-chemotherapy; 63% post-chemotherapy
 **Staging Imaging:**

- CT chest/abdomen/pelvis: No evidence of distant metastasis
- Bone scan: No abnormal findings

**6. Neoadjuvant Treatment**
 **Regimen:** Anthracycline followed by taxane

- Doxorubicin 60 mg/m² + cyclophosphamide 600 mg/m², 4 cycles every 21 days (08/14/2023 – 10/09/2023)
- Docetaxel 75 mg/m², 4 cycles every 21 days (10/30/2023 – 12/25/2023)
   **Total Duration:** 20 weeks
   **Adverse Events:** Grade 1 fatigue, grade 2 constipation, no neutropenia
   **Clinical Response:** Partial response with approximately 40% tumor size reduction
   **Interval Between Last Chemotherapy and Surgery:** 6 weeks

**7. Surgical and Pathological Data**
 **Surgical Procedure:** Right quadrantectomy with sentinel lymph node biopsy
 **Incision Type:** Periareolar vertical
 **Laterality:** Right
 **Lymph Nodes Resected:** 7 total, 1 positive
 **Residual Tumor Size:** 1.6 cm
 **Surgical Margins:** Clear (≥3 mm)
 **Histologic Grade Post-Chemotherapy:** Grade II
 **Lymphovascular Invasion:** Present
 **Pathologic Response:** Partial
 **Final Pathological Staging:** ypT1cN1mi

**PATIENT 11 — SYNTHETIC EXCLUSION RECORD**

**1. Identification**
 **Synthetic ID:** OLY-011
 **Age:** 38 years and 2 months
 **Sex / Ethnicity:** Female, Indigenous
 **Weight / Height / BMI:** 58 kg / 1.55 m / 24.1 kg/m²
 **Marital Status / Occupation:** Single / Community health agent
 **Treatment Center:** Hospital Universitário da Universidade Federal do Amazonas (HU-UFAM) – Manaus, AM, Brazil
 **Date of Diagnosis:** 02/03/2023
 **Date of Surgery:** 09/04/2023
 **Date of Study Inclusion:** 11/10/2023
 **ECOG Performance Status:** 1

**2. Present Illness History (PIH)**
 Patient presented with progressive pain and swelling in the right breast since December 2022.
 Ultrasound on 01/20/2023 showed a 4.3 cm irregular mass with posterior shadowing and axillary lymphadenopathy.
 Core biopsy on 02/03/2023 revealed invasive ductal carcinoma, histologic grade III.
 **Initial Clinical TNM Staging:** cT2N1M0 – Stage IIB (AJCC 8th edition)
 **Immunohistochemistry:** ER 0%, PR 0%, HER2 0, Ki-67 80%
 **Histological Subtype:** Triple-negative
 **Genetic Testing:** BRCA1 mutation c.5263insC (deleterious), confirmed 02/25/2023

**3. Personal and Family History**
 **Relevant Comorbidities:** None reported
 **Drug Allergies:** No known allergies
 **Gynecological History:** Menarche at 11, 2 pregnancies, 2 deliveries, no hormonal contraceptive use, premenopausal
 **Family History of Cancer:** Maternal aunt with breast cancer; sister with ovarian cancer at 41
 **Prior Chest Radiotherapy:** None
 **Lifestyle Habits:** Non-smoker, no alcohol use

**4. Physical Examination**
 Patient in good general condition, reports moderate breast pain
 **Right breast:** Irregular mass in upper outer quadrant measuring approximately 4.0 cm; mild skin thickening
 **Lymph Nodes:** Two palpable, mobile, firm axillary nodes (up to 2.0 cm)
 No erythema, nipple discharge, or signs of inflammation

**5. Complementary Tests**
 **Complete Blood Count:** Hb 12.3 g/dL; leukocytes 7,200/mm³; platelets 251,000/mm³
 **Liver Function:** AST 31 U/L; ALT 29 U/L
 **Renal Function:** Creatinine 0.78 mg/dL; urea 24 mg/dL
 **Beta-HCG:** Negative
 **Serologies:** HIV, HCV, and HBsAg negative
 **LVEF (Echocardiogram):** 65% pre-chemotherapy; 62% post-chemotherapy
 **Staging Imaging:**

- CT chest/abdomen/pelvis: No distant metastases
- Bone scan: Normal

**6. Neoadjuvant Treatment**
 **Regimen:** Taxane + platinum + anthracycline

- Paclitaxel 80 mg/m² weekly + carboplatin AUC 5 (03/06/2023 – 05/29/2023)
- Doxorubicin 60 mg/m² + cyclophosphamide 600 mg/m², 4 cycles (06/19/2023 – 08/14/2023)
   **Total Duration:** 23 weeks
   **Adverse Events:** Grade 2 peripheral neuropathy, grade 1 anemia
   **Clinical Response:** Near-complete response on imaging
   **Interval Between Last Chemotherapy and Surgery:** 3 weeks

**7. Surgical and Pathological Data**
 **Surgical Procedure:** Right mastectomy with level I–II axillary dissection
 **Incision Type:** Elliptical
 **Laterality:** Right
 **Lymph Nodes Resected:** 15 total, 0 positive
 **Residual Tumor Size:** No residual invasive carcinoma
 **Surgical Margins:** Negative
 **Histologic Grade Post-Chemotherapy:** Not applicable (pCR)
 **Lymphovascular Invasion:** Absent
 **Pathologic Response:** Complete (pCR)
 **Final Pathological Staging:** ypT0N0

**PATIENT 12 — SYNTHETIC EXCLUSION RECORD**

**1. Identification**
 **Synthetic ID:** OLY-012
 **Age:** 48 years and 6 months
 **Sex / Ethnicity:** Female, Black
 **Weight / Height / BMI:** 72 kg / 1.67 m / 25.8 kg/m²
 **Marital Status / Occupation:** Divorced / Public school teacher
 **Treatment Center:** Instituto Nacional de Câncer (INCA) – Rio de Janeiro, RJ, Brazil
 **Date of Diagnosis:** 03/27/2023
 **Date of Surgery:** 10/09/2023
 **Date of Study Inclusion:** 01/03/2024
 **ECOG Performance Status:** 0

**2. Present Illness History (PIH)**
 Patient reported detection of a lump in the left breast during self-examination in early March 2023.
 Breast ultrasound and mammography performed on 03/15/2023 revealed a 3.5 cm irregular hypoechoic lesion with associated axillary adenopathy.
 Core biopsy on 03/27/2023 confirmed invasive ductal carcinoma, histologic grade II.
 **Initial Clinical TNM Staging:** cT2N1M0 – Stage IIB (AJCC 8th edition)
 **Immunohistochemistry:** ER 90%, PR 60%, HER2 negative (IHC 1+), Ki-67 30%
 **Histological Subtype:** Luminal B-like (HER2 negative)
 **Genetic Testing:** BRCA2 mutation c.7008-1G>A (deleterious), confirmed on 04/14/2023

**3. Personal and Family History**
 **Relevant Comorbidities:** Type 2 diabetes mellitus (on metformin), systemic arterial hypertension
 **Drug Allergies:** No known drug allergies
 **Gynecological History:** Menarche at 12, menopause at 47 (spontaneous), 2 pregnancies, 2 deliveries
 **Family History of Cancer:** Sister with breast cancer at age 51; maternal uncle with colorectal cancer
 **Prior Chest Radiotherapy:** None
 **Lifestyle Habits:** Non-smoker, does not consume alcohol

**4. Physical Examination**
 Patient in good overall condition, no systemic symptoms
 **Left breast:** Firm, mobile mass of approximately 3.3 cm in upper outer quadrant; no skin retraction or nipple changes
 **Lymph Nodes:** Palpable mobile axillary lymph node (~1.7 cm), no supraclavicular adenopathy
 No lymphedema, erythema, or inflammatory signs

**5. Complementary Tests**
 **Complete Blood Count:** Hb 12.6 g/dL; leukocytes 6,900/mm³; platelets 272,000/mm³
 **Liver Function:** AST 26 U/L; ALT 30 U/L
 **Renal Function:** Creatinine 0.83 mg/dL; urea 28 mg/dL
 **Beta-HCG:** Negative
 **Serologies:** Non-reactive for HIV, HCV, and HBsAg
 **LVEF (Echocardiogram):** 65% pre-chemotherapy; 63% post-chemotherapy
 **Staging Imaging:**

- CT chest/abdomen/pelvis: No evidence of metastasis
- Bone scintigraphy: Normal

**6. Neoadjuvant Treatment**
 **Regimen:** Anthracycline + taxane

- Doxorubicin 60 mg/m² + cyclophosphamide 600 mg/m², 4 cycles every 21 days (04/24/2023 – 06/19/2023)
- Docetaxel 75 mg/m², 4 cycles every 21 days (07/10/2023 – 09/04/2023)
   **Total Duration:** 20 weeks
   **Adverse Events:** Grade 1 fatigue, grade 2 myalgia, no hematologic toxicity
   **Clinical Response:** Partial tumor reduction (~50%)
   **Interval Between Last Chemotherapy and Surgery:** 5 weeks

**7. Surgical and Pathological Data**
 **Surgical Procedure:** Left quadrantectomy with sentinel lymph node biopsy
 **Incision Type:** Radial lateral
 **Laterality:** Left
 **Lymph Nodes Resected:** 6 total, 1 positive
 **Residual Tumor Size:** 1.9 cm
 **Surgical Margins:** Free (≥3 mm)
 **Histologic Grade Post-Chemotherapy:** Grade II
 **Lymphovascular Invasion:** Absent
 **Pathologic Response:** Partial
 **Final Pathological Staging:** ypT1cN1mi

**PATIENT 10 — SYNTHETIC EXCLUSION RECORD**

**1. Identification**
 **Synthetic ID:** OLY-013
 **Age:** 43 years and 5 months
 **Sex / Ethnicity:** Female, Mixed race (parda)
 **Weight / Height / BMI:** 69 kg / 1.60 m / 26.9 kg/m²
 **Marital Status / Occupation:** Married / Supermarket cashier
 **Treatment Center:** Hospital Universitário Professor Alberto Antunes – UFAL, Maceió, AL, Brazil
 **Date of Diagnosis:** 05/02/2023
 **Date of Surgery:** 12/04/2023
 **Date of Study Inclusion:** 02/10/2024
 **ECOG Performance Status:** 1

**2. Present Illness History (PIH)**
 Patient noted a nodule in the right breast during self-examination in April 2023.
 Breast ultrasound and mammography performed on 04/24/2023 revealed a solid, irregular nodule measuring 3.7 cm and enlarged axillary lymph nodes.
 Core needle biopsy performed on 05/02/2023 confirmed invasive ductal carcinoma, histologic grade III.
 **Initial Clinical TNM Staging:** cT2N1M0 – Stage IIB (AJCC 8th edition)
 **Immunohistochemistry:** ER 0%, PR 0%, HER2 0, Ki-67 82%
 **Histological Subtype:** Triple-negative
 **Genetic Testing:** BRCA1 mutation c.68_69delAG (deleterious), confirmed on 05/25/2023

**3. Personal and Family History**
 **Relevant Comorbidities:** Obesity (BMI > 30), dyslipidemia
 **Drug Allergies:** None reported
 **Gynecological History:** Menarche at 11, 2 pregnancies, 2 deliveries, premenopausal, no hormone therapy
 **Family History of Cancer:** Mother with breast cancer at age 48; maternal grandfather with pancreatic cancer
 **Prior Chest Radiotherapy:** None
 **Lifestyle Habits:** Smoker (15 cigarettes/day), light alcohol consumption

**4. Physical Examination**
 Patient in good general condition, no weight loss or systemic symptoms
 **Right breast:** Palpable mass ~3.5–4.0 cm in the upper outer quadrant, firm and fixed, no skin involvement
 **Lymph Nodes:** Two palpable axillary nodes, ~2.0 cm, mobile
 No inflammatory signs or nipple discharge

**5. Complementary Tests**
 **Complete Blood Count:** Hb 12.1 g/dL; leukocytes 7,800/mm³; platelets 276,000/mm³
 **Liver Function:** AST 33 U/L; ALT 35 U/L
 **Renal Function:** Creatinine 0.87 mg/dL; urea 29 mg/dL
 **Beta-HCG:** Negative
 **Serologies:** HIV, HCV, and HBsAg all non-reactive
 **LVEF (Echocardiogram):** 67% pre-chemotherapy; 64% post-chemotherapy
 **Staging Imaging:**

- CT chest/abdomen/pelvis: No metastatic lesions
- Bone scan: Normal

**6. Neoadjuvant Treatment**
 **Regimen:** Taxane + platinum + anthracycline

- Weekly paclitaxel 80 mg/m² + carboplatin AUC 5, 12 weeks (06/05/2023 – 08/28/2023)
- Doxorubicin 60 mg/m² + cyclophosphamide 600 mg/m², 4 cycles (09/18/2023 – 11/13/2023)
   **Total Duration:** 24 weeks
   **Adverse Events:** Grade 2 nausea, grade 2 alopecia, grade 1 neutropenia
   **Clinical Response:** Good clinical response with >70% tumor shrinkage
   **Interval Between Last Chemotherapy and Surgery:** 3 weeks

**7. Surgical and Pathological Data**
 **Surgical Procedure:** Right mastectomy with level I–II axillary dissection
 **Incision Type:** Oblique inferolateral
 **Laterality:** Right
 **Lymph Nodes Resected:** 14 total, 1 positive
 **Residual Tumor Size:** 0.9 cm
 **Surgical Margins:** Free (≥4 mm)
 **Histologic Grade Post-Chemotherapy:** Grade II
 **Lymphovascular Invasion:** Absent
 **Pathologic Response:** Near-complete response (residual minimal disease)
 **Final Pathological Staging:** ypT1bN1mi

**PATIENT 14 — SYNTHETIC EXCLUSION RECORD**

**1. Identification**
 **Synthetic ID:** OLY-014
 **Age:** 51 years and 3 months
 **Sex / Ethnicity:** Female, White
 **Weight / Height / BMI:** 63 kg / 1.58 m / 25.2 kg/m²
 **Marital Status / Occupation:** Married / Primary school teacher
 **Treatment Center:** Hospital das Clínicas da Universidade Federal do Paraná (HC-UFPR) – Curitiba, PR, Brazil
 **Date of Diagnosis:** 06/06/2023
 **Date of Surgery:** 01/08/2024
 **Date of Study Inclusion:** 03/15/2024
 **ECOG Performance Status:** 0

**2. Present Illness History (PIH)**
 Patient noticed localized thickening in the lower inner quadrant of the left breast in May 2023.
 Breast imaging (ultrasound and mammogram) on 05/29/2023 identified a 2.8 cm irregular mass, associated with one suspicious axillary node.
 Core biopsy on 06/06/2023 confirmed invasive ductal carcinoma, histologic grade II.
 **Initial Clinical TNM Staging:** cT2N1M0 – Stage IIB (AJCC 8th edition)
 **Immunohistochemistry:** ER 95%, PR 80%, HER2 negative (IHC 1+), Ki-67 32%
 **Histological Subtype:** Luminal B-like (HER2 negative)
 **Genetic Testing:** BRCA2 mutation c.8488-1G>A (deleterious), confirmed 06/28/2023

**3. Personal and Family History**
 **Relevant Comorbidities:** Hypertension (controlled), hypothyroidism
 **Drug Allergies:** No known drug allergies
 **Gynecological History:** Menarche at 12, menopause at 49, 2 pregnancies, 2 deliveries, no hormone replacement therapy
 **Family History of Cancer:** Mother with breast cancer at 58; sister with ovarian cancer
 **Prior Chest Radiotherapy:** None
 **Lifestyle Habits:** Non-smoker, light alcohol use

**4. Physical Examination**
 Patient in good general health, no systemic complaints
 **Left breast:** Firm, mobile nodule in the lower inner quadrant, ~2.7 cm, no skin or nipple changes
 **Lymph Nodes:** One mobile axillary node ~1.5 cm
 No signs of inflammation or lymphedema

**5. Complementary Tests**
 **Complete Blood Count:** Hb 12.9 g/dL; leukocytes 6,700/mm³; platelets 258,000/mm³
 **Liver Function:** AST 28 U/L; ALT 26 U/L
 **Renal Function:** Creatinine 0.81 mg/dL; urea 25 mg/dL
 **Beta-HCG:** Negative
 **Serologies:** HIV, HCV, and HBsAg non-reactive
 **LVEF (Echocardiogram):** 66% before chemotherapy; 63% after
 **Staging Imaging:**

- Chest/abdomen/pelvis CT: No metastatic disease
- Bone scan: Normal

**6. Neoadjuvant Treatment**
 **Regimen:** Anthracycline followed by taxane

- Doxorubicin 60 mg/m² + cyclophosphamide 600 mg/m², 4 cycles (07/10/2023 – 09/04/2023)
- Docetaxel 75 mg/m², 4 cycles (09/25/2023 – 11/20/2023)
   **Total Duration:** 20 weeks
   **Adverse Events:** Grade 1 fatigue, grade 2 arthralgia
   **Clinical Response:** Partial response with ~40% reduction
   **Interval Between Last Chemotherapy and Surgery:** 7 weeks

**7. Surgical and Pathological Data**
 **Surgical Procedure:** Left quadrantectomy with sentinel lymph node biopsy
 **Incision Type:** Medial radial
 **Laterality:** Left
 **Lymph Nodes Resected:** 5 total, 0 positive
 **Residual Tumor Size:** 1.4 cm
 **Surgical Margins:** Free (≥3 mm)
 **Histologic Grade Post-Chemotherapy:** Grade II
 **Lymphovascular Invasion:** Absent
 **Pathologic Response:** Partial
 **Final Pathological Staging:** ypT1cN0

**PATIENT 15 — SYNTHETIC EXCLUSION RECORD**

**1. Identification**
 **Synthetic ID:** OLY-015
 **Age:** 46 years and 7 months
 **Sex / Ethnicity:** Female, Black
 **Weight / Height / BMI:** 74 kg / 1.64 m / 27.5 kg/m²
 **Marital Status / Occupation:** Divorced / Cook
 **Treatment Center:** Hospital Universitário da Universidade Federal de Santa Catarina (HU-UFSC) – Florianópolis, SC, Brazil
 **Date of Diagnosis:** 08/18/2023
 **Date of Surgery:** 03/11/2024
 **Date of Study Inclusion:** 05/06/2024
 **ECOG Performance Status:** 1

**2. Present Illness History (PIH)**
 Patient presented to primary care after noting a painful mass in the right breast in late July 2023.
 Ultrasound and mammography on 08/10/2023 showed a 3.9 cm irregular mass in the upper outer quadrant and one suspicious axillary lymph node.
 Core biopsy performed on 08/18/2023 revealed invasive ductal carcinoma, histologic grade III.
 **Initial Clinical TNM Staging:** cT2N1M0 – Stage IIB (AJCC 8th edition)
 **Immunohistochemistry:** ER 0%, PR 0%, HER2 0, Ki-67 88%
 **Histological Subtype:** Triple-negative
 **Genetic Testing:** BRCA1 mutation c.5123C>A (deleterious), confirmed on 09/05/2023

**3. Personal and Family History**
 **Relevant Comorbidities:** Type 2 diabetes mellitus, obesity
 **Drug Allergies:** No known allergies
 **Gynecological History:** Menarche at 13, 3 pregnancies, 3 deliveries, premenopausal, no use of hormone therapy
 **Family History of Cancer:** Paternal aunt with breast cancer at age 50; no other known neoplasms
 **Prior Chest Radiotherapy:** None
 **Lifestyle Habits:** Smoker (5 cigarettes/day), no alcohol consumption

**4. Physical Examination**
 Patient in fair general condition, mild fatigue reported
 **Right breast:** Palpable, firm, partially mobile mass in upper outer quadrant, ~3.8–4.0 cm
 **Lymph Nodes:** One firm axillary node, ~2.0 cm, mobile
 No skin retraction, erythema, or nipple discharge

**5. Complementary Tests**
 **Complete Blood Count:** Hb 11.8 g/dL; leukocytes 8,100/mm³; platelets 269,000/mm³
 **Liver Function:** AST 36 U/L; ALT 33 U/L
 **Renal Function:** Creatinine 0.92 mg/dL; urea 29 mg/dL
 **Beta-HCG:** Negative
 **Serologies:** HBsAg, anti-HCV, and HIV non-reactive
 **LVEF (Echocardiogram):** 65% before chemotherapy; 61% after
 **Staging Imaging:**

- Chest/abdomen/pelvis CT: No evidence of distant metastasis
- Bone scan: No abnormalities

**6. Neoadjuvant Treatment**
 **Regimen:** Taxane + platinum + anthracycline

- Paclitaxel 80 mg/m² weekly + carboplatin AUC 5 (09/18/2023 – 12/11/2023)
- Doxorubicin 60 mg/m² + cyclophosphamide 600 mg/m², 4 cycles every 21 days (01/02/2024 – 02/26/2024)
   **Total Duration:** 24 weeks
   **Adverse Events:** Grade 2 neutropenia, grade 1 nausea, grade 2 alopecia
   **Clinical Response:** Partial reduction (~60%)
   **Interval Between Last Chemotherapy and Surgery:** 2 weeks

**7. Surgical and Pathological Data**
 **Surgical Procedure:** Right mastectomy with level I–II axillary dissection
 **Incision Type:** Transverse
 **Laterality:** Right
 **Lymph Nodes Resected:** 17 total, 2 positive
 **Residual Tumor Size:** 1.2 cm
 **Surgical Margins:** Negative (≥4 mm)
 **Histologic Grade Post-Chemotherapy:** Grade III
 **Lymphovascular Invasion:** Present
 **Pathologic Response:** Partial
 **Final Pathological Staging:** ypT1cN1a

**PATIENT 16 — SYNTHETIC EXCLUSION RECORD**

**1. Identification**
 **Synthetic ID:** OLY-016
 **Age:** 39 years and 0 months
 **Sex / Ethnicity:** Female, White
 **Weight / Height / BMI:** 60 kg / 1.63 m / 22.6 kg/m²
 **Marital Status / Occupation:** Single / Biologist
 **Treatment Center:** Hospital Universitário Walter Cantídio – UFC, Fortaleza, CE, Brazil
 **Date of Diagnosis:** 03/30/2023
 **Date of Surgery:** 10/23/2023
 **Date of Study Inclusion:** 01/02/2024
 **ECOG Performance Status:** 0

**2. Present Illness History (PIH)**
 Patient identified a nodule in the left breast during routine self-exam in March 2023.
 Ultrasound and mammogram on 03/20/2023 revealed a 3.3 cm hypoechoic solid mass with irregular borders in the upper outer quadrant, BI-RADS 5.
 Core biopsy performed on 03/30/2023 confirmed invasive ductal carcinoma, histologic grade III.
 **Initial Clinical TNM Staging:** cT2N0M0 – Stage IIA (AJCC 8th edition)
 **Immunohistochemistry:** ER 0%, PR 0%, HER2 0, Ki-67 77%
 **Histological Subtype:** Triple-negative
 **Genetic Testing:** BRCA1 mutation c.5266dupC (deleterious), confirmed on 04/18/2023

**3. Personal and Family History**
 **Relevant Comorbidities:** None reported
 **Drug Allergies:** None known
 **Gynecological History:** Menarche at 11, nulligravida, regular cycles, no hormonal therapy
 **Family History of Cancer:** Mother with breast cancer at 44; maternal aunt with ovarian cancer
 **Prior Chest Radiotherapy:** None
 **Lifestyle Habits:** Non-smoker, does not consume alcohol

**4. Physical Examination**
 Patient in good general condition, no systemic symptoms
 **Left breast:** Firm, irregular, mobile mass in upper outer quadrant, ~3.2 cm, no skin retraction or discharge
 **Lymph Nodes:** No clinically palpable axillary or supraclavicular nodes
 No signs of lymphedema or inflammation

**5. Complementary Tests**
 **Complete Blood Count:** Hb 12.8 g/dL; leukocytes 6,600/mm³; platelets 265,000/mm³
 **Liver Function:** AST 27 U/L; ALT 29 U/L
 **Renal Function:** Creatinine 0.78 mg/dL; urea 26 mg/dL
 **Beta-HCG:** Negative
 **Serologies:** HIV, HCV, and HBsAg non-reactive
 **LVEF (Echocardiogram):** 67% pre-chemotherapy; 65% post-chemotherapy
 **Staging Imaging:**

- CT chest/abdomen/pelvis: No signs of metastasis
- Bone scan: Normal

**6. Neoadjuvant Treatment**
 **Regimen:** Platinum + taxane + anthracycline

- Paclitaxel 80 mg/m² weekly + carboplatin AUC 5 (05/01/2023 – 07/24/2023)
- Doxorubicin 60 mg/m² + cyclophosphamide 600 mg/m², 4 cycles (08/14/2023 – 10/09/2023)
   **Total Duration:** 23 weeks
   **Adverse Events:** Grade 1 nausea, grade 2 fatigue, grade 2 alopecia
   **Clinical Response:** Excellent response, tumor no longer palpable after treatment
   **Interval Between Last Chemotherapy and Surgery:** 2 weeks

**7. Surgical and Pathological Data**
 **Surgical Procedure:** Left mastectomy with sentinel lymph node biopsy
 **Incision Type:** Elliptical
 **Laterality:** Left
 **Lymph Nodes Resected:** 5 total, 0 positive
 **Residual Tumor Size:** No residual invasive carcinoma
 **Surgical Margins:** Clear
 **Histologic Grade Post-Chemotherapy:** Not applicable (pCR)
 **Lymphovascular Invasion:** Absent
 **Pathologic Response:** Complete
 **Final Pathological Staging:** ypT0N0

**PATIENT 17 — SYNTHETIC EXCLUSION RECORD**

**1. Identification**
 **Synthetic ID:** OLY-017
 **Age:** 54 years and 10 months
 **Sex / Ethnicity:** Female, Mixed race (parda)
 **Weight / Height / BMI:** 69 kg / 1.60 m / 27.0 kg/m²
 **Marital Status / Occupation:** Widowed / Rural worker
 **Treatment Center:** Hospital Universitário da Universidade Federal de Pernambuco (UFPE) – Recife, PE, Brazil
 **Date of Diagnosis:** 10/20/2022
 **Date of Surgery:** 05/15/2023
 **Date of Study Inclusion:** 08/01/2023
 **ECOG Performance Status:** 1

**2. Present Illness History (PIH)**
 Patient sought care in October 2022 with complaints of a lump and discomfort in the right breast for approximately two months.
 Ultrasound on 10/12/2022 showed a solid, irregular mass measuring 4.2 cm in the upper outer quadrant.
 Core needle biopsy on 10/20/2022 confirmed invasive ductal carcinoma, histologic grade II.
 **Initial Clinical TNM Staging:** cT2N1M0 – Stage IIB (AJCC 8th edition)
 **Immunohistochemistry:** ER 98%, PR 90%, HER2 negative (IHC 0), Ki-67 22%
 **Histological Subtype:** Luminal B-like (HER2 negative, Ki-67 >20%)
 **Genetic Testing:** BRCA2 mutation c.8537_8538delAG (deleterious), confirmed on 11/14/2022

**3. Personal and Family History**
 **Relevant Comorbidities:** Hypertension, dyslipidemia
 **Drug Allergies:** No known allergies
 **Gynecological History:** Menarche at 12, menopause at 50, 4 pregnancies, 4 deliveries, no hormone therapy
 **Family History of Cancer:** Mother with breast cancer at 63; older sister with ovarian cancer at 58
 **Prior Chest Radiotherapy:** None
 **Lifestyle Habits:** Former smoker (quit 2015); no alcohol consumption

**4. Physical Examination**
 Patient alert, oriented, no signs of systemic illness
 **Right breast:** Tumor ~4.0 cm, firm, mobile, no skin changes or nipple retraction
 **Axillary Lymph Nodes:** One palpable, mobile, ~1.5 cm
 No inflammatory signs or lymphedema

**5. Complementary Tests**
 **Complete Blood Count:** Hb 12.2 g/dL; leukocytes 7,100/mm³; platelets 255,000/mm³
 **Liver Function:** AST 31 U/L; ALT 29 U/L
 **Renal Function:** Creatinine 0.84 mg/dL; urea 28 mg/dL
 **Beta-HCG:** Negative
 **Serologies:** HIV, HCV, HBsAg negative
 **LVEF (Echocardiogram):** 65% before chemotherapy; 63% after
 **Staging Imaging:**

- CT chest/abdomen/pelvis: No distant metastases
- Bone scan: No suspicious findings

**6. Neoadjuvant Treatment**
 **Regimen:** Anthracycline + taxane

- Doxorubicin 60 mg/m² + cyclophosphamide 600 mg/m², 4 cycles (11/28/2022 – 01/23/2023)
- Paclitaxel 80 mg/m² weekly for 12 weeks (02/13/2023 – 05/08/2023)
   **Total Duration:** 24 weeks
   **Adverse Events:** Grade 2 neuropathy, grade 1 arthralgia
   **Clinical Response:** Tumor reduced by approximately 50%
   **Interval Between Last Chemotherapy and Surgery:** 1 week

**7. Surgical and Pathological Data**
 **Surgical Procedure:** Right quadrantectomy with axillary lymph node dissection
 **Incision Type:** Oblique lateral
 **Laterality:** Right
 **Lymph Nodes Resected:** 12 total, 2 positive
 **Residual Tumor Size:** 1.8 cm
 **Surgical Margins:** Free (≥5 mm)
 **Histologic Grade Post-Chemotherapy:** Grade II
 **Lymphovascular Invasion:** Absent
 **Pathologic Response:** Partial
 **Final Pathological Staging:** ypT1cN1a

**PATIENT 18 — SYNTHETIC EXCLUSION RECORD**

**1. Identification**
 **Synthetic ID:** OLY-018
 **Age:** 42 years and 6 months
 **Sex / Ethnicity:** Female, Indigenous
 **Weight / Height / BMI:** 64 kg / 1.60 m / 25.0 kg/m²
 **Marital Status / Occupation:** Single / School janitor
 **Treatment Center:** Hospital Universitário da Universidade Federal do Maranhão (HU-UFMA) – São Luís, MA, Brazil
 **Date of Diagnosis:** 02/13/2023
 **Date of Surgery:** 09/11/2023
 **Date of Study Inclusion:** 11/27/2023
 **ECOG Performance Status:** 0

**2. Present Illness History (PIH)**
 Patient reported a rapidly growing nodule in the left breast, first noticed in January 2023.
 Ultrasound and mammography performed on 02/06/2023 showed a 4.5 cm mass in the upper inner quadrant, with associated axillary adenopathy.
 Core biopsy on 02/13/2023 revealed invasive ductal carcinoma, histologic grade III.
 **Initial Clinical TNM Staging:** cT2N1M0 – Stage IIB (AJCC 8th edition)
 **Immunohistochemistry:** ER 0%, PR 0%, HER2 0, Ki-67 85%
 **Histological Subtype:** Triple-negative
 **Genetic Testing:** BRCA1 mutation c.181T>G (deleterious), confirmed on 03/02/2023

**3. Personal and Family History**
 **Relevant Comorbidities:** None reported
 **Drug Allergies:** None known
 **Gynecological History:** Menarche at 12, 3 pregnancies, 2 deliveries, 1 abortion, premenopausal, no hormone therapy
 **Family History of Cancer:** No known history of breast, ovarian, or prostate cancer
 **Prior Chest Radiotherapy:** None
 **Lifestyle Habits:** Non-smoker, no alcohol consumption

**4. Physical Examination**
 Patient in good overall health, no signs of systemic illness
 **Left breast:** Palpable, irregular, firm mass in upper inner quadrant measuring ~4.4 cm; no skin involvement
 **Lymph Nodes:** One mobile, firm axillary lymph node ~1.8 cm
 No erythema, nipple discharge, or lymphedema

**5. Complementary Tests**
 **Complete Blood Count:** Hb 12.9 g/dL; leukocytes 7,200/mm³; platelets 264,000/mm³
 **Liver Function:** AST 28 U/L; ALT 30 U/L
 **Renal Function:** Creatinine 0.86 mg/dL; urea 27 mg/dL
 **Beta-HCG:** Negative
 **Serologies:** HIV, HCV, HBsAg non-reactive
 **LVEF (Echocardiogram):** 67% before chemotherapy; 65% after
 **Staging Imaging:**

- CT chest/abdomen/pelvis: No distant metastases
- Bone scan: No evidence of bone lesions

**6. Neoadjuvant Treatment**
 **Regimen:** Platinum + taxane + anthracycline

- Weekly paclitaxel 80 mg/m² + carboplatin AUC 5 (03/13/2023 – 06/05/2023)
- Doxorubicin 60 mg/m² + cyclophosphamide 600 mg/m², 4 cycles every 21 days (06/26/2023 – 08/21/2023)
   **Total Duration:** 24 weeks
   **Adverse Events:** Grade 1 nausea, grade 2 alopecia, grade 2 anemia
   **Clinical Response:** Near-complete response; tumor no longer palpable at end of treatment
   **Interval Between Last Chemotherapy and Surgery:** 3 weeks

**7. Surgical and Pathological Data**
 **Surgical Procedure:** Left mastectomy with sentinel lymph node biopsy
 **Incision Type:** Elliptical
 **Laterality:** Left
 **Lymph Nodes Resected:** 6 total, 0 positive
 **Residual Tumor Size:** No residual invasive carcinoma
 **Surgical Margins:** Negative
 **Histologic Grade Post-Chemotherapy:** Not applicable (pCR)
 **Lymphovascular Invasion:** Absent
 **Pathologic Response:** Complete
 **Final Pathological Staging:** ypT0N0

**PATIENT 19 — SYNTHETIC EXCLUSION RECORD**

**1. Identification**
 **Synthetic ID:** OLY-019
 **Age:** 47 years and 11 months
 **Sex / Ethnicity:** Female, Black
 **Weight / Height / BMI:** 68 kg / 1.59 m / 26.9 kg/m²
 **Marital Status / Occupation:** Married / Bus driver
 **Treatment Center:** Hospital das Clínicas da Universidade Federal de Goiás (HC-UFG) – Goiânia, GO, Brazil
 **Date of Diagnosis:** 01/17/2023
 **Date of Surgery:** 08/14/2023
 **Date of Study Inclusion:** 10/20/2023
 **ECOG Performance Status:** 1

**2. Present Illness History (PIH)**
 Patient presented with a painful lump in the right breast, noticed in December 2022.
 Ultrasound and mammography on 01/10/2023 revealed a solid, irregular lesion measuring 3.6 cm, with axillary lymphadenopathy.
 Core biopsy on 01/17/2023 confirmed invasive ductal carcinoma, histologic grade III.
 **Initial Clinical TNM Staging:** cT2N1M0 – Stage IIB (AJCC 8th edition)
 **Immunohistochemistry:** ER 0%, PR 0%, HER2 0, Ki-67 90%
 **Histological Subtype:** Triple-negative
 **Genetic Testing:** BRCA1 mutation c.68_69delAG (deleterious), confirmed on 02/03/2023

**3. Personal and Family History**
 **Relevant Comorbidities:** Systemic arterial hypertension
 **Drug Allergies:** Sulfa (rash)
 **Gynecological History:** Menarche at 13, 2 pregnancies, 2 deliveries, premenopausal, no hormonal therapy
 **Family History of Cancer:** Sister with breast cancer at 39; maternal aunt with ovarian cancer
 **Prior Chest Radiotherapy:** None
 **Lifestyle Habits:** Former smoker (quit 2020), light alcohol use

**4. Physical Examination**
 Patient alert, oriented, no signs of systemic illness
 **Right breast:** Palpable mass in upper outer quadrant, firm, poorly mobile, ~3.5 cm; skin intact
 **Lymph Nodes:** One palpable axillary lymph node ~2.0 cm, mobile
 No lymphedema or inflammatory signs

**5. Complementary Tests**
 **Complete Blood Count:** Hb 12.4 g/dL; leukocytes 7,300/mm³; platelets 278,000/mm³
 **Liver Function:** AST 34 U/L; ALT 36 U/L
 **Renal Function:** Creatinine 0.89 mg/dL; urea 30 mg/dL
 **Beta-HCG:** Negative
 **Serologies:** HIV, HCV, and HBsAg non-reactive
 **LVEF (Echocardiogram):** 64% pre-chemotherapy; 62% post-chemotherapy
 **Staging Imaging:**

- CT chest/abdomen/pelvis: No evidence of metastasis
- Bone scan: Normal

**6. Neoadjuvant Treatment**
 **Regimen:** Platinum + taxane + anthracycline

- Paclitaxel 80 mg/m² weekly + carboplatin AUC 5 (02/20/2023 – 05/15/2023)
- Doxorubicin 60 mg/m² + cyclophosphamide 600 mg/m², 4 cycles (06/05/2023 – 07/31/2023)
   **Total Duration:** 24 weeks
   **Adverse Events:** Grade 2 fatigue, grade 2 neuropathy, grade 1 neutropenia
   **Clinical Response:** Tumor reduction >70%, clinically near-complete response
   **Interval Between Last Chemotherapy and Surgery:** 2 weeks

**7. Surgical and Pathological Data**
 **Surgical Procedure:** Right mastectomy with level I–II axillary dissection
 **Incision Type:** Transverse
 **Laterality:** Right
 **Lymph Nodes Resected:** 13 total, 1 positive
 **Residual Tumor Size:** 0.7 cm
 **Surgical Margins:** Free (≥5 mm)
 **Histologic Grade Post-Chemotherapy:** Grade II
 **Lymphovascular Invasion:** Absent
 **Pathologic Response:** Near-complete (residual minimal disease)
 **Final Pathological Staging:** ypT1bN1mi

**PATIENT 20 — SYNTHETIC EXCLUSION RECORD**

**1. Identification**
 **Synthetic ID:** OLY-020
 **Age:** 36 years and 9 months
 **Sex / Ethnicity:** Female, White
 **Weight / Height / BMI:** 59 kg / 1.61 m / 22.8 kg/m²
 **Marital Status / Occupation:** Single / Physical education teacher
 **Treatment Center:** Hospital de Câncer de Pernambuco (HCP) – Recife, PE, Brazil
 **Date of Diagnosis:** 06/05/2023
 **Date of Surgery:** 01/15/2024
 **Date of Study Inclusion:** 03/25/2024
 **ECOG Performance Status:** 0

**2. Present Illness History (PIH)**
 Patient reported detection of a lump in the left breast during self-exam in May 2023.
 Ultrasound and mammography on 05/25/2023 identified a 3.1 cm irregular solid lesion in the lower outer quadrant, with one suspicious axillary node.
 Core needle biopsy on 06/05/2023 confirmed invasive ductal carcinoma, histologic grade III.
 **Initial Clinical TNM Staging:** cT2N1M0 – Stage IIB (AJCC 8th edition)
 **Immunohistochemistry:** ER 0%, PR 0%, HER2 0, Ki-67 86%
 **Histological Subtype:** Triple-negative
 **Genetic Testing:** BRCA1 mutation c.4035delA (deleterious), confirmed on 06/26/2023

**3. Personal and Family History**
 **Relevant Comorbidities:** None reported
 **Drug Allergies:** No known drug allergies
 **Gynecological History:** Menarche at 12, nulligravida, regular cycles, no hormonal contraceptives
 **Family History of Cancer:** Mother with breast cancer at age 42; maternal grandmother with ovarian cancer
 **Prior Chest Radiotherapy:** None
 **Lifestyle Habits:** Non-smoker, physically active, no alcohol use

**4. Physical Examination**
 Patient in excellent general condition, asymptomatic
 **Left breast:** Firm, mobile nodule ~3.0 cm in lower outer quadrant; skin and nipple normal
 **Axillary Lymph Nodes:** One mobile, firm node ~1.7 cm
 No inflammatory signs, lymphedema, or pain

**5. Complementary Tests**
 **Complete Blood Count:** Hb 13.1 g/dL; leukocytes 6,800/mm³; platelets 259,000/mm³
 **Liver Function:** AST 25 U/L; ALT 24 U/L
 **Renal Function:** Creatinine 0.77 mg/dL; urea 26 mg/dL
 **Beta-HCG:** Negative
 **Serologies:** HIV, HCV, and HBsAg non-reactive
 **LVEF (Echocardiogram):** 68% before chemotherapy; 66% after
 **Staging Imaging:**

- CT chest/abdomen/pelvis: No metastases
- Bone scan: Normal

**6. Neoadjuvant Treatment**

**Regimen:** Platinum + taxane + anthracycline

- Paclitaxel 80 mg/m² weekly + carboplatin AUC 5 (07/10/2023 – 10/02/2023)
- Doxorubicin 60 mg/m² + cyclophosphamide 600 mg/m², 4 cycles (10/23/2023 – 12/18/2023)
   **Total Duration:** 24 weeks
   **Adverse Events:** Grade 2 alopecia, grade 1 nausea, no neutropenia
   **Clinical Response:** Complete clinical response by imaging
   **Interval Between Last Chemotherapy and Surgery:** 4 weeks

**7. Surgical and Pathological Data**
 **Surgical Procedure:** Left mastectomy with sentinel lymph node biopsy
 **Incision Type:** Oblique
 **Laterality:** Left
 **Lymph Nodes Resected:** 6 total, 0 positive
 **Residual Tumor Size:** No residual invasive carcinoma
 **Surgical Margins:** Clear
 **Histologic Grade Post-Chemotherapy:** Not applicable (pCR)
 **Lymphovascular Invasion:** Absent
 **Pathologic Response:** Complete
 **Final Pathological Staging:** ypT0N0

**PATIENT 21 — SYNTHETIC EXCLUSION RECORD**

**1. Identification**
 **Synthetic ID:** OLY-021
 **Age:** 49 years and 2 months
 **Sex / Ethnicity:** Female, Mixed race (parda)
 **Weight / Height / BMI:** 70 kg / 1.63 m / 26.4 kg/m²
 **Marital Status / Occupation:** Married / Supermarket stock clerk
 **Treatment Center:** Hospital Universitário Júlio Müller – UFMT, Cuiabá, MT, Brazil
 **Date of Diagnosis:** 07/03/2023
 **Date of Surgery:** 02/05/2024
 **Date of Study Inclusion:** 04/11/2024
 **ECOG Performance Status:** 0

**2. Present Illness History (PIH)**
 Patient presented with a self-detected lump in the right breast in June 2023.
 Ultrasound and mammography on 06/28/2023 showed a 3.4 cm irregular mass in the upper outer quadrant, with suspicious ipsilateral axillary node.
 Core biopsy on 07/03/2023 confirmed invasive ductal carcinoma, histologic grade II.
 **Initial Clinical TNM Staging:** cT2N1M0 – Stage IIB (AJCC 8th edition)
 **Immunohistochemistry:** ER 92%, PR 70%, HER2 negative (IHC 0), Ki-67 28%
 **Histological Subtype:** Luminal B-like (HER2 negative)
 **Genetic Testing:** BRCA2 mutation c.5946delT (deleterious), confirmed on 07/25/2023

**3. Personal and Family History**
 **Relevant Comorbidities:** Hypertension (on losartan), obesity
 **Drug Allergies:** None reported
 **Gynecological History:** Menarche at 13, 3 pregnancies, 3 deliveries, menopause at 48
 **Family History of Cancer:** Sister with breast cancer at age 53; maternal uncle with colorectal cancer
 **Prior Chest Radiotherapy:** None
 **Lifestyle Habits:** Non-smoker, occasional alcohol consumption

**4. Physical Examination**
 Patient in good general health, no weight loss or systemic symptoms
 **Right breast:** Firm, partially mobile tumor ~3.3 cm in upper outer quadrant, no skin changes or nipple discharge
 **Lymph Nodes:** One mobile axillary lymph node ~1.5 cm
 No signs of inflammation, edema, or lymphedema

**5. Complementary Tests**
 **Complete Blood Count:** Hb 12.7 g/dL; leukocytes 7,000/mm³; platelets 268,000/mm³
 **Liver Function:** AST 29 U/L; ALT 27 U/L
 **Renal Function:** Creatinine 0.84 mg/dL; urea 25 mg/dL
 **Beta-HCG:** Negative
 **Serologies:** Non-reactive for HIV, HCV, and HBsAg
 **LVEF (Echocardiogram):** 66% before chemotherapy; 64% after
 **Staging Imaging:**

- CT chest/abdomen/pelvis: No evidence of metastasis
- Bone scan: Normal

**6. Neoadjuvant Treatment**
 **Regimen:** Anthracycline + taxane

- Doxorubicin 60 mg/m² + cyclophosphamide 600 mg/m², 4 cycles every 21 days (08/07/2023 – 10/02/2023)
- Docetaxel 75 mg/m², 4 cycles every 21 days (10/23/2023 – 12/18/2023)
   **Total Duration:** 20 weeks
   **Adverse Events:** Grade 2 arthralgia, grade 1 constipation, no hematologic toxicity
   **Clinical Response:** Partial response (~50% reduction)
   **Interval Between Last Chemotherapy and Surgery:** 7 weeks

**7. Surgical and Pathological Data**
 **Surgical Procedure:** Right quadrantectomy with sentinel lymph node biopsy
 **Incision Type:** Oblique upper lateral
 **Laterality:** Right
 **Lymph Nodes Resected:** 5 total, 1 positive
 **Residual Tumor Size:** 1.5 cm
 **Surgical Margins:** Negative (≥3 mm)
 **Histologic Grade Post-Chemotherapy:** Grade II
 **Lymphovascular Invasion:** Present
 **Pathologic Response:** Partial
 **Final Pathological Staging:** ypT1cN1mi

**PATIENT 22 — SYNTHETIC EXCLUSION RECORD**

**1. Identification**
 **Synthetic ID:** OLY-022
 **Age:** 41 years and 10 months
 **Sex / Ethnicity:** Female, White
 **Weight / Height / BMI:** 61 kg / 1.62 m / 23.2 kg/m²
 **Marital Status / Occupation:** Married / Graphic designer
 **Treatment Center:** Hospital Universitário da Universidade Federal do Rio Grande do Sul (HCPA-UFRGS) – Porto Alegre, RS, Brazil
 **Date of Diagnosis:** 09/04/2023
 **Date of Surgery:** 03/18/2024
 **Date of Study Inclusion:** 05/10/2024
 **ECOG Performance Status:** 0

**2. Present Illness History (PIH)**
 Patient presented with a palpable lump in the left breast in August 2023.
 Ultrasound and mammography on 08/28/2023 showed a 2.9 cm irregular mass in the upper inner quadrant and no enlarged axillary lymph nodes.
 Core biopsy on 09/04/2023 revealed invasive ductal carcinoma, histologic grade III.
 **Initial Clinical TNM Staging:** cT2N0M0 – Stage IIA (AJCC 8th edition)
 **Immunohistochemistry:** ER 0%, PR 0%, HER2 0, Ki-67 82%
 **Histological Subtype:** Triple-negative
 **Genetic Testing:** BRCA1 mutation c.3700_3704delGTAAA (deleterious), confirmed on 09/26/2023

**3. Personal and Family History**
 **Relevant Comorbidities:** None reported
 **Drug Allergies:** No known allergies
 **Gynecological History:** Menarche at 12, 2 pregnancies, 2 deliveries, premenopausal, no use of hormone therapy
 **Family History of Cancer:** Maternal aunt with ovarian cancer; mother with breast cancer at age 55
 **Prior Chest Radiotherapy:** None
 **Lifestyle Habits:** Non-smoker, regular physical activity, does not consume alcohol

**4. Physical Examination**
 Patient in good general condition, no systemic symptoms
 **Left breast:** Solid, mobile mass ~2.8–3.0 cm in upper inner quadrant; no skin retraction or nipple discharge
 **Lymph Nodes:** No palpable axillary or supraclavicular nodes
 No signs of lymphedema or inflammation

**5. Complementary Tests**
 **Complete Blood Count:** Hb 13.0 g/dL; leukocytes 6,400/mm³; platelets 260,000/mm³
 **Liver Function:** AST 25 U/L; ALT 27 U/L
 **Renal Function:** Creatinine 0.79 mg/dL; urea 24 mg/dL
 **Beta-HCG:** Negative
 **Serologies:** HIV, HCV, and HBsAg non-reactive
 **LVEF (Echocardiogram):** 68% before chemotherapy; 65% after
 **Staging Imaging:**

- Chest/abdomen/pelvis CT: No evidence of metastasis
- Bone scan: Normal

**6. Neoadjuvant Treatment**
 **Regimen:** Platinum + taxane + anthracycline

- Paclitaxel 80 mg/m² weekly + carboplatin AUC 5 (10/02/2023 – 12/25/2023)
- Doxorubicin 60 mg/m² + cyclophosphamide 600 mg/m², 4 cycles (01/15/2024 – 03/11/2024)
   **Total Duration:** 24 weeks
   **Adverse Events:** Grade 2 nausea, grade 2 alopecia, grade 1 mucositis
   **Clinical Response:** Clinical complete response, tumor no longer palpable by end of treatment
   **Interval Between Last Chemotherapy and Surgery:** 1 week

**7. Surgical and Pathological Data**
 **Surgical Procedure:** Left mastectomy with sentinel lymph node biopsy
 **Incision Type:** Elliptical
 **Laterality:** Left
 **Lymph Nodes Resected:** 5 total, 0 positive
 **Residual Tumor Size:** No residual invasive carcinoma
 **Surgical Margins:** Negative
 **Histologic Grade Post-Chemotherapy:** Not applicable (pCR)
 **Lymphovascular Invasion:** Absent
 **Pathologic Response:** Complete
 **Final Pathological Staging:** ypT0N0

**PATIENT 23 — SYNTHETIC EXCLUSION RECORD**

**1. Identification**
 **Synthetic ID:** OLY-023
 **Age:** 52 years and 5 months
 **Sex / Ethnicity:** Female, Black
 **Weight / Height / BMI:** 75 kg / 1.65 m / 27.5 kg/m²
 **Marital Status / Occupation:** Married / Cleaning professional
 **Treatment Center:** Hospital Universitário da Universidade Federal do Espírito Santo (HUCAM-UFES) – Vitória, ES, Brazil
 **Date of Diagnosis:** 05/16/2023
 **Date of Surgery:** 12/04/2023
 **Date of Study Inclusion:** 02/01/2024
 **ECOG Performance Status:** 1

**2. Present Illness History (PIH)**
 Patient sought evaluation due to a palpable mass in the right breast noted during bathing in early May 2023.
 Ultrasound and mammography on 05/09/2023 revealed a 3.6 cm irregular mass in the upper outer quadrant, associated with ipsilateral axillary lymphadenopathy.
 Core needle biopsy on 05/16/2023 confirmed invasive ductal carcinoma, histologic grade II.
 **Initial Clinical TNM Staging:** cT2N1M0 – Stage IIB (AJCC 8th edition)
 **Immunohistochemistry:** ER 95%, PR 80%, HER2 negative (IHC 1+), Ki-67 26%
 **Histological Subtype:** Luminal B-like (HER2 negative)
 **Genetic Testing:** BRCA2 mutation c.9097dupA (deleterious), confirmed on 06/06/2023

**3. Personal and Family History**
 **Relevant Comorbidities:** Hypertension, type 2 diabetes mellitus (on metformin and losartan)
 **Drug Allergies:** None reported
 **Gynecological History:** Menarche at 13, menopause at 50, 4 pregnancies, 4 deliveries, no hormone therapy
 **Family History of Cancer:** Older sister with breast cancer at 59; maternal cousin with prostate cancer
 **Prior Chest Radiotherapy:** None
 **Lifestyle Habits:** Non-smoker, no alcohol use

**4. Physical Examination**
 Patient in good general condition, reports occasional fatigue
 **Right breast:** Firm, irregular mass ~3.5 cm in upper outer quadrant; overlying skin intact
 **Axillary Lymph Nodes:** One palpable, firm, mobile node ~1.8 cm
 No signs of inflammation, discharge, or lymphedema

**5. Complementary Tests**
 **Complete Blood Count:** Hb 12.5 g/dL; leukocytes 7,200/mm³; platelets 261,000/mm³
 **Liver Function:** AST 30 U/L; ALT 28 U/L
 **Renal Function:** Creatinine 0.91 mg/dL; urea 27 mg/dL
 **Beta-HCG:** Negative
 **Serologies:** HIV, HCV, and HBsAg non-reactive
 **LVEF (Echocardiogram):** 65% pre-chemotherapy; 63% post-chemotherapy
 **Staging Imaging:**

- CT chest/abdomen/pelvis: No evidence of metastases
- Bone scintigraphy: Normal

**6. Neoadjuvant Treatment**
 **Regimen:** Anthracycline followed by taxane

- Doxorubicin 60 mg/m² + cyclophosphamide 600 mg/m², 4 cycles (06/19/2023 – 08/14/2023)
- Docetaxel 75 mg/m², 4 cycles (09/04/2023 – 10/30/2023)
   **Total Duration:** 20 weeks
   **Adverse Events:** Grade 2 arthralgia, grade 1 peripheral edema
   **Clinical Response:** Partial reduction (~50%)
   **Interval Between Last Chemotherapy and Surgery:** 5 weeks

**7. Surgical and Pathological Data**
 **Surgical Procedure:** Right quadrantectomy with axillary dissection
 **Incision Type:** Upper lateral oblique
 **Laterality:** Right
 **Lymph Nodes Resected:** 14 total, 2 positive
 **Residual Tumor Size:** 1.9 cm
 **Surgical Margins:** Negative (≥4 mm)
 **Histologic Grade Post-Chemotherapy:** Grade II
 **Lymphovascular Invasion:** Present
 **Pathologic Response:** Partial
 **Final Pathological Staging:** ypT1cN1a

**PATIENT 24 — SYNTHETIC EXCLUSION RECORD**

**1. Identification**
 **Synthetic ID:** OLY-024
 **Age:** 38 years and 4 months
 **Sex / Ethnicity:** Female, White
 **Weight / Height / BMI:** 58 kg / 1.60 m / 22.7 kg/m²
 **Marital Status / Occupation:** Single / Lawyer
 **Treatment Center:** Instituto do Câncer do Estado de São Paulo (ICESP) – São Paulo, SP, Brazil
 **Date of Diagnosis:** 04/11/2023
 **Date of Surgery:** 11/13/2023
 **Date of Study Inclusion:** 01/29/2024
 **ECOG Performance Status:** 0

**2. Present Illness History (PIH)**
 Patient presented after noticing a lump in the upper outer quadrant of the left breast in late March 2023.
 Imaging studies (US and mammography) performed on 04/05/2023 showed a 2.7 cm hypoechoic mass with irregular borders; BI-RADS 5.
 Core needle biopsy on 04/11/2023 confirmed invasive ductal carcinoma, histologic grade III.
 **Initial Clinical TNM Staging:** cT2N0M0 – Stage IIA (AJCC 8th edition)
 **Immunohistochemistry:** ER 0%, PR 0%, HER2 0, Ki-67 84%
 **Histological Subtype:** Triple-negative
 **Genetic Testing:** BRCA1 mutation c.1961delA (deleterious), confirmed on 04/28/2023

**3. Personal and Family History**
 **Relevant Comorbidities:** None reported
 **Drug Allergies:** No known allergies
 **Gynecological History:** Menarche at 12, nulligravida, regular menstrual cycles, no hormonal therapy
 **Family History of Cancer:** Mother with breast cancer at age 45; maternal aunt with ovarian cancer
 **Prior Chest Radiotherapy:** None
 **Lifestyle Habits:** Non-smoker, does not consume alcohol, physically active

**4. Physical Examination**
 Patient in good general condition, no systemic symptoms
 **Left breast:** Firm, mobile nodule in upper outer quadrant, ~2.8 cm, no skin retraction or discharge
 **Lymph Nodes:** No palpable axillary or supraclavicular nodes
 No signs of inflammation, pain, or lymphedema

**5. Complementary Tests**
 **Complete Blood Count:** Hb 13.3 g/dL; leukocytes 6,900/mm³; platelets 262,000/mm³
 **Liver Function:** AST 24 U/L; ALT 26 U/L
 **Renal Function:** Creatinine 0.79 mg/dL; urea 25 mg/dL
 **Beta-HCG:** Negative
 **Serologies:** HIV, HCV, and HBsAg non-reactive
 **LVEF (Echocardiogram):** 69% before chemotherapy; 67% after
 **Staging Imaging:**

- CT chest/abdomen/pelvis: No signs of metastatic disease
- Bone scintigraphy: Normal

**6. Neoadjuvant Treatment**
 **Regimen:** Platinum + taxane + anthracycline

- Paclitaxel 80 mg/m² weekly + carboplatin AUC 5 (05/08/2023 – 07/31/2023)
- Doxorubicin 60 mg/m² + cyclophosphamide 600 mg/m², 4 cycles (08/21/2023 – 10/16/2023)
   **Total Duration:** 24 weeks
   **Adverse Events:** Grade 2 nausea, grade 2 alopecia, no hematologic toxicity
   **Clinical Response:** Excellent clinical response; mass no longer palpable after treatment
   **Interval Between Last Chemotherapy and Surgery:** 4 weeks

**7. Surgical and Pathological Data**
 **Surgical Procedure:** Left mastectomy with sentinel lymph node biopsy
 **Incision Type:** Oblique inferolateral
 **Laterality:** Left
 **Lymph Nodes Resected:** 6 total, 0 positive
 **Residual Tumor Size:** No residual invasive carcinoma
 **Surgical Margins:** Negative
 **Histologic Grade Post-Chemotherapy:** Not applicable (pCR)
 **Lymphovascular Invasion:** Absent
 **Pathologic Response:** Complete
 **Final Pathological Staging:** ypT0N0

**PATIENT 25 — SYNTHETIC EXCLUSION RECORD**

**1. Identification**
 **Synthetic ID:** OLY-025
 **Age:** 45 years and 1 month
 **Sex / Ethnicity:** Female, Mixed race (parda)
 **Weight / Height / BMI:** 72 kg / 1.62 m / 27.4 kg/m²
 **Marital Status / Occupation:** Divorced / Taxi driver
 **Treatment Center:** Hospital Universitário Antônio Pedro – UFF, Niterói, RJ, Brazil
 **Date of Diagnosis:** 08/22/2023
 **Date of Surgery:** 03/11/2024
 **Date of Study Inclusion:** 05/06/2024
 **ECOG Performance Status:** 1

**2. Present Illness History (PIH)**
 Patient noticed a growing lump in the right breast during self-exam in August 2023.
 Ultrasound and mammography on 08/15/2023 identified a 4.1 cm irregular hypoechoic mass and an enlarged axillary lymph node on the same side.
 Core biopsy on 08/22/2023 confirmed invasive ductal carcinoma, histologic grade III.
 **Initial Clinical TNM Staging:** cT2N1M0 – Stage IIB (AJCC 8th edition)
 **Immunohistochemistry:** ER 0%, PR 0%, HER2 0, Ki-67 90%
 **Histological Subtype:** Triple-negative
 **Genetic Testing:** BRCA1 mutation c.5263insC (deleterious), confirmed on 09/12/2023

**3. Personal and Family History**
 **Relevant Comorbidities:** Obesity, systemic arterial hypertension
 **Drug Allergies:** Dipyrone (rash)
 **Gynecological History:** Menarche at 11, 3 pregnancies, 2 deliveries, 1 abortion, no hormone therapy
 **Family History of Cancer:** Mother with breast cancer at 48; maternal grandmother with ovarian cancer
 **Prior Chest Radiotherapy:** None
 **Lifestyle Habits:** Smoker (10 cigarettes/day); no alcohol use

**4. Physical Examination**
 Patient in regular general condition; mild fatigue
 **Right breast:** Firm, poorly mobile mass in upper outer quadrant measuring ~4.0 cm; skin and nipple intact
 **Axillary Lymph Nodes:** One palpable, firm, mobile node ~2.1 cm
 No signs of inflammation or lymphedema

**5. Complementary Tests**
 **Complete Blood Count:** Hb 12.0 g/dL; leukocytes 7,500/mm³; platelets 275,000/mm³
 **Liver Function:** AST 34 U/L; ALT 36 U/L
 **Renal Function:** Creatinine 0.88 mg/dL; urea 29 mg/dL
 **Beta-HCG:** Negative
 **Serologies:** HIV, HCV, and HBsAg non-reactive
 **LVEF (Echocardiogram):** 63% before chemotherapy; 61% after
 **Staging Imaging:**

- CT chest/abdomen/pelvis: No evidence of metastasis
- Bone scan: Normal

**6. Neoadjuvant Treatment**
 **Regimen:** Platinum + taxane + anthracycline

- Paclitaxel 80 mg/m² weekly + carboplatin AUC 5 (09/18/2023 – 12/11/2023)
- Doxorubicin 60 mg/m² + cyclophosphamide 600 mg/m², 4 cycles (01/02/2024 – 02/26/2024)
   **Total Duration:** 24 weeks
   **Adverse Events:** Grade 2 nausea, grade 2 fatigue, grade 1 anemia
   **Clinical Response:** Tumor reduced by >70%; no palpable mass after last cycle
   **Interval Between Last Chemotherapy and Surgery:** 2 weeks

**7. Surgical and Pathological Data**
 **Surgical Procedure:** Right mastectomy with level I–II axillary dissection
 **Incision Type:** Oblique inferolateral
 **Laterality:** Right
 **Lymph Nodes Resected:** 15 total, 1 positive
 **Residual Tumor Size:** 0.6 cm
 **Surgical Margins:** Negative
 **Histologic Grade Post-Chemotherapy:** Grade III
 **Lymphovascular Invasion:** Absent
 **Pathologic Response:** Near-complete (residual minimal disease)
 **Final Pathological Staging:** ypT1bN1mi

**FINAL PROMPT FOR PATIENT SENTENCE GENERATION (P2S)**

**CONTEXT:** You are a specialist in Natural Language Processing (NLP) and information extraction, focusing on oncology data.

**GOAL:** Process an electronic medical record to generate a "patient sentence" (P2S). The goal of this sentence is to create a concise, structured clinical “fingerprint” that accelerates initial screening for clinical trials. The sentence should be a factual and chronological representation of the patient’s positive clinical data.

**INPUT:** A patient’s electronic medical record file.

**OUTPUT FORMAT:** A single text string, with each clinical detail separated by the character |.

**EXTRACTION AND FORMATTING RULES:**

**Mandatory Start:** The sentence MUST begin with the location of the primary tumor.

**Chronological Order:** Events must be listed from oldest to most recent.

**Date Format:** Each event or report MUST include its date in the format (DD/MM/YYYY).

**"Key: Value" Structure:** Use a Key: Value format for all information.

**Variable Sentence Length, Focus on Positive Data:** The sentence will vary in length. Include only positive findings and data. Do NOT include negative results (e.g., negative genetic tests), minor comorbidities, or incidental findings.

**Maximum Detail for Reports:**

- **Pathology:** Extract all details: histological type, grade, tumor size (Tu), margin status, angiolymphatic and perineural invasion.
- **Immunohistochemistry (IHC):** Extract exact percentages for ER, PR, and Ki-67, and HER2 score.
- **Genetics:** Transcribe the full results of genetic tests (e.g., BRCA1/2, MLPA).

**Handling Dynamic Data:**

- **Clinical Data (e.g., ECOG status):** Include ONLY the most recent record.
- **Tumor Markers (e.g., CEA, CA 15-3):** Include the baseline AND most recent value. Use the format: “Marker: Initial_Value (DD/MM/YYYY) -> Recent_Value (DD/MM/YYYY)”.

**Essential Information List (Extract if present and positive):**

- **Diagnosis:** Histological type, grade, size, invasions (angiolymphatic/perineural), etc. (with date).
- **Pathological Staging:** pTNM (with date).
- **IHC:** ER, PR, HER2, Ki-67 (with date).
- **Genetic Tests:** Test result (with date).
- **Surgery(ies):** Procedure type, margin status (with date).
- **Systemic Treatments (CT, HT, etc.):** Protocol/medication and period (start and end).
- **Radiotherapy (RT):** Period (start and end).
- **Performance Status:** Latest ECOG (with date).
- **Tumor Markers:** First and last value (with dates).
- **Major Comorbidities:** Only those listed as significant prior conditions (e.g., Hypertension, Diabetes).

**FINAL OUTPUT EXAMPLE:**

*Right Breast | Diagnosis: Invasive ductal carcinoma NST, Histologic grade II, Size: 2.0 cm, Margins: Clear, Angiolymphatic/peritumoral embolization: Not detected (26/08/2019) | Pathological Staging: pT1c pN1 pMx (26/08/2019) | IHC: ER positive 100%, PR positive 100%, HER2 score 0, Ki67 positive 10% (18/09/2019) | Adjuvant CT: TC protocol (Start: NOV/2019 - End: 16/01/2020) | Adjuvant RT: (Start: 17/02/2020 - End: 10/03/2020) | Adjuvant HT: Anastrozole (Start: MAR/2020) | Bone Therapy: Zoledronic Acid every 6 months (Start: MAR/2020) | Comorbidities: Systemic arterial hypertension | Tumor Marker CA15-3: 15.7 U/ml (31/10/2019) -> 16.7 U/ml (05/01/2024) | Tumor Marker CEA: 1.6 ng/ml (31/10/2019) | ECOG: 0 (22/04/2024)*

**SYNTHESIS OF SYNTHETIC PATIENTS - OLYMPIA**

**PATIENT 1 — SYNTHETIC ENROLLMENT RECORD**
 Left Breast | Diagnosis: Invasive ductal carcinoma, Histologic grade III (Nottingham 8/9), Size: 3.8 cm, Lymphovascular invasion present (04/17/2023) | Genetic Test: Germline BRCA1 mutation c.5266dupC (deleterious) confirmed (05/05/2023) | Neoadjuvant CT: AC-T protocol – Doxorubicin 60 mg/m² + Cyclophosphamide 600 mg/m² (05/15/2023 – 07/10/2023) and Docetaxel 75 mg/m² (07/31/2023 – 09/25/2023), total duration 20 weeks, partial clinical response (~40% tumor reduction) | Surgery: Left quadrantectomy + Axillary lymph node dissection levels I–II, clear margins ≥5 mm, 15 lymph nodes resected (3 positive), residual tumor 1.8 cm, Post-CT histologic grade II, partial pathological response (11/10/2023) | Pathological Staging: ypT1cN1aM0 (11/10/2023) | IHC: ER 0%, PR 0%, HER2 0, Ki-67 65% (04/17/2023) | Comorbidities: Systemic arterial hypertension | ECOG: 0 (02/20/2024)

**PATIENT 2 — SYNTHETIC ENROLLMENT RECORD**
 Right Breast | Diagnosis: Invasive ductal carcinoma, Histologic grade II (Nottingham 7/9), Size: 3.5 cm, Lymphovascular invasion present (03/09/2023) | Genetic Test: Germline BRCA2 mutation c.5946delT (deleterious) confirmed (04/18/2023) | Adjuvant CT: AC protocol followed by Paclitaxel – Doxorubicin 60 mg/m² + Cyclophosphamide 600 mg/m² (04/10/2023 – 06/05/2023) and weekly Paclitaxel 80 mg/m² (06/19/2023 – 09/04/2023), total duration 24 weeks, adverse events: grade 1 peripheral neuropathy, grade 2 fatigue, grade 2 neutropenia | Surgery: Right quadrantectomy + Axillary lymph node dissection levels I–II, clear margins ≥3 mm, 18 lymph nodes resected (2 positive), residual tumor 2.0 cm, Histologic grade II, partial pathological response (12/02/2023) | Pathological Staging: ypT2N1aM0 (12/02/2023) | IHC: ER 95%, PR 90%, HER2 1+, Ki-67 30% (03/09/2023) | Planned HT: Anastrozole 1 mg/day (to start after RT) | Comorbidities: Dyslipidemia | ECOG: 1 (02/28/2024)

**PATIENT 3 — SYNTHETIC ENROLLMENT RECORD**
 Left Breast | Diagnosis: Invasive ductal carcinoma, Histologic grade III (Nottingham 8/9), Size: 2.9 cm (05/11/2023) | Genetic Test: Germline BRCA1 mutation c.68_69delAG (deleterious) confirmed (06/20/2023) | Neoadjuvant CT: Carboplatin + Paclitaxel → AC protocol – Carboplatin AUC6 + weekly Paclitaxel 80 mg/m² (06/19/2023 – 09/04/2023) followed by Doxorubicin 60 mg/m² + Cyclophosphamide 600 mg/m² (09/18/2023 – 11/13/2023), total duration 24 weeks, partial clinical response (~60% tumor reduction) | Surgery: Left simple mastectomy + Axillary lymph node dissection levels I–II, clear margins ≥7 mm, 14 lymph nodes resected (2 positive), residual tumor 0.9 cm, Post-CT histologic grade II, near-complete pathological response (12/15/2023) | Pathological Staging: ypT1bN1aM0 (12/15/2023) | IHC: ER 0%, PR 0%, HER2 0, Ki-67 70% (05/11/2023) | ECOG: 0 (03/10/2024)

**PATIENT 4 — SYNTHETIC ENROLLMENT RECORD**
 Right Breast | Diagnosis: Invasive ductal carcinoma, Histologic grade II (Nottingham 7/9), Size: 4.0 cm, Lymphovascular invasion present (04/03/2023) | Genetic Test: Germline BRCA2 mutation c.8537_8538delAG (deleterious) confirmed (05/12/2023) | Adjuvant CT: AC protocol followed by Paclitaxel – Doxorubicin 60 mg/m² + Cyclophosphamide 600 mg/m² (05/08/2023 – 07/03/2023) and weekly Paclitaxel 80 mg/m² (07/17/2023 – 10/09/2023), total duration 22 weeks, adverse events: grade 2 fatigue, grade 1 peripheral neuropathy, grade 1 nausea | Surgery: Right quadrantectomy + Axillary lymph node dissection levels I–II, clear margins ≥4 mm, 16 lymph nodes resected (2 positive), residual tumor 2.3 cm, Histologic grade II, partial pathological response (11/20/2023) | Pathological Staging: ypT2N1aM0 (11/20/2023) | IHC: ER 80%, PR 60%, HER2 0, Ki-67 35% (04/03/2023) | Planned HT: Letrozole 2.5 mg/day (to start after RT) | Comorbidities: Systemic arterial hypertension | ECOG: 0 (03/05/2024)

**PATIENT 5 — SYNTHETIC ENROLLMENT RECORD**
 Left Breast | Diagnosis: Invasive ductal carcinoma, Histologic grade III (Nottingham 8/9), Size: 3.1 cm (02/20/2023) | Genetic Test: Germline BRCA1 mutation c.4035delA (deleterious) confirmed (04/04/2023) | Neoadjuvant CT: Carboplatin + Paclitaxel → AC protocol – Carboplatin AUC6 + weekly Paclitaxel 80 mg/m² (04/03/2023 – 06/26/2023) followed by Doxorubicin 60 mg/m² + Cyclophosphamide 600 mg/m² (07/10/2023 – 08/28/2023), total duration 23 weeks, significant clinical response (~80% tumor reduction) | Surgery: Left simple mastectomy + Axillary lymph node dissection levels I–II, clear margins ≥10 mm, 17 lymph nodes resected (1 positive), residual tumor 0.4 cm, Post-CT histologic grade II, near-complete pathological response (09/25/2023) | Pathological Staging: ypT1aN1aM0 (09/25/2023) | IHC: ER 0%, PR 0%, HER2 0, Ki-67 80% (02/20/2023) | ECOG: 0 (02/15/2024)

**PATIENT 6 — SYNTHETIC EXCLUSION RECORD**

Right Breast | Diagnosis: Invasive ductal carcinoma, Histologic grade II (Nottingham 7/9), Size: 3.6 cm, Lymphovascular invasion present (02/28/2023) | IHC: ER 95%, PR 90%, HER2 2+ (FISH positive), Ki-67 35% (02/28/2023) | Genetic Test: BRCA1/BRCA2 – no mutations detected (06/02/2023) | Neoadjuvant CT: TCHP protocol – Docetaxel 75 mg/m² + Carboplatin AUC6 + Trastuzumab + Pertuzumab (6 cycles, 04/17/2023 – 09/11/2023), adverse events: grade 2 diarrhea, grade 2 fatigue, grade 3 neutropenia (treated with filgrastim), partial clinical response (~50%) | Surgery: Right modified radical mastectomy, clear margins ≥3 mm, 19 lymph nodes resected (4 positive), residual tumor 1.7 cm, Histologic grade II, lymphovascular invasion present (10/10/2023) | Pathological Staging: ypT1cN2aM0 (10/10/2023) | Comorbidities: Type 2 diabetes mellitus, Systemic arterial hypertension | ECOG: 1 (03/05/2024)

**PATIENT 7 — SYNTHETIC EXCLUSION RECORD**

Left Breast | Diagnosis: Invasive ductal carcinoma, Histologic grade II (Nottingham 7/9), Size: 4.5 cm (03/15/2023) | IHC: ER 90%, PR 85%, HER2 1+, Ki-67 25% (03/15/2023) | Genetic Test: BRCA1/BRCA2 negative (04/25/2023) | Adjuvant CT: AC protocol followed by Paclitaxel – Doxorubicin 60 mg/m² + Cyclophosphamide 600 mg/m² (05/01/2023 – 06/26/2023) and weekly Paclitaxel 80 mg/m² (07/10/2023 – 09/25/2023), total duration 24 weeks, mild adverse events (fatigue, neuropathy, grade 1 nausea) | Surgery: Left quadrantectomy + axillary lymph node dissection levels I–II, clear margins ≥3 mm, 17 lymph nodes resected (2 positive), residual tumor 2.2 cm, Histologic grade II, partial pathological response (12/05/2023) | Pathological Staging: ypT2N1aM0 (12/05/2023) | HT: Anastrozole 1 mg/day (started 12/15/2023) | Comorbidities: Systemic arterial hypertension, Dyslipidemia | ECOG: 1 (02/20/2024)

**PATIENT 8 — SYNTHETIC EXCLUSION RECORD**

Right Breast | Diagnosis: Invasive ductal carcinoma, Histologic grade III (Nottingham 8/9), Size: 3.2 cm, Lymphovascular invasion present (05/05/2023) | IHC: ER 0%, PR 0%, HER2 3+ (positive), Ki-67 55% (05/05/2023) | Genetic Test: BRCA1/BRCA2 negative (06/10/2023) | Neoadjuvant CT: TCHP protocol – Docetaxel + Carboplatin + Trastuzumab + Pertuzumab (6 cycles, 06/21/2023 – 10/02/2023), adverse events: grade 2 diarrhea, grade 2 neutropenia, grade 1 mucositis, partial clinical response (~50%) | Surgery: Right quadrantectomy + axillary lymph node dissection levels I–II, clear margins ≥4 mm, 16 lymph nodes resected (3 positive), residual tumor 1.6 cm, Histologic grade II, partial pathological response (12/14/2023) | Pathological Staging: ypT1cN1aM0 (12/14/2023) | Comorbidities: Mild dyslipidemia | ECOG: 1 (02/22/2024)

**PATIENT 9 — SYNTHETIC EXCLUSION RECORD**

Left Breast | Diagnosis: Invasive ductal carcinoma, Histologic grade II (Nottingham 7/9), Size: 5.2 cm, Lymphovascular invasion present (01/12/2023) | IHC: ER 95%, PR 80%, HER2 1+, Ki-67 22% (01/12/2023) | Genetic Test: BRCA1/BRCA2 – no mutations detected (03/05/2023) | Systemic Treatment: Letrozole + Palbociclib (started 03/01/2023, palliative treatment), disease under control at evaluation date, adverse events: mild fatigue, grade 2 neutropenia | Surgery: Left simple mastectomy, clear margins ≥5 mm, 12 lymph nodes resected (3 positive), residual tumor 3.2 cm, Histologic grade II, lymphovascular invasion present (09/05/2023) | Pathological Staging: pT2N1aM1 (09/05/2023) | Comorbidities: Type 2 diabetes mellitus, Systemic arterial hypertension, Dyslipidemia | ECOG: 1 (02/28/2024)

**PATIENT 10 — SYNTHETIC EXCLUSION RECORD**

Right Breast | Diagnosis: Invasive ductal carcinoma, Histologic grade II (Nottingham 7/9), Size: 3.8 cm, Lymphovascular invasion present (02/09/2023) | IHC: ER 100%, PR 95%, HER2 0, Ki-67 12% (02/09/2023) | Genetic Test: BRCA1/BRCA2 – no mutations detected (04/10/2023) | Adjuvant CT: AC protocol followed by Paclitaxel – Doxorubicin 60 mg/m² + Cyclophosphamide 600 mg/m² (03/15/2023 – 05/10/2023) and weekly Paclitaxel 80 mg/m² (05/22/2023 – 08/07/2023), mild adverse events (grade 1 fatigue, grade 2 alopecia) | Surgery: Right quadrantectomy + axillary lymph node dissection levels I–II, clear margins ≥3 mm, 15 lymph nodes resected (2 positive), residual tumor 2.4 cm, Histologic grade II, partial pathological response (10/03/2023) | Pathological Staging: ypT2N1aM0 (10/03/2023) | HT: Anastrozole 1 mg/day (started 11/01/2023) | Comorbidities: Systemic arterial hypertension, Hypothyroidism | ECOG: 1 (03/12/2024)

**PATIENT 11 — SYNTHETIC EXCLUSION RECORD**
 Left Breast | Diagnosis: Invasive ductal carcinoma, Histologic grade II (Nottingham 7/9), Size: 3.9 cm, Lymphovascular invasion present (03/02/2023) | IHC: ER 0%, PR 0%, HER2 3+ (positive), Ki-67 60% (03/02/2023) | Genetic Test: BRCA1/BRCA2 negative (05/05/2023) | Neoadjuvant CT: TCHP protocol – Docetaxel + Carboplatin + Trastuzumab + Pertuzumab (6 cycles, 04/10/2023 – 08/25/2023), adverse events: grade 2 nausea, grade 1 mucositis, grade 2 neutropenia, partial clinical response (~40%) | Surgery: Left modified radical mastectomy, clear margins ≥5 mm, 18 lymph nodes resected (3 positive), residual tumor 2.1 cm, Histologic grade II, partial pathological response (10/11/2023) | Pathological Staging: ypT2N1aM0 (10/11/2023) | Comorbidities: Systemic arterial hypertension, Grade I obesity | ECOG: 1 (02/18/2024)

**PATIENT 12 — SYNTHETIC EXCLUSION RECORD**
 Right Breast | Diagnosis: Invasive ductal carcinoma, Histologic grade III (Nottingham 8/9), Size: 4.8 cm, Lymphovascular invasion present (01/10/2023) | IHC: ER 10%, PR 0%, HER2 2+ (FISH positive), Ki-67 45% (01/10/2023) | Genetic Test: BRCA1/BRCA2 – no mutations detected (02/28/2023) | Neoadjuvant CT: TCHP protocol – Docetaxel + Carboplatin + Trastuzumab + Pertuzumab (6 cycles, 03/20/2023 – 08/15/2023), adverse events: grade 2 diarrhea, grade 3 neutropenia, LVEF reduction of 13 p.p., partial clinical response (~40%) | Surgery: Right simple mastectomy + axillary lymph node dissection levels I–II, clear margins ≥5 mm, 19 lymph nodes resected (5 positive), residual tumor 2.5 cm, Histologic grade II, partial pathological response (09/04/2023) | Pathological Staging: ypT2N2aM0 (09/04/2023) | Comorbidities: Systemic arterial hypertension, Peripheral venous insufficiency, Knee osteoarthritis | ECOG: 1 (02/15/2024)

**PATIENT 13 — SYNTHETIC EXCLUSION RECORD**
 Right Breast | Diagnosis: Invasive ductal carcinoma, Histologic grade III (Nottingham 8/9), Size: 4.9 cm, Lymphovascular invasion present (03/22/2023) | IHC: ER 100%, PR 95%, HER2 1+, Ki-67 10% (03/22/2023) | Genetic Test: BRCA1/BRCA2 negative (06/05/2023) | Neoadjuvant HT: Letrozole 2.5 mg/day (04/01/2023 – 09/30/2023), adverse events: grade 2 arthralgia, hot flashes, mild fatigue, partial clinical response (~25%) | Surgery: Right simple mastectomy + axillary lymph node dissection levels I–II, clear margins ≥5 mm, 15 lymph nodes resected (2 positive), residual tumor 3.8 cm, Histologic grade II, minimal pathological response (10/10/2023) | Pathological Staging: ypT3N1aM0 (10/10/2023) | Comorbidities: Systemic arterial hypertension, Type 2 diabetes mellitus, Compensated heart failure | ECOG: 2 (02/28/2024)

**PATIENT 14 — SYNTHETIC EXCLUSION RECORD**
 Left Breast | Diagnosis: Invasive ductal carcinoma, Histologic grade II (Nottingham 7/9), Size: 3.6 cm, Lymphovascular invasion present (04/20/2023) | IHC: ER 90%, PR 85%, HER2 2+ (FISH positive), Ki-67 28% (04/20/2023) | Genetic Test: BRCA1/BRCA2 – no mutations detected (06/05/2023) | Neoadjuvant CT: TCHP protocol – Docetaxel + Carboplatin + Trastuzumab + Pertuzumab (6 cycles, 06/10/2023 – 10/20/2023), adverse events: grade 2 fatigue, grade 2 neutropenia, grade 1 mucositis, partial clinical response (~40%) | Surgery: Left quadrantectomy + axillary lymph node dissection levels I–II, clear margins ≥4 mm, 16 lymph nodes resected (3 positive), residual tumor 2.1 cm, Histologic grade II, partial pathological response (12/15/2023) | Pathological Staging: ypT2N1aM0 (12/15/2023) | Comorbidities: Type 2 diabetes mellitus, Systemic arterial hypertension, Obesity | ECOG: 1 (02/22/2024)

**PATIENT 15 — SYNTHETIC EXCLUSION RECORD**
 Right Breast | Diagnosis: Invasive ductal carcinoma, Histologic grade II (Nottingham 7/9), Size: 5.1 cm, Lymphovascular invasion present (03/06/2023) | IHC: ER 95%, PR 90%, HER2 2+ (FISH positive), Ki-67 20% (03/06/2023) | Genetic Test: BRCA1/BRCA2 – no mutations detected (06/01/2023) | Neoadjuvant CT: TCHP protocol – Docetaxel + Carboplatin + Trastuzumab + Pertuzumab (6 cycles, 04/15/2023 – 09/30/2023), adverse events: grade 2 fatigue, grade 3 neutropenia, borderline cardiac toxicity (LVEF drop >10 p.p.), partial clinical response (~45%) | Surgery: Right modified radical mastectomy, clear margins ≥5 mm, 18 lymph nodes resected (4 positive), residual tumor 2.5 cm, Histologic grade II, partial pathological response (10/09/2023) | Pathological Staging: ypT2N2aM0 (10/09/2023) | Comorbidities: Type 2 diabetes mellitus, Systemic arterial hypertension, Grade I obesity | ECOG: 2 (02/25/2024)

**PATIENT 16 — SYNTHETIC EXCLUSION RECORD**
 Left Breast | Diagnosis: Invasive ductal carcinoma, Histologic grade III (Nottingham 8/9), Size: 3.3 cm, Lymphovascular invasion present (04/14/2023) | IHC: ER 0%, PR 0%, HER2 3+ (positive), Ki-67 65% (04/14/2023) | Genetic Test: BRCA1/BRCA2 negative (06/05/2023) | Neoadjuvant CT: TCHP protocol – Docetaxel + Carboplatin + Trastuzumab + Pertuzumab (6 cycles, 06/19/2023 – 10/20/2023), adverse events: grade 2 neutropenia, grade 1 fatigue, grade 1 mucositis, partial clinical response (~50%) | Surgery: Left quadrantectomy + axillary lymph node dissection levels I–II, clear margins ≥5 mm, 17 lymph nodes resected (3 positive), residual tumor 1.8 cm, Histologic grade II, partial pathological response (12/07/2023) | Pathological Staging: ypT1cN1aM0 (12/07/2023) | Comorbidities: Mild dyslipidemia | ECOG: 1 (02/26/2024)

**PATIENT 17 — SYNTHETIC EXCLUSION RECORD**
 Right Breast | Diagnosis: Invasive ductal carcinoma, Histologic grade II (Nottingham 7/9), Size: 5.0 cm, Lymphovascular invasion present (02/27/2023) | IHC: ER 90%, PR 85%, HER2 2+ (FISH positive), Ki-67 18% (02/27/2023) | Genetic Test: BRCA1/BRCA2 negative (04/15/2023) | Neoadjuvant CT: TCHP protocol – Docetaxel + Carboplatin + Trastuzumab + Pertuzumab (6 cycles, 05/01/2023 – 09/10/2023), adverse events: grade 2 fatigue, grade 2 nausea, grade 3 neutropenia, grade 2 cardiac toxicity (LVEF drop >10 p.p.), partial clinical response (~40%) | Surgery: Right simple mastectomy + axillary lymph node dissection levels I–II, clear margins ≥6 mm, 20 lymph nodes resected (5 positive), residual tumor 2.6 cm, Histologic grade II, partial pathological response (11/06/2023) | Pathological Staging: ypT2N2aM0 (11/06/2023) | Comorbidities: Systemic arterial hypertension, Type 2 diabetes mellitus, Mild heart failure | ECOG: 2 (02/29/2024)

**PATIENT 18 — SYNTHETIC EXCLUSION RECORD**
 Left Breast | Diagnosis: Invasive ductal carcinoma, Histologic grade II (Nottingham 7/9), Size: 5.0 cm, Involved margins (01/18/2023) | IHC: ER 100%, PR 90%, HER2 1+, Ki-67 15% (01/18/2023) | Genetic Test: BRCA1/BRCA2 negative (04/10/2023) | Palliative Systemic Treatment: Letrozole 2.5 mg/day + Palbociclib 125 mg/day (21/28 day cycles), started 03/01/2023, adverse events: grade 2 neutropenia, mild fatigue, stable clinical response | Surgery: Extended biopsy with partial excision, involved margins, lymph nodes not resected, residual tumor 4.6 cm (09/20/2023) | Pathological Staging: pT3NxM1 (single bone metastasis in L3) (09/20/2023) | Comorbidities: Systemic arterial hypertension, Osteoporosis | ECOG: 2 (02/10/2024)

**PATIENT 19 — SYNTHETIC EXCLUSION RECORD**
 Right Breast | Diagnosis: Invasive ductal carcinoma, Histologic grade III (Nottingham 8/9), Size: 4.2 cm, Lymphovascular invasion present (03/22/2023) | IHC: ER 0%, PR 0%, HER2 3+ (positive), Ki-67 70% (03/22/2023) | Genetic Test: BRCA1/BRCA2 negative (05/05/2023) | Neoadjuvant CT: TCHP protocol – Docetaxel + Carboplatin + Trastuzumab + Pertuzumab (6 cycles, 05/08/2023 – 09/18/2023), adverse events: grade 2 diarrhea, grade 2 fatigue, grade 3 neutropenia, significant LVEF drop, partial clinical response (~45%) | Surgery: Right modified radical mastectomy, clear margins ≥5 mm, 20 lymph nodes resected (5 positive), residual tumor 1.9 cm, Histologic grade II, partial pathological response (10/18/2023) | Pathological Staging: ypT1cN2aM0 (10/18/2023) | Comorbidities: Mild arterial hypertension | ECOG: 1 (03/03/2024)

**PATIENT 20 — SYNTHETIC EXCLUSION RECORD**
 Left Breast | Diagnosis: Invasive ductal carcinoma, Histologic grade II (Nottingham 7/9), Size: 4.6 cm, Lymphovascular invasion present (03/03/2023) | IHC: ER 90%, PR 90%, HER2 2+ (FISH positive), Ki-67 22% (03/03/2023) | Genetic Test: BRCA1/BRCA2 negative (05/10/2023) | Neoadjuvant CT: TCHP protocol – Docetaxel + Carboplatin + Trastuzumab + Pertuzumab (6 cycles, 05/20/2023 – 09/05/2023), adverse events: grade 2 neutropenia, grade 2 diarrhea, grade 2 fatigue, cardiac toxicity (LVEF drop >10 p.p.), partial clinical response (~45%) | Surgery: Left modified radical mastectomy, clear margins ≥5 mm, 19 lymph nodes resected (4 positive), residual tumor 2.3 cm, Histologic grade II, partial pathological response (11/14/2023) | Pathological Staging: ypT2N2aM0 (11/14/2023) | Comorbidities: Arterial hypertension, Type 2 diabetes mellitus | ECOG: 1 (02/25/2024)

**PATIENT 21 — SYNTHETIC EXCLUSION RECORD**
 Right Breast | Diagnosis: Invasive ductal carcinoma, Histologic grade III (Nottingham 8/9), Size: 5.3 cm, Lymphovascular invasion present (02/12/2023) | IHC: ER 95%, PR 90%, HER2 1+ (negative), Ki-67 12% (02/12/2023) | Genetic Test: BRCA1/BRCA2 negative (04/30/2023) | Systemic Treatment: Neoadjuvant hormone therapy with Letrozole 2.5 mg/day (03/15/2023 – 08/10/2023), adverse events: grade 2 arthralgia, hot flashes and mild fatigue, slight clinical response (~20%) | Surgery: Right simple mastectomy + axillary lymph node dissection levels I–II, clear margins ≥5 mm, 17 lymph nodes resected (4 positive), residual tumor 4.2 cm, Histologic grade II, minimal pathological response (09/01/2023) | Pathological Staging: ypT3N2aM0 (09/01/2023) | Comorbidities: Mild heart failure (ejection fraction 45%), Systemic arterial hypertension, Type 2 diabetes mellitus | ECOG: 2 (02/25/2024)

**PATIENT 22 — SYNTHETIC EXCLUSION RECORD**
 Left Breast | Diagnosis: Invasive ductal carcinoma, Histologic grade II (Nottingham 7/9), Size: 4.1 cm, Lymphovascular invasion present (03/08/2023) | IHC: ER 100%, PR 95%, HER2 1+ (negative), Ki-67 10% (03/08/2023) | Genetic Test: BRCA1/BRCA2 negative (05/25/2023) | Neoadjuvant CT: AC-T protocol — Doxorubicin 60 mg/m² + Cyclophosphamide 600 mg/m² (4 cycles, 04/15/2023 – 06/30/2023) followed by weekly Paclitaxel 80 mg/m² (12 weeks, 07/10/2023 – 09/30/2023); adverse events: grade 1 fatigue, grade 2 alopecia; partial clinical response (~40%) | Surgery: Left quadrantectomy + axillary lymph node dissection levels I–II, clear margins ≥5 mm, 15 lymph nodes resected (2 positive), residual tumor 2.5 cm, Histologic grade II, partial pathological response (10/22/2023) | Pathological Staging: ypT2N1aM0 (10/22/2023) | Comorbidities: Mild arterial hypertension, Dyslipidemia | ECOG: 1 (03/02/2024)

**PATIENT 23 — SYNTHETIC EXCLUSION RECORD**
 Right Breast | Diagnosis: Invasive ductal carcinoma, Histologic grade II (Nottingham 7/9), Size: 5.2 cm, Lymphovascular invasion present (02/03/2023) | IHC: ER 100%, PR 90%, HER2 1+ (negative), Ki-67 14% (02/03/2023) | Genetic Test: BRCA1/BRCA2 negative (04/15/2023) | Systemic Treatment: Neoadjuvant hormone therapy with Letrozole 2.5 mg/day (04/01/2023 – 09/15/2023), adverse events: hot flashes and grade 2 arthralgia, modest clinical response (~25%) | Surgery: Right simple mastectomy + axillary lymph node dissection levels I–II, clear margins ≥4 mm, 16 lymph nodes resected (3 positive), residual tumor 4.1 cm, Histologic grade II, minimal pathological response (09/12/2023) | Pathological Staging: ypT3N1aM0 (09/12/2023) | Comorbidities: Mild heart failure (LVEF 47%), Systemic arterial hypertension, Type 2 diabetes mellitus | ECOG: 2 (02/27/2024)

**PATIENT 24 — SYNTHETIC EXCLUSION RECORD**
 Left Breast | Diagnosis: Invasive ductal carcinoma, Histologic grade II (Nottingham 7/9), Size: 5.2 cm, Lymphovascular invasion present (01/18/2023) | IHC: ER 100%, PR 95%, HER2 1+ (negative), Ki-67 12% (01/18/2023) | Genetic Test: BRCA1/BRCA2 negative (04/05/2023) | Systemic Treatment: Neoadjuvant hormone therapy with Letrozole 2.5 mg/day (04/01/2023 – 08/31/2023); adverse events: mild arthralgia and hot flashes; modest clinical response (~30%) | Surgery: Left simple mastectomy + axillary lymph node dissection levels I–II, clear margins ≥4 mm, 18 lymph nodes resected (4 positive), residual tumor 4.3 cm, Histologic grade II, minimal pathological response (09/12/2023) | Pathological Staging: ypT3N2aM0 (09/12/2023) | Comorbidities: Mild heart failure (LVEF 46%), Arterial hypertension, Type 2 diabetes mellitus | ECOG: 2 (02/27/2024)

**PATIENT 25 — SYNTHETIC EXCLUSION RECORD**
 Right Breast | Diagnosis: Invasive ductal carcinoma, Histologic grade II (Nottingham 7/9), Size: 4.4 cm, Lymphovascular invasion present (04/10/2023) | IHC: ER 95%, PR 90%, HER2 3+ (positive), Ki-67 35% (04/10/2023) | Genetic Test: BRCA1/BRCA2 negative (06/20/2023) | Neoadjuvant CT: TCHP regimen — Docetaxel + Carboplatin + Trastuzumab + Pertuzumab (6 cycles, 06/20/2023 – 10/10/2023); adverse events: grade 3 neutropenia, grade 2 diarrhea, grade 2 fatigue, cardiac toxicity (LVEF drop >10 p.p.); partial clinical response (~50%) | Surgery: Right modified radical mastectomy, clear margins ≥5 mm, 21 lymph nodes resected (4 positive), residual tumor 2.0 cm, Histologic grade II, partial pathological response (12/18/2023) | Pathological Staging: ypT1cN2aM0 (12/18/2023) | Comorbidities: Systemic arterial hypertension, Dyslipidemia | ECOG: 1 (03/01/2024)

**SYNTHETIC PATIENT GENERATOR PROMPT - OLYMPIA**

**Context:**
You are a healthcare professional specialized in clinical oncology and translational research, responsible for screening oncology patients according to the inclusion and exclusion criteria of a clinical study. Your role is to accurately analyze clinical records, ensuring full compliance with the study protocol.

**Objective (your task):**
Carefully review the patient’s medical record and determine eligibility based on the following inclusion and exclusion criteria:

**Inclusion Criteria:**

- Histologically confirmed non-metastatic primary invasive adenocarcinoma of the breast, of one of the following phenotypes:
  - Triple-negative breast cancer defined as ER and PgR negative and HER2 negative (not eligible for anti-HER2 therapy)
  - ER and/or PgR positive, HER2 negative
- Documented germline mutation in BRCA1 or BRCA2 that is predicted or suspected to be deleterious (i.e., known or likely to be detrimental or lead to loss of function)
- Completion of adequate surgery of the breast and axilla
- Completion of at least 6 cycles of neoadjuvant or adjuvant chemotherapy containing anthracyclines, taxanes, or both. Prior use of platinum agents is permitted if given with curative intent for another cancer (e.g., ovarian), or as neoadjuvant or adjuvant treatment for breast cancer
- ECOG performance status 0 or 1

**Exclusion Criteria:**

- Any prior treatment with a PARP inhibitor (including olaparib) or known hypersensitivity to any of the excipients in the study treatment
- Diagnosis of a second primary malignancy, except for:
  - Adequately treated non-melanoma skin cancer
  - Curatively treated in situ carcinoma of the cervix
  - Ductal carcinoma in situ (DCIS) of the breast
  - Stage 1, grade 1 endometrial carcinoma
  - Other solid tumors or lymphomas (without bone marrow involvement) diagnosed at least 5 years prior to randomization and treated without recurrence, with no more than one line of chemotherapy
- Concomitant use of known strong CYP3A inhibitors (such as itraconazole, telithromycin, clarithromycin, protease inhibitors boosted with ritonavir or cobicistat, indinavir, saquinavir, nelfinavir, boceprevir, or telaprevir), or moderate inhibitors (such as ciprofloxacin, erythromycin, diltiazem, fluconazole, or verapamil). A washout period of 2 weeks is required prior to starting study treatment
- Concomitant use of strong CYP3A inducers (such as phenobarbital, enzalutamide, phenytoin, rifampin, rifabutin, rifapentine, carbamazepine, nevirapine, or St. John’s Wort), or moderate inducers (such as bosentan, efavirenz, or modafinil). A washout period of 5 weeks is required for enzalutamide or phenobarbital, and 3 weeks for other agents
- Evidence of metastatic breast cancer

**Decision Logic:**
Patients will be classified as:

- **Included** if they meet all inclusion criteria and none of the exclusion criteria
- **Excluded** if they do not meet at least one inclusion criterion or meet at least one exclusion criterion

**Style:**
The screening report should follow the formal, medical-technical style used in oncology records, employing standardized, objective medical terminology. Sentences should be short, precise, and avoid unnecessary adjectives or subjective language.

**Tone:**
Formal and clinical. The tone should reflect that of oncologists and clinical researchers.

**Target Audience:**
Healthcare professionals involved in oncology research and clinical screening activities, especially research nurses, physician-investigators, and clinical trial coordinators.

**Response Format:**
The screening report should be structured as follows:

Screening outcome: Included or Excluded
Inclusion criteria met: Clear, objective summary of which inclusion criteria are met
Exclusion criteria met: Clear, objective summary of which exclusion criteria are met (if any)
Decision summary: Concise justification for the screening decision, written in clinical language

**Purpose of the Prompt:**
To generate oncological screening reports using standardized medical language, based on the eligibility criteria defined by the study protocol. These reports are intended to support the training and validation of artificial intelligence models applied to oncology clinical research and trial screening.
